# Shared genetic architecture between brain iron deposition and Alzheimer’s disease

**DOI:** 10.64898/2026.09.07.26362470

**Authors:** Md Rezanur Rahman, Ying Xia, Parnesh Raniga, Amir Fazlollahi, Tania Islam, Aishwarya Dhruva, Jue-Sheng Ong, Stuart MacGregor, Ashley I. Bush, Jurgen Fripp, Michelle K Lupton

## Abstract

Brain iron accumulation is well established in Alzheimer’s disease (AD), but whether the two share a genetic basis remains unclear. Here, we performed genome-wide association analyses of brain iron across six iron-rich brain regions, quantified via quantitative susceptibility mapping (QSM) in 38,142 UK Biobank participants. Genetic pleiotropic overlap, genetic correlation and genomic structural equation modelling were used to identify shared genetic architecture between brain iron and risk of AD, and bidirectional Mendelian randomisation was used to test for causal relationships. We identified 116 independent genome-wide significant loci associated with brain iron (*r^2^*<0.1; p <5 × 10^-8^) and validated these signals in an independent cohort using polygenic scores, which explained between 8.9% and 25.4% of variance in regional iron. Genetic pleiotropy and correlation analyses revealed that brain iron and AD share a polygenic basis extending well beyond the canonical *APOE* locus. This shared liability is specific to basal ganglia structures relevant to cognition, specifically, iron deposition across the caudate nucleus, putamen, and globus pallidus loaded onto a single latent factor that correlated significantly with AD (*r*g = 0.13, *P* = 0.019). Multivariate genome-wide association analysis of the shared liability factor between AD and brain iron revealed 27 independent genome-wide significant loci implicating genes that modulate neuroimmune function. Mendelian randomisation provided no evidence of a causal effect in either direction. In summary, we identified region-specific shared genetic liability between brain iron and AD, and a putative neuroimmune mechanism underlying it.

## Introduction

Alzheimer’s disease (AD) is the predominant progressive neurodegenerative disease characterised by impaired cognitive function and decreased quality of life^1^. It has been observed that in AD patients, higher levels of brain amyloid-β identified by PET correlate with cognitive decline, however, large variability in the rate of cognitive decline suggests other molecular players in collaboration with amyloid-β might be associated with the accelerated clinical decline^2,3^. Accumulation of increased iron in cortical brain regions has been associated with neurodegeneration, potentially involving mitochondrial dysfunction, oxidative stress, and/or neuroinflammation pathways^4,5^. Previous studies have shown that higher levels of cerebrospinal fluid ferritin, a marker of brain iron burden, predicted decreased cognition and an increased risk of AD^6,7^. Recent studies leveraging quantitative susceptibility mapping (QSM) of MRI data and amyloid-β PET have shown that higher QSM values in the hippocampus, temporal, and frontal lobes predicted greater decline in the longitudinal cognitive performance of subjects who had underlying amyloid-β pathology^8^. In addition, higher QSM susceptibility was observed in caudate and putamen basal ganglia regions in mild to moderate AD patients compared to healthy controls, and elevated QSM susceptibility in the left caudate showed correlation with cognitive decline^9,10^.

Despite growing evidence implicating iron dysregulation in neurodegenerative diseases, the molecular mechanisms linking brain iron accumulation to AD pathogenesis remain poorly characterised^11^. Iron is an essential trace element for normal brain function, supporting neurotransmitter synthesis, oxygen transport, and myelin formation^12^. However, when iron accumulates excessively in brain tissue, particularly within the basal ganglia, it may promote oxidative stress through the generation of toxic reactive oxygen species^4^. These reactive species in turn may interfere with mitochondrial function and trigger neuroinflammatory responses, including microglial activation^13^. There is also emerging evidence that excess iron may enhance amyloid-beta aggregation and tau phosphorylation, which are key pathological features of AD^14–16^. Consistent with this, higher subcortical iron deposition measured by QSM has been associated with increased amyloid burden in the pallidum and putamen^14^, and with tau aggregation in regions typically affected in AD^16^. However, other studies suggest that QSM may more strongly reflect neuroinflammatory processes rather than direct protein aggregation^17^, and this relationship remains an area of active investigation. Taken together, brain iron dysregulation may contribute to neurodegenerative disease pathogenesis through converging pathways involving oxidative damage and neuroinflammation. However, the causal role of iron is not settled. Recent evidence suggests that iron elevation may be a downstream consequence of disease rather than a primary driver. Notably, iron chelation with brain permeable deferiprone worsened cognition and executive function in AD, which argues against a simple model where iron drives neurodegeneration^5^. Separating cause from consequence therefore benefits from genetic approaches. Investigating the genetic relationship between regional brain iron levels and AD risk is an important step toward clarifying the role of iron in neurodegeneration.

QSM is an MRI-based non-invasive technique that measures tissue magnetic susceptibility^18^. It has been shown to provide reliable quantification of brain iron content in deep grey matter structures^18,19^. Compared to other iron-sensitive MRI methods such as susceptibility-weighted imaging and R2*, QSM is more selective for iron load, and offers superior spatial specificity and is less dependent on the object orientation and field strength^20,21^. These properties suggest that QSM could be a superior tool for studying regional brain iron accumulation in large population cohorts. Previous studies have reported that brain iron levels measured by QSM are substantially heritable, indicating a genetic component underlying regional iron deposition^22,23^. Genome-wide association studies (GWAS) of QSM-derived brain iron have emerged to identify genetic loci associated with subcortical iron accumulation^23,24^, however, systematically examination of the shared genetic liability between regional brain iron and AD remains to be investigated. A prior QSM study reported elevated iron in the caudate and putamen in AD patients, with iron levels correlating with cognitive decline^9^. Another study found that neocortical amyloid-β burden colocalized with elevated cortical QSM, and that in subjects positive for amyloid-β, hippocampal QSM predicted cognitive decline over 6 years^8^. The genetic underpinnings of these associations have remained unknown. Addressing this gap may reveal causal pathways and mechanisms linking iron dysregulation to AD and could nominate therapeutic targets. Thus, in the present study, we investigated the shared genetic architecture between iron-rich subcortical regions and AD risk.

## Materials and Methods

### Study sample

UK Biobank (UKB) is a large prospective cohort study of approximately 500,000 UK adults from across the United Kingdom aged 40–70 years at baseline assessment (2006–2010)^25^. The UKB data resource includes demographic and lifestyle characteristics, biomarkers, genetics, and linked electronic health records. A subset of participants completed brain MRI data acquisition (data collected 2014–2022). UKB data are available to researchers following application and ethics approval. The use of UKB data was approved by the QIMR Berghofer Research Ethics Committee. The present analyses were conducted under linked UKB Applications 27483 and 25331.

### MRI data processing

Regional brain iron quantification was performed using T1-weighted structural and susceptibility-weighted MRI acquisitions. The susceptibility-weighted data were acquired using a 3D dual-echo gradient echo (GRE) sequence on identical 3T Siemens Skyra scanners with 32-channel head receive coils. Acquisition parameters included echo times (TE_1_/TE_2_) = 9.4/19.7 *ms*, repetition time = 27 *ms*, flip angle = 15°, an in-plane parallel imaging acceleration factor of 2, voxel size = 0.8 × 0.8 × 3.0 *mm^3^*, and matrix size = 256 × 288 × 48.

QSM images were reconstructed from the raw phase and magnitude images of the second echo (TE_2_ = 19.7 *ms*) using a containerized in-house pipeline based on the STI Suite toolbox. Briefly, a 3D magnitude image was generated via root-sum-of-squares across channels, followed by N4 bias field correction, and then used for brain mask extraction. Each channel’s phase data were independently rescaled and unwrapped using Laplacian phase unwrapping^26^, then combined into a single combined phase image via sensitivity-weighted coil combination, where sensitivity maps were derived from per-coil magnitude images. Background field removal was performed using V-SHARP^27^, and dipole inversion was performed using the iterative least-squares (iLSQR) algorithm^28^ with 50 regularisation iterations. Susceptibility values are reported in parts-per-billion (ppb).

Regional QSM quantification was performed by co-registering the JHU QSM atlas to each QSM map using a combined affine and non-rigid registration framework. The JHU QSM atlas delineates iron-rich deep grey matter structures using the Eve Parcellation Map (EvePM)^29^. Median susceptibility values were extracted from the registered images across twelve highly heritable, iron-rich subcortical regions of interest, including the bilateral caudate nucleus, putamen, globus pallidus, substantia nigra, red nucleus and dentate nucleus. The median susceptibility values extracted from a region within the middle-frontal white matter were used for normalizing QSM measures^8^. An automated quality control protocol was applied to exclude participants with extreme statistical outliers (i.e., outside the central 99.6% distribution) in more than two regions. Lastly, regional QSM measures from the left and right hemisphere were averaged to yield a single, bilateral phenotype for each subcortical structure in subsequent analyses.

### Genotype data and genome-wide association studies

Genotyping was used the UK Biobank Axiom Array (Affymetrix). UK Biobank genotyping was imputed to the Haplotype Reference Consortium (HRC) panel using IMPUTE2. Analyses were restricted to individuals of White British ancestry inferred via ancestral principal component clustering, using the standard exclusion list applied in our laboratory. We did not exclude related individuals, because BOLT-LMM^30^ accounts for relatedness through the mixed model. In the association step, we kept imputed variants with INFO ≥ 0.3 and MAF ≥ 0.01. We ran a separate GWAS for each of the six QSM-derived regional brain iron phenotypes. We used BOLT-LMM and applied the non-infinitesimal Bayesian mixture-of-normals model, which improves power over the standard infinitesimal model^30^. BOLT-LMM builds a genetic relationship matrix from directly genotyped SNPs and applies a leave-one-chromosome-out scheme to account for population structure and cryptic relatedness. We adjusted each GWAS for age, genetic sex, genotyping array, and the first ten genetic principal components. We also included the middle-frontal white matter susceptibility value as a per-participant quantitative covariate, which normalises each regional phenotype against the QSM reference region and accounts for between-subject differences.

### Functional annotation and gene mapping

We uploaded the GWAS summary statistics from each regional analysis to FUMA for post-GWAS annotation and locus definition^31^. We used FUMA default settings with the 1000 Genomes European reference panel. First, we selected genome-wide significant SNPs (P < 5×10⁻⁸). From these, we defined independent significant SNPs as those independent from each other at r² < 0.6. Lead SNPs were a subset of the independent significant SNPs, further pruned at r² < 0.1. We then defined genomic risk loci by merging lead SNPs that were within 250 kb of each other. All lead SNPs were therefore independent. To identify unique loci across the six regional GWAS, loci identified across all six regional GWAS were pooled and merged into a set of unique genomic intervals. Two loci were considered the same if they fell within 500 kb of each other. All merged loci were assessed for replication against two prior QSM GWAS^23,24^. For each prior study, we extracted QSM-associated SNPs and built ±500 kb windows around each SNP, then merged these into replication intervals. A merged locus replicated in a study if it overlapped any interval from that study. We considered a locus novel when it overlapped no interval from either study.

### Polygenic risk score analysis

Polygenic risk scores (PRS) for each of the six QSM-derived iron phenotypes were constructed using SBayesRC^32^. SBayesRC is a Bayesian shrinkage method that incorporates functional genomic annotations to improve prediction accuracy. PRS weights were derived from the regional GWAS summary statistics obtained from the UKB discovery sample and applied to genotype data from an independent cohort, the Prospective Imaging Study of Ageing (PISA)^33^ for out-of-sample validation. PRS were standardised prior to analysis. PRS performance was assessed as the incremental variance explained (ΔR²), calculated as the difference in R² between a full model including the PRS and a null model containing age, sex, years of education, and the first ten ancestry principal components. Both same-region (PRS for region *i* predicting QSM for region *i*) and cross-region (PRS for region *i* predicting QSM for region *j*, *i* ≠ *j*) predictions were evaluated. Statistical significance was assessed using a Bonferroni-corrected threshold of *P* < 1.39 × 10⁻³.

### Gene-based analyses

We used two complementary gene-based methods, Multi-marker Analysis of GenoMic Annotation (MAGMA) and Summary data-based Mendelian randomisation (SMR) to prioritise genes associated with traits. Firstly, we performed MAGMA^34^ within FUMA^31^. MAGMA aggregates SNP-level *P*-values within gene boundaries into a gene-level statistic using a multiple regression model. Gene windows were defined as 35 kb upstream and 10 kb downstream of each protein-coding gene boundary to capture regulatory variants in proximal promoter regions. Gene-based tests were performed using the 1000 Genomes Phase 3 European panel as the LD reference. Statistical significance was defined by Bonferroni correction for the number of protein-coding genes tested in MAGMA.

We also performed SMR to prioritise putative causal genes associated with trait of interest^35^. SMR integrates GWAS summary statistics with cis-eQTL data to test whether the association between the GWAS signal and gene expression is consistent with a shared causal variant. Two eQTL datasets, specifically whole blood (eQTLGen consortium; *n* = 31,684) and brain cortex (BrainMeta v2; *n* = 2,865), were used. To distinguish true pleiotropy from confounding by linkage disequilibrium (LD), the HEIDI (Heterogeneity in Dependent Instruments) test was applied. Genes were retained as statistically significant if *P*_HEIDI_ > 0.01 with ≥ 5 SNPs in the HEIDI test. Bonferroni-corrected significance thresholds were applied separately by eQTL dataset (eQTLGen: *P*_SMR < 3.19 × 10⁻⁶; BrainMeta: *P*_SMR < 3.07 × 10⁻⁶).

### AD GWAS summary statistics

We used two published AD GWAS summary statistics datasets throughout this study. Kunkle et al. (2019) included only clinically diagnosed AD cases and controls (21,982 cases / 41,944 controls), providing high diagnostic specificity^36^. Jansen et al. (2019) augmented confirmed cases with proxy cases defined by parental history of AD from the UK Biobank (71,880 cases including proxies and 383,378 controls), increasing statistical power at the cost of phenotypic precision^37^. Because each dataset captures different aspects of AD genetic liability, we applied both to improve robustness of inference across analyses.

### Genetic pleiotropy analysis with AD

We tested for shared genetic effects between regional brain iron and AD using GPA (Genetic analysis incorporating Pleiotropy and Annotation)^38^. GPA requires only SNP-level p-values from two GWAS as input. For each SNP, it models association as one of four latent groups: associated with neither trait, iron only, AD only, or both. It estimates the proportion of SNPs in each group and SNP ranking is achieved by an EM algorithm (π₁₀ iron only, π₀₁ AD only, π₁₁ both). We paired each regional iron GWAS with each AD GWAS^36,37^, giving six regions across two datasets. We tested for pleiotropy using a likelihood-ratio test that compares the fitted model against a null in which iron and AD association are independent. A significant test indicates more shared associations than expected by chance. We summarised the magnitude of overlap using the pleiotropic association ratio (PAR), defined as π₁₁ / (π₁₀ + π₀₁ + π₁₁). PAR is the proportion of all iron- or AD-associated variants that are associated with both traits. We ran each analysis with and without the *APOE* region (chr19: 44.9–45.9 Mb, GRCh37).

### LD score regression and genetic correlation analysis

We estimated genetic correlations between regional brain iron measures and AD using two complementary methods: bivariate linkage disequilibrium score regression (LDSC)^39^ and high-definition likelihood (HDL)^40^. LDSC estimates genetic correlation using the relationship between test statistics and LD scores. HDL uses a larger proportion of the genome, thereby reducing standard errors and improving precision. For LDSC, we pre-processed AD GWAS summary statistics and QSM GWAS summary statistics, respectively, using munge_sumstats.py and restricted analyses to HapMap3 SNPs with MAF > 0.01. We used pre-computed LD scores from the 1000 Genomes European reference panel. For HDL, we used the UK Biobank-based LD reference panel. From the same LDSC pipeline, we also estimated SNP-based heritability for each regional iron phenotype using univariate LDSC. QSM-derived iron is a quantitative trait, so we report heritability estimate on the observed scale. We ran all analyses twice, including and excluding the *APOE* locus (chr19: 44.9–45.9 Mb, GRCh37). Significance of genetic correlation estimates was defined at *P* < 0.05.

### Genomic structural equation modelling

To model shared genetic architecture between AD and QSM GWAS, we used Genomic structural equation modelling (Genomic SEM)^41^. Using a genetic covariance matrix, we modelled latent factor structures via multivariable LDSC. GWAS summary statistics for all six QSM phenotypes and AD datasets were harmonised to HapMap3 SNPs. We first tested a global latent factor model (F_Iron) in which all six regional iron measures loaded onto a single common factor. We also specified two circuit-specific factors: a factor with cognitive relevant regions comprising basal ganglia structures (caudate nucleus, putamen, globus pallidus), and a motor circuit-relevant factor comprising brainstem and cerebellar regions (substantia nigra, red nucleus, dentate nucleus). Model fit was assessed using the Comparative Fit Index (CFI ≥ 0.90) and Standardised Root Mean Square Residual (SRMR ≤ 0.08). Next, to quantify the shared genetic liability between basal ganglia iron and AD, a higher-order latent factor model was constructed in which basal ganglia iron latent factor and AD statistics loaded onto a shared latent factor. A multivariate GWAS of AD-Iron was performed in Genomic SEM using genome-wide SNP data.

### GO and pathway enrichment analysis

We performed all enrichment analyses using Metascape^42^, using genes underlying the AD-iron shared latent factor as input and all genes in the genome as the enrichment background. For pathway and process enrichment, we tested gene lists against ontology sources. For protein– protein interaction (PPI) network analysis, we used STRING (physical score > 0.132) and BioGrid, retaining only physical interactions. The resulting network included proteins forming at least one physical interaction with another member of the input gene list. We applied the Molecular Complex Detection (MCODE) algorithm to identify densely connected network modules. For each MCODE module, we retained the three best-scoring pathway terms as functional descriptors. For gene list enrichment, we tested the AD-iron gene set against three annotation categories: Cell Type Signatures, DisGeNET disease associations and PaGenBase tissue-specific expression. For pathway and gene list enrichment analyses, *P*-values were calculated using the hypergeometric distribution. Terms were retained if they met all three criteria: *P* < 0.01, a minimum gene count of 3, and an enrichment factor > 1.5. Statistical significance was considered after adjusted for multiple testing using the Benjamini–Hochberg method.

### Mendelian randomisation analysis

We performed bidirectional two-sample Mendelian randomisation (MR) to evaluate causal relationships between regional brain iron deposition and AD using MendelianRandomisation R package. Genetic instruments for each QSM-derived iron phenotype and the basal ganglia common factor were selected from the respective GWAS as genome-wide significant SNPs (*P* < 5 × 10⁻⁸), followed by linkage disequilibrium (LD) clumping (r^2^ <0.1) using the 1000 Genomes European reference panel. Instruments with F-statistic < 10 were considered weak and were excluded from the analysis. We used AD GWAS summary statistics from Jansen et al. (2019) as the primary outcome in forward direction (iron to AD). We used Kunkle et al. (2019) as a sensitivity dataset, because it has no UK Biobank overlap and gives replication free from overlap bias. We removed the *APOE* region from the AD summary statistics before MR. We used these APOE-free statistics in both directions: as the outcome in forward direction, and as the instrument source in reverse direction (AD to iron). This removed *APOE* as an AD instrument in reverse MR, where *APOE* is the major AD risk locus and is strongly pleiotropic. In the iron GWAS, no SNP reached genome-wide significance in the *APOE* region (chr19:44.9–45.9 Mb, GRCh37; minimum P = 4×10⁻⁵ across the six regional iron phenotypes and the basal ganglia factor), so *APOE* did not enter the iron instruments in forward MR. We applied the same estimators in both directions. IVW was the primary estimator. We ran MR-Egger regression, weighted median, simple mode and weighted mode as sensitivity estimators. We used the MR-Egger intercept to test directional pleiotropy. We applied Cochran’s Q to detect heterogeneous instruments and re-ran IVW and MR-Egger on the Q-homogeneous instruments only. The proxy-augmented dataset may overlap partially with our discovery analysis. We therefore applied MRlap to all Jansen-based analyses, to correct for sample-overlap bias. We did not apply MRlap to the clinically diagnosed dataset, which has no UK Biobank overlap. We controlled FDR using Benjamini Hochberg method across MR tests.

## Results

### Genome-wide association study of brain iron

To identify genetic loci associated with brain iron levels, we performed a GWAS of brain iron-rich regions leveraging QSM susceptibility measures in the UKB. Briefly, QSM of brain iron levels were estimated from MRI images of deep subcortical regions in UKB participants. Next, we performed GWAS analyses of QSM-derived brain iron using imputed genotype data. Our GWAS analysis identified 211 genome-wide significant independent loci (*r²* < 0.1, 250 kb window; p < 5×10⁻⁸) across six subcortical regions (**Fig. 1 and Fig. 2, Supplementary Table 1**). After merging loci within 500 kb across regions, these reduced to 116 independent loci (**Supplementary Table 2**). Of the 116 unique loci, 41 were novel, 75 replicated in prior studies^23,24^ (**Supplementary Table 2**).

**Figure 1.**
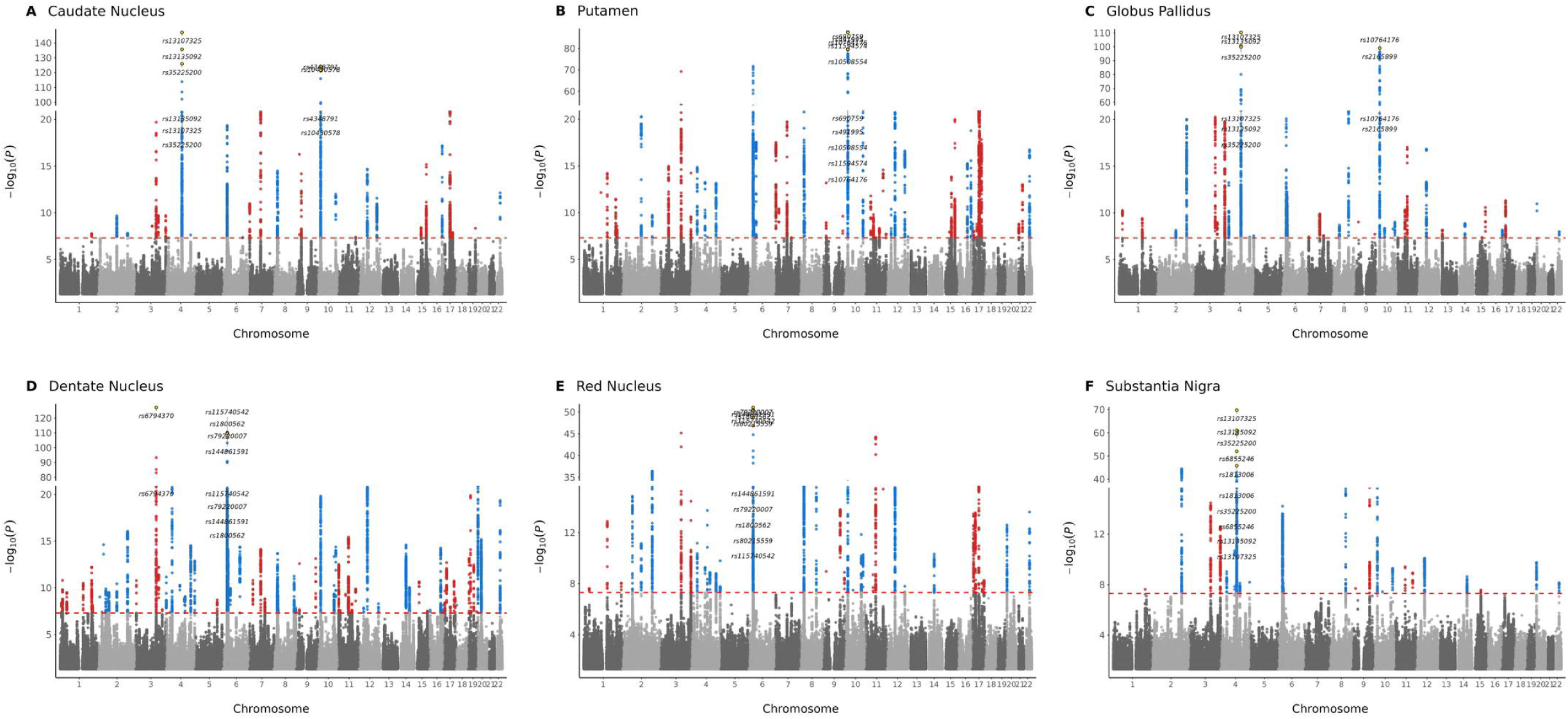
Genome-wide association results of brain iron across iron rich subcortical regions. Manhattan plots for (**A**) caudate nucleus, (**B**) putamen, (**C**) globus pallidus, (**D**) dentate nucleus, (**E**) red nucleus and (**F**) substantia nigra. The *x*-axis shows genomic position and the y-axis shows statistical significance as *−log₁₀(P). The r*ed dashed line marks the genome-wide significance threshold (*P <* 5 × 10^−8^)

**Figure 2.**
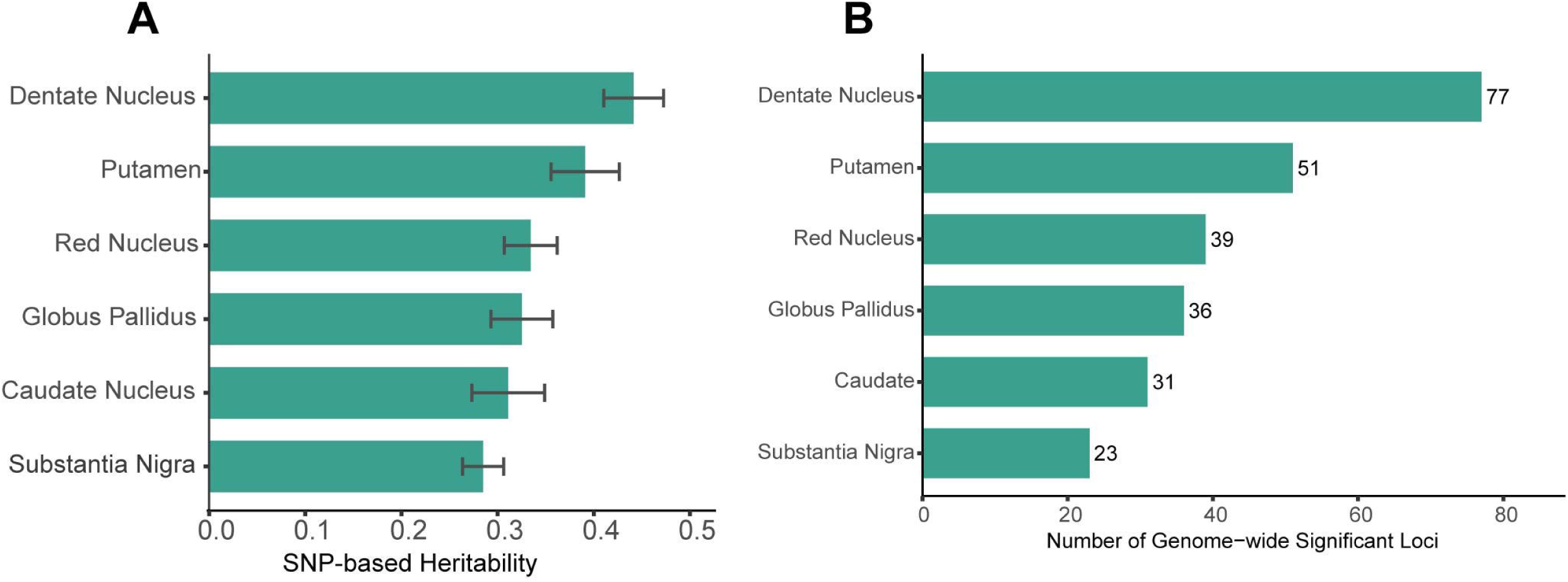
Genetic architecture of QSM derived brain iron across iron rich subcortical regions. (A) Bar plot represents SNP-based heritability estimate, (B) Number of genome-wide significant independent loci.

### Polygenic risk score of brain iron

To validate the genetic architecture identified in our GWAS, we constructed genome-wide PRS for each of the six QSM-derived iron phenotypes using SBayesRC. PRS were derived from the regional GWAS summary statistics and validated in an independent cohort, the Prospective Imaging Study of Ageing (n = 268). Brain iron PRS significantly predicted QSM-derived iron levels across all six subcortical regions in PISA participants (**Fig. 3**, **Supplementary Table 3**). Same-region PRS explained between 8.9% (substantia nigra) and 25.4% (dentate nucleus) of variance in regional iron levels (ΔR², all Bonferroni-corrected *p* < 0.05) (**Fig. 3**). Next, we evaluated cross-regional PRS and QSM, in which PRS derived from one region was tested against QSM in another to characterise the extent of shared genetic architecture across subcortical structures (**Supplementary Table 3**). Most cross-region pairs were significant (27 of 30, Bonferroni-corrected), with the strongest prediction from the putamen PRS to caudate iron (ΔR² = 19.9%) (**Fig. 3A**).

**Figure 3.**
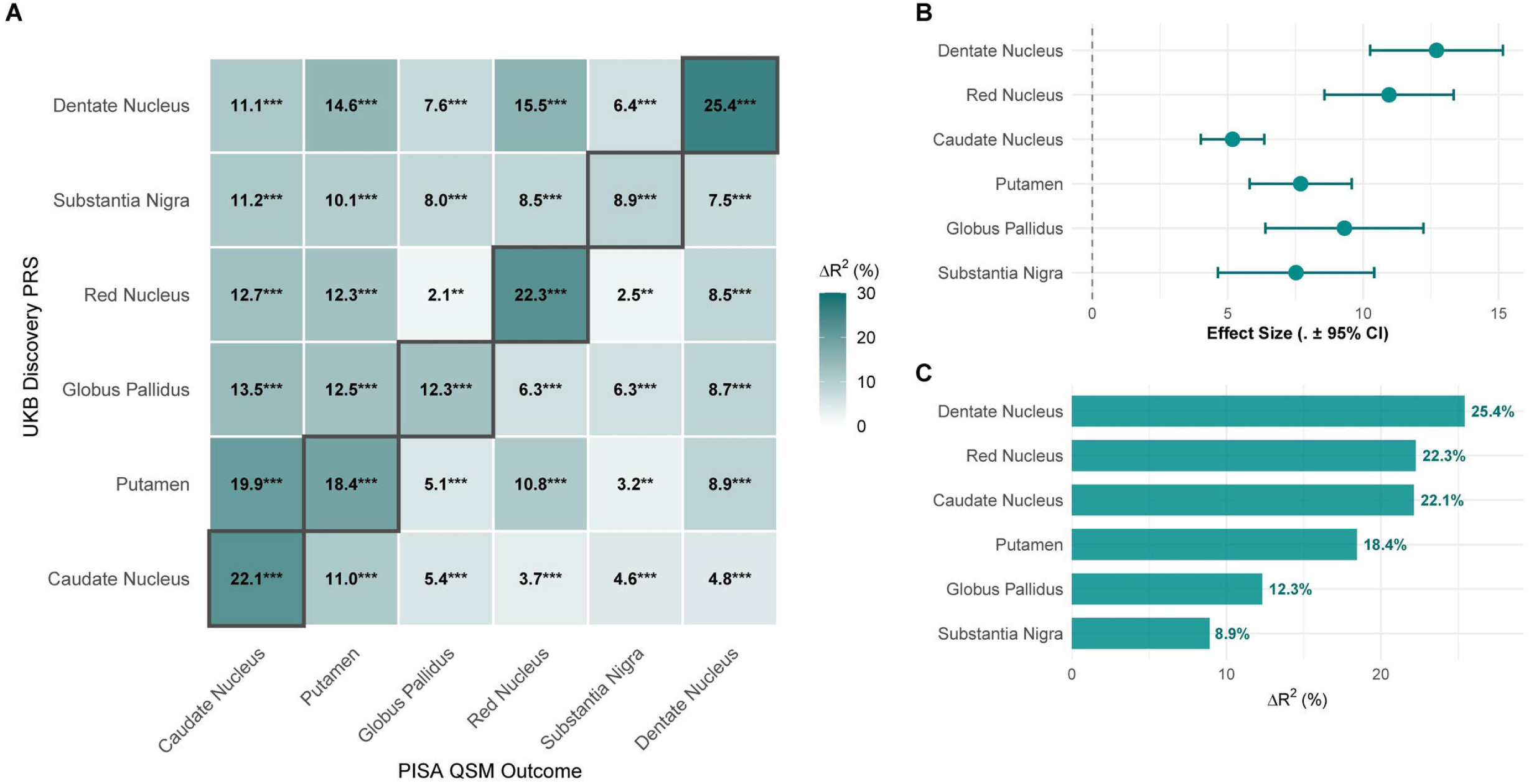
Independent validation of brain iron polygenic risk scores. (**A**) Heatmap showing incremental variance explained (ΔR²) by each regional PRS in predicting QSM-measured brain iron in the independent PISA cohort (N=268). Diagonal cells (black border) represent matched-region validation where the PRS discovery and outcome regions are identical. All matched-region associations and multiple cross-regional associations survived Bonferroni correction for 36 tests. ***P < 0.05/36; **FDR < 0.05; *P < 0.05. (**B**) Bar plot shows the effect size, (C) Incremental ΔR² by each regional PRS in predicting QSM-measured brain iron in the independent PISA cohort.

### Prioritisation of brain iron genes

To prioritise genes associated with brain iron, we performed MAGMA gene-based analysis. After Bonferroni correction (P < 2.59 × 10⁻⁶), we identified 154, 239, 163, 339, 145, and 81 significant genes, for caudate nucleus, putamen, globus pallidus, dentate nucleus, red nucleus and substantia nigra, respectively (**Supplementary Table 4**). Next, we performed SMR analysis to prioritise putative causal genes for brain iron levels using gene expression eQTL datasets from whole blood (eQTLGen, n = 31684) and brain cortex (BrainMeta, n = 2865). The HEIDI test was applied to distinguish pleiotropy from linkage. After Bonferroni correction for the number of unique genes tested (eQTLGen: P < 3.19 × 10⁻⁶; BrainMeta: P < 3.07 × 10⁻⁶) and HEIDI filtering (P > 0.01, nSNP ≥ 5), we identified 22, 40, 29, 56, 28 and 15 genes for caudate nucleus, putamen, globus pallidus, dentate nucleus, red nucleus and substantia nigra, respectively (**Supplementary Table 5**).

### Genetic overlap and correlation analysis with AD

To investigate the shared genetic basis between regional brain iron deposition and AD, we performed genetic pleiotropy analysis using GPA, with two independent AD GWAS datasets, Jansen et al. (2019)^37^ and Kunkle et al. (2019)^38^. We ran each analysis with and without the *APOE* region to isolate the contribution of this AD risk locus. Genetic pleiotropy between brain iron and AD was highly significant across all six subcortical regions in both AD datasets (**Fig. 4A, Supplementary Table 6**). The pleiotropic association ratio (PAR) measures the proportion of iron- and AD-associated variants that affect both traits, and varied across regions (**Supplementary Fig. 1, Supplementary Table 7**). In the Jansen et al. dataset with *APOE* excluded, PAR was highest for the globus pallidus (26.5%), followed by the substantia nigra (19.3%), putamen (17.9%), dentate nucleus (17.1%), red nucleus (16.2%) and caudate nucleus (12.9%). Including *APOE* lowered PAR across all regions. Because *APOE* has strong AD effect, it dominates the genome-wide mixture and compresses the polygenic estimate; we therefore report the *APOE*-excluded results as primary. The same pattern was observed in the Kunkle et al. dataset, where including *APOE* again attenuated all estimates. Because pleiotropy remained significant and PAR remained substantial (12.9-26.5%) after removing *APOE*, this shared signal is not driven by the *APOE* locus. Taken together, a substantial proportion of variants associated with brain iron and AD have shared effects on both traits, beyond APOE.

**Figure 4.**
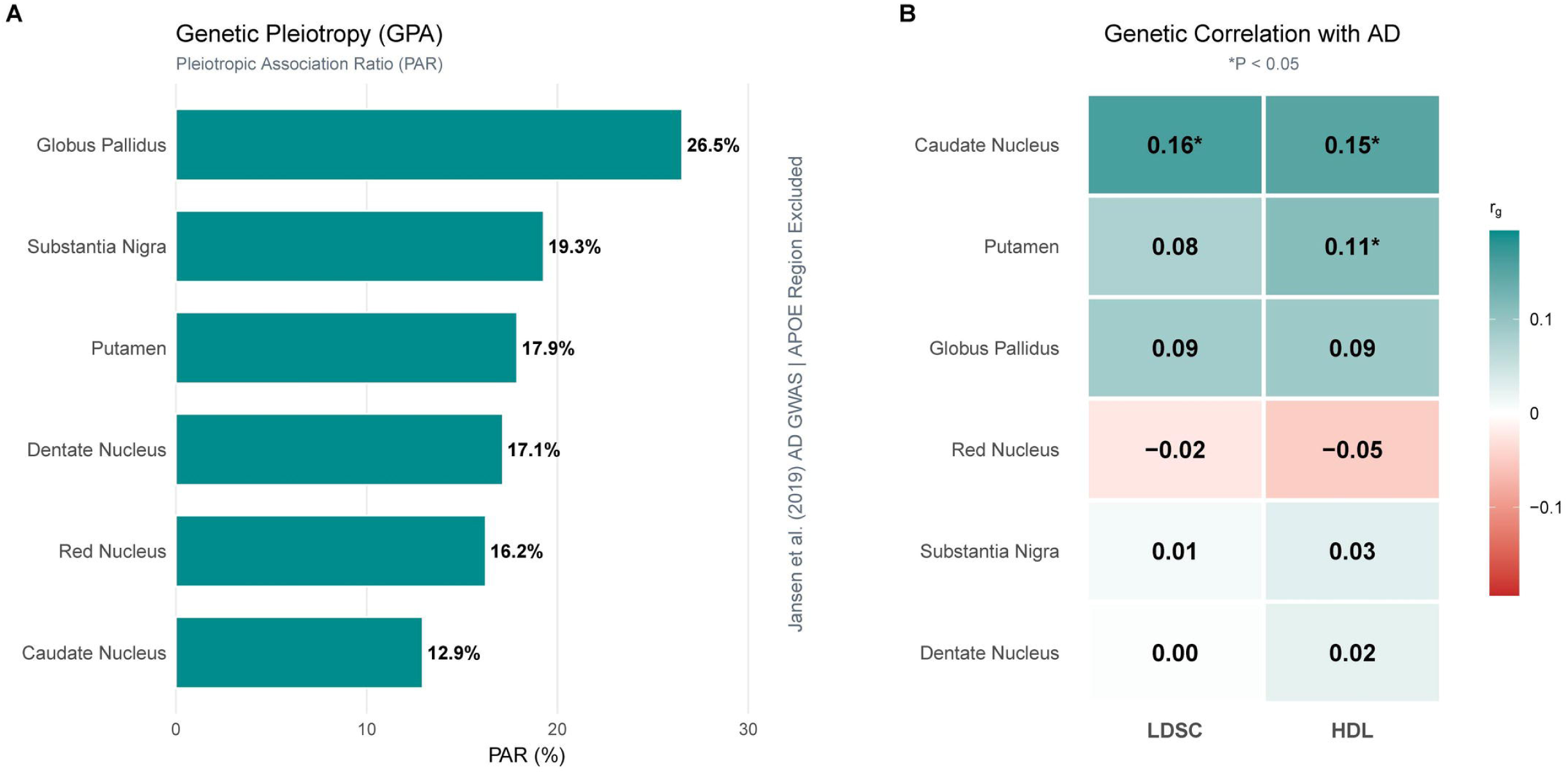
Genetic overlap and genetic correlation with Alzheimer’s Disease. (**A**) Heatmap shows pleiotropic association ratio (PAR) from genetic pleiotropy analysis (GPA). (**B**) Heatmap shows LDSC and HDL based genetic correlations of brain iron and Alzheimer’s disease.

Moving beyond genetic overlap, we performed genetic correlation analyses using LDSC and HDL methods. Although genetic overlap was substantial across all regions, genetic correlations were weaker and dataset specific. Using the Jansen et al. AD GWAS, the caudate nucleus showed a positive genetic correlation of brain iron with AD (*APOE* included: r = 0.159, SE = 0.056, *P* = 0.004; *APOE* excluded: r = 0.151, SE = 0.053, *P* = 0.005; **Fig. 4B, Supplementary Fig. 2**). The putamen was also positive (*APOE* included: r = 0.116, SE = 0.055, *P* = 0.034; *APOE* excluded: r = 0.111, SE = 0.053, *P* = 0.037), and the globus pallidus was nominally positive but non-significant (r ≈ 0.09, *P* ≈ 0.10). None survived Bonferroni correction, and no region reached significance in the clinically diagnosed dataset **(Supplementary Fig. 2, Supplementary Table 7)**.

### Shared genetic liability between brain iron and AD

To further characterise the genetic overlap, we modelled shared genetic liability using GenomicSEM. We fitted a common latent factor model incorporating all six QSM-derived regional iron measures, representing a global brain iron latent factor (**Supplementary Fig. 3**). The model showed acceptable fit (CFI = 0.937, SRMR = 0.066) with factor loadings ranging from 0.66 to 0.85. The global brain iron latent factor showed no significant genetic correlation with AD (*r* = 0.071, *P* = 0.20). This led us to hypothesise that brain iron accumulation in certain regions may have a shared liability, as previous studies have shown elevated levels in caudate, putamen, and globus pallidus of AD patients^43,44^. We therefore partitioned regions into two circuit-specific factors based on established neuroanatomical distinctions (**Supplementary Fig. 3**). The cognition relevant latent circuit factor comprised basal ganglia structures including the caudate nucleus, putamen and globus pallidus. This model showed excellent fit (CFI = 0.99, SRMR = 0.022) and sufficient factor loadings. The cognitive relevant latent circuit factor showed a significant positive genetic correlation with AD (*r* = 0.13, *P* = 0.019). We built a motor function relevant circuit latent factor comprised of substantia nigra, red nucleus and dentate nucleus (**Supplementary Fig. 3**). This model showed perfect fit (CFI = 1.0, SRMR = 0.006), however, no significant genetic correlation was observed with AD.

Next, we explicitly modelled shared genetic liability between brain iron in cognitive relevant regions and AD using a higher order latent factor model (**Fig. 5)**. This model captured the genetic variance shared between the basal ganglia iron latent factor and AD in a higher-order AD-iron latent factor. AD and basal ganglia iron factor loaded almost equally onto this shared factor. The model showed excellent fit (CFI = 0.99, SRMR = 0.022) with sufficient regional loading (putamen = 0.93, caudate = 0.80, globus pallidus = 0.73). The higher-order factor explained 14% of shared genetic variance (*P* = 0.019). A GWAS of AD-iron identified 27 independent genome-wide significant SNPs (r^2^ <0.1, **Supplementary Table 8**). We excluded genome-wide significant SNPs that showed a significant Q heterogeneity statistic and their partners in high LD, as their effects were more consistent with an independent pathway model than a common pathway model.

**Figure 5.**
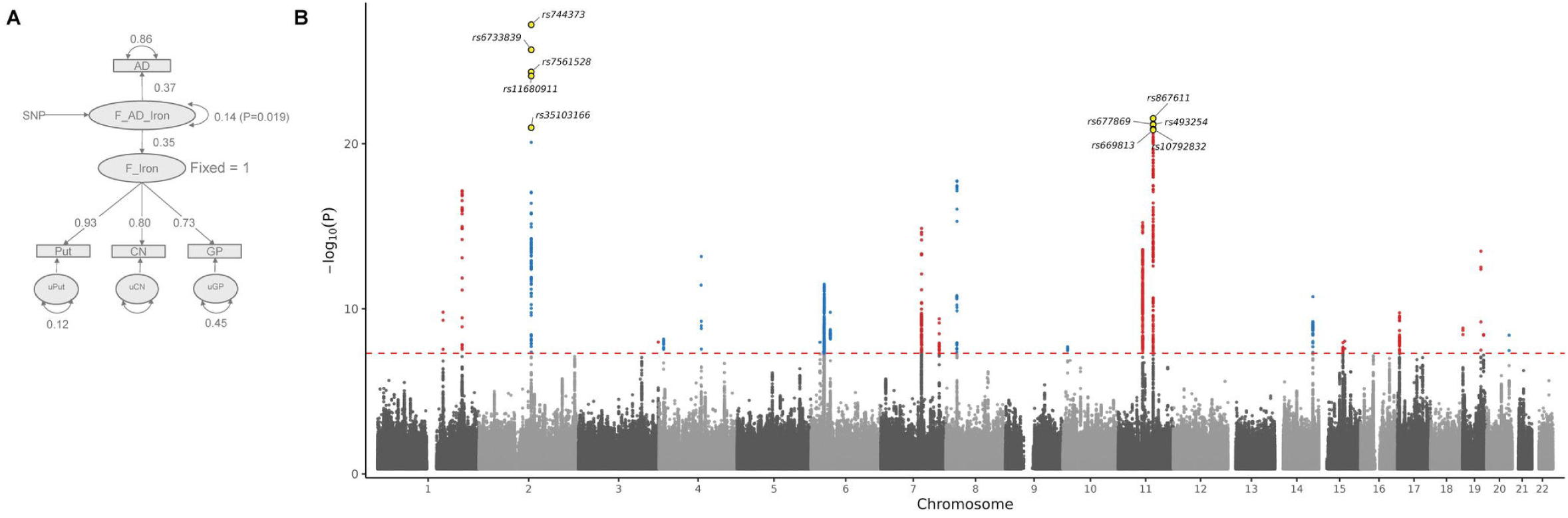
Shared liability latent factor. (**A**) Schematic diagram of the higher-order latent factor model. (**B**) Manhattan plot of the genome-wide association study (GWAS) for the shared AD-iron latent factor. The x-axis shows genomic position by chromosome. The y-axis shows statistical significance as −log₁₀(P). The red dashed line marks the genome-wide significance threshold (P < 5 × 10⁻⁸). Significant SNPs were retained after filtering for heterogeneity (Q_SNP). SNP rs4663105 (chr2; P = 1.26 × 10⁻⁴³) was excluded from the plot for display clarity. Yellow circles indicate the top 10 most significant SNPs.

### Gene prioritisation and functional enrichment of of the AD-iron latent factor

We next characterised the genes and overrepresented biological pathways underlying this shared factor. Multivariate GWAS of shared latent factor was used to identify genes using MAGMA and SMR. MAGMA identified 55 genes significantly associated with the AD-iron latent factor GWAS (Bonferroni corrected *P* < 2.62 × 10⁻⁶; **Supplementary Table 9**). SMR analysis prioritised 19 unique genes (Bonferroni corrected *P*_SMR_ < 3.55 × 10⁻⁶, *P*_HEIDI > 0.01; **Supplementary Table 10**), of which 11 derived from blood eQTLs and 8 from brain cortex eQTLs. Six genes (*ACE*, *SLC24A4*, *KAT8*, *CD2AP*, *EPHA1*, and *CR1*) were identified by both MAGMA and SMR analyses (**Fig. 6A**).

**Figure 6.**
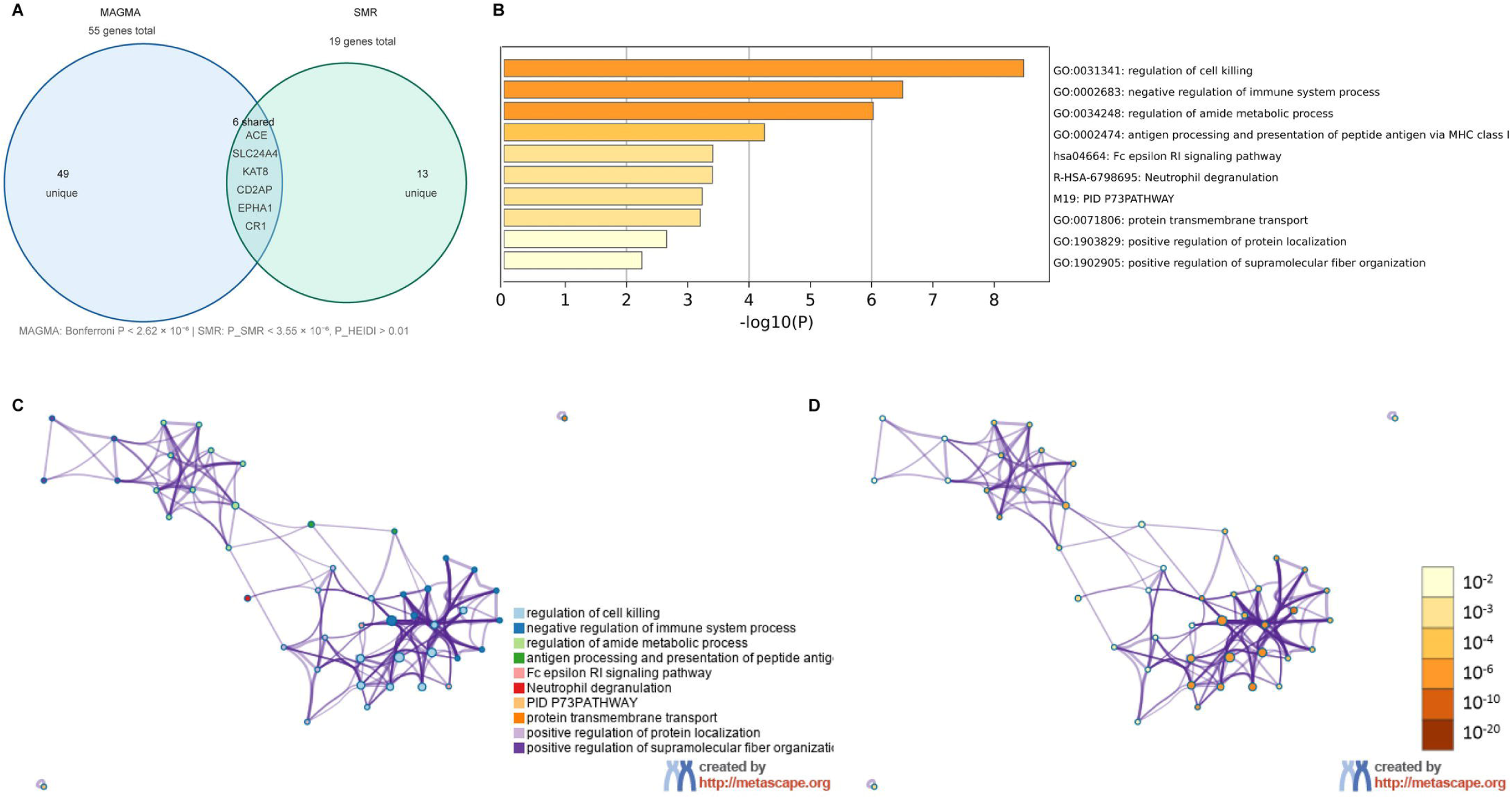
Gene-based and pathway enrichment results for the shared Alzheimer’s disease-iron latent factor. (**A**) Venn diagram of genes identified by MAGMA and summary data based Mendelian randomisation (SMR). ((**B**) Gene Ontology and pathway terms enriched among the latent factor genes. (C-D) Enrichment network of the same terms coloured by cluster (C) and by significance (D).

Using gene-level statistics, we next performed competitive gene-set analysis in MAGMA across all 17,009 MSigDB gene sets. MAGMA enrichment analysis showed seven gene sets associated with shared AD-iron latent factor (Bonferroni correction *P* < 2.94 × 10⁻⁶, **Supplementary Table 11A**), including complement and coagulation cascades (53 genes, β = 0.71, SE = 0.13, *P* = 1.1 × 10⁻⁸), immune complex clearance (5 genes, *P* = 1.4 × 10⁻⁸), neurofibrillary tangle (5 genes, *P* = 3.0 × 10⁻⁸), negative regulation of amyloid precursor protein catabolic process (18 genes, *P* = 4.0 × 10⁻⁸), negative regulation of metalloendopeptidase activity (4 genes, *P* = 2.6 × 10⁻⁷), microglial cell proliferation (8 genes, *P* = 1.5 × 10⁻⁶) and macrophage activation involved in immune response (19 genes, *P* = 2.9 × 10⁻⁶). Given the established role of iron-dependent lipid peroxidation in neurodegeneration, we examined whether the shared factor showed evidence of enrichment for ferroptosis-related biology within the same analysis. No gene set relating to ferroptosis, oxidative stress-induced cell death, lipid peroxidation, glutathione metabolism or iron handling was enriched (42 sets; **Supplementary Table 11B**). The ferroptosis pathway ranked 8,970 of 17,009 sets (58 genes, β = 0.007, standardised β = 0.0004, *P* = 0.47), and regulation of oxidative stress-induced neuron death was similarly null (*P* = 0.49).

Pathway and biological process enrichment analysis of genes underlying the AD-iron latent factor GWAS revealed consistent enrichment for immune-related biological processes across (**Fig. 6-7; Supplementary Table 12**). The strongest was regulation of cell killing (GO:0031341), followed by regulation of leukocyte-mediated immunity (GO:0002703) and a cluster of related terms at q = 9.7 × 10⁻⁴ comprising regulation of immune effector process, regulation of lymphocyte-mediated immunity, negative regulation of immune system process, immune response-regulating cell surface receptor signalling, and antigen processing and presentation of peptide antigen. Complement-related processes were also enriched (regulation of complement-dependent cytotoxicity, negative regulation of complement activation), as was antigen presentation via MHC class I (GO:0002474). Consistent with MAGMA based enrichment findings, over-representation analysis of the prioritised gene set identified immune-related biological processes (**Fig. 6-7; Supplementary Table 12**).

**Figure 7.**
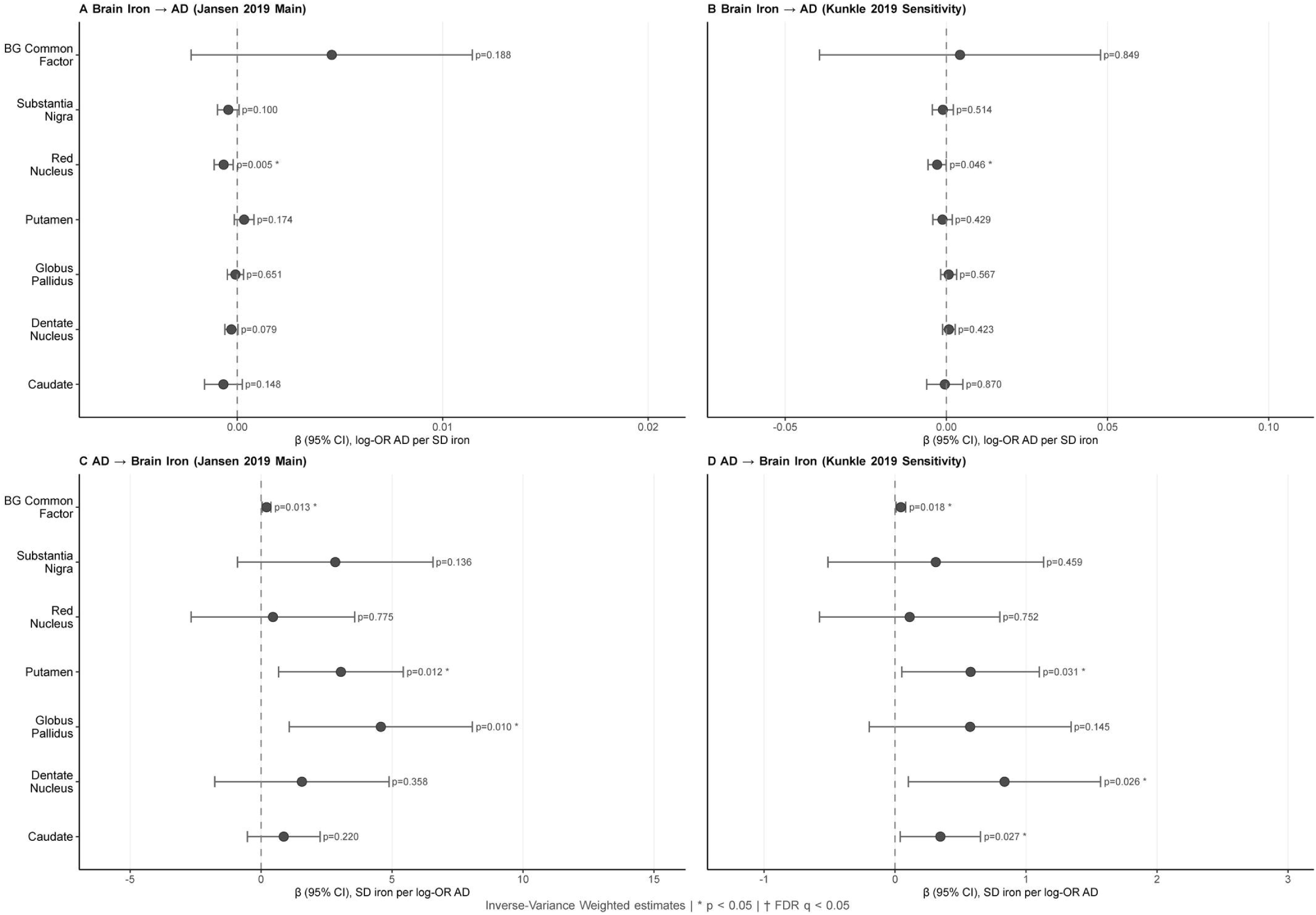
Bidirectional Mendelian randomisation between brain iron and Alzheimer’s disease. Inverse-variance weighted estimates with 95% confidence intervals. (**A**) Brain iron to Alzheimer’s disease, main analysis. (**B**) Brain iron to Alzheimer’s disease, sensitivity analysis in the overlap-free dataset. (**C**) Alzheimer’s disease to brain iron, main analysis. (**D**) Alzheimer’s disease to brain iron, sensitivity analysis. Estimates are the log-odds of Alzheimer’s disease per standard deviation of iron in **A** and **B**, and the standard deviation of iron per log-odds of Alzheimer’s disease in **C** and **D**. The dashed vertical line marks the null. Asterisks denote P < 0.05; no estimate survived correction for the false discovery rate. The APOE region was excluded throughout. BG = basal ganglia; IVW = inverse-variance weighted.

Protein-protein interaction network analysis reinforced the immune signal, with the primary interaction module enriched for regulation of cell killing (log₁₀P = −11.7), regulation of lymphocyte-mediated immunity (log₁₀P = −9.9), and regulation of immune effector process (log₁₀P = −9.4; **Supplementary Fig. 4A**). Cell-type signature analysis further indicated that the AD-iron gene set was most strongly expressed in myeloid cell populations, including fetal myeloid cells across multiple tissues (log₁₀P = −12.0 to −7.0) and fetal brain immune cells and microglia (log₁₀P = −6.2 to −5.9; **Supplementary Fig. 4B**). Tissue-specific expression analysis confirmed predominant enrichment in bone marrow and spleen (log₁₀P up to −3.0; **Supplementary Fig. 4C**), consistent with the myeloid cell-type signature. Disease association enrichment analysis using DisGeNET showed that AD phenotypes dominated the top associations of the AD-iron gene set (log₁₀P = −16.0 to −9.0) (**Supplementary Fig. 4D**), consistent with the known disease architecture targeted by the shared latent factor.

### Causal pathways linking AD and brain iron

We investigated putative causal relationships between regional brain iron and AD using bidirectional two-sample Mendelian randomisation, with the *APOE* region excluded throughout. Genetic instruments were drawn from our QSM GWAS for the iron-to-AD direction and from two AD GWAS (Jansen et al. 2019; Kunkle et al. 2019) for the reverse direction. In the iron-to-AD direction, no region showed a causal effect on AD that survived multiple testing correction (**Fig. 7, Supplementary Table 13-17**). The strongest nominal signal-an inverse association between red nucleus iron and AD risk- was directionally consistent across IVW, MR-Egger, and Weighted Median estimators and strengthened after MRlap correction (β = -0.014, *P* = 0.012) but did not survive FDR (q = 0.093). In the AD-to-iron direction, nominal effects of AD liability on putamen and globus pallidus iron were substantially attenuated after MRlap correction (all p > 0.10), indicating inflation by sample overlap rather than true causal effects (**Fig. 7, Supplementary Fig. 5, Supplementary Table 13-17**). A residual signal for putamen using the overlap-free Kunkle instruments did not survive FDR correction either. Instruments were strong in both directions (all F > 10; mean F = 54-77 forward, ∼48 reverse; **Supplementary Table 17**). In the forward direction (iron → AD), the genome-wide significant instruments explained a substantial share of variance in regional iron: 20.4% (caudate), 26.3% (dentate nucleus), 22.6% (putamen), 16.7% (globus pallidus), 13.7% (red nucleus) and 7.5% (substantia nigra) in the Jansen dataset, with near-identical values for Kunkle (**Supplementary Table 17**). These are comparable to the full genome-wide polygenic scores (approximately ∼25%; **Supplementary Table 3**). Despite this power, MRlap estimates for the iron-to-disease direction were null with narrow confidence intervals across all regions (**Supplementary Table 17**), excluding all but small causal effects. In the reverse direction (AD → iron), *APOE*-excluded AD instruments explained only 0.35% (Jansen) and 1.28% (Kunkle) of variance in AD, and the reverse estimates had wide confidence intervals; reverse causal effects could therefore not be reliably estimated. Between-instrument heterogeneity was low-to-moderate in the forward direction (I² = 11–55%; **Supplementary Table 17**), consistent with pleiotropy, and MR-Egger and weighted-median estimates were concordant with IVW (**Supplementary Table 13**). Taken together, these analyses do not provide robust evidence for a causal relationship in either direction at current sample sizes, suggesting that basal ganglia iron accumulation may partly reflect downstream consequences of AD-related neurodegeneration.

## Discussion

Brain iron accumulation has long been observed in AD, but whether this association reflects a shared genetic basis has remained unresolved. This study provides one of the largest genome-wide association analyses of QSM brain iron, identifying 116 independent loci. Using genetic overlap analysis, genetic correlation and genomic SEM, we show that brain iron and AD share a common genetic factor concentrated within the basal ganglia-cognitive circuit. Pathway analyses of this shared factor implicate neuroinflammation and complement activation as candidate mechanistic links between iron accumulation and AD risk.

Of the 116 independent loci associated with subcortical brain iron, 41 are novel and 75 replicated against earlier analyses of quantitative susceptibility mapping^23,24^. This replication rate is consistent with observations from other neuroimaging GWAS, where differences in sample size, ancestry composition and QSM processing protocols typically drive partial overlap. Earlier studies established that subcortical iron measures are substantially heritable and identified an initial set of associated loci^23,24^. The present analysis expands this catalogue and demonstrates its predictive validity. A polygenic risk score derived from our GWAS predicted brain iron levels in the independent PISA cohort, providing out-of-sample evidence that these signals are robust and generalisable. PRS explained between 8.9% and 25.4% of variance in QSM-derived iron levels, which is consistent with reported heritability for neuroimaging endophenotypes^23^. These findings suggest PRS of QSM-derived iron measures may be used for genetic risk stratification of healthy individuals, potentially aiding early intervention strategies related to iron deposition and neurodegeneration prevention.

Beyond expanding the genetic architecture of brain iron, we identified a novel point of genetic convergence with AD. Genomic SEM revealed that iron deposition across the caudate nucleus, putamen and globus pallidus loads onto a single latent factor, and that this factor shows a significant positive genetic correlation with AD. Notably, a global iron factor incorporating all six subcortical regions showed no significant genetic correlation with AD. The association is therefore not a property of brain iron in general, but of iron within a specific basal ganglion-cognitive circuit. This circuit-specific signal aligns with observational QSM findings of elevated basal ganglia iron in AD patients, where the magnitude of accumulation tracks with cognitive decline^45,46^. Our genetic evidence complements these observations by indicating that the link is not purely a downstream consequence of disease but reflects shared genetic liability.

To identify the molecular basis of this shared liability, we performed a multivariate GWAS of the higher order latent factor and identified 27 independent loci. Competitive gene-set analysis across all gene-level statistics implicated complement regulation, endocytic handling of amyloid precursor protein and microglial survival and proliferation, suggesting that the shared liability potentially acts through complement-mediated opsonisation and phagocytic signalling. Over-representation analysis of the prioritised genes and cell-type signature analysis converged on the same conclusion, pointing consistently to myeloid populations including fetal myeloid cells, fetal brain immune cells and microglia. Together these results indicate that the genetic overlap between basal ganglia iron and AD is concentrated in genes governing neuroimmune function. While lipid peroxidation and iron accumulation, signatures of ferroptosis, have been documented in post-mortem AD brain tissues^47,48^, and iron dysregulation in the presence of neuroinflammation has been shown to accelerate ferroptosis, a form of regulated cell death implicated in synaptic loss and neurodegeneration^49,50^, we did not, however, observe enrichment of ferroptosis-related gene sets in the shared factor. We observed enrichment of complement cascade (*P* = 1.1 × 10⁻⁸). This does not argue against a role for ferroptosis in iron-related neurodegeneration but indicates that the common variation shared between basal ganglia iron and AD is concentrated in neuroimmune rather than ferroptotic genes. One interpretation is that genetically determined microglial function modulates iron handling within the basal ganglia, and that ferroptotic injury arises downstream of iron loading in those cells rather than being encoded in the shared genetic architecture itself.

Bidirectional MR did not detect causal effects between brain iron and AD in either direction after multiple-testing correction and MRlap adjustment for sample overlap. In the forward direction this is an informative null rather than a power limitation: instruments were strong (mean F 54-77) and explained 7-26% of variance in regional iron, comparable to the full genome-wide PRS, because iron heritability is concentrated in a few large-effect loci. Against this well-powered background, the tight null confidence intervals exclude all but small causal effects of iron on AD. The reverse direction was underpowered instruments with *APOE* excluded explained less than 1.3% of variance. This pattern- a well-powered forward null alongside strong, direction-mixed pleiotropy and a shared genetic factor - indicates that the genetic overlap between basal ganglia iron and AD most plausibly reflects shared genetic architecture rather than a direct causal effect in either direction.

Our analysis covered six iron-rich subcortical nuclei and did not include the cortex or hippocampus, the regions where Alzheimer disease pathology is most pronounced. This was a deliberate choice driven by measurement reliability. QSM gives dependable susceptibility estimates in structures with high iron content, but in lower-iron and geometrically complex regions such as the hippocampus, and in areas prone to signal dropout near air–tissue boundaries, the estimates are far noisier. The same pattern appears in the UK Biobank QSM reference data of Wang and colleagues, where susceptibility in the putamen and substantia nigra was among the most heritable of all brain measures while regions such as the accumbens and amygdala were among the least. We therefore restricted the analysis to nuclei where the iron signal is strong and reproducible. A consequence is that our conclusions describe the genetics of basal ganglia iron and its overlap with AD and cannot say whether iron in cortical or hippocampal regions carries a different genetic relationship with AD. Measuring those regions reliably, and extending this analysis to them, is an important next step.

We excluded *APOE* from the pleiotropy, correlation and Mendelian randomisation analyses. This was a methodological decision: *APOE* has an extreme effect on AD that dominates genome-wide polygenic estimates, and in Mendelian randomisation it acts as a strongly pleiotropic, invalid instrument. Removing it let us test whether iron and AD share genetic signal beyond this single locus. This should not be read as treating APOE as unimportant to the iron–AD link. APOE genotype is itself a regulator of brain iron: CSF ferritin, a marker of brain iron burden, is regulated by APOE and predicts AD outcomes^6^. APOE is therefore likely one of the biologically meaningful connections between iron and AD, which we set aside specifically to reveal the additional sharing that remains without it. Consistent with a genuine APOE contribution, the pleiotropy signal stayed significant when the region was retained, and the reduced power of our reverse MR partly reflects its removal, since APOE is the strongest AD instrument and one with documented iron effects.

Overall, this study provides a comprehensive genetic characterisation of subcortical brain iron and identifies the basal ganglia-cognitive circuit as a key locus of shared genetic susceptibility with AD. Iron accumulation in the basal ganglia-cognitive circuit shows significant shared genetic architecture with AD, with the overlap concentrated in complement regulation, endocytic processing and microglial genes. Gene-set analysis found no evidence that this shared liability is encoded in ferroptosis machinery, indicating that the overlap we identify is a distinct neuroimmune axis. The PRS derived here predicts brain iron levels in an independent cohort, supporting the robustness of the identified genetic architecture. Although MR did not support a direct causal effect in either direction, the well-powered forward null together with the shared genetic factor suggests that the observed relationship is driven by shared genetic architecture rather than causation. Together, these findings position basal ganglia iron as a genetically regulated marker of AD biology and provide new directions for mechanism-targeted prevention research.

## Data availability

All generated datasets are provided within the manuscript and associated supplementary tables. Individual-level UK Biobank data (linked Applications 27483 and 25331) cannot be redistributed by the authors and are available to bona fide researchers via application to the UK Biobank (https://www.ukbiobank.ac.uk).

## Acknowledgements

This research has been conducted using the UK Biobank Resource under linked Applications 27483 and 25331. A large language model (Claude, Anthropic) was used to check the manuscript against the journal’s formatting and style requirements, to assist with language editing and to draw the schematic thumbnail figure. It was not used to design the study, to generate, process or analyse data, or to interpret the results.

## Supplementary material

Supplementary material is available at online.

