## Supplementary Figures for "Shared genetic architecture between brain iron deposition and Alzheimer’s disease"

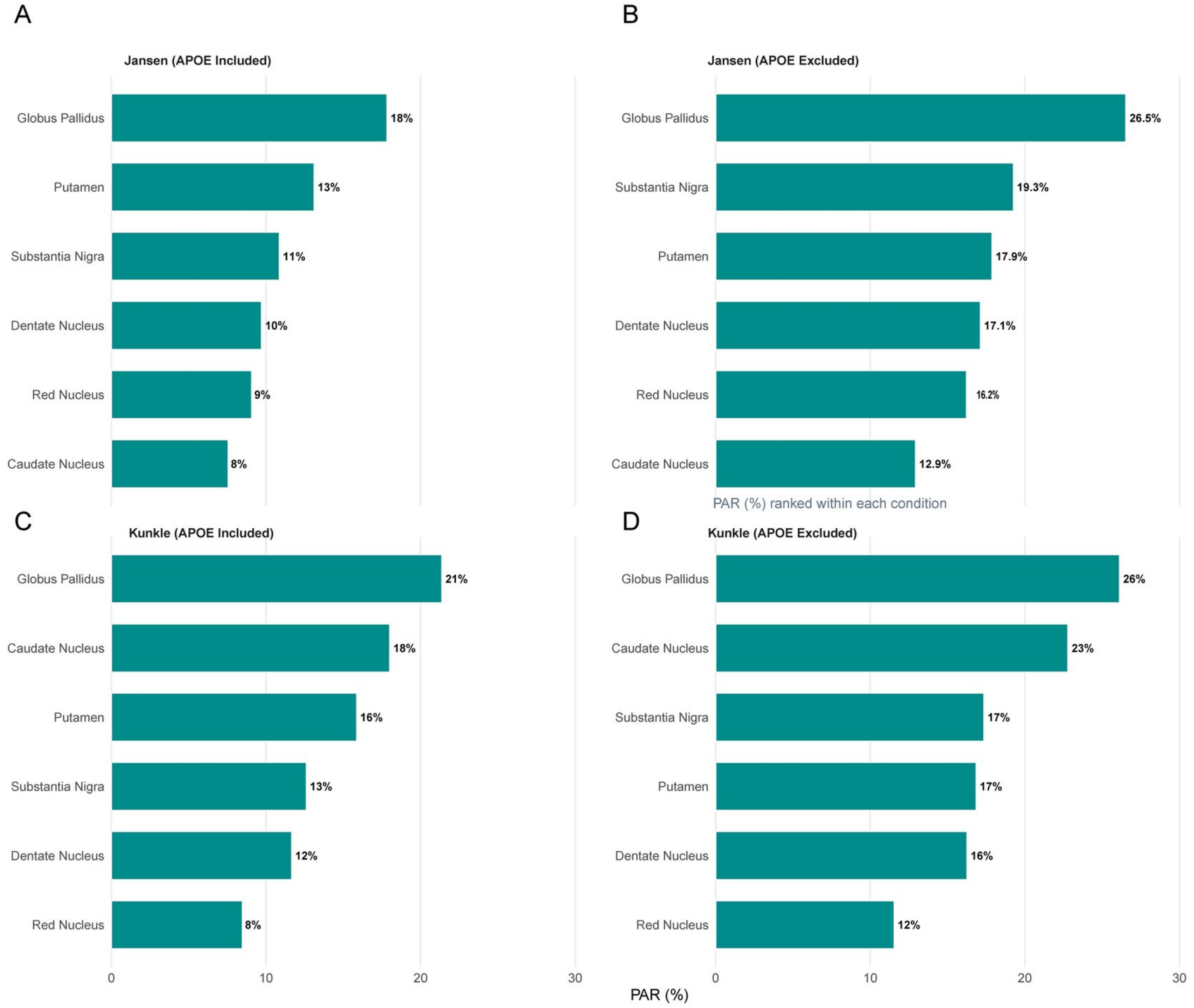


**Fig. S1**. Genetic pleiotropy between brain iron and Alzheimer's disease across APOE conditions. Bar charts show the pleiotropic association ratio (PAR, %) between regional brain iron levels (QSM) and Alzheimer's disease (AD), estimated using the genetic pleiotropy analysis (GPA) framework. Regions are ranked by PAR within each condition. Results are shown for two independent AD GWAS datasets - Jansen et al. (**A, B**) and Kunkle et al. (**C, D**) - each analysed with APOE included (**A, C**) and excluded (**B, D**). PAR represents the proportion of brain iron-associated SNPs that also show enrichment for AD genetic signal. CN = caudate nucleus; GP = globus pallidus; DN = dentate nucleus; SN = substantia nigra; RN = red nucleus.

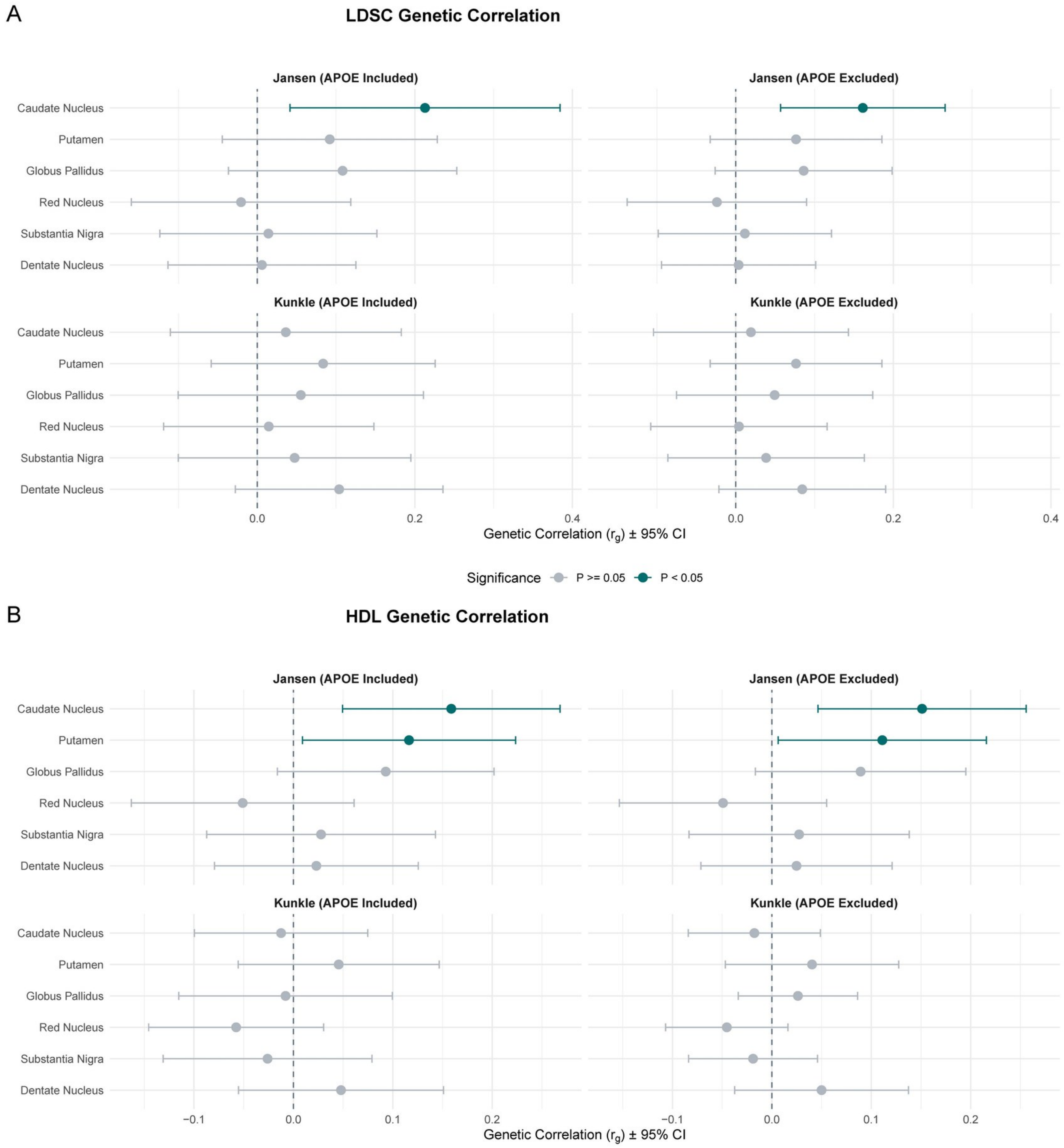


**Fig. S2**. Sensitivity analysis of genetic correlations between brain iron and Alzheimer's disease using LDSC and HDL. Forest plots show genetic correlations between six brain iron regions (QSM) and Alzheimer's disease (AD), estimated using LD score regression (LDSC; **A**) and High-Definition Likelihood (HDL; **B**). Results are shown for two independent AD GWAS datasets - Jansen et al. and Kunkle et al. - each analysed with APOE included (left panels) and excluded (right panels). Teal circles indicate statistically significant correlations ($P < 0.05$); grey circles indicate non-significant results.

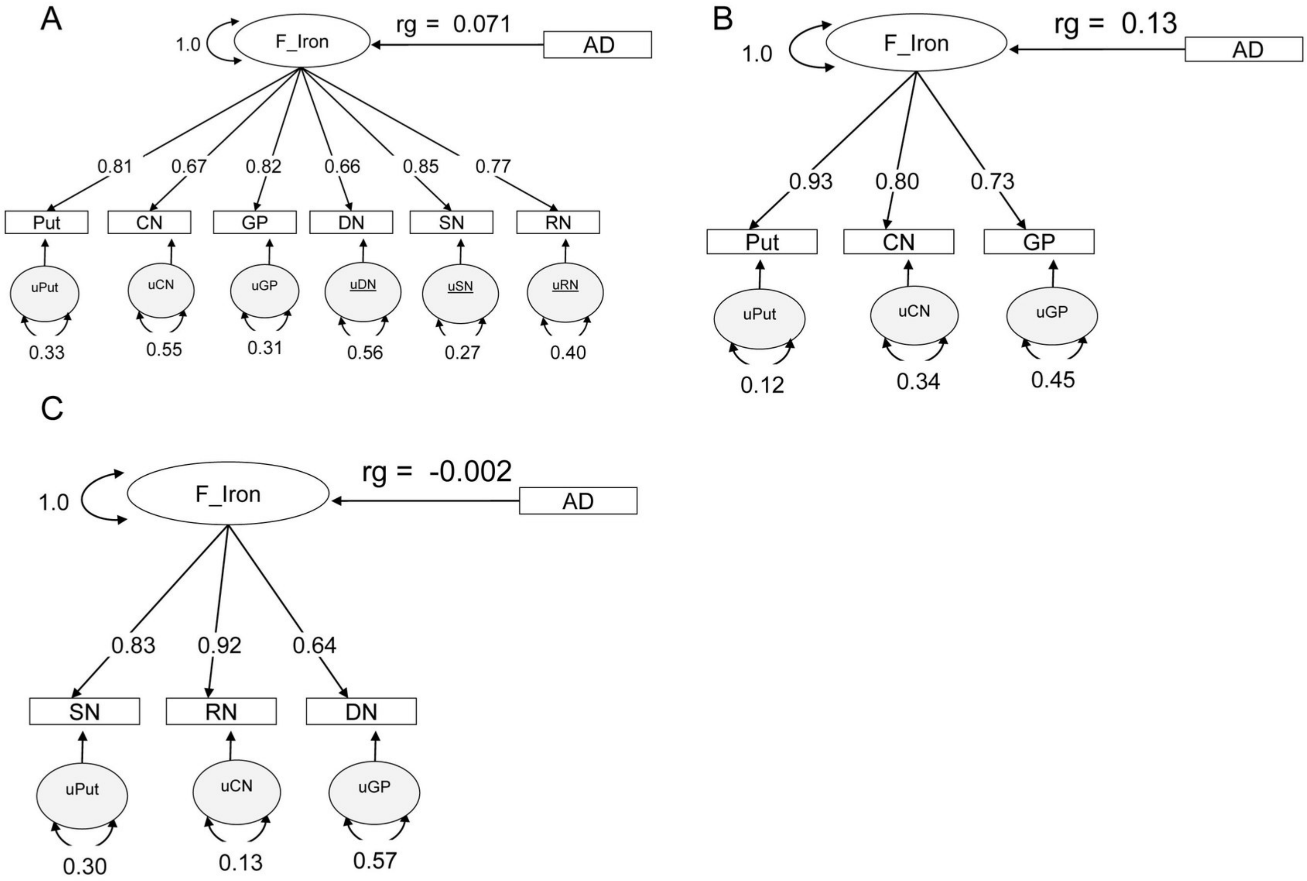


**Fig. S3. Genomic SEM latent factor models for brain iron and Alzheimer's disease genetic correlation.** Path diagrams from Genomic structural equation modelling (Genomic SEM) show three alternative latent factor structures for shared genetic architecture between brain iron and Alzheimer's disease (AD). (**A**) Global brain iron factor (F_Iron) constructed from all six QSM regions (putamen, caudate nucleus, globus pallidus, dentate nucleus, substantia nigra, red nucleus). Factor loadings ranged from 0.66 to 0.85. (**B**) Cognitive circuit factor comprising basal ganglia regions only (putamen, caudate nucleus, globus pallidus; loadings: 0.93, 0.80, 0.73). (**C**) Motor circuit factor comprising cerebellar and brainstem regions (substantia nigra, red nucleus, dentate nucleus; loadings: 0.83, 0.92, 0.64). Numbers on paths represent standardised loadings (rectangles to factor) and residual variances (ovals). AD = Alzheimer's disease; F_Iron = common brain iron latent factor; Put = putamen; CN = caudate nucleus; GP = globus pallidus; DN = dentate nucleus; SN = substantia nigra; RN = red nucleus.

**A**

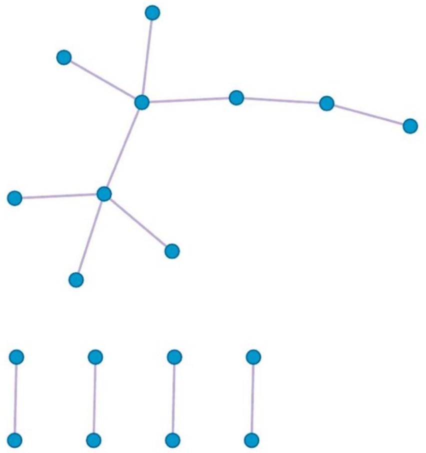

**Protein-protein interaction (PPI) network.** Physical interactions among genes associated with the AD-iron latent factor identified using STRING (physical score > 0.132) and BioGrid. Densely connected modules were identified using the MCODE algorithm. The primary module was enriched for regulation of cell killing ($\log_{10}P = -11.7$), regulation of lymphocyte-mediated immunity ($\log_{10}P = -9.9$), and regulation of immune effector process ($\log_{10}P = -9.4$).

**B**

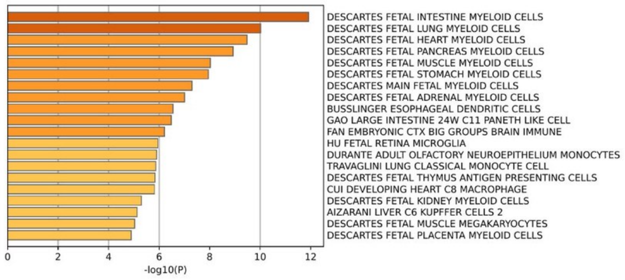


**Cell-type signature enrichment.** Bar chart showing enrichment of the AD-iron gene set against reference cell-type expression signatures. Consistent enrichment was observed across fetal myeloid cell populations ($\log_{10}P = -12.0$ to $-7.0$) and fetal brain immune cells including microglia ($\log_{10}P = -6.2$ to $-5.9$), confirming the neuroinflammatory and myeloid character of the shared AD-iron genetic signal.

**C**

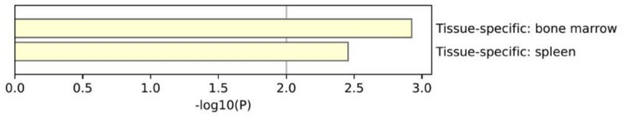


**Tissue-specific expression (PaGenBase).** Bar chart showing tissue specificity of the AD-iron gene set. Enrichment was restricted to bone marrow and spleen ($\log_{10}P$ up to $-3.0$), consistent with the predominantly immune and myeloid cell-type signature observed in panel B.

**D**

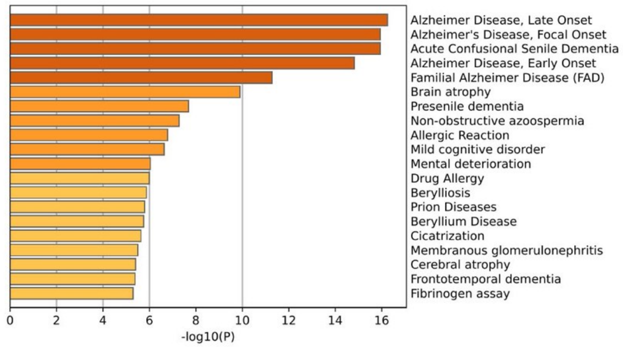


**Disease association enrichment (DisGeNET).** Bar chart showing enrichment of the AD-iron gene set for curated disease-gene associations. Alzheimer disease phenotypes dominate the top associations ($\log_{10}P = -16.0$ to $-9.0$), encompassing late-onset, early-onset, and familial AD, presenile dementia, and brain atrophy, consistent with the known disease architecture targeted by the shared latent factor.

**Fig. S4. Functional enrichment of the AD–brain iron shared genetic signal.** Results from Metascape enrichment analyses using 64 genes associated with the AD–iron shared latent factor. (**A**) Protein-protein interaction network constructed using STRING. Densely connected modules were identified using the MCODE algorithm. (**B**) Cell-type signature enrichment against reference expression profiles (DESCARTES and other datasets). (**C**) Tissue-specific expression (PaGenBase). (**D**) Disease association enrichment (DisGeNET). Bar colour indicates enrichment strength (darker = more significant).

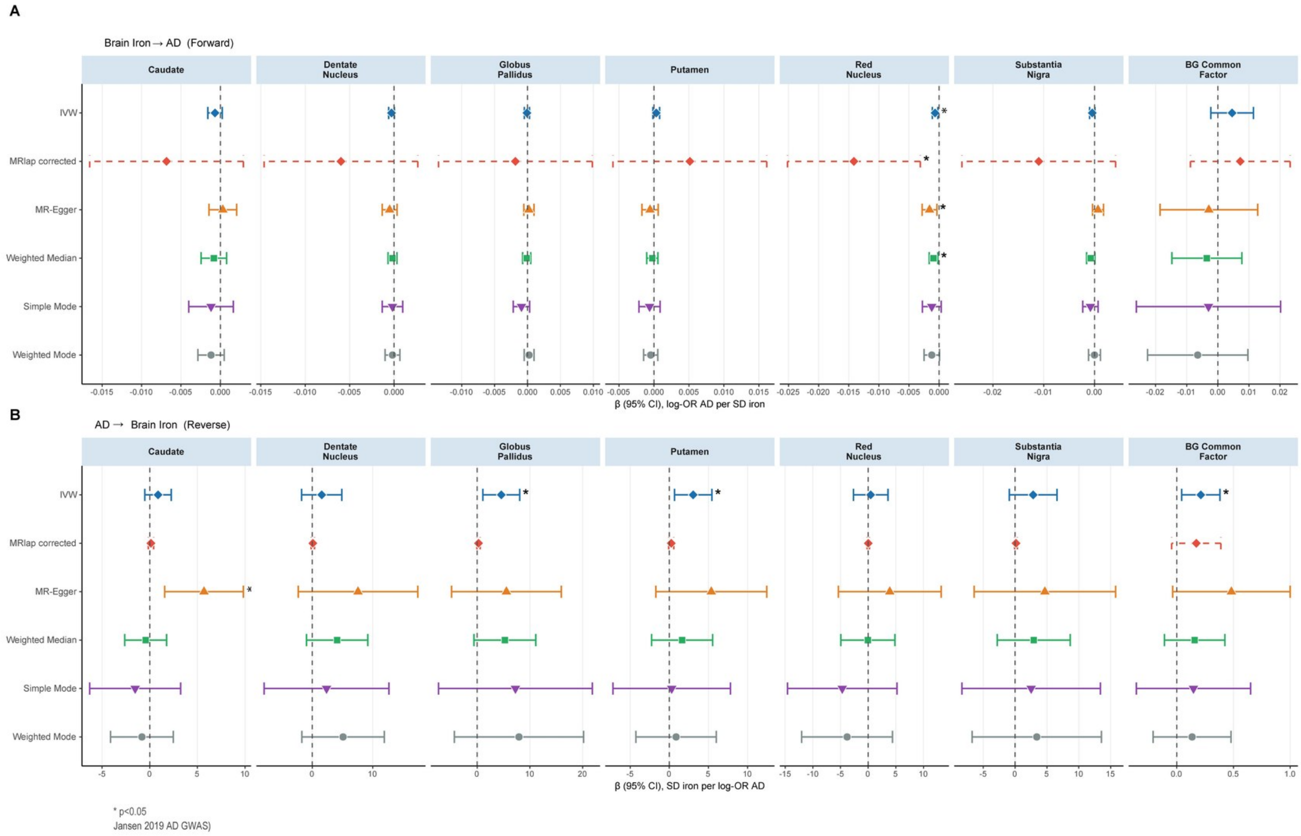


**Fig. S5. Mendelian randomisation sensitivity analyses for the bidirectional relationship between brain iron and Alzheimer's disease.** Forest plots show effect estimates (β ± 95% CI) from multiple MR methods for the bidirectional relationship between regional brain iron (QSM) and Alzheimer's disease (AD; Jansen et al. 2019). (A) Forward direction (brain iron → AD). (B) Reverse direction (AD → brain iron). Results are shown for six brain regions and the basal ganglia common factor. Six MR methods are displayed for each region: inverse-variance weighted (IVW), MRlap-corrected IVW, MR-Egger, weighted median, simple mode, and weighted mode. Asterisks (*) denote statistically significant results ($P < 0.05$).
