## Supplementary Tables for "Shared genetic architecture between brain iron deposition and Alzheimer’s disease"

Rahman *et al.* Shared genetic architecture between brain iron deposition and Alzheimer's disease. Abbreviations are defined in a footnote beneath every table. The same tables are supplied as a spreadsheet workbook for readers who wish to sort or filter them.

| Table | Title | Rows |
|---|---|---|
| S1 | Regional genome-wide significant loci | 257 |
| S2 | Independent loci after cross-regional merging | 116 |
| S3 | Polygenic risk score validation in PISA | 36 |
| S4 | MAGMA genes, regional brain iron | 1126 |
| S5 | SMR genes, regional brain iron | 267 |
| S6 | Genetic pleiotropy with Alzheimer's disease | 24 |
| S7 | Pleiotropic association ratios and genetic correlations | 48 |
| S8 | Multivariate GWAS loci, shared factor | 27 |
| S9 | MAGMA genes, shared factor | 56 |
| S10 | SMR genes, shared factor | 20 |
| S11 | Competitive gene-set analysis | 92 |
| S12 | Pathway and process enrichment | 50 |
| S13 | IVW Mendelian randomisation, main analysis | 14 |
| S14 | IVW Mendelian randomisation, Kunkle sensitivity | 16 |
| S15 | All MR estimators, forward direction | 42 |
| S16 | All MR estimators, reverse direction | 43 |
| S17 | Instrument strength and heterogeneity | 28 |

### Supplementary Table 1. Genome-wide significant loci from the regional genome-wide association analyses of quantitative susceptibility mapping-derived brain iron across six subcortical regions (P < 5 x 10-8).

| Region | Locus ID | Unique variant ID | Chromosome | Start (bp) | End (bp) | Top SNP | Top P | Total SNPs | Genome-wide significant SNPs | Independent significant SNPs | Lead SNPs | Independent SNPs |
|---|---|---|---|---|---|---|---|---|---|---|---|---|
| Caudate nucleus | 1 | 1:205248032:A:G | 1 | 205012106 | 205297880 | rs1172134 | 1.7e-08 | 76 | 55 | 1 | 1 | rs1172134 |
| Caudate nucleus | 2 | 2:120155348:A:G | 2 | 120087644 | 120208802 | rs72833227 | 2.1e-10 | 95 | 70 | 3 | 2 | rs72833227;rs6722204;rs2579633 |
| Caudate nucleus | 3 | 2:190382493:A:G | 2 | 190374283 | 190423785 | rs12616162 | 1.6e-08 | 20 | 16 | 1 | 1 | rs12616162 |
| Caudate nucleus | 4 | 3:107464170:A:G | 3 | 107464170 | 107470613 | rs11710737 | 2.6e-09 | 2 | 2 | 1 | 1 | rs11710737 |
| Caudate nucleus | 5 | 3:133466374:A:G | 3 | 133282706 | 133571004 | rs4428180 | 7.5e-25 | 181 | 128 | 15 | 2 | rs4428180;rs1799852;rs6762415;rs1405022; rs6794370;rs9824452;rs77653936;rs6439425 ... |
| Caudate nucleus | 6 | 3:147032401:A:G | 3 | 147020326 | 147082740 | rs60852193 | 2.1e-10 | 40 | 29 | 3 | 1 | rs60852193;rs66507831;rs4681356 |
| Caudate nucleus | 7 | 3:195841644:C:T | 3 | 195776017 | 195844238 | rs7432894 | 2e-10 | 79 | 58 | 5 | 1 | rs7432894;rs9849934;rs41298089;rs6583290; rs12498103 |
| Caudate nucleus | 8 | 4:15737348:A:G | 4 | 15710330 | 15743332 | rs4698412 | 4.9e-08 | 11 | 11 | 1 | 1 | rs4698412 |
| Caudate nucleus | 9 | 4:103188709:C:T | 4 | 101920593 | 104163862 | rs13107325 | 7.3e-148 | 1076 | 805 | 69 | 17 | rs2063058;rs4698918;rs17030740;rs76062146; rs17249745;rs17199964;rs34592089;rs779 ... |
| Caudate nucleus | 10 | 4:159650783:A:G | 4 | 159621185 | 159650783 | rs58093123 | 2.4e-08 | 9 | 6 | 1 | 1 | rs58093123 |
| Caudate nucleus | 11 | 6:26093141:A:G | 6 | 25331455 | 28103409 | rs1800562 | 2.9e-23 | 748 | 547 | 31 | 8 | rs12196438;rs9467478;rs75580845;rs28393611; rs78151190;rs62393683;rs72838866;rs93 ... |
| Caudate nucleus | 12 | 6:126683594:A:G | 6 | 126659043 | 127080700 | rs9398803 | 4.3e-08 | 248 | 173 | 1 | 1 | rs9398803 |
| Caudate nucleus | 13 | 7:1563046:C:T | 7 | 1553373 | 1595839 | rs6463895 | 1.1e-11 | 129 | 112 | 1 | 1 | rs6463895 |
| Caudate nucleus | 14 | 7:72977249:C:T | 7 | 72376843 | 73134026 | rs34594435 | 5.1e-26 | 315 | 244 | 11 | 2 | rs34594435;rs57978469;rs799169;rs56377543; rs4717763;rs62465131;rs42238;rs1411361 ... |
| Caudate nucleus | 15 | 8:23417091:A:G | 8 | 23220509 | 23417918 | rs2004645 | 3.3e-15 | 178 | 137 | 10 | 2 | rs4260902;rs2048528;rs36104352;rs721183; rs201643157;rs11782783;rs28656888;rs7813 ... |
| Caudate nucleus | 16 | 8:27103024:A:G | 8 | 27098436 | 27119538 | rs17366947 | 7.7e-11 | 8 | 7 | 1 | 1 | rs17366947 |
| Caudate nucleus | 17 | 9:19210177:A:G | 9 | 19210177 | 19378074 | rs465423 | 5.4e-17 | 12 | 8 | 2 | 1 | rs465423;rs62560298 |
| Caudate nucleus | 18 | 9:32448436:A:G | 9 | 32347719 | 32579454 | rs10970982 | 6.6e-15 | 148 | 104 | 7 | 3 | rs1023087;rs10970982;rs13289402;rs75438929; rs7043497;rs4879586;rs4487862 |
| Caudate nucleus | 19 | 10:18221908:C:T | 10 | 17845995 | 18720109 | rs4348791 | 5.9e-125 | 911 | 640 | 115 | 31 | rs530434517;rs555045010;rs2436671; rs367834034;rs201843067;rs149410244;rs1926732 ... |
| Caudate nucleus | 20 | 10:116237760:C:T | 10 | 116228747 | 116275097 | rs11196758 | 1e-12 | 18 | 16 | 1 | 1 | rs11196758 |
| Caudate nucleus | 21 | 11:109936687:G:T | 11 | 109936687 | 109953618 | rs4027098 | 4.1e-08 | 15 | 11 | 1 | 1 | rs4027098 |
| Caudate nucleus | 22 | 12:51484646:C:T | 12 | 51326702 | 51581740 | rs7979705 | 2.1e-15 | 349 | 253 | 6 | 2 | rs11169664;rs7980874;rs7979705;rs224567; rs4768952;rs1112392 |
| Caudate nucleus | 23 | 12:113583863:A:C | 12 | 113536863 | 113752802 | rs146487929 | 2.7e-12 | 261 | 198 | 4 | 1 | rs146487929;rs61943591;rs77625177;rs73208705 |
| Caudate nucleus | 24 | 15:45388382:C:T | 15 | 45365790 | 45409271 | rs2554451 | 3.9e-09 | 35 | 29 | 2 | 1 | rs2554451;rs269859 |
| Caudate nucleus | 25 | 15:79237293:C:T | 15 | 78714008 | 79257278 | rs2289702 | 6.7e-16 | 297 | 229 | 8 | 2 | rs12912022;rs11636753;rs8038920;rs12916769; rs12910090;rs2289702;rs12901830;rs620 ... |
| Caudate nucleus | 26 | 16:79606520:C:T | 16 | 79586108 | 79614116 | rs434626 | 6.7e-18 | 52 | 40 | 8 | 1 | rs434626;rs425628;rs30388;rs250164;rs250151; rs250150;rs250148;rs821225 |
| Caudate nucleus | 27 | 17:40716235:C:T | 17 | 40287106 | 41459301 | rs668799 | 2.5e-41 | 994 | 729 | 22 | 4 | rs76899038;rs601999;rs668799;rs1474040; rs12937081;rs76189032;rs1554227;rs7863005 ... |
| Caudate nucleus | 28 | 17:42989063:C:T | 17 | 42901466 | 43056905 | rs1126642 | 1e-09 | 12 | 10 | 2 | 1 | rs1126642;rs77136668 |
| Caudate nucleus | 29 | 17:57609588:C:T | 17 | 57557750 | 58052162 | rs8073905 | 1.3e-08 | 205 | 146 | 1 | 1 | rs8073905 |
| Caudate nucleus | 30 | 19:46464137:A:G | 19 | 46459728 | 46489105 | rs57398989 | 4.5e-09 | 14 | 10 | 1 | 1 | rs57398989 |
| Caudate nucleus | 31 | 22:37445668:C:T | 22 | 37409226 | 37470604 | rs9610638 | 7.4e-13 | 22 | 19 | 2 | 1 | rs9610638;rs5756491 |
| Dentate nucleus | 1 | 1:10315297:A:G | 1 | 10271688 | 10483606 | rs12137527 | 4.1e-09 | 50 | 35 | 1 | 1 | rs12137527 |
| Dentate nucleus | 2 | 1:15890189:A:C | 1 | 15790974 | 16049893 | rs60524694 | 1.6e-11 | 327 | 238 | 3 | 1 | rs60524694;rs927206;rs78232475 |
| Dentate nucleus | 3 | 1:43760070:C:T | 1 | 43760070 | 43856410 | rs7520981 | 3.2e-10 | 63 | 49 | 1 | 1 | rs7520981 |
| Dentate nucleus | 4 | 1:150650194:C:T | 1 | 150571149 | 150954671 | rs72702521 | 3.2e-11 | 19 | 15 | 2 | 1 | rs72702521;rs56661631 |
| Dentate nucleus | 5 | 1:205723572:C:T | 1 | 205646278 | 205803284 | rs823118 | 5.9e-13 | 110 | 79 | 5 | 1 | rs823118;rs7522056;rs823143;rs12747475; rs823084 |
| Dentate nucleus | 6 | 1:206846263:A:G | 1 | 206830925 | 206912501 | rs11119281 | 7.5e-09 | 16 | 11 | 1 | 1 | rs11119281 |
| Dentate nucleus | 7 | 1:212515613:C:T | 1 | 212408780 | 212588206 | rs1774249 | 6.8e-09 | 63 | 45 | 1 | 1 | rs1774249 |
| Dentate nucleus | 8 | 2:9777602:A:G | 2 | 9777602 | 9777602 | rs75981534 | 9.5e-10 | 1 | 1 | 1 | 1 | rs75981534 |

| Region | Locus ID | Unique variant ID | Chromosome | Start (bp) | End (bp) | Top SNP | Top P | Total SNPs | Genome-wide significant SNPs | Independent significant SNPs | Lead SNPs | Independent SNPs |
|---|---|---|---|---|---|---|---|---|---|---|---|---|
| Dentate nucleus | 9 | 2:24431184:C:T | 2 | 24366829 | 24627074 | rs2303291 | 3.9e-08 | 118 | 95 | 1 | 1 | rs2303291 |
| Dentate nucleus | 10 | 2:33764176:A:G | 2 | 33764176 | 33764176 | rs13388394 | 2.4e-15 | 1 | 1 | 1 | 1 | rs13388394 |
| Dentate nucleus | 11 | 2:48495780:C:T | 2 | 48187695 | 48766078 | rs72818477 | 1.3e-10 | 65 | 49 | 4 | 1 | rs72818477;rs6705802;rs4293602;rs11695374 |
| Dentate nucleus | 12 | 2:61368532:G:T | 2 | 61331376 | 61851845 | rs1177274 | 3.4e-10 | 354 | 230 | 3 | 1 | rs1177274;rs778756;rs7593702 |
| Dentate nucleus | 13 | 2:69705677:A:G | 2 | 69525721 | 69742989 | rs4852768 | 3.1e-10 | 276 | 204 | 3 | 2 | rs6546508;rs7577851;rs4852768 |
| Dentate nucleus | 14 | 2:120113520:A:G | 2 | 120080765 | 120155348 | rs6751553 | 3.7e-11 | 82 | 58 | 2 | 1 | rs6751553;rs72829483 |
| Dentate nucleus | 15 | 2:190358288:C:T | 2 | 189889675 | 190474215 | rs77973398 | 8.6e-17 | 592 | 449 | 8 | 2 | rs77973398;rs115369115;rs13424586;rs1123109; rs11899607;rs17271204;rs726093;rs758 ... |
| Dentate nucleus | 16 | 3:4410534:C:T | 3 | 4137745 | 4430443 | rs11915920 | 1.7e-11 | 60 | 45 | 2 | 2 | rs75118083;rs11915920 |
| Dentate nucleus | 17 | 3:133490334:A:C | 3 | 132962425 | 133581459 | rs6794370 | 6.4e-128 | 549 | 409 | 63 | 15 | rs73207717;rs144907099;rs1867506;rs6439425; rs6439426;rs77009352;rs1880669;rs9824 ... |
| Dentate nucleus | 18 | 3:148925566:A:G | 3 | 148918944 | 148991232 | rs36021368 | 6e-13 | 31 | 23 | 1 | 1 | rs36021368 |
| Dentate nucleus | 19 | 3:172431547:C:T | 3 | 172418184 | 172459863 | rs56261559 | 2.2e-08 | 34 | 26 | 1 | 1 | rs56261559 |
| Dentate nucleus | 20 | 4:827585:C:T | 4 | 810093 | 939883 | rs11727899 | 7.1e-12 | 69 | 49 | 4 | 1 | rs11727899;rs11248050;rs17781378;rs142677280 |
| Dentate nucleus | 21 | 4:26442109:A:G | 4 | 26308097 | 26465449 | rs73245789 | 4.2e-08 | 19 | 15 | 1 | 1 | rs73245789 |
| Dentate nucleus | 22 | 4:38654681:C:T | 4 | 38556399 | 38758432 | rs4833079 | 2.7e-25 | 173 | 115 | 15 | 1 | rs4833079;rs7686818;rs6531656;rs7685520; rs2045767;rs12510962;rs2381198;rs1000803 ... |
| Dentate nucleus | 23 | 4:89052323:G:T | 4 | 89039082 | 89054667 | rs2231142 | 5.2e-11 | 9 | 6 | 1 | 1 | rs2231142 |
| Dentate nucleus | 24 | 4:120019660:C:T | 4 | 120018333 | 120044474 | rs4565063 | 1.3e-09 | 47 | 34 | 1 | 1 | rs4565063 |
| Dentate nucleus | 25 | 4:123353432:A:G | 4 | 123013266 | 123517432 | rs17454584 | 9e-10 | 56 | 38 | 1 | 1 | rs17454584 |
| Dentate nucleus | 26 | 4:159689895:C:T | 4 | 159651875 | 159944851 | rs13135442 | 3.1e-15 | 277 | 192 | 3 | 1 | rs13135442;rs4600878;rs2068625 |
| Dentate nucleus | 27 | 4:185688754:C:T | 4 | 185673134 | 185731940 | rs4862415 | 1.3e-13 | 29 | 24 | 4 | 1 | rs4862415;rs2046814;rs13137179;rs13126272 |
| Dentate nucleus | 28 | 5:140884527:C:T | 5 | 140797630 | 141016288 | rs9324851 | 2e-09 | 42 | 26 | 2 | 1 | rs9324851;rs3844598 |
| Dentate nucleus | 29 | 6:26123502:C:T | 6 | 25075717 | 29603136 | rs115740542 | 6.3e-111 | 2929 | 2152 | 126 | 27 | rs12110409;rs10807000;rs72828704;rs181058325; rs34706906;rs58333539;rs35191161;rs ... |
| Dentate nucleus | 30 | 6:45447558:A:G | 6 | 45407654 | 45511945 | rs10214519 | 1.2e-10 | 74 | 53 | 1 | 1 | rs10214519 |
| Dentate nucleus | 31 | 6:108390978:A:G | 6 | 108335345 | 108411010 | rs956208 | 4.2e-15 | 154 | 111 | 3 | 1 | rs956208;rs7749081;rs11153101 |
| Dentate nucleus | 32 | 7:22301192:C:T | 7 | 22292706 | 22337720 | rs2686473 | 1.6e-11 | 32 | 22 | 2 | 1 | rs2686473;rs10225069 |
| Dentate nucleus | 33 | 7:72984220:C:T | 7 | 72823741 | 73058025 | rs2573 | 7.4e-15 | 213 | 163 | 5 | 1 | rs2573;rs11974409;rs57978469;rs142883694; rs42238 |
| Dentate nucleus | 34 | 7:99835613:G:T | 7 | 99527977 | 100345106 | rs62482175 | 1.5e-09 | 77 | 55 | 3 | 1 | rs62482175;rs13227444;rs10231649 |
| Dentate nucleus | 35 | 8:22271995:G:T | 8 | 22226052 | 22388897 | rs2280521 | 3.4e-12 | 221 | 168 | 5 | 1 | rs2280521;rs4872491;rs1989320;rs7828089; rs13276612 |
| Dentate nucleus | 36 | 8:23382985:A:G | 8 | 23220509 | 23441219 | rs6557678 | 2.1e-14 | 211 | 150 | 15 | 2 | rs6557678;rs10992;rs11782783;rs7840451; rs6557672;rs7843529;rs7813840;rs9644076;r ... |
| Dentate nucleus | 37 | 8:24742458:A:G | 8 | 24399360 | 24779437 | rs1545627 | 9.9e-14 | 185 | 129 | 6 | 2 | rs9314292;rs1545627;rs1457266;rs196864; rs1016446;rs926105 |
| Dentate nucleus | 38 | 8:27104727:A:G | 8 | 27098436 | 27119538 | rs10481349 | 2.9e-08 | 8 | 7 | 1 | 1 | rs10481349 |
| Dentate nucleus | 39 | 8:131213570:A:G | 8 | 131027338 | 131361477 | rs4733775 | 5.7e-12 | 137 | 99 | 3 | 1 | rs4733775;rs6470799;rs10956511 |
| Dentate nucleus | 40 | 8:142005245:C:T | 8 | 141645267 | 142018581 | rs306960 | 1.5e-08 | 400 | 280 | 1 | 1 | rs306960 |
| Dentate nucleus | 41 | 9:19210177:A:G | 9 | 19182872 | 19378074 | rs465423 | 1.9e-25 | 26 | 19 | 3 | 1 | rs465423;rs62560298;rs408403 |
| Dentate nucleus | 42 | 9:125550272:C:T | 9 | 125266929 | 125957759 | rs542119549 | 7.5e-14 | 12 | 10 | 9 | 4 | rs16911860;rs149587545;rs142238458; rs145918200;rs117773745;rs76707591;rs54211954 ... |
| Dentate nucleus | 43 | 10:18240466:C:T | 10 | 17862529 | 18348030 | rs2165899 | 1.9e-68 | 357 | 252 | 33 | 11 | rs530434517;rs200594495;rs143940620; rs2461184;rs2014349;rs10430577;rs72784708;rs ... |
| Dentate nucleus | 44 | 10:104697781:C:T | 10 | 104565199 | 105176914 | rs12414777 | 4.5e-14 | 269 | 196 | 4 | 2 | rs12414777;rs11191514;rs7082288;rs79780963 |
| Dentate nucleus | 45 | 10:116237760:C:T | 10 | 116228747 | 116275097 | rs11196758 | 4.2e-15 | 25 | 19 | 2 | 1 | rs11196758;rs2475232 |
| Dentate nucleus | 46 | 11:526478:A:G | 11 | 414416 | 665995 | rs35333170 | 3.2e-13 | 230 | 173 | 6 | 1 | rs35333170;rs1870726;rs2396545;rs61879160; rs10794323;rs61742833 |
| Dentate nucleus | 47 | 11:9287095:C:T | 11 | 9130147 | 9343781 | rs61877916 | 2.7e-08 | 83 | 57 | 1 | 1 | rs61877916 |
| Dentate nucleus | 48 | 11:44636833:A:G | 11 | 44619040 | 44636833 | rs2303865 | 4.9e-10 | 8 | 5 | 1 | 1 | rs2303865 |
| Dentate nucleus | 49 | 11:62523559:A:G | 11 | 62450588 | 62523559 | rs2510075 | 3.2e-09 | 83 | 57 | 1 | 1 | rs2510075 |
| Dentate nucleus | 50 | 11:63615554:C:T | 11 | 63585804 | 63797679 | rs10897452 | 3.7e-16 | 248 | 176 | 7 | 1 | rs10897452;rs11231629;rs145192830;rs3832717; rs568930;rs643634;rs4963432 |

| Region | Locus ID | Unique variant ID | Chromosome | Start (bp) | End (bp) | Top SNP | Top P | Total SNPs | Genome-wide significant SNPs | Independent significant SNPs | Lead SNPs | Independent SNPs |
|---|---|---|---|---|---|---|---|---|---|---|---|---|
| Dentate nucleus | 51 | 11:73460030:C:T | 11 | 73460030 | 73572495 | rs113450689 | 4.9e-08 | 2 | 2 | 1 | 1 | rs113450689 |
| Dentate nucleus | 52 | 11:85809825:C:T | 11 | 85657759 | 85868640 | rs645293 | 5.3e-09 | 143 | 104 | 3 | 1 | rs645293;rs629343;rs4944560 |
| Dentate nucleus | 53 | 11:109943972:C:T | 11 | 109936687 | 109953618 | rs7113219 | 1.3e-10 | 15 | 11 | 1 | 1 | rs7113219 |
| Dentate nucleus | 54 | 12:27425366:C:T | 12 | 27408778 | 27483318 | rs10771328 | 1.5e-08 | 12 | 10 | 1 | 1 | rs10771328 |
| Dentate nucleus | 55 | 12:51357542:A:G | 12 | 51326702 | 51581740 | rs12304921 | 1.6e-21 | 417 | 300 | 9 | 2 | rs12304921;rs77050960;rs3190077;rs12813728; rs1038526;rs2956428;rs11169744;rs1116 ... |
| Dentate nucleus | 56 | 12:123751339:A:G | 12 | 123492112 | 123911789 | rs2030401 | 5.1e-09 | 17 | 13 | 1 | 1 | rs2030401 |
| Dentate nucleus | 57 | 14:53827087:C:T | 14 | 53476090 | 53996341 | rs72673671 | 2.6e-15 | 680 | 510 | 11 | 2 | rs72673671;rs113034904;rs10151033;rs10149802; rs28617616;rs68148011;rs17253124;rs ... |
| Dentate nucleus | 58 | 14:76207794:A:G | 14 | 76109364 | 76444469 | rs1126190 | 9.5e-11 | 305 | 224 | 4 | 1 | rs1126190;rs999718;rs4252328;rs12887886 |
| Dentate nucleus | 59 | 15:32145735:C:T | 15 | 32068396 | 32177326 | rs4238558 | 2.3e-11 | 98 | 65 | 2 | 1 | rs4238558;rs34315282 |
| Dentate nucleus | 60 | 15:85667983:G:T | 15 | 85602142 | 85668644 | rs11852268 | 3.2e-08 | 19 | 16 | 1 | 1 | rs11852268 |
| Dentate nucleus | 61 | 16:1035548:A:G | 16 | 1004554 | 1035548 | rs3751669 | 2.4e-08 | 5 | 4 | 1 | 1 | rs3751669 |
| Dentate nucleus | 62 | 16:69622762:A:G | 16 | 69547741 | 70130803 | rs244418 | 5.5e-15 | 346 | 261 | 5 | 1 | rs244418;rs45488899;rs4985451;rs2650522; rs4783718 |
| Dentate nucleus | 63 | 16:87258088:A:G | 16 | 87220694 | 87264385 | rs11640181 | 7.2e-09 | 71 | 50 | 1 | 1 | rs11640181 |
| Dentate nucleus | 64 | 17:7167776:C:T | 17 | 7093396 | 7185861 | rs57508443 | 2.2e-10 | 72 | 52 | 2 | 1 | rs57508443;rs17671352 |
| Dentate nucleus | 65 | 17:18199150:A:G | 17 | 18040404 | 18268113 | rs8081343 | 2e-13 | 157 | 120 | 1 | 1 | rs8081343 |
| Dentate nucleus | 66 | 17:42673858:C:T | 17 | 42667406 | 43022994 | rs12451761 | 1.1e-08 | 44 | 32 | 2 | 1 | rs12451761;rs736866 |
| Dentate nucleus | 67 | 17:66282050:A:G | 17 | 66244088 | 66301495 | rs2041130 | 2e-11 | 42 | 35 | 2 | 1 | rs2041130;rs8080714 |
| Dentate nucleus | 68 | 17:73810674:A:C | 17 | 73761839 | 73974293 | rs9913385 | 1.2e-09 | 48 | 36 | 3 | 1 | rs9913385;rs112253169;rs56936829 |
| Dentate nucleus | 69 | 18:3017136:C:T | 18 | 2944809 | 3017136 | rs4798054 | 7.3e-09 | 3 | 3 | 1 | 1 | rs4798054 |
| Dentate nucleus | 70 | 19:5908163:C:T | 19 | 5872782 | 5999684 | rs146033453 | 6.9e-14 | 44 | 34 | 3 | 1 | rs146033453;rs274786;rs75566110 |
| Dentate nucleus | 71 | 19:14573327:A:G | 19 | 14557821 | 14592476 | rs8105847 | 1.3e-20 | 44 | 34 | 8 | 1 | rs8105847;rs3760702;rs1127307;rs2241357; rs2420536;rs11673475;rs12974686;rs382675 ... |
| Dentate nucleus | 72 | 19:17596144:A:G | 19 | 17580707 | 17611261 | rs62126783 | 1.2e-08 | 31 | 25 | 1 | 1 | rs62126783 |
| Dentate nucleus | 73 | 19:36204831:C:T | 19 | 36094205 | 36287869 | rs23595 | 3.6e-13 | 88 | 67 | 8 | 1 | rs23595;rs23595;rs807479;rs807479;rs7254601; rs10401695;rs10401695;rs23595 |
| Dentate nucleus | 74 | 19:53617818:C:T | 19 | 53608032 | 53621561 | rs12985137 | 3.6e-08 | 42 | 31 | 1 | 1 | rs12985137 |
| Dentate nucleus | 75 | 20:4026818:A:G | 20 | 3840539 | 4051202 | rs6037730 | 1.3e-21 | 242 | 181 | 12 | 4 | rs6084506;rs6037695;rs241642;rs6084569; rs12480920;rs149211152;rs6037730;rs812572 ... |
| Dentate nucleus | 76 | 20:25238173:C:T | 20 | 25177920 | 25991371 | rs6132824 | 6.7e-16 | 915 | 677 | 12 | 1 | rs6132824;rs2227891;rs6076335;rs2500406; rs6115218;rs4813576;rs6037243;rs6107130 ... |
| Dentate nucleus | 77 | 22:37462936:A:G | 22 | 37377094 | 37492201 | rs855791 | 1.8e-33 | 154 | 124 | 12 | 2 | rs2235320;rs877908;rs5756491;rs5750373; rs855791;rs6000550;rs9610643;rs855788;rs5 ... |
| Globus pallidus | 1 | 1:21853745:A:G | 1 | 21852961 | 21891861 | rs4654959 | 5.5e-11 | 35 | 28 | 1 | 1 | rs4654959 |
| Globus pallidus | 2 | 1:150742542:C:T | 1 | 150345558 | 151136354 | rs75581187 | 4e-10 | 91 | 70 | 2 | 1 | rs75581187;rs6667900 |
| Globus pallidus | 3 | 2:120151013:A:G | 2 | 120087644 | 120169886 | rs10179740 | 7.3e-09 | 82 | 60 | 2 | 1 | rs10179740;rs6542526 |
| Globus pallidus | 4 | 2:190109206:C:T | 2 | 189882901 | 190706339 | rs141100688 | 7.3e-53 | 941 | 707 | 28 | 6 | rs141100688;rs7599216;rs77798223;rs143278843; rs115176763;rs744556;rs11689109;rs7 ... |
| Globus pallidus | 5 | 3:133466374:A:G | 3 | 133282706 | 133536655 | rs4428180 | 1.5e-33 | 129 | 99 | 13 | 2 | rs4428180;rs8177205;rs1799852;rs6762415; rs2692695;rs6794370;rs77653936;rs6439425 ... |
| Globus pallidus | 6 | 3:147020326:C:T | 3 | 147020326 | 147076791 | rs10935665 | 1.1e-09 | 36 | 26 | 1 | 1 | rs10935665 |
| Globus pallidus | 7 | 3:195784997:A:G | 3 | 195463764 | 195844238 | rs3933 | 1.7e-20 | 292 | 221 | 19 | 2 | rs6804822;rs2688560;rs4927702;rs13093022; rs7630875;rs12498103;rs9881504;rs678503 ... |
| Globus pallidus | 8 | 4:23965563:A:G | 4 | 23926087 | 24034973 | rs13147029 | 5.9e-14 | 105 | 78 | 4 | 1 | rs13147029;rs1316862;rs655582;rs13101254 |
| Globus pallidus | 9 | 4:103188709:C:T | 4 | 102195416 | 104032368 | rs13107325 | 4.5e-111 | 1071 | 801 | 63 | 16 | rs72682927;rs17249745;rs17199964;rs34592089; rs141501067;rs201081507;rs4699257;rs ... |
| Globus pallidus | 10 | 4:159685743:C:T | 4 | 159651875 | 159874588 | rs11100197 | 4.1e-08 | 229 | 158 | 1 | 1 | rs11100197 |
| Globus pallidus | 11 | 4:185710838:C:T | 4 | 185673134 | 185712679 | rs7659090 | 2.9e-08 | 20 | 15 | 1 | 1 | rs7659090 |
| Globus pallidus | 12 | 6:26093141:A:G | 6 | 25331455 | 28941688 | rs1800562 | 8.3e-21 | 631 | 463 | 22 | 5 | rs169219;rs9467656;rs6922603;rs9968910; rs198851;rs9366626;rs12191655;rs6905614;r ... |
| Globus pallidus | 13 | 7:1574812:C:T | 7 | 1551262 | 1595839 | rs4720836 | 3.8e-08 | 128 | 113 | 1 | 1 | rs4720836 |
| Globus pallidus | 14 | 7:73007606:C:T | 7 | 72823741 | 73058025 | rs1051943 | 1.4e-10 | 209 | 160 | 2 | 1 | rs1051943;rs34594435 |
| Globus pallidus | 15 | 7:97967556:A:C | 7 | 97845713 | 98039459 | rs12671301 | 2.8e-08 | 249 | 185 | 1 | 1 | rs12671301 |

| Region | Locus ID | Unique variant ID | Chromosome | Start (bp) | End (bp) | Top SNP | Top P | Total SNPs | Genome-wide significant SNPs | Independent significant SNPs | Lead SNPs | Independent SNPs |
|---|---|---|---|---|---|---|---|---|---|---|---|---|
| Globus pallidus | 16 | 8:42428852:A:G | 8 | 42351176 | 42457590 | rs35295075 | 2.1e-09 | 184 | 142 | 1 | 1 | rs35295075 |
| Globus pallidus | 17 | 8:101676675:A:C | 8 | 101668920 | 101760211 | rs2978098 | 1.2e-33 | 107 | 73 | 8 | 1 | rs2978098;rs1693566;rs879391;rs1693565; rs1786349;rs7002825;rs1693546;rs2935545 |
| Globus pallidus | 18 | 9:19210177:A:G | 9 | 19210177 | 19210177 | rs465423 | 9.8e-10 | 1 | 1 | 1 | 1 | rs465423 |
| Globus pallidus | 19 | 9:125694309:C:T | 9 | 125694309 | 125694309 | rs138036195 | 2e-08 | 1 | 1 | 1 | 1 | rs138036195 |
| Globus pallidus | 20 | 10:18242311:A:G | 10 | 17451731 | 19011804 | rs10764176 | 9.9e-100 | 382 | 258 | 43 | 15 | rs145922999;rs72782503;rs72782510;rs11254698; rs201843067;rs555045010;rs181922661 ... |
| Globus pallidus | 21 | 10:49822099:A:G | 10 | 49818441 | 49860258 | rs4114458 | 4.5e-09 | 35 | 24 | 1 | 1 | rs4114458 |
| Globus pallidus | 22 | 10:104697781:C:T | 10 | 104697781 | 104741114 | rs12414777 | 5.4e-09 | 4 | 2 | 1 | 1 | rs12414777 |
| Globus pallidus | 23 | 10:116237760:C:T | 10 | 116228747 | 116275097 | rs11196758 | 9.9e-10 | 18 | 16 | 1 | 1 | rs11196758 |
| Globus pallidus | 24 | 11:970503:C:T | 11 | 924904 | 991109 | rs7395773 | 4.7e-08 | 180 | 142 | 1 | 1 | rs7395773 |
| Globus pallidus | 25 | 11:47581443:A:G | 11 | 47229316 | 47943419 | rs11039297 | 4.8e-11 | 117 | 88 | 4 | 1 | rs11039297;rs7944419;rs71475909;rs3740688 |
| Globus pallidus | 26 | 11:61769588:C:T | 11 | 61710538 | 61809046 | rs907638 | 9.5e-18 | 74 | 57 | 8 | 2 | rs2009875;rs4963279;rs77133046;rs2727269; rs12288004;rs907638;rs112691937;rs57280 ... |
| Globus pallidus | 27 | 12:51454786:C:T | 12 | 51325298 | 51551680 | rs11169690 | 1.5e-17 | 424 | 305 | 5 | 2 | rs71445771;rs11169690;rs224567;rs4768952; rs7975628 |
| Globus pallidus | 28 | 13:21457395:C:T | 13 | 21285496 | 21492997 | rs9506568 | 7.2e-09 | 123 | 91 | 1 | 1 | rs9506568 |
| Globus pallidus | 29 | 14:53662014:A:G | 14 | 53479695 | 53692090 | rs2552402 | 1.5e-09 | 238 | 182 | 1 | 1 | rs2552402 |
| Globus pallidus | 30 | 15:57114233:A:C | 15 | 57042063 | 57579746 | rs72747142 | 4.2e-08 | 16 | 10 | 1 | 1 | rs72747142 |
| Globus pallidus | 31 | 15:79234470:C:T | 15 | 79219188 | 79257278 | rs34843303 | 2.7e-11 | 10 | 8 | 2 | 1 | rs34843303;rs12901830 |
| Globus pallidus | 32 | 16:87235916:A:G | 16 | 87220694 | 87264534 | rs4240791 | 7e-09 | 76 | 55 | 3 | 1 | rs4240791;rs62056167;rs12711473 |
| Globus pallidus | 33 | 16:90170095:A:G | 16 | 90134240 | 90173553 | rs56407236 | 2e-08 | 13 | 13 | 1 | 1 | rs56407236 |
| Globus pallidus | 34 | 17:18057907:A:G | 17 | 18040404 | 18268113 | rs62073602 | 4.9e-12 | 154 | 116 | 2 | 1 | rs62073602;rs28364628 |
| Globus pallidus | 35 | 20:3971079:G:T | 20 | 3894497 | 4029974 | rs117255962 | 1.1e-11 | 4 | 3 | 1 | 1 | rs117255962 |
| Globus pallidus | 36 | 22:37445668:C:T | 22 | 37409226 | 37470604 | rs9610638 | 1.1e-08 | 22 | 19 | 2 | 1 | rs9610638;rs4821585 |
| Putamen | 1 | 1:110431378:C:T | 1 | 110431020 | 110432565 | rs433192 | 7.1e-13 | 3 | 1 | 1 | 1 | rs433192 |
| Putamen | 2 | 1:150725401:A:C | 1 | 150540181 | 150987886 | rs61386199 | 6.2e-15 | 48 | 38 | 3 | 1 | rs61386199;rs1355637;rs72702561 |
| Putamen | 3 | 1:205241659:C:T | 1 | 205012106 | 205297880 | rs17728 | 3.6e-12 | 85 | 63 | 3 | 2 | rs1572993;rs17728;rs56323810 |
| Putamen | 4 | 1:212639003:C:T | 1 | 212639003 | 212686682 | rs902438 | 2.7e-09 | 54 | 44 | 2 | 1 | rs902438;rs748517 |
| Putamen | 5 | 2:120111287:C:T | 2 | 120084174 | 120244703 | rs7598408 | 5.1e-22 | 179 | 121 | 9 | 2 | rs7598408;rs11888528;rs10179740;rs6542528; rs62158463;rs2579633;rs2579642;rs25796 ... |
| Putamen | 6 | 2:190431875:A:C | 2 | 190361911 | 190441066 | rs4667287 | 1.9e-10 | 45 | 33 | 3 | 1 | rs4667287;rs10206543;rs11896326 |
| Putamen | 7 | 3:52506426:C:T | 3 | 52213454 | 53138254 | rs6784615 | 1.1e-15 | 363 | 274 | 6 | 1 | rs6784615;rs1011063;rs13326165;rs111434579; rs59583591;rs352155 |
| Putamen | 8 | 3:133466374:A:G | 3 | 133252430 | 133571004 | rs4428180 | 6.1e-70 | 364 | 261 | 37 | 8 | rs1867506;rs4854733;rs6439425;rs6439426; rs77009352;rs2715634;rs9824452;rs6768696 ... |
| Putamen | 9 | 3:195840318:C:T | 3 | 195640352 | 195918266 | rs12498103 | 5.7e-15 | 67 | 51 | 6 | 4 | rs497787;rs7432894;rs9849934;rs7375124; rs12498103;rs7617766 |
| Putamen | 10 | 4:38659594:A:G | 4 | 38556399 | 38758432 | rs34722008 | 1.4e-15 | 82 | 55 | 6 | 1 | rs34722008;rs9522;rs10008032;rs6531639; rs6827279;rs3980 |
| Putamen | 11 | 4:89052323:G:T | 4 | 88913121 | 89054667 | rs2231142 | 5.9e-14 | 44 | 33 | 2 | 1 | rs2231142;rs2054576 |
| Putamen | 12 | 4:103146888:A:C | 4 | 103112470 | 103198082 | rs35225200 | 4.1e-08 | 5 | 4 | 1 | 1 | rs35225200 |
| Putamen | 13 | 4:159777764:A:C | 4 | 159651875 | 159944851 | rs7663520 | 7.3e-14 | 277 | 192 | 3 | 1 | rs7663520;rs4600878;rs2068625 |
| Putamen | 14 | 6:26093141:A:G | 6 | 25075717 | 29510630 | rs1800562 | 2.4e-72 | 1528 | 1108 | 71 | 14 | rs58333539;rs72828704;rs181058325;rs58924292; rs34706906;rs12196438;rs7762836;rs7 ... |
| Putamen | 15 | 6:45420847:C:T | 6 | 45407654 | 45519382 | rs2677100 | 2.9e-18 | 97 | 70 | 4 | 1 | rs2677100;rs2396441;rs6930053;rs4714858 |
| Putamen | 16 | 7:1574812:C:T | 7 | 1550455 | 1595839 | rs4720836 | 3e-18 | 138 | 120 | 3 | 1 | rs4720836;rs6942779;rs11981930 |
| Putamen | 17 | 7:20394948:C:T | 7 | 20371273 | 20442863 | rs73086586 | 4e-11 | 11 | 8 | 1 | 1 | rs73086586 |
| Putamen | 18 | 7:73042614:A:G | 7 | 72308916 | 73239219 | rs7786376 | 1.7e-36 | 420 | 322 | 21 | 6 | rs62465071;rs4717763;rs42238;rs62465131; rs141136126;rs35797675;rs73134963;rs5797 ... |
| Putamen | 19 | 7:73495390:C:T | 7 | 73489643 | 73537628 | rs810547 | 1.2e-09 | 20 | 15 | 1 | 1 | rs810547 |
| Putamen | 20 | 7:100456595:A:G | 7 | 100422481 | 100528223 | rs314373 | 4.7e-08 | 28 | 24 | 1 | 1 | rs314373 |
| Putamen | 21 | 8:23415655:C:T | 8 | 23217420 | 23472541 | rs11778179 | 3e-29 | 323 | 244 | 22 | 2 | rs4872141;rs721183;rs10866828;rs10992; rs11782589;rs11782783;rs62501410;rs2865688 ... |
| Putamen | 22 | 8:27104727:A:G | 8 | 27075395 | 27119538 | rs10481349 | 1.1e-12 | 28 | 25 | 3 | 1 | rs10481349;rs11990926;rs879403 |

| Region | Locus ID | Unique variant ID | Chromosome | Start (bp) | End (bp) | Top SNP | Top P | Total SNPs | Genome-wide significant SNPs | Independent significant SNPs | Lead SNPs | Independent SNPs |
|---|---|---|---|---|---|---|---|---|---|---|---|---|
| Putamen | 23 | 9:19210177:A:G | 9 | 19210177 | 19378074 | rs465423 | 8.1e-31 | 37 | 24 | 3 | 1 | rs465423;rs62560298;rs7855666 |
| Putamen | 24 | 9:125694309:C:T | 9 | 125550272 | 125957759 | rs138036195 | 2.4e-10 | 5 | 3 | 3 | 2 | rs542119549;rs138036195;rs143663770 |
| Putamen | 25 | 10:18243703:A:G | 10 | 17451731 | 18418644 | rs690759 | 2.5e-88 | 376 | 259 | 40 | 13 | rs145922999;rs72782503;rs201843067; rs555045010;rs181922661;rs530434517;rs3705906 ... |
| Putamen | 26 | 10:116237760:C:T | 10 | 116228747 | 116360503 | rs11196758 | 9.3e-23 | 123 | 98 | 6 | 2 | rs11196758;rs2475232;rs56339171;rs4255462; rs2483569;rs2475241 |
| Putamen | 27 | 11:30510599:C:T | 11 | 30492070 | 30569363 | rs35827570 | 1.6e-12 | 41 | 29 | 1 | 1 | rs35827570 |
| Putamen | 28 | 11:47434986:A:G | 11 | 47377283 | 47943419 | rs2293576 | 1.6e-10 | 299 | 208 | 3 | 1 | rs2293576;rs11039409;rs10838777 |
| Putamen | 29 | 11:63587117:C:T | 11 | 63585804 | 63598802 | rs630486 | 4.6e-08 | 20 | 16 | 1 | 1 | rs630486 |
| Putamen | 30 | 11:85800279:A:G | 11 | 85652251 | 85868640 | rs561655 | 4.7e-09 | 139 | 96 | 2 | 1 | rs561655;rs592314 |
| Putamen | 31 | 11:109940267:A:G | 11 | 109936687 | 109953618 | rs10891050 | 2.7e-15 | 15 | 11 | 1 | 1 | rs10891050 |
| Putamen | 32 | 11:128531739:A:C | 11 | 128525423 | 128555379 | rs694438 | 1.9e-08 | 9 | 7 | 1 | 1 | rs694438 |
| Putamen | 33 | 12:26507016:A:G | 12 | 26310234 | 26529242 | rs10842717 | 4.2e-11 | 51 | 35 | 6 | 3 | rs11048420;rs9804706;rs76829083;rs10842718; rs10842717;rs11048473 |
| Putamen | 34 | 12:51484646:C:T | 12 | 51223579 | 51581740 | rs7979705 | 2.8e-40 | 632 | 454 | 12 | 3 | rs11169664;rs7135059;rs2956428;rs829022; rs7979705;rs10160844;rs117234840;rs22456 ... |
| Putamen | 35 | 12:113740871:A:G | 12 | 113536863 | 113752802 | rs77625177 | 2.6e-17 | 261 | 198 | 5 | 1 | rs77625177;rs75426961;rs146487929;rs61941404; rs34649612 |
| Putamen | 36 | 12:133780309:G:T | 12 | 133688707 | 133781163 | rs36127550 | 3.6e-08 | 4 | 4 | 1 | 1 | rs36127550 |
| Putamen | 37 | 15:45388382:C:T | 15 | 45365790 | 45409271 | rs2554451 | 1.1e-08 | 34 | 28 | 1 | 1 | rs2554451 |
| Putamen | 38 | 15:57295579:A:G | 15 | 56876088 | 57601813 | rs72749499 | 1.5e-14 | 250 | 180 | 4 | 1 | rs72749499;rs72736018;rs72744924;rs13379960 |
| Putamen | 39 | 15:79237293:C:T | 15 | 78714008 | 79257278 | rs2289702 | 2.4e-25 | 568 | 429 | 14 | 3 | rs12910090;rs12912022;rs34684276;rs4887070; rs11636753;rs12905641;rs56227704;rs11 ... |
| Putamen | 40 | 16:57319143:A:C | 16 | 57158679 | 57320113 | rs7187527 | 5.6e-16 | 75 | 61 | 4 | 1 | rs7187527;rs7205444;rs4784771;rs113885775 |
| Putamen | 41 | 16:79606520:C:T | 16 | 79586108 | 79631946 | rs434626 | 1e-27 | 86 | 64 | 13 | 2 | rs434626;rs434443;rs1559341;rs30388;rs250164; rs250150;rs250148;rs13337519;rs3042 ... |
| Putamen | 42 | 16:90170811:A:G | 16 | 90125822 | 90173553 | rs201497512 | 1.6e-09 | 15 | 15 | 2 | 1 | rs201497512;rs7197795 |
| Putamen | 43 | 17:7172609:A:G | 17 | 7105399 | 7187123 | rs12601936 | 9.5e-09 | 65 | 46 | 1 | 1 | rs12601936 |
| Putamen | 44 | 17:27678484:G:T | 17 | 27660490 | 27881366 | rs60018793 | 1.7e-09 | 85 | 65 | 1 | 1 | rs60018793 |
| Putamen | 45 | 17:40741013:C:T | 17 | 40287106 | 41018481 | rs12951632 | 6.1e-49 | 565 | 415 | 24 | 4 | rs144993221;rs117559165;rs117292219; rs17320971;rs11871731;rs72823055;rs12600758 ... |
| Putamen | 46 | 17:42281282:G:T | 17 | 42197213 | 42290015 | rs4473241 | 4.6e-08 | 83 | 59 | 1 | 1 | rs4473241 |
| Putamen | 47 | 17:44051589:G:T | 17 | 43460181 | 44874453 | rs118087478 | 1.6e-30 | 4124 | 3099 | 50 | 3 | rs2696471;rs2693361;rs3976763;rs2462838; rs114316249;rs2693366;rs2696467;rs559381 ... |
| Putamen | 48 | 17:57696251:A:G | 17 | 57556414 | 58226089 | rs1881442 | 1.5e-18 | 471 | 321 | 13 | 1 | rs1881442;rs8069123;rs10853015;rs4968391; rs11654074;rs150581172;rs180536;rs57964 ... |
| Putamen | 49 | 21:16798091:G:T | 21 | 16781136 | 16838662 | rs2403966 | 1.9e-09 | 91 | 67 | 2 | 1 | rs2403966;rs2823286 |
| Putamen | 50 | 21:40292946:A:G | 21 | 40289167 | 40316902 | rs7278605 | 1e-13 | 41 | 31 | 3 | 1 | rs7278605;rs13053080;rs6517487 |
| Putamen | 51 | 22:37462936:A:G | 22 | 37390400 | 37508507 | rs855791 | 6.2e-27 | 157 | 127 | 13 | 3 | rs2235320;rs56270606;rs5756491;rs5750373; rs135167;rs855791;rs9610643;rs855788;rs ... |
| Red nucleus | 1 | 1:35253844:C:T | 1 | 35253844 | 35266192 | rs6697367 | 2.4e-08 | 11 | 8 | 1 | 1 | rs6697367 |
| Red nucleus | 2 | 1:150868102:A:C | 1 | 150526406 | 150954671 | rs78132593 | 1.3e-13 | 24 | 17 | 4 | 1 | rs78132593;rs6681639;rs1355637;rs66958685 |
| Red nucleus | 3 | 1:240695636:G:T | 1 | 240695532 | 240706107 | rs7547238 | 8.9e-09 | 12 | 7 | 3 | 2 | rs7547238;rs10495471;rs10926294 |
| Red nucleus | 4 | 2:63111289:C:T | 2 | 62742841 | 63934167 | rs76536971 | 1.4e-15 | 150 | 113 | 5 | 2 | rs13017269;rs76536971;rs2162011;rs76262922; rs6714241 |
| Red nucleus | 5 | 2:120120877:A:G | 2 | 120080765 | 120169886 | rs6723209 | 7.1e-12 | 84 | 61 | 3 | 1 | rs6723209;rs28478111;rs12612942 |
| Red nucleus | 6 | 2:190175090:G:T | 2 | 189882901 | 190706339 | rs114335166 | 4e-37 | 899 | 677 | 22 | 5 | rs73051219;rs375880845;rs115176763; rs78262626;rs6434308;rs114335166;rs13424586;r ... |
| Red nucleus | 7 | 3:133490334:A:C | 3 | 133253719 | 133571004 | rs6794370 | 6.3e-46 | 335 | 249 | 35 | 10 | rs1867506;rs6439425;rs6439426;rs77009352; rs2692695;rs9824452;rs149591457;rs81771 ... |
| Red nucleus | 8 | 3:195840318:C:T | 3 | 195468430 | 195844238 | rs12498103 | 3.4e-15 | 207 | 158 | 16 | 3 | rs9880934;rs2688560;rs4927702;rs56314672; rs11925423;rs10452050;rs73208061;rs9817 ... |
| Red nucleus | 9 | 4:38654681:C:T | 4 | 38592440 | 38716721 | rs4833079 | 6.7e-12 | 60 | 36 | 3 | 1 | rs4833079;rs337637;rs4634234 |
| Red nucleus | 10 | 4:89052323:G:T | 4 | 89039082 | 89054667 | rs2231142 | 5.3e-10 | 9 | 6 | 1 | 1 | rs2231142 |
| Red nucleus | 11 | 4:103188709:C:T | 4 | 102702364 | 103387161 | rs13107325 | 1.8e-14 | 30 | 25 | 4 | 1 | rs13107325;rs34333163;rs17199964;rs34592089 |

| Region | Locus ID | Unique variant ID | Chromosome | Start (bp) | End (bp) | Top SNP | Top P | Total SNPs | Genome-wide significant SNPs | Independent significant SNPs | Lead SNPs | Independent SNPs |
|---|---|---|---|---|---|---|---|---|---|---|---|---|
| Red nucleus | 12 | 4:120019660:C:T | 4 | 120018333 | 120044474 | rs4565063 | 1.3e-09 | 47 | 34 | 1 | 1 | rs4565063 |
| Red nucleus | 13 | 4:159674887:C:T | 4 | 159651875 | 159944851 | rs35782846 | 1.6e-09 | 288 | 198 | 2 | 1 | rs35782846;rs9784569 |
| Red nucleus | 14 | 4:185704176:G:T | 4 | 185673134 | 185712679 | rs2046814 | 1.6e-10 | 21 | 16 | 2 | 1 | rs2046814;rs7659090 |
| Red nucleus | 15 | 6:26093141:A:G | 6 | 25075717 | 28147855 | rs1800562 | 8.5e-52 | 1227 | 901 | 57 | 15 | rs7773244;rs5009714;rs77483728;rs55731467; rs73387233;rs77042711;rs75580845;rs347 ... |
| Red nucleus | 16 | 8:22279863:A:C | 8 | 22112929 | 22410157 | rs1051708 | 1.1e-25 | 449 | 332 | 11 | 3 | rs4266653;rs7826061;rs1051647;rs1051708; rs10099515;rs2461480;rs754270;rs2005080 ... |
| Red nucleus | 17 | 8:23382985:A:G | 8 | 23356364 | 23384559 | rs6557678 | 1.9e-12 | 30 | 20 | 3 | 1 | rs6557678;rs9644076;rs62503281 |
| Red nucleus | 18 | 8:101679224:G:T | 8 | 101673265 | 101760211 | rs2978101 | 1.8e-24 | 105 | 70 | 6 | 1 | rs2978101;rs1786338;rs1786316;rs1693546; rs1693549;rs2935545 |
| Red nucleus | 19 | 9:19210177:A:G | 9 | 19210177 | 19378074 | rs465423 | 9.1e-20 | 12 | 8 | 2 | 1 | rs465423;rs62560298 |
| Red nucleus | 20 | 9:109137188:C:T | 9 | 108901453 | 109144194 | rs76289637 | 1.6e-14 | 80 | 51 | 2 | 1 | rs76289637;rs75089243 |
| Red nucleus | 21 | 9:136142203:A:C | 9 | 136132908 | 136149500 | rs514659 | 1.3e-09 | 48 | 32 | 1 | 1 | rs514659 |
| Red nucleus | 22 | 10:18243703:A:G | 10 | 17928439 | 19011804 | rs690759 | 1e-34 | 196 | 138 | 20 | 6 | rs200594495;rs143940620;rs2014349;rs10430578; rs72784708;rs56336322;rs12263430;rs ... |
| Red nucleus | 23 | 10:51563993:A:G | 10 | 51563700 | 51563993 | rs10994675 | 3.2e-08 | 2 | 2 | 1 | 1 | rs10994675 |
| Red nucleus | 24 | 10:104697781:C:T | 10 | 104565199 | 105176914 | rs12414777 | 1.4e-12 | 269 | 196 | 4 | 2 | rs12414777;rs11191574;rs7082288;rs79780963 |
| Red nucleus | 25 | 10:116255019:G:T | 10 | 116228747 | 116275097 | rs9421175 | 5.3e-11 | 25 | 19 | 3 | 1 | rs9421175;rs2475232;rs11196752 |
| Red nucleus | 26 | 11:44635208:A:G | 11 | 44619040 | 44636833 | rs7930904 | 2e-09 | 9 | 6 | 2 | 1 | rs7930904;rs2303865 |
| Red nucleus | 27 | 11:61738545:G:T | 11 | 61728651 | 61810183 | rs12799017 | 6.1e-45 | 198 | 153 | 14 | 4 | rs113017397;rs10897193;rs112262426; rs57937104;rs78256861;rs4963283;rs12799017;rs ... |
| Red nucleus | 28 | 11:109940267:A:G | 11 | 109936687 | 109953618 | rs10891050 | 3.8e-18 | 15 | 11 | 1 | 1 | rs10891050 |
| Red nucleus | 29 | 12:51357542:A:G | 12 | 51223579 | 51581740 | rs12304921 | 6.9e-24 | 501 | 361 | 8 | 2 | rs12304921;rs77050960;rs3190077;rs4768938; rs11169672;rs1038526;rs11169754;rs1116 ... |
| Red nucleus | 30 | 12:113661258:G:T | 12 | 113588828 | 113752802 | rs73192811 | 1.6e-08 | 221 | 167 | 1 | 1 | rs73192811 |
| Red nucleus | 31 | 14:53827087:C:T | 14 | 53696031 | 53992930 | rs72673671 | 4.8e-11 | 162 | 124 | 4 | 2 | rs72673671;rs113034904;rs9796362;rs17253326 |
| Red nucleus | 32 | 17:7127718:C:T | 17 | 7092704 | 7187123 | rs17671352 | 3.3e-14 | 86 | 61 | 6 | 1 | rs17671352;rs222837;rs113086489;rs2654189; rs188777;rs3826408 |
| Red nucleus | 33 | 17:18057301:A:G | 17 | 18040404 | 18268113 | rs735960 | 2.8e-14 | 161 | 123 | 3 | 1 | rs735960;rs2061214;rs8080168 |
| Red nucleus | 34 | 17:19802247:C:T | 17 | 19799698 | 19911649 | rs9914273 | 1.4e-09 | 58 | 44 | 1 | 1 | rs9914273 |
| Red nucleus | 35 | 17:40696915:C:T | 17 | 40568094 | 40824834 | rs684214 | 1.9e-16 | 119 | 84 | 3 | 1 | rs684214;rs4796657;rs647397 |
| Red nucleus | 36 | 17:62284510:A:G | 17 | 62274324 | 62318545 | rs8072646 | 8.9e-09 | 9 | 7 | 1 | 1 | rs8072646 |
| Red nucleus | 37 | 17:73935233:G:T | 17 | 73820601 | 73974293 | rs73355708 | 5.6e-09 | 24 | 19 | 1 | 1 | rs73355708 |
| Red nucleus | 38 | 20:3996208:C:T | 20 | 3840539 | 4028706 | rs6116153 | 2.5e-13 | 222 | 166 | 6 | 1 | rs6116153;rs6052246;rs8125729;rs6116164; rs11906481;rs241596 |
| Red nucleus | 39 | 22:37462936:A:G | 22 | 37390400 | 37492201 | rs855791 | 8.9e-23 | 105 | 83 | 8 | 2 | rs2235320;rs877908;rs5756489;rs5750373; rs855791;rs9610643;rs855788;rs130605 |
| Substantia nigra | 1 | 1:205045087:A:G | 1 | 205044339 | 205261963 | rs1572993 | 2.4e-08 | 8 | 7 | 1 | 1 | rs1572993 |
| Substantia nigra | 2 | 2:190097538:A:G | 2 | 189882901 | 190706339 | rs73980192 | 3.8e-45 | 838 | 635 | 18 | 5 | rs73051219;rs375880845;rs115176763; rs78262626;rs12617111;rs73980192;rs76762748;r ... |
| Substantia nigra | 3 | 3:133466374:A:G | 3 | 133282706 | 133536655 | rs4428180 | 9e-25 | 142 | 108 | 16 | 4 | rs189448514;rs4428180;rs1799852;rs6762415; rs2692695;rs6794370;rs9824452;rs776539 ... |
| Substantia nigra | 4 | 3:195784997:A:G | 3 | 195645945 | 195844238 | rs3933 | 2.6e-13 | 177 | 133 | 8 | 1 | rs3933;rs111625920;rs6785037;rs2261192; rs75307539;rs4927689;rs4927707;rs9817514 |
| Substantia nigra | 5 | 4:38654681:C:T | 4 | 38594895 | 38718732 | rs4833079 | 9.2e-10 | 55 | 34 | 3 | 1 | rs4833079;rs13149231;rs4634234 |
| Substantia nigra | 6 | 4:89054667:A:G | 4 | 88913121 | 89054667 | rs4148155 | 2.1e-11 | 44 | 33 | 2 | 1 | rs4148155;rs2054576 |
| Substantia nigra | 7 | 4:103188709:C:T | 4 | 102195416 | 104069822 | rs13107325 | 1.8e-70 | 767 | 581 | 44 | 12 | rs78009918;rs17249398;rs17249745;rs201081507; rs4371620;rs13136118;rs10032160;rs1 ... |
| Substantia nigra | 8 | 4:123067808:A:G | 4 | 123013266 | 123541341 | rs56035021 | 7.5e-09 | 88 | 58 | 2 | 1 | rs56035021;rs45610037 |
| Substantia nigra | 9 | 4:185704176:G:T | 4 | 185704176 | 185704176 | rs2046814 | 6.5e-09 | 1 | 1 | 1 | 1 | rs2046814 |
| Substantia nigra | 10 | 6:26098474:C:T | 6 | 25452783 | 27249439 | rs79220007 | 3.6e-24 | 578 | 413 | 26 | 5 | rs13203202;rs3804112;rs78151190;rs62393683; rs72838866;rs116272812;rs9348689;rs45 ... |
| Substantia nigra | 11 | 8:101676675:A:C | 8 | 101668920 | 101760211 | rs2978098 | 8.4e-24 | 107 | 72 | 7 | 1 | rs2978098;rs1693566;rs1786338;rs1693565; rs1786349;rs1693546;rs2935545 |
| Substantia nigra | 12 | 9:19210177:A:G | 9 | 19210177 | 19210177 | rs465423 | 2e-08 | 1 | 1 | 1 | 1 | rs465423 |

| Region | Locus ID | Unique variant ID | Chromosome | Start (bp) | End (bp) | Top SNP | Top P | Total SNPs | Genome-wide significant SNPs | Independent significant SNPs | Lead SNPs | Independent SNPs |
|---|---|---|---|---|---|---|---|---|---|---|---|---|
| Substantia nigra | 13 | 9:109112718:C:T | 9 | 108838212 | 109151678 | rs117197034 | 2.3e-21 | 187 | 127 | 5 | 1 | rs117197034;rs57617805;rs75089243; rs117532980;rs1484374 |
| Substantia nigra | 14 | 10:18239344:G:T | 10 | 18192889 | 19011804 | rs12249671 | 1e-18 | 126 | 92 | 8 | 1 | rs12249671;rs11592271;rs1926740;rs11011865; rs11594139;rs10508553;rs691068;rs7278 ... |
| Substantia nigra | 15 | 10:51565548:C:T | 10 | 51565548 | 51649753 | rs7909927 | 3.7e-08 | 10 | 6 | 1 | 1 | rs7909927 |
| Substantia nigra | 16 | 10:116252408:C:T | 10 | 116228747 | 116275097 | rs11196764 | 5.2e-10 | 18 | 16 | 1 | 1 | rs11196764 |
| Substantia nigra | 17 | 11:61744342:C:T | 11 | 61739142 | 61812942 | rs195445 | 3.8e-10 | 13 | 9 | 2 | 2 | rs195445;rs7948767 |
| Substantia nigra | 18 | 11:109943972:C:T | 11 | 109936687 | 109953618 | rs7113219 | 1.2e-09 | 15 | 11 | 1 | 1 | rs7113219 |
| Substantia nigra | 19 | 12:51335777:G:T | 12 | 51326702 | 51581007 | rs73090629 | 8.2e-11 | 365 | 265 | 4 | 2 | rs73090629;rs4768952;rs11169690;rs11169672 |
| Substantia nigra | 20 | 14:75108290:C:T | 14 | 75062398 | 75377692 | rs12892228 | 2.3e-09 | 172 | 132 | 2 | 1 | rs12892228;rs11159098 |
| Substantia nigra | 21 | 15:57218850:A:G | 15 | 57046107 | 57583301 | rs7178030 | 2.7e-08 | 178 | 125 | 1 | 1 | rs7178030 |
| Substantia nigra | 22 | 20:3923758:C:T | 20 | 3846843 | 4018314 | rs6084521 | 1.8e-10 | 92 | 69 | 2 | 1 | rs6084521;rs6052248 |
| Substantia nigra | 23 | 22:37462936:A:G | 22 | 37409226 | 37470604 | rs855791 | 7e-09 | 22 | 19 | 2 | 1 | rs855791;rs5756491 |

*Abbreviations: Chr = chromosome; GWAS = genome-wide association study; IndSig = independent significant; SNP = single-nucleotide polymorphism; uniqID = unique variant identifier (chromosome:position:reference:alternate). Loci are reported per region; the same locus may appear for more than one region. Positions are given on GRCh37. Long variant and gene lists are abbreviated in this document; complete values are given in the accompanying spreadsheet workbook.*

### Supplementary Table 2. Independent loci associated with quantitative susceptibility mapping-derived brain iron after cross-regional merging, with replication status against previously published quantitative susceptibility mapping analyses.

| Locus ID | Unique variant ID | Chromosome | Start (bp) | End (bp) | Width (kb) | Top SNP | Top P | Top region | Number of regions | Regions | Total SNPs | Genome-wide significant SNPs | Independent significant SNPs | Lead SNPs | Independent SNPs | Pleiotropy | Replication (Casanova) | Replication (Wang) | Overall status |
|---|---|---|---|---|---|---|---|---|---|---|---|---|---|---|---|---|---|---|---|
| 1 | 1:10315297:A:G | 1 | 10271688 | 10483606 | 211.9 | rs12137527 | 4.1e-09 | Dentate nucleus | 1 | Dentate | 50 | 35 | 1 | 1 | rs12137527 | Region-specific | Novel | Novel | Novel |
| 2 | 1:15890189:A:C | 1 | 15790974 | 16049893 | 258.9 | rs60524694 | 1.6e-11 | Dentate nucleus | 1 | Dentate | 327 | 238 | 3 | 1 | rs60524694;rs927206;rs78232475 | Region-specific | Novel | Novel | Novel |
| 3 | 1:21853745:A:G | 1 | 21852961 | 21891861 | 38.9 | rs4654959 | 5.5e-11 | Globus pallidus | 1 | Globus_Pallidus | 35 | 28 | 1 | 1 | rs4654959 | Region-specific | Replicated | Novel | Replicated (Casanova) |
| 4 | 1:35253844:C:T | 1 | 35253844 | 35266192 | 12.3 | rs6697367 | 2.4e-08 | Red nucleus | 1 | Red_Nucleus | 11 | 8 | 1 | 1 | rs6697367 | Region-specific | Novel | Novel | Novel |
| 5 | 1:43760070:C:T | 1 | 43760070 | 43856410 | 96.3 | rs7520981 | 3.2e-10 | Dentate nucleus | 1 | Dentate | 63 | 49 | 1 | 1 | rs7520981 | Region-specific | Replicated | Novel | Replicated (Casanova) |
| 6 | 1:110431378:C:T | 1 | 110431020 | 110432565 | 1.5 | rs433192 | 7.1e-13 | Putamen | 1 | Putamen | 3 | 1 | 1 | 1 | rs433192 | Region-specific | Replicated | Novel | Replicated (Casanova) |
| 7 | 1:150725401:A:C;1:150742542:C:T;<br>1:150868102:A:C;1:150650194:C:T | 1 | 150345558 | 151136354 | 790.8 | rs61386199 | 6.2e-15 | Putamen | 4 | Dentate;Globus_Pallidus;Putamen;<br>Red_Nucleus | 182 | 140 | 11 | 4 | rs61386199;rs1355637;rs72702561;<br>rs75581187;rs6667900;rs78132593;<br>rs6681639;rs13 ... | Pleiotropic | Replicated | Novel | Replicated (Casanova) |
| 8 | 1:205248032:A:G;1:205241659:C:T;<br>1:205045087:A:G;1:205723572:C:T | 1 | 205012106 | 205803284 | 791.2 | rs823118 | 5.9e-13 | Dentate nucleus | 4 | Caudate;Dentate;Putamen;Substantia_Nigra | 279 | 204 | 10 | 5 | rs1172134;rs1572993;rs17728;rs56323810;<br>rs1572993;rs823118;rs7522056;rs823143 ... | Pleiotropic | Replicated | Novel | Replicated (Casanova) |
| 9 | 1:206846263:A:G | 1 | 206830925 | 206912501 | 81.6 | rs11119281 | 7.5e-09 | Dentate nucleus | 1 | Dentate | 16 | 11 | 1 | 1 | rs11119281 | Region-specific | Novel | Novel | Novel |
| 10 | 1:212639003:C:T;1:212515613:C:T | 1 | 212408780 | 212686682 | 277.9 | rs902438 | 2.7e-09 | Putamen | 2 | Dentate;Putamen | 117 | 89 | 3 | 2 | rs902438;rs748517;rs1774249 | Pleiotropic | Replicated | Novel | Replicated (Casanova) |
| 11 | 1:240695636:G:T | 1 | 240695532 | 240706107 | 10.6 | rs7547238 | 8.9e-09 | Red nucleus | 1 | Red_Nucleus | 12 | 7 | 3 | 2 | rs7547238;rs10495471;rs10926294 | Region-specific | Novel | Novel | Novel |
| 12 | 2:9777602:A:G | 2 | 9777602 | 9777602 | 0 | rs75981534 | 9.5e-10 | Dentate nucleus | 1 | Dentate | 1 | 1 | 1 | 1 | rs75981534 | Region-specific | Novel | Novel | Novel |
| 13 | 2:24431184:C:T | 2 | 24366829 | 24627074 | 260.2 | rs2303291 | 3.9e-08 | Dentate nucleus | 1 | Dentate | 118 | 95 | 1 | 1 | rs2303291 | Region-specific | Replicated | Novel | Replicated (Casanova) |
| 14 | 2:33764176:A:G | 2 | 33764176 | 33764176 | 0 | rs13388394 | 2.4e-15 | Dentate nucleus | 1 | Dentate | 1 | 1 | 1 | 1 | rs13388394 | Region-specific | Novel | Novel | Novel |
| 15 | 2:48495780:C:T | 2 | 48187695 | 48766078 | 578.4 | rs72818477 | 1.3e-10 | Dentate nucleus | 1 | Dentate | 65 | 49 | 4 | 1 | rs72818477;rs6705802;rs4293602;<br>rs11695374 | Region-specific | Novel | Novel | Novel |
| 16 | 2:63111289:C:T;2:61368532:G:T | 2 | 61331376 | 63934167 | 2602.8 | rs76536971 | 1.4e-15 | Red nucleus | 2 | Dentate;Red_Nucleus | 504 | 343 | 8 | 3 | rs13017269;rs76536971;rs2162011;<br>rs76262922;rs6714241;rs1177274;rs778756;<br>rs75937 ... | Pleiotropic | Novel | Novel | Novel |
| 17 | 2:69705677:A:G | 2 | 69525721 | 69742989 | 217.3 | rs4852768 | 3.1e-10 | Dentate nucleus | 1 | Dentate | 276 | 204 | 3 | 2 | rs6546508;rs7577851;rs4852768 | Region-specific | Novel | Novel | Novel |
| 18 | 2:120155348:A:G;2:120111287:C:T;<br>2:120151013:A:G;2:120120877:A:G;<br>2:120113520: ... | 2 | 120080765 | 120244703 | 163.9 | rs7598408 | 5.1e-22 | Putamen | 5 | Caudate;Dentate;Globus_Pallidus;Putamen;<br>Red_Nucleus | 522 | 370 | 19 | 7 | rs72833227;rs6722204;rs2579633;<br>rs7598408;rs11888528;rs10179740;<br>rs6542528;rs6215 ... | Pleiotropic | Replicated | Novel | Replicated (Casanova) |
| 19 | 2:190382493:A:G;2:190431875:A:C;<br>2:190109206:C:T;2:190097538:A:G;<br>2:190175090: ... | 2 | 189882901 | 190706339 | 823.4 | rs141100688 | 7.3e-53 | Globus pallidus | 6 | Caudate;Dentate;Globus_Pallidus;Putamen;<br>Red_Nucleus;Substantia_Nigra | 3335 | 2517 | 80 | 20 | rs12616162;rs4667287;rs10206543;<br>rs11896326;rs141100688;rs7599216;<br>rs77798223;rs ... | Pan-regional | Replicated | Novel | Replicated (Casanova) |
| 20 | 3:4410534:C:T | 3 | 4137745 | 4430443 | 292.7 | rs11915920 | 1.7e-11 | Dentate nucleus | 1 | Dentate | 60 | 45 | 2 | 2 | rs75118083;rs11915920 | Region-specific | Novel | Novel | Novel |
| 21 | 3:52506426:C:T | 3 | 52213454 | 53138254 | 924.8 | rs6784615 | 1.1e-15 | Putamen | 1 | Putamen | 363 | 274 | 6 | 1 | rs6784615;rs1011063;rs13326165;<br>rs111434579;rs59583591;rs352155 | Region-specific | Replicated | Novel | Replicated (Casanova) |
| 22 | 3:107464170:A:G | 3 | 107464170 | 107470613 | 6.4 | rs11710737 | 2.6e-09 | Caudate nucleus | 1 | Caudate | 2 | 2 | 1 | 1 | rs11710737 | Region-specific | Novel | Novel | Novel |
| 23 | 3:133466374:A:G;3:133490334:A:C | 3 | 132962425 | 133581459 | 619 | rs6794370 | 6.4e-128 | Dentate nucleus | 6 | Caudate;Dentate;Globus_Pallidus;Putamen;<br>Red_Nucleus;Substantia_Nigra | 1700 | 1254 | 179 | 41 | rs4428180;rs1799852;rs6762415;rs1405022;<br>rs6794370;rs9824452;rs77653936;<br>rs6439425 ... | Pan-regional | Replicated | Novel | Replicated (Casanova) |
| 24 | 3:147032401:A:G;3:147020326:C:T | 3 | 147020326 | 147082740 | 62.4 | rs60852193 | 2.1e-10 | Caudate nucleus | 2 | Caudate;Globus_Pallidus | 76 | 55 | 4 | 2 | rs60852193;rs66507831;rs4681356;<br>rs10935665 | Pleiotropic | Novel | Novel | Novel |
| 25 | 3:148925566:A:G | 3 | 148918944 | 148991232 | 72.3 | rs36021368 | 6e-13 | Dentate nucleus | 1 | Dentate | 31 | 23 | 1 | 1 | rs36021368 | Region-specific | Replicated | Novel | Replicated (Casanova) |
| 26 | 3:172431547:C:T | 3 | 172418184 | 172459863 | 41.7 | rs56261559 | 2.2e-08 | Dentate nucleus | 1 | Dentate | 34 | 26 | 1 | 1 | rs56261559 | Region-specific | Novel | Novel | Novel |
| 27 | 3:195841644:C:T;3:195840318:C:T;<br>3:195784997:A:G | 3 | 195463764 | 195918266 | 454.5 | rs3933 | 1.7e-20 | Globus pallidus | 5 | Caudate;Globus_Pallidus;Putamen;<br>Red_Nucleus;Substantia_Nigra | 822 | 621 | 54 | 11 | rs7432894;rs9849934;rs41298089;<br>rs6583290;rs12498103;rs497787;rs7432894;<br>rs984993 ... | Pleiotropic | Replicated | Novel | Replicated (Casanova) |
| 28 | 4:827585:C:T | 4 | 810093 | 939883 | 129.8 | rs11727899 | 7.1e-12 | Dentate nucleus | 1 | Dentate | 69 | 49 | 4 | 1 | rs11727899;rs11248050;rs17781378;<br>rs142677280 | Region-specific | Novel | Novel | Novel |
| 29 | 4:15737348:A:G | 4 | 15710330 | 15743332 | 33 | rs4698412 | 4.9e-08 | Caudate nucleus | 1 | Caudate | 11 | 11 | 1 | 1 | rs4698412 | Region-specific | Replicated | Novel | Replicated (Casanova) |
| 30 | 4:23965563:A:G | 4 | 23926087 | 24034973 | 108.9 | rs13147029 | 5.9e-14 | Globus pallidus | 1 | Globus_Pallidus | 105 | 78 | 4 | 1 | rs13147029;rs1316862;rs655582;<br>rs13101254 | Region-specific | Replicated | Novel | Replicated (Casanova) |
| 31 | 4:26442109:A:G | 4 | 26308097 | 26465449 | 157.4 | rs73245789 | 4.2e-08 | Dentate nucleus | 1 | Dentate | 19 | 15 | 1 | 1 | rs73245789 | Region-specific | Novel | Novel | Novel |
| 32 | 4:38659594:A:G;4:38654681:C:T | 4 | 38556399 | 38758432 | 202 | rs4833079 | 2.7e-25 | Dentate nucleus | 4 | Dentate;Putamen;Red_Nucleus;<br>Substantia_Nigra | 370 | 240 | 27 | 4 | rs34722008;rs9522;rs10008032;rs6531639;<br>rs6827279;rs3980;rs4833079;rs13149231;rs<br>... | Pleiotropic | Replicated | Novel | Replicated (Casanova) |
| 33 | 4:89052323:G:T;4:89054667:A:G | 4 | 88913121 | 89054667 | 141.5 | rs2231142 | 5.9e-14 | Putamen | 4 | Dentate;Putamen;Red_Nucleus;<br>Substantia_Nigra | 106 | 78 | 6 | 4 | rs2231142;rs2054576;rs4148155;rs2054576;<br>rs2231142 | Pleiotropic | Replicated | Novel | Replicated (Casanova) |
| 34 | 4:103188709:C:T;4:103146888:A:C | 4 | 101920593 | 104163862 | 2243.3 | rs13107325 | 7.3e-148 | Caudate nucleus | 5 | Caudate;Globus_Pallidus;Putamen;<br>Red_Nucleus;Substantia_Nigra | 2949 | 2216 | 181 | 47 | rs2063058;rs4698918;rs17030740;<br>rs76062146;rs17249745;rs17199964;<br>rs34592089;rs779 ... | Pleiotropic | Replicated | Novel | Replicated (Casanova) |
| 35 | 4:120019660:C:T | 4 | 120018333 | 120044474 | 26.1 | rs4565063 | 1.3e-09 | Red nucleus | 2 | Dentate;Red_Nucleus | 94 | 68 | 2 | 2 | rs4565063 | Pleiotropic | Replicated | Novel | Replicated (Casanova) |
| 36 | 4:123067808:A:G;4:123353432:A:G | 4 | 123013266 | 123541341 | 528.1 | rs17454584 | 9e-10 | Dentate nucleus | 2 | Dentate;Substantia_Nigra | 144 | 96 | 3 | 2 | rs56035021;rs45610037;rs17454584 | Pleiotropic | Novel | Novel | Novel |
| 37 | 4:159650783:A:G;4:159777764:A:C;<br>4:159685743:C:T;4:159674887:C:T;<br>4:159689895: ... | 4 | 159621185 | 159944851 | 323.7 | rs13135442 | 3.1e-15 | Dentate nucleus | 5 | Caudate;Dentate;Globus_Pallidus;Putamen;<br>Red_Nucleus | 1080 | 746 | 10 | 5 | rs58093123;rs7663520;rs4600878;<br>rs2068625;rs11100197;rs35782846;<br>rs9784569;rs1 ... | Pleiotropic | Replicated | Novel | Replicated (Casanova) |
| 38 | 4:185710838:C:T;4:185704176:G:T;<br>4:185688754:C:T | 4 | 185673134 | 185731940 | 58.8 | rs4862415 | 1.3e-13 | Dentate nucleus | 4 | Dentate;Globus_Pallidus;Red_Nucleus;<br>Substantia_Nigra | 71 | 56 | 8 | 4 | rs7659090;rs2046814;rs2046814;rs7659090;<br>rs4862415;rs2046814;rs13137179;rs1312 ... | Pleiotropic | Replicated | Novel | Replicated (Casanova) |
| 39 | 5:140884527:C:T | 5 | 140797630 | 141016288 | 218.7 | rs9324851 | 2e-09 | Dentate nucleus | 1 | Dentate | 42 | 26 | 2 | 1 | rs9324851;rs3844598 | Region-specific | Replicated | Novel | Replicated (Casanova) |
| 40 | 6:26093141:A:G;6:26098474:C:T;<br>6:26123502:C:T | 6 | 25075717 | 29603136 | 4527.4 | rs115740542 | 6.3e-111 | Dentate nucleus | 6 | Caudate;Dentate;Globus_Pallidus;Putamen;<br>Red_Nucleus;Substantia_Nigra | 7641 | 5584 | 333 | 74 | rs12196438;rs9467478;rs75580845;<br>rs28393611;rs78151190;rs62393683;<br>rs72838866;rs93 ... | Pan-regional | Replicated | Novel | Replicated (Casanova) |

| Locus ID | Unique variant ID | Chromosome | Start (bp) | End (bp) | Width (kb) | Top SNP | Top P | Top region | Number of regions | Regions | Total SNPs | Genome-wide significant SNPs | Independent significant SNPs | Lead SNPs | Independent SNPs | Pleiotropy | Replication (Casanova) | Replication (Wang) | Overall status |
|---|---|---|---|---|---|---|---|---|---|---|---|---|---|---|---|---|---|---|---|
| 41 | 6:45420847:C:T;6:45447558:A:G | 6 | 45407654 | 45519382 | 111.7 | rs2677100 | 2.9e-18 | Putamen | 2 | Dentate;Putamen | 171 | 123 | 5 | 2 | rs2677100;rs2396441;rs6930053;rs4714858; rs10214519 | Pleiotropic | Replicated | Novel | Replicated (Casanova) |
| 42 | 6:108390978:A:G | 6 | 108335345 | 108411010 | 75.7 | rs956208 | 4.2e-15 | Dentate nucleus | 1 | Dentate | 154 | 111 | 3 | 1 | rs956208;rs7749081;rs11153101 | Region-specific | Novel | Novel | Novel |
| 43 | 6:126683594:A:G | 6 | 126659043 | 127080700 | 421.7 | rs9398803 | 4.3e-08 | Caudate nucleus | 1 | Caudate | 248 | 173 | 1 | 1 | rs9398803 | Region-specific | Novel | Novel | Novel |
| 44 | 7:1563046:C:T;7:1574812:C:T | 7 | 1550455 | 1595839 | 45.4 | rs4720836 | 3e-18 | Putamen | 3 | Caudate;Globus_Pallidus;Putamen | 395 | 345 | 5 | 3 | rs6463895;rs4720836;rs6942779; rs11981930;rs4720836 | Pleiotropic | Replicated | Novel | Replicated (Casanova) |
| 45 | 7:20394948:C:T | 7 | 20371273 | 20442863 | 71.6 | rs73086586 | 4e-11 | Putamen | 1 | Putamen | 11 | 8 | 1 | 1 | rs73086586 | Region-specific | Replicated | Novel | Replicated (Casanova) |
| 46 | 7:22301192:C:T | 7 | 22292706 | 22337720 | 45 | rs2686473 | 1.6e-11 | Dentate nucleus | 1 | Dentate | 32 | 22 | 2 | 1 | rs2686473;rs10225069 | Region-specific | Novel | Novel | Novel |
| 47 | 7:72977249:C:T;7:73042614:A:G; 7:73495390:C:T;7:73007606:C:T; 7:72984220:C:T | 7 | 72308916 | 73537628 | 1228.7 | rs7786376 | 1.7e-36 | Putamen | 4 | Caudate;Dentate;Globus_Pallidus;Putamen | 1177 | 904 | 40 | 11 | rs34594435;rs57978469;rs799169; rs56377543;rs4717763;rs62465131;rs42238; rs1411361 ... | Pleiotropic | Replicated | Novel | Replicated (Casanova) |
| 48 | 7:97967556:A:C | 7 | 97845713 | 98039459 | 193.7 | rs12671301 | 2.8e-08 | Globus pallidus | 1 | Globus_Pallidus | 249 | 185 | 1 | 1 | rs12671301 | Region-specific | Novel | Novel | Novel |
| 49 | 7:100456595:A:G;7:99835613:G:T | 7 | 99527977 | 100528223 | 1000.2 | rs62482175 | 1.5e-09 | Dentate nucleus | 2 | Dentate;Putamen | 105 | 79 | 4 | 2 | rs314373;rs62482175;rs13227444; rs10231649 | Pleiotropic | Replicated | Novel | Replicated (Casanova) |
| 50 | 8:23417091:A:G;8:23415655:C:T; 8:22279863:A:C;8:23382985:A:G; 8:22271995:G:T ... | 8 | 22112929 | 24779437 | 2666.5 | rs11778179 | 3e-29 | Putamen | 4 | Caudate;Dentate;Putamen;Red_Nucleus | 1597 | 1180 | 72 | 13 | rs4260902;rs2048528;rs36104352;rs721183; rs201643157;rs11782783;rs28656888; rs7813 ... | Pleiotropic | Replicated | Novel | Replicated (Casanova) |
| 51 | 8:27103024:A:G;8:27104727:A:G | 8 | 27075395 | 27119538 | 44.1 | rs10481349 | 1.1e-12 | Putamen | 3 | Caudate;Dentate;Putamen | 44 | 39 | 5 | 3 | rs17366947;rs10481349;rs11990926; rs879403;rs10481349 | Pleiotropic | Replicated | Novel | Replicated (Casanova) |
| 52 | 8:42428852:A:G | 8 | 42351176 | 42457590 | 106.4 | rs35295075 | 2.1e-09 | Globus pallidus | 1 | Globus_Pallidus | 184 | 142 | 1 | 1 | rs35295075 | Region-specific | Replicated | Novel | Replicated (Casanova) |
| 53 | 8:101676675:A:C;8:101679224:G:T | 8 | 101668920 | 101760211 | 91.3 | rs2978098 | 1.2e-33 | Globus pallidus | 3 | Globus_Pallidus;Red_Nucleus; Substantia_Nigra | 319 | 215 | 21 | 3 | rs2978098;rs1693566;rs879391;rs1693565; rs1786349;rs7002825;rs1693546;rs2935545 ... | Pleiotropic | Replicated | Novel | Replicated (Casanova) |
| 54 | 8:131213570:A:G | 8 | 131027338 | 131361477 | 334.1 | rs4733775 | 5.7e-12 | Dentate nucleus | 1 | Dentate | 137 | 99 | 3 | 1 | rs4733775;rs6470799;rs10956511 | Region-specific | Replicated | Novel | Replicated (Casanova) |
| 55 | 8:142005245:C:T | 8 | 141645267 | 142018581 | 373.3 | rs306960 | 1.5e-08 | Dentate nucleus | 1 | Dentate | 400 | 280 | 1 | 1 | rs306960 | Region-specific | Novel | Novel | Novel |
| 56 | 9:19210177:A:G | 9 | 19182872 | 19378074 | 195.2 | rs465423 | 8.1e-31 | Putamen | 6 | Caudate;Dentate;Globus_Pallidus;Putamen; Red_Nucleus;Substantia_Nigra | 89 | 61 | 12 | 6 | rs465423;rs62560298;rs465423;rs62560298; rs7855666;rs465423;rs465423;rs6256029 ... | Pan-regional | Replicated | Novel | Replicated (Casanova) |
| 57 | 9:32448436:A:G | 9 | 32347719 | 32579454 | 231.7 | rs10970982 | 6.6e-15 | Caudate nucleus | 1 | Caudate | 148 | 104 | 7 | 3 | rs1023087;rs10970982;rs13289402; rs75438929;rs7043497;rs4879586; rs4487862 | Region-specific | Replicated | Novel | Replicated (Casanova) |
| 58 | 9:109112718:C:T;9:109137188:C:T | 9 | 108838212 | 109151678 | 313.5 | rs117197034 | 2.3e-21 | Substantia nigra | 2 | Red_Nucleus;Substantia_Nigra | 267 | 178 | 7 | 2 | rs117197034;rs57617805;rs75089243; rs117532980;rs1484374;rs76289637; rs75089243 | Pleiotropic | Replicated | Novel | Replicated (Casanova) |
| 59 | 9:125694309:C:T;9:125550272:C:T | 9 | 125266929 | 125957759 | 690.8 | rs542119549 | 7.5e-14 | Dentate nucleus | 3 | Dentate;Globus_Pallidus;Putamen | 18 | 14 | 13 | 7 | rs542119549;rs138036195;rs143663770; rs138036195;rs16911860;rs149587545; rs14223 ... | Pleiotropic | Replicated | Novel | Replicated (Casanova) |
| 60 | 9:136142203:A:C | 9 | 136132908 | 136149500 | 16.6 | rs514659 | 1.3e-09 | Red nucleus | 1 | Red_Nucleus | 48 | 32 | 1 | 1 | rs514659 | Region-specific | Novel | Novel | Novel |
| 61 | 10:18221908:C:T;10:18243703:A:G; 10:18242311:A:G;10:18239344:G:T; 10:18240466: ... | 10 | 17451731 | 19011804 | 1560.1 | rs4348791 | 5.9e-125 | Caudate nucleus | 6 | Caudate;Dentate;Globus_Pallidus;Putamen; Red_Nucleus;Substantia_Nigra | 2348 | 1639 | 259 | 77 | rs530434517;rs555045010;rs2436671; rs367834034;rs201843067;rs149410244; rs1926732 ... | Pan-regional | Replicated | Replicated | Replicated (both) |
| 62 | 10:49822099:A:G | 10 | 49818441 | 49860258 | 41.8 | rs4114458 | 4.5e-09 | Globus pallidus | 1 | Globus_Pallidus | 35 | 24 | 1 | 1 | rs4114458 | Region-specific | Novel | Novel | Novel |
| 63 | 10:51565548:C:T;10:51563993:A:G | 10 | 51563700 | 51649753 | 86.1 | rs10994675 | 3.2e-08 | Red nucleus | 2 | Red_Nucleus;Substantia_Nigra | 12 | 8 | 2 | 2 | rs7909927;rs10994675 | Pleiotropic | Replicated | Novel | Replicated (Casanova) |
| 64 | 10:104697781:C:T | 10 | 104565199 | 105176914 | 611.7 | rs12414777 | 4.5e-14 | Dentate nucleus | 3 | Dentate;Globus_Pallidus;Red_Nucleus | 542 | 394 | 9 | 5 | rs12414777;rs12414777;rs11191574; rs7082288;rs79780963;rs12414777; rs11191514;rs ... | Pleiotropic | Replicated | Novel | Replicated (Casanova) |
| 65 | 10:116237760:C:T;10:116252408:C:T; 10:116255019:G:T | 10 | 116228747 | 116360503 | 131.8 | rs11196758 | 9.3e-23 | Putamen | 6 | Caudate;Dentate;Globus_Pallidus;Putamen; Red_Nucleus;Substantia_Nigra | 227 | 184 | 14 | 7 | rs11196758;rs11196758;rs2475232; rs56339171;rs4255462;rs2483569; rs2475241;rs111 ... | Pan-regional | Replicated | Replicated | Replicated (both) |
| 66 | 11:970503:C:T;11:526478:A:G | 11 | 414416 | 991109 | 576.7 | rs35333170 | 3.2e-13 | Dentate nucleus | 2 | Dentate;Globus_Pallidus | 410 | 315 | 7 | 2 | rs7395773;rs35333170;rs1870726; rs2396545;rs61879160;rs10794323; rs61742833 | Pleiotropic | Novel | Novel | Novel |
| 67 | 11:9287095:C:T | 11 | 9130147 | 9343781 | 213.6 | rs61877916 | 2.7e-08 | Dentate nucleus | 1 | Dentate | 83 | 57 | 1 | 1 | rs61877916 | Region-specific | Novel | Novel | Novel |
| 68 | 11:30510599:C:T | 11 | 30492070 | 30569363 | 77.3 | rs35827570 | 1.6e-12 | Putamen | 1 | Putamen | 41 | 29 | 1 | 1 | rs35827570 | Region-specific | Replicated | Novel | Replicated (Casanova) |
| 69 | 11:44635208:A:G;11:44636833:A:G | 11 | 44619040 | 44636833 | 17.8 | rs2303865 | 4.9e-10 | Dentate nucleus | 2 | Dentate;Red_Nucleus | 17 | 11 | 3 | 2 | rs7930904;rs2303865;rs2303865 | Pleiotropic | Replicated | Replicated | Replicated (both) |
| 70 | 11:47434986:A:G;11:47581443:A:G | 11 | 47229316 | 47943419 | 714.1 | rs11039297 | 4.8e-11 | Globus pallidus | 2 | Globus_Pallidus;Putamen | 416 | 296 | 7 | 2 | rs2293576;rs11039409;rs10838777; rs11039297;rs7944419;rs71475909; rs3740688 | Pleiotropic | Replicated | Novel | Replicated (Casanova) |
| 71 | 11:61769588:C:T;11:61744342:C:T; 11:61738545:G:T;11:62523559:A:G | 11 | 61710538 | 62523559 | 813 | rs12799017 | 6.1e-45 | Red nucleus | 4 | Dentate;Globus_Pallidus;Red_Nucleus; Substantia_Nigra | 368 | 276 | 25 | 9 | rs2009875;rs4963279;rs77133046; rs2727269;rs12288004;rs907638; rs112691937;rs57280 ... | Pleiotropic | Replicated | Replicated | Replicated (both) |
| 72 | 11:63587117:C:T;11:63615554:C:T | 11 | 63585804 | 63797679 | 211.9 | rs10897452 | 3.7e-16 | Dentate nucleus | 2 | Dentate;Putamen | 268 | 192 | 8 | 2 | rs630486;rs10897452;rs11231629; rs145192830;rs3832717;rs568930;rs643634; rs496343 ... | Pleiotropic | Replicated | Novel | Replicated (Casanova) |
| 73 | 11:73460030:C:T | 11 | 73460030 | 73572495 | 112.5 | rs113450689 | 4.9e-08 | Dentate nucleus | 1 | Dentate | 2 | 2 | 1 | 1 | rs113450689 | Region-specific | Novel | Novel | Novel |
| 74 | 11:85800279:A:G;11:85809825:C:T | 11 | 85652251 | 85868640 | 216.4 | rs561655 | 4.7e-09 | Putamen | 2 | Dentate;Putamen | 282 | 200 | 5 | 2 | rs561655;rs592314;rs645293;rs629343; rs4944560 | Pleiotropic | Replicated | Novel | Replicated (Casanova) |
| 75 | 11:109936687:G:T;11:109940267:A:G; 11:109943972:C:T | 11 | 109936687 | 109953618 | 16.9 | rs10891050 | 3.8e-18 | Red nucleus | 5 | Caudate;Dentate;Putamen;Red_Nucleus; Substantia_Nigra | 75 | 55 | 5 | 5 | rs4027098;rs10891050;rs7113219 | Pleiotropic | Replicated | Replicated | Replicated (both) |
| 76 | 11:128531739:A:C | 11 | 128525423 | 128555379 | 30 | rs694438 | 1.9e-08 | Putamen | 1 | Putamen | 9 | 7 | 1 | 1 | rs694438 | Region-specific | Replicated | Novel | Replicated (Casanova) |
| 77 | 12:26507016:A:G;12:27425366:C:T | 12 | 26310234 | 27483318 | 1173.1 | rs10842717 | 4.2e-11 | Putamen | 2 | Dentate;Putamen | 63 | 45 | 7 | 4 | rs11048420;rs9804706;rs76829083; rs10842718;rs10842717;rs11048473; rs10771328 | Pleiotropic | Replicated | Replicated | Replicated (both) |
| 78 | 12:51484646:C:T;12:51454786:C:T; 12:51335777:G:T;12:51357542:A:G | 12 | 51223579 | 51581740 | 358.2 | rs7979705 | 2.8e-40 | Putamen | 6 | Caudate;Dentate;Globus_Pallidus;Putamen; Red_Nucleus;Substantia_Nigra | 2688 | 1938 | 44 | 13 | rs11169664;rs7980874;rs7979705;rs224567; rs4768952;rs1112392;rs11169664;rs713505 ... | Pan-regional | Replicated | Replicated | Replicated (both) |

| Locus ID | Unique variant ID | Chromosome | Start (bp) | End (bp) | Width (kb) | Top SNP | Top P | Top region | Number of regions | Regions | Total SNPs | Genome-wide significant SNPs | Independent significant SNPs | Lead SNPs | Independent SNPs | Pleiotropy | Replication (Casanova) | Replication (Wang) | Overall status |
|---|---|---|---|---|---|---|---|---|---|---|---|---|---|---|---|---|---|---|---|
| 79 | 12:113583863:A:C;12:113740871:A:G;12:113661258:G:T | 12 | 113536863 | 113752802 | 215.9 | rs77625177 | 2.6e-17 | Putamen | 3 | Caudate;Putamen;Red_Nucleus | 743 | 563 | 10 | 3 | rs146487929;rs61943591;rs77625177;rs73208705;rs77625177;rs75426961;rs146487929 ... | Pleiotropic | Replicated | Replicated | Replicated (both) |
| 80 | 12:123751339:A:G | 12 | 123492112 | 123911789 | 419.7 | rs2030401 | 5.1e-09 | Dentate nucleus | 1 | Dentate | 17 | 13 | 1 | 1 | rs2030401 | Region-specific | Novel | Novel | Novel |
| 81 | 12:133780309:G:T | 12 | 133688707 | 133781163 | 92.5 | rs36127550 | 3.6e-08 | Putamen | 1 | Putamen | 4 | 4 | 1 | 1 | rs36127550 | Region-specific | Replicated | Novel | Replicated (Casanova) |
| 82 | 13:21457395:C:T | 13 | 21285496 | 21492997 | 207.5 | rs9506568 | 7.2e-09 | Globus pallidus | 1 | Globus_Pallidus | 123 | 91 | 1 | 1 | rs9506568 | Region-specific | Novel | Novel | Novel |
| 83 | 14:53662014:A:G;14:53827087:C:T | 14 | 53476090 | 53996341 | 520.3 | rs72673671 | 2.6e-15 | Dentate nucleus | 3 | Dentate;Globus_Pallidus;Red_Nucleus | 1080 | 816 | 16 | 5 | rs2552402;rs72673671;rs113034904;rs9796362;rs17253326;rs72673671;rs113034904;r ... | Pleiotropic | Replicated | Novel | Replicated (Casanova) |
| 84 | 14:75108290:C:T;14:76207794:A:G | 14 | 75062398 | 76444469 | 1382.1 | rs1126190 | 9.5e-11 | Dentate nucleus | 2 | Dentate;Substantia_Nigra | 477 | 356 | 6 | 2 | rs12892228;rs11159098;rs1126190;rs999718;rs4252328;rs12887886 | Pleiotropic | Replicated | Novel | Replicated (Casanova) |
| 85 | 15:32145735:C:T | 15 | 32068396 | 32177326 | 108.9 | rs4238558 | 2.3e-11 | Dentate nucleus | 1 | Dentate | 98 | 65 | 2 | 1 | rs4238558;rs34315282 | Region-specific | Novel | Novel | Novel |
| 86 | 15:45388382:C:T | 15 | 45365790 | 45409271 | 43.5 | rs2554451 | 3.9e-09 | Caudate nucleus | 2 | Caudate;Putamen | 69 | 57 | 3 | 2 | rs2554451;rs269859;rs2554451 | Pleiotropic | Replicated | Novel | Replicated (Casanova) |
| 87 | 15:57295579:A:G;15:57114233:A:C;15:57218850:A:G | 15 | 56876088 | 57601813 | 725.7 | rs72749499 | 1.5e-14 | Putamen | 3 | Globus_Pallidus;Putamen;Substantia_Nigra | 444 | 315 | 6 | 3 | rs72749499;rs72736018;rs72744924;rs13379960;rs72747142;rs7178030 | Pleiotropic | Replicated | Replicated | Replicated (both) |
| 88 | 15:79237293:C:T;15:79234470:C:T | 15 | 78714008 | 79257278 | 543.3 | rs2289702 | 2.4e-25 | Putamen | 3 | Caudate;Globus_Pallidus;Putamen | 875 | 666 | 24 | 6 | rs12912022;rs11636753;rs8038920;rs12916769;rs12910090;rs2289702;rs12901830;rs620 ... | Pleiotropic | Replicated | Novel | Replicated (Casanova) |
| 89 | 15:85667983:G:T | 15 | 85602142 | 85668644 | 66.5 | rs11852268 | 3.2e-08 | Dentate nucleus | 1 | Dentate | 19 | 16 | 1 | 1 | rs11852268 | Region-specific | Replicated | Novel | Replicated (Casanova) |
| 90 | 16:1035548:A:G | 16 | 1004554 | 1035548 | 31 | rs3751669 | 2.4e-08 | Dentate nucleus | 1 | Dentate | 5 | 4 | 1 | 1 | rs3751669 | Region-specific | Replicated | Novel | Replicated (Casanova) |
| 91 | 16:57319143:A:C | 16 | 57158679 | 57320113 | 161.4 | rs7187527 | 5.6e-16 | Putamen | 1 | Putamen | 75 | 61 | 4 | 1 | rs7187527;rs7205444;rs4784771;rs113885775 | Region-specific | Replicated | Replicated | Replicated (both) |
| 92 | 16:69622762:A:G | 16 | 69547741 | 70130803 | 583.1 | rs244418 | 5.5e-15 | Dentate nucleus | 1 | Dentate | 346 | 261 | 5 | 1 | rs244418;rs45488899;rs4985451;rs2650522;rs4783718 | Region-specific | Novel | Novel | Novel |
| 93 | 16:79606520:C:T | 16 | 79586108 | 79631946 | 45.8 | rs434626 | 1e-27 | Putamen | 2 | Caudate;Putamen | 138 | 104 | 21 | 3 | rs434626;rs425628;rs30388;rs250164;rs250151;rs250150;rs250148;rs821225;rs434626 ... | Pleiotropic | Replicated | Replicated | Replicated (both) |
| 94 | 16:87235916:A:G;16:87258088:A:G | 16 | 87220694 | 87264534 | 43.8 | rs4240791 | 7e-09 | Globus pallidus | 2 | Dentate;Globus_Pallidus | 147 | 105 | 4 | 2 | rs4240791;rs62056167;rs12711473;rs11640181 | Pleiotropic | Replicated | Novel | Replicated (Casanova) |
| 95 | 16:90170811:A:G;16:90170095:A:G | 16 | 90125822 | 90173553 | 47.7 | rs201497512 | 1.6e-09 | Putamen | 2 | Globus_Pallidus;Putamen | 28 | 28 | 3 | 2 | rs201497512;rs7197795;rs56407236 | Pleiotropic | Replicated | Novel | Replicated (Casanova) |
| 96 | 17:7172609:A:G;17:7127718:C:T;17:7167776:C:T | 17 | 7092704 | 7187123 | 94.4 | rs17671352 | 3.3e-14 | Red nucleus | 3 | Dentate;Putamen;Red_Nucleus | 223 | 159 | 9 | 3 | rs12601936;rs17671352;rs222837;rs113086489;rs2654189;rs188777;rs3826408;rs5750 ... | Pleiotropic | Replicated | Novel | Replicated (Casanova) |
| 97 | 17:18057907:A:G;17:18057301:A:G;17:18199150:A:G | 17 | 18040404 | 18268113 | 227.7 | rs735960 | 2.8e-14 | Red nucleus | 3 | Dentate;Globus_Pallidus;Red_Nucleus | 472 | 359 | 6 | 3 | rs62073602;rs28364628;rs735960;rs2061214;rs8080168;rs8081343 | Pleiotropic | Replicated | Novel | Replicated (Casanova) |
| 98 | 17:19802247:C:T | 17 | 19799698 | 19911649 | 112 | rs9914273 | 1.4e-09 | Red nucleus | 1 | Red_Nucleus | 58 | 44 | 1 | 1 | rs9914273 | Region-specific | Novel | Novel | Novel |
| 99 | 17:27678484:G:T | 17 | 27660490 | 27881366 | 220.9 | rs60018793 | 1.7e-09 | Putamen | 1 | Putamen | 85 | 65 | 1 | 1 | rs60018793 | Region-specific | Novel | Novel | Novel |
| 100 | 17:40716235:C:T;17:42989063:C:T;17:40741013:C:T;17:42281282:G:T;17:44051589: ... | 17 | 40287106 | 44874453 | 4587.3 | rs12951632 | 6.1e-49 | Putamen | 4 | Caudate;Dentate;Putamen;Red_Nucleus | 5941 | 4428 | 104 | 15 | rs76899038;rs601999;rs668799;rs1474040;rs12937081;rs76189032;rs1554227;rs7863005 ... | Pleiotropic | Replicated | Replicated | Replicated (both) |
| 101 | 17:57609588:C:T;17:57696251:A:G | 17 | 57556414 | 58226089 | 669.7 | rs1881442 | 1.5e-18 | Putamen | 2 | Caudate;Putamen | 676 | 467 | 14 | 2 | rs8073905;rs1881442;rs8069123;rs10853015;rs4968391;rs11654074;rs150581172;rs180 ... | Pleiotropic | Replicated | Replicated | Replicated (both) |
| 102 | 17:62284510:A:G | 17 | 62274324 | 62318545 | 44.2 | rs8072646 | 8.9e-09 | Red nucleus | 1 | Red_Nucleus | 9 | 7 | 1 | 1 | rs8072646 | Region-specific | Novel | Novel | Novel |
| 103 | 17:66282050:A:G | 17 | 66244088 | 66301495 | 57.4 | rs2041130 | 2e-11 | Dentate nucleus | 1 | Dentate | 42 | 35 | 2 | 1 | rs2041130;rs8080714 | Region-specific | Replicated | Novel | Replicated (Casanova) |
| 104 | 17:73935233:G:T;17:73810674:A:C | 17 | 73761839 | 73974293 | 212.5 | rs9913385 | 1.2e-09 | Dentate nucleus | 2 | Dentate;Red_Nucleus | 72 | 55 | 4 | 2 | rs73355708;rs9913385;rs112253169;rs56936829 | Pleiotropic | Replicated | Replicated | Replicated (both) |
| 105 | 18:3017136:C:T | 18 | 2944809 | 3017136 | 72.3 | rs4798054 | 7.3e-09 | Dentate nucleus | 1 | Dentate | 3 | 3 | 1 | 1 | rs4798054 | Region-specific | Novel | Novel | Novel |
| 106 | 19:5908163:C:T | 19 | 5872782 | 5999684 | 126.9 | rs146033453 | 6.9e-14 | Dentate nucleus | 1 | Dentate | 44 | 34 | 3 | 1 | rs146033453;rs274786;rs75566110 | Region-specific | Novel | Novel | Novel |
| 107 | 19:14573327:A:G | 19 | 14557821 | 14592476 | 34.7 | rs8105847 | 1.3e-20 | Dentate nucleus | 1 | Dentate | 44 | 34 | 8 | 1 | rs8105847;rs3760702;rs1127307;rs2241357;rs2420536;rs11673475;rs12974686;rs382675 ... | Region-specific | Novel | Novel | Novel |
| 108 | 19:17596144:A:G | 19 | 17580707 | 17611261 | 30.6 | rs62126783 | 1.2e-08 | Dentate nucleus | 1 | Dentate | 31 | 25 | 1 | 1 | rs62126783 | Region-specific | Novel | Novel | Novel |
| 109 | 19:36204831:C:T | 19 | 36094205 | 36287869 | 193.7 | rs23595 | 3.6e-13 | Dentate nucleus | 1 | Dentate | 88 | 67 | 8 | 1 | rs23595;rs23595;rs807479;rs807479;rs7254601;rs10401695;rs10401695;rs23595 | Region-specific | Replicated | Novel | Replicated (Casanova) |
| 110 | 19:46464137:A:G | 19 | 46459728 | 46489105 | 29.4 | rs57398989 | 4.5e-09 | Caudate nucleus | 1 | Caudate | 14 | 10 | 1 | 1 | rs57398989 | Region-specific | Novel | Novel | Novel |
| 111 | 19:53617818:C:T | 19 | 53608032 | 53621561 | 13.5 | rs12985137 | 3.6e-08 | Dentate nucleus | 1 | Dentate | 42 | 31 | 1 | 1 | rs12985137 | Region-specific | Novel | Novel | Novel |
| 112 | 20:3971079:G:T;20:3923758:C:T;20:3996208:C:T;20:4026818:A:G | 20 | 3840539 | 4051202 | 210.7 | rs6037730 | 1.3e-21 | Dentate nucleus | 4 | Dentate;Globus_Pallidus;Red_Nucleus;Substantia_Nigra | 560 | 419 | 21 | 7 | rs117255962;rs6084521;rs6052248;rs6116153;rs6052246;rs8125729;rs6116164;rs1190 ... | Pleiotropic | Replicated | Replicated | Replicated (both) |
| 113 | 20:25238173:C:T | 20 | 25177920 | 25991371 | 813.5 | rs6132824 | 6.7e-16 | Dentate nucleus | 1 | Dentate | 915 | 677 | 12 | 1 | rs6132824;rs2227891;rs6076335;rs2500406;rs6115218;rs4813576;rs6037243;rs6107130 ... | Region-specific | Novel | Novel | Novel |
| 114 | 21:16798091:G:T | 21 | 16781136 | 16838662 | 57.5 | rs2403966 | 1.9e-09 | Putamen | 1 | Putamen | 91 | 67 | 2 | 1 | rs2403966;rs2823286 | Region-specific | Replicated | Replicated | Replicated (both) |
| 115 | 21:40292946:A:G | 21 | 40289167 | 40316902 | 27.7 | rs7278605 | 1e-13 | Putamen | 1 | Putamen | 41 | 31 | 3 | 1 | rs7278605;rs13053080;rs6517487 | Region-specific | Replicated | Replicated | Replicated (both) |
| 116 | 22:37445668:C:T;22:37462936:A:G | 22 | 37377094 | 37508507 | 131.4 | rs855791 | 1.8e-33 | Dentate nucleus | 6 | Caudate;Dentate;Globus_Pallidus;Putamen;Red_Nucleus;Substantia_Nigra | 482 | 391 | 39 | 10 | rs9610638;rs5756491;rs2235320;rs56270606;rs5756491;rs5750373;rs135167;rs855791 ... | Pan-regional | Replicated | Replicated | Replicated (both) |

*Abbreviations: Chr = chromosome; SNP = single-nucleotide polymorphism; uniqID = unique variant identifier. Loci within 500 kb of one another across regions were merged. A locus was counted as replicated where it overlapped a +/-500 kb window around a variant reported in either prior quantitative susceptibility mapping analysis. Positions are given on GRCh37. Long variant and gene lists are abbreviated in this document; complete values are given in the accompanying spreadsheet workbook.*

### Supplementary Table 3. Polygenic risk score associations with quantitative susceptibility mapping-derived brain iron across same-region and cross-regional subcortical pairs in an independent cohort (Prospective Imaging Study of Ageing; n = 268).

| Polygenic score region | Outcome region | Comparison | Beta | SE | 95% CI lower | 95% CI upper | P | Bonferroni-adjusted P | FDR q | Incremental R2 | n |
|---|---|---|---|---|---|---|---|---|---|---|---|
| Caudate nucleus | Caudate nucleus | Same region | 5.1763 | 0.5986 | 4.0031 | 6.3494 | 6.14e-16 | 2.21e-14 | 7.37e-15 | 0.2213 | 268 |
| Caudate nucleus | Dentate nucleus | Cross region | 5.9872 | 1.5672 | 2.9154 | 9.0589 | 0.0001678 | 0.006041 | 0.0001949 | 0.0479 | 268 |
| Caudate nucleus | Globus pallidus | Cross region | 5.9866 | 1.5136 | 3.02 | 8.9531 | 9.93e-05 | 0.003575 | 0.0001233 | 0.0535 | 268 |
| Caudate nucleus | Putamen | Cross region | 6.0411 | 1.0288 | 4.0247 | 8.0576 | 1.35e-08 | 4.85e-07 | 3.73e-08 | 0.1095 | 268 |
| Caudate nucleus | Red nucleus | Cross region | 4.7216 | 1.4384 | 1.9024 | 7.5409 | 0.001174 | 0.04225 | 0.00128 | 0.0375 | 268 |
| Caudate nucleus | Substantia nigra | Cross region | 5.3691 | 1.4924 | 2.444 | 8.2942 | 0.0003861 | 0.0139 | 0.0004344 | 0.0464 | 268 |
| Dentate nucleus | Caudate nucleus | Cross region | 3.3829 | 0.5903 | 2.2258 | 4.54 | 2.84e-08 | 1.02e-06 | 6.39e-08 | 0.1114 | 268 |
| Dentate nucleus | Dentate nucleus | Same region | 12.707 | 1.2512 | 10.2547 | 15.1593 | 1.51e-20 | 5.43e-19 | 5.43e-19 | 0.2541 | 268 |
| Dentate nucleus | Globus pallidus | Cross region | 6.586 | 1.3755 | 3.89 | 9.2819 | 2.87e-06 | 0.0001032 | 4.13e-06 | 0.0763 | 268 |
| Dentate nucleus | Putamen | Cross region | 6.4189 | 0.9259 | 4.6041 | 8.2336 | 3.43e-11 | 1.23e-09 | 1.76e-10 | 0.1457 | 268 |
| Dentate nucleus | Red nucleus | Cross region | 8.8432 | 1.2331 | 6.4262 | 11.2602 | 8.17e-12 | 2.94e-10 | 4.9e-11 | 0.155 | 268 |
| Dentate nucleus | Substantia nigra | Cross region | 5.7914 | 1.3614 | 3.1231 | 8.4597 | 2.96e-05 | 0.001065 | 3.94e-05 | 0.0636 | 268 |
| Globus pallidus | Caudate nucleus | Cross region | 4.144 | 0.6476 | 2.8747 | 5.4132 | 7.52e-10 | 2.71e-08 | 3.39e-09 | 0.1351 | 268 |
| Globus pallidus | Dentate nucleus | Cross region | 8.2712 | 1.5673 | 5.1992 | 11.3431 | 2.82e-07 | 1.02e-05 | 5.34e-07 | 0.087 | 268 |
| Globus pallidus | Globus pallidus | Same region | 9.3074 | 1.4869 | 6.3931 | 12.2218 | 1.64e-09 | 5.91e-08 | 5.38e-09 | 0.1232 | 268 |
| Globus pallidus | Putamen | Cross region | 6.6238 | 1.0436 | 4.5785 | 8.6692 | 1.01e-09 | 3.62e-08 | 4.02e-09 | 0.1254 | 268 |
| Globus pallidus | Red nucleus | Cross region | 6.2883 | 1.4519 | 3.4427 | 9.134 | 2.14e-05 | 0.0007703 | 2.96e-05 | 0.0633 | 268 |
| Globus pallidus | Substantia nigra | Cross region | 6.4027 | 1.5151 | 3.4331 | 9.3722 | 3.32e-05 | 0.001197 | 4.27e-05 | 0.0629 | 268 |
| Putamen | Caudate nucleus | Cross region | 4.8215 | 0.5957 | 3.6538 | 5.9891 | 2.45e-14 | 8.81e-13 | 2.2e-13 | 0.1995 | 268 |
| Putamen | Dentate nucleus | Cross region | 7.9932 | 1.4993 | 5.0545 | 10.9319 | 2.16e-07 | 7.78e-06 | 4.32e-07 | 0.0886 | 268 |
| Putamen | Globus pallidus | Cross region | 5.731 | 1.4871 | 2.8162 | 8.6458 | 0.0001476 | 0.005312 | 0.0001771 | 0.0509 | 268 |
| Putamen | Putamen | Same region | 7.6921 | 0.9611 | 5.8083 | 9.5759 | 4.39e-14 | 1.58e-12 | 3.16e-13 | 0.1844 | 268 |
| Putamen | Red nucleus | Cross region | 7.8578 | 1.3536 | 5.2047 | 10.5108 | 1.92e-08 | 6.91e-07 | 4.94e-08 | 0.1079 | 268 |
| Putamen | Substantia nigra | Cross region | 4.383 | 1.4757 | 1.4907 | 7.2753 | 0.003262 | 0.1174 | 0.003454 | 0.0321 | 268 |
| Red nucleus | Caudate nucleus | Cross region | 3.7383 | 0.6047 | 2.5532 | 4.9235 | 2.52e-09 | 9.05e-08 | 7.55e-09 | 0.1273 | 268 |
| Red nucleus | Dentate nucleus | Cross region | 7.6048 | 1.4584 | 4.7464 | 10.4632 | 3.83e-07 | 1.38e-05 | 6.9e-07 | 0.0852 | 268 |
| Red nucleus | Globus pallidus | Cross region | 3.597 | 1.4678 | 0.7202 | 6.4739 | 0.01494 | 0.5377 | 0.01494 | 0.0213 | 268 |
| Red nucleus | Putamen | Cross region | 6.0979 | 0.9713 | 4.1941 | 8.0017 | 1.48e-09 | 5.34e-08 | 5.34e-09 | 0.1231 | 268 |
| Red nucleus | Red nucleus | Same region | 10.9546 | 1.2172 | 8.5689 | 13.3402 | 5.53e-17 | 1.99e-15 | 9.96e-16 | 0.2225 | 268 |
| Red nucleus | Substantia nigra | Cross region | 3.7427 | 1.4378 | 0.9245 | 6.5608 | 0.009787 | 0.3523 | 0.01007 | 0.0249 | 268 |
| Substantia nigra | Caudate nucleus | Cross region | 3.7215 | 0.6472 | 2.453 | 4.9901 | 2.56e-08 | 9.23e-07 | 6.15e-08 | 0.1121 | 268 |
| Substantia nigra | Dentate nucleus | Cross region | 7.5864 | 1.5567 | 4.5352 | 10.6375 | 1.94e-06 | 6.98e-05 | 2.91e-06 | 0.0753 | 268 |
| Substantia nigra | Globus pallidus | Cross region | 7.378 | 1.5056 | 4.427 | 10.329 | 1.71e-06 | 6.16e-05 | 2.68e-06 | 0.0796 | 268 |
| Substantia nigra | Putamen | Cross region | 5.8672 | 1.0446 | 3.8198 | 7.9145 | 5.11e-08 | 1.84e-06 | 1.08e-07 | 0.1012 | 268 |
| Substantia nigra | Red nucleus | Cross region | 7.1652 | 1.4135 | 4.3946 | 9.9357 | 7.73e-07 | 2.78e-05 | 1.27e-06 | 0.0846 | 268 |
| Substantia nigra | Substantia nigra | Same region | 7.5211 | 1.4715 | 4.6369 | 10.4052 | 6.32e-07 | 2.27e-05 | 1.08e-06 | 0.0892 | 268 |

*Abbreviations: CI = confidence interval; FDR = false discovery rate; PRS = polygenic risk score; QSM = quantitative susceptibility mapping; R2 incremental = variance explained by the polygenic score over a covariate-only model. The Bonferroni threshold was P < 1.39 x 10-3 (0.05/36).*

**Supplementary Table 4. Significant genes from MAGMA gene-based analysis of quantitative susceptibility mapping-derived brain iron across six subcortical regions (Bonferroni-corrected P < 2.59 x 10-6).**

| Trait | Gene symbol | Ensembl gene ID | Chromosome | Start (bp) | Stop (bp) | SNPs (n) | Z | P | Bonferroni-adjusted P |
|---|---|---|---|---|---|---|---|---|---|
| Caudate nucleus | BANK1 | ENSG00000153064 | 4 | 102297443 | 103005969 | 1655 | 10.29 | 3.9e-25 | 7.54e-21 |
| Caudate nucleus | CACNB2 | ENSG00000165995 | 10 | 18394606 | 18840798 | 1507 | 10.15 | 1.69e-24 | 3.27e-20 |
| Caudate nucleus | SLC39A8 | ENSG00000138821 | 4 | 103162198 | 103387415 | 653 | 8.806 | 6.46e-19 | 1.25e-14 |
| Caudate nucleus | MRC1 | ENSG00000120586 | 10 | 18063352 | 18210091 | 7 | 7.941 | 9.99e-16 | 1.93e-11 |
| Caudate nucleus | SLC25A37 | ENSG00000147454 | 8 | 23351318 | 23442976 | 425 | 7.855 | 2e-15 | 3.87e-11 |
| Caudate nucleus | SLC39A12 | ENSG00000148482 | 10 | 18205768 | 18342221 | 469 | 7.727 | 5.52e-15 | 1.07e-10 |
| Caudate nucleus | PTRF | ENSG00000177469 | 17 | 40544470 | 40610535 | 170 | 7.673 | 8.38e-15 | 1.62e-10 |
| Caudate nucleus | HIST1H2AB | ENSG00000137259 | 6 | 26023320 | 26068796 | 82 | 7.664 | 9.05e-15 | 1.75e-10 |
| Caudate nucleus | TUBG1 | ENSG00000131462 | 17 | 40726694 | 40777252 | 78 | 7.602 | 1.45e-14 | 2.81e-10 |
| Caudate nucleus | HIST1H1C | ENSG00000187837 | 6 | 26045968 | 26091699 | 101 | 7.594 | 1.55e-14 | 2.99e-10 |
| Caudate nucleus | TF | ENSG00000091513 | 3 | 133429800 | 133507850 | 256 | 7.503 | 3.13e-14 | 6.04e-10 |
| Caudate nucleus | CCR10 | ENSG00000184451 | 17 | 40820907 | 40870935 | 45 | 7.498 | 3.24e-14 | 6.25e-10 |
| Caudate nucleus | HFE | ENSG00000010704 | 6 | 26052509 | 26108571 | 134 | 7.497 | 3.26e-14 | 6.29e-10 |
| Caudate nucleus | VPS37D | ENSG00000176428 | 7 | 73047155 | 73096442 | 133 | 7.405 | 6.54e-14 | 1.26e-09 |
| Caudate nucleus | SCGN | ENSG00000079689 | 6 | 25617464 | 25712011 | 282 | 7.402 | 6.73e-14 | 1.3e-09 |
| Caudate nucleus | CNTNAP1 | ENSG00000108797 | 17 | 40799631 | 40861832 | 73 | 7.343 | 1.04e-13 | 2.01e-09 |
| Caudate nucleus | HIGD1C | ENSG00000214511 | 12 | 51312705 | 51374289 | 197 | 7.295 | 1.49e-13 | 2.87e-09 |
| Caudate nucleus | SLC11A2 | ENSG00000110911 | 12 | 51363184 | 51457349 | 233 | 7.287 | 1.58e-13 | 3.05e-09 |
| Caudate nucleus | HIST1H3C | ENSG00000196532 | 6 | 26010639 | 26056097 | 90 | 7.285 | 1.61e-13 | 3.12e-09 |
| Caudate nucleus | TBL2 | ENSG00000106638 | 7 | 72973262 | 73028121 | 117 | 7.245 | 2.17e-13 | 4.19e-09 |
| Caudate nucleus | STAT3 | ENSG00000168610 | 17 | 40455342 | 40575586 | 230 | 7.206 | 2.88e-13 | 5.56e-09 |
| Caudate nucleus | LETMD1 | ENSG00000050426 | 12 | 51406745 | 51464207 | 134 | 7.143 | 4.56e-13 | 8.8e-09 |
| Caudate nucleus | TRIM38 | ENSG00000112343 | 6 | 25928030 | 25997384 | 160 | 7.086 | 6.92e-13 | 1.34e-08 |
| Caudate nucleus | HIST1H2BB | ENSG00000196226 | 6 | 26033455 | 26078885 | 94 | 7.071 | 7.69e-13 | 1.49e-08 |
| Caudate nucleus | COASY | ENSG00000068120 | 17 | 40678485 | 40728295 | 102 | 7.066 | 7.97e-13 | 1.54e-08 |
| Caudate nucleus | AOC2 | ENSG00000131480 | 17 | 40961617 | 41012724 | 23 | 7.038 | 9.76e-13 | 1.89e-08 |
| Caudate nucleus | NAGLU | ENSG00000108784 | 17 | 40653190 | 40706467 | 84 | 7.03 | 1.04e-12 | 2e-08 |
| Caudate nucleus | LRRC16A | ENSG00000079691 | 6 | 25244306 | 25630758 | 1409 | 6.986 | 1.42e-12 | 2.74e-08 |
| Caudate nucleus | BECN1 | ENSG00000126581 | 17 | 40952152 | 41020367 | 33 | 6.971 | 1.57e-12 | 3.04e-08 |
| Caudate nucleus | SRPRB | ENSG00000144867 | 3 | 133467877 | 133554616 | 218 | 6.927 | 2.15e-12 | 4.15e-08 |
| Caudate nucleus | MLX | ENSG00000108788 | 17 | 40684086 | 40735257 | 95 | 6.826 | 4.37e-12 | 8.45e-08 |
| Caudate nucleus | SLC17A1 | ENSG00000124568 | 6 | 25773125 | 25867287 | 232 | 6.815 | 4.72e-12 | 9.12e-08 |
| Caudate nucleus | ACO1 | ENSG00000122729 | 9 | 32349618 | 32464767 | 262 | 6.811 | 4.83e-12 | 9.33e-08 |
| Caudate nucleus | AOC3 | ENSG00000131471 | 17 | 40968201 | 41020147 | 24 | 6.715 | 9.39e-12 | 1.81e-07 |
| Caudate nucleus | CSRNP2 | ENSG00000110925 | 12 | 51444990 | 51512447 | 166 | 6.611 | 1.91e-11 | 3.69e-07 |
| Caudate nucleus | SLC17A2 | ENSG00000112337 | 6 | 25902982 | 25965946 | 182 | 6.537 | 3.15e-11 | 6.08e-07 |
| Caudate nucleus | FP15737 | ENSG00000215298 | 8 | 23395157 | 23442974 | 291 | 6.534 | 3.2e-11 | 6.18e-07 |
| Caudate nucleus | RITA1 | ENSG00000139405 | 12 | 113588331 | 113640173 | 87 | 6.533 | 3.22e-11 | 6.22e-07 |
| Caudate nucleus | DDX54 | ENSG00000123064 | 12 | 113584979 | 113658284 | 117 | 6.518 | 3.56e-11 | 6.87e-07 |
| Caudate nucleus | PSME3 | ENSG00000131467 | 17 | 40941402 | 41005774 | 33 | 6.505 | 3.87e-11 | 7.48e-07 |
| Caudate nucleus | IQCD | ENSG00000166578 | 12 | 113623246 | 113693899 | 110 | 6.43 | 6.36e-11 | 1.23e-06 |
| Caudate nucleus | HYKK | ENSG00000188266 | 15 | 78764906 | 78839714 | 161 | 6.427 | 6.5e-11 | 1.25e-06 |
| Caudate nucleus | TPCN1 | ENSG00000186815 | 12 | 113623855 | 113746390 | 208 | 6.402 | 7.67e-11 | 1.48e-06 |
| Caudate nucleus | PSMA4 | ENSG00000041357 | 15 | 78797747 | 78851604 | 127 | 6.39 | 8.32e-11 | 1.61e-06 |
| Caudate nucleus | AC027228.1 | ENSG00000268838 | 15 | 78795023 | 78841288 | 110 | 6.381 | 8.79e-11 | 1.7e-06 |
| Caudate nucleus | CHRNA5 | ENSG00000169684 | 15 | 78822862 | 78897611 | 161 | 6.221 | 2.47e-10 | 4.76e-06 |
| Caudate nucleus | CCDC42B | ENSG00000186710 | 12 | 113552663 | 113607081 | 120 | 6.212 | 2.61e-10 | 5.04e-06 |
| Caudate nucleus | PPID | ENSG00000171497 | 4 | 159620286 | 159679548 | 76 | 6.2 | 2.82e-10 | 5.44e-06 |
| Caudate nucleus | MAFK | ENSG00000198517 | 7 | 1535350 | 1592679 | 213 | 6.188 | 3.04e-10 | 5.87e-06 |

| Trait | Gene symbol | Ensembl gene ID | Chromosome | Start (bp) | Stop (bp) | SNPs (n) | Z | P | Bonferroni-adjusted P |
|---|---|---|---|---|---|---|---|---|---|
| Caudate nucleus | SLC17A3 | ENSG00000124564 | 6 | 25823294 | 25917514 | 233 | 6.109 | 5e-10 | 9.65e-06 |
| Caudate nucleus | HIST1H3A | ENSG00000198366 | 6 | 25985718 | 26031186 | 80 | 6.109 | 5e-10 | 9.65e-06 |
| Caudate nucleus | HIST1H4A | ENSG00000196176 | 6 | 25986907 | 26032278 | 79 | 6.109 | 5e-10 | 9.65e-06 |
| Caudate nucleus | HIST1H1A | ENSG00000124610 | 6 | 26007260 | 26053040 | 93 | 6.109 | 5e-10 | 9.65e-06 |
| Caudate nucleus | HIST1H4B | ENSG00000124529 | 6 | 26017124 | 26062480 | 89 | 6.109 | 5e-10 | 9.65e-06 |
| Caudate nucleus | HIST1H3B | ENSG00000124693 | 6 | 26021817 | 26067288 | 86 | 6.109 | 5e-10 | 9.65e-06 |
| Caudate nucleus | HIST1H4C | ENSG00000197061 | 6 | 26069104 | 26114518 | 128 | 6.109 | 5e-10 | 9.65e-06 |
| Caudate nucleus | FZD9 | ENSG00000188763 | 7 | 72813109 | 72860450 | 69 | 6.109 | 5e-10 | 9.65e-06 |
| Caudate nucleus | BAZ1B | ENSG00000009954 | 7 | 72844728 | 72971608 | 174 | 6.109 | 5e-10 | 9.65e-06 |
| Caudate nucleus | BCL7B | ENSG00000106635 | 7 | 72940686 | 73007332 | 124 | 6.109 | 5e-10 | 9.65e-06 |
| Caudate nucleus | MLXIPL | ENSG00000009950 | 7 | 72997524 | 73073873 | 203 | 6.109 | 5e-10 | 9.65e-06 |
| Caudate nucleus | MRC1L1 | ENSG00000183748 | 10 | 17816362 | 17963178 | 46 | 6.109 | 5e-10 | 9.65e-06 |
| Caudate nucleus | ATP6V0A1 | ENSG00000033627 | 17 | 40575862 | 40684629 | 183 | 6.109 | 5e-10 | 9.65e-06 |
| Caudate nucleus | HSD17B1 | ENSG00000108786 | 17 | 40666232 | 40717231 | 91 | 6.109 | 5e-10 | 9.65e-06 |
| Caudate nucleus | PSMC3IP | ENSG00000131470 | 17 | 40714333 | 40764849 | 75 | 6.109 | 5e-10 | 9.65e-06 |
| Caudate nucleus | FAM134C | ENSG00000141699 | 17 | 40721531 | 40797641 | 129 | 6.109 | 5e-10 | 9.65e-06 |
| Caudate nucleus | TUBG2 | ENSG00000037042 | 17 | 40776323 | 40829024 | 91 | 6.109 | 5e-10 | 9.65e-06 |
| Caudate nucleus | PLEKHH3 | ENSG00000068137 | 17 | 40809932 | 40864048 | 54 | 6.109 | 5e-10 | 9.65e-06 |
| Caudate nucleus | CNTD1 | ENSG00000176563 | 17 | 40915810 | 40973605 | 29 | 6.034 | 7.98e-10 | 1.54e-05 |
| Caudate nucleus | DBI | ENSG00000155368 | 2 | 120089497 | 120140126 | 163 | 5.985 | 1.08e-09 | 2.09e-05 |
| Caudate nucleus | CHRNA3 | ENSG00000080644 | 15 | 78875394 | 78948637 | 155 | 5.944 | 1.39e-09 | 2.68e-05 |
| Caudate nucleus | AC051642.1 | ENSG00000268085 | 8 | 23402287 | 23455651 | 319 | 5.809 | 3.14e-09 | 6.06e-05 |
| Caudate nucleus | KCTD17 | ENSG00000100379 | 22 | 37412779 | 37469430 | 102 | 5.808 | 3.16e-09 | 6.1e-05 |
| Caudate nucleus | METTL7A | ENSG00000185432 | 12 | 51282255 | 51336300 | 120 | 5.776 | 3.82e-09 | 7.37e-05 |
| Caudate nucleus | HIST1H2AC | ENSG00000180573 | 6 | 26089373 | 26149344 | 158 | 5.771 | 3.94e-09 | 7.6e-05 |
| Caudate nucleus | HCRT | ENSG00000161610 | 17 | 40326078 | 40372470 | 30 | 5.764 | 4.1e-09 | 7.91e-05 |
| Caudate nucleus | GHDC | ENSG00000167925 | 17 | 40330817 | 40381531 | 35 | 5.746 | 4.58e-09 | 8.84e-05 |
| Caudate nucleus | TMEM184A | ENSG00000164855 | 7 | 1571871 | 1635457 | 206 | 5.706 | 5.77e-09 | 0.000111 |
| Caudate nucleus | C2orf76 | ENSG00000186132 | 2 | 120049801 | 120159404 | 351 | 5.674 | 6.99e-09 | 0.000135 |
| Caudate nucleus | INTS1 | ENSG00000164880 | 7 | 1499913 | 1580489 | 258 | 5.668 | 7.24e-09 | 0.00014 |
| Caudate nucleus | ABLIM1 | ENSG00000099204 | 10 | 116180872 | 116479762 | 786 | 5.667 | 7.25e-09 | 0.00014 |
| Caudate nucleus | WNK4 | ENSG00000126562 | 17 | 40897696 | 40958954 | 35 | 5.665 | 7.35e-09 | 0.000142 |
| Caudate nucleus | IFI35 | ENSG00000068079 | 17 | 41123742 | 41176473 | 73 | 5.659 | 7.6e-09 | 0.000147 |
| Caudate nucleus | TFRC | ENSG00000072274 | 3 | 195744054 | 195844060 | 428 | 5.632 | 8.93e-09 | 0.000172 |
| Caudate nucleus | DUOX2 | ENSG00000140279 | 15 | 45374848 | 45441542 | 124 | 5.568 | 1.29e-08 | 0.000249 |
| Caudate nucleus | PPP3CA | ENSG00000138814 | 4 | 101934566 | 102304435 | 719 | 5.563 | 1.33e-08 | 0.000256 |
| Caudate nucleus | WBSCR22 | ENSG00000071462 | 7 | 73062355 | 73129491 | 180 | 5.528 | 1.62e-08 | 0.000313 |
| Caudate nucleus | TMPRSS6 | ENSG00000187045 | 22 | 37451476 | 37540603 | 267 | 5.514 | 1.75e-08 | 0.000338 |
| Caudate nucleus | TFCP2 | ENSG00000135457 | 12 | 51477446 | 51601926 | 308 | 5.495 | 1.96e-08 | 0.000378 |
| Caudate nucleus | ENTPD4 | ENSG00000197217 | 8 | 23233296 | 23350208 | 333 | 5.493 | 1.98e-08 | 0.000382 |
| Caudate nucleus | DUOX1 | ENSG00000137857 | 15 | 45387131 | 45467774 | 158 | 5.484 | 2.07e-08 | 0.0004 |
| Caudate nucleus | PTGES3L | ENSG00000267060 | 17 | 41110105 | 41167545 | 81 | 5.468 | 2.28e-08 | 0.00044 |
| Caudate nucleus | RND2 | ENSG00000108830 | 17 | 41142258 | 41194057 | 76 | 5.466 | 2.3e-08 | 0.000444 |
| Caudate nucleus | HIST1H1T | ENSG00000187475 | 6 | 26097640 | 26143364 | 127 | 5.432 | 2.79e-08 | 0.000539 |
| Caudate nucleus | RPL27 | ENSG00000131469 | 17 | 41115290 | 41164976 | 73 | 5.426 | 2.89e-08 | 0.000557 |
| Caudate nucleus | TMEM37 | ENSG00000171227 | 2 | 120152477 | 120206096 | 139 | 5.409 | 3.16e-08 | 0.000611 |
| Caudate nucleus | COA3 | ENSG00000183978 | 17 | 40937165 | 40985722 | 23 | 5.406 | 3.21e-08 | 0.000621 |
| Caudate nucleus | IREB2 | ENSG00000136381 | 15 | 78694773 | 78803798 | 208 | 5.367 | 3.99e-08 | 0.000771 |
| Caudate nucleus | PTGES3L-AARSD1 | ENSG00000108825 | 17 | 41092543 | 41167545 | 95 | 5.352 | 4.36e-08 | 0.000841 |
| Caudate nucleus | DUOXA2 | ENSG00000140274 | 15 | 45371519 | 45420619 | 100 | 5.321 | 5.16e-08 | 0.000996 |
| Caudate nucleus | HIST1H2BA | ENSG00000146047 | 6 | 25692137 | 25737573 | 144 | 5.275 | 6.63e-08 | 0.00128 |
| Caudate nucleus | PHF2 | ENSG00000197724 | 9 | 96303689 | 96451869 | 491 | 5.272 | 6.76e-08 | 0.00131 |

| Trait | Gene symbol | Ensembl gene ID | Chromosome | Start (bp) | Stop (bp) | SNPs (n) | Z | P | Bonferroni-adjusted P |
|---|---|---|---|---|---|---|---|---|---|
| Caudate nucleus | CLTC | ENSG00000141367 | 17 | 57662219 | 57783671 | 166 | 5.268 | 6.89e-08 | 0.00133 |
| Caudate nucleus | TST | ENSG00000128311 | 22 | 37396900 | 37450681 | 164 | 5.259 | 7.26e-08 | 0.0014 |
| Caudate nucleus | MAF | ENSG00000178573 | 16 | 79609740 | 79669611 | 168 | 5.242 | 7.94e-08 | 0.00153 |
| Caudate nucleus | HIST1H2BI | ENSG00000168242 | 6 | 26238144 | 26283622 | 100 | 5.236 | 8.22e-08 | 0.00159 |
| Caudate nucleus | SHMT2 | ENSG00000182199 | 12 | 57588110 | 57638718 | 60 | 5.231 | 8.44e-08 | 0.00163 |
| Caudate nucleus | VPS25 | ENSG00000131475 | 17 | 40890454 | 40941617 | 32 | 5.204 | 9.74e-08 | 0.00188 |
| Caudate nucleus | HIST1H3F | ENSG00000256316 | 6 | 26240370 | 26285835 | 106 | 5.198 | 1.01e-07 | 0.00195 |
| Caudate nucleus | STMN4 | ENSG00000015592 | 8 | 27082840 | 27150937 | 161 | 5.197 | 1.01e-07 | 0.00196 |
| Caudate nucleus | HIST1H4G | ENSG00000124578 | 6 | 26236886 | 26282259 | 102 | 5.192 | 1.04e-07 | 0.00201 |
| Caudate nucleus | DDX58 | ENSG00000107201 | 9 | 32445300 | 32561322 | 242 | 5.19 | 1.05e-07 | 0.00203 |
| Caudate nucleus | NXPH4 | ENSG00000182379 | 12 | 57575578 | 57630232 | 77 | 5.18 | 1.11e-07 | 0.00214 |
| Caudate nucleus | PTRH2 | ENSG00000141378 | 17 | 57741997 | 57819987 | 100 | 5.179 | 1.12e-07 | 0.00215 |
| Caudate nucleus | SCTR | ENSG00000080293 | 2 | 120187419 | 120317070 | 358 | 5.167 | 1.19e-07 | 0.0023 |
| Caudate nucleus | G6PC | ENSG00000131482 | 17 | 41017814 | 41075386 | 78 | 5.096 | 1.73e-07 | 0.00334 |
| Caudate nucleus | HIST1H1E | ENSG00000168298 | 6 | 26121559 | 26167343 | 86 | 5.074 | 1.94e-07 | 0.00375 |
| Caudate nucleus | DHX40 | ENSG00000108406 | 17 | 57607886 | 57695706 | 92 | 5.066 | 2.03e-07 | 0.00392 |
| Caudate nucleus | RASAL1 | ENSG00000111344 | 12 | 113526624 | 113609044 | 229 | 5.066 | 2.03e-07 | 0.00392 |
| Caudate nucleus | HIST1H2BD | ENSG00000158373 | 6 | 26123349 | 26181577 | 98 | 5.058 | 2.12e-07 | 0.00409 |
| Caudate nucleus | LPIN1 | ENSG00000134324 | 2 | 11782721 | 11977535 | 560 | 5.054 | 2.17e-07 | 0.00418 |
| Caudate nucleus | RUNDC1 | ENSG00000198863 | 17 | 41097582 | 41155707 | 67 | 5.051 | 2.2e-07 | 0.00425 |
| Caudate nucleus | HIST1H2BC | ENSG00000180596 | 6 | 26105101 | 26159154 | 129 | 5.043 | 2.29e-07 | 0.00443 |
| Caudate nucleus | MARK2 | ENSG00000072518 | 11 | 63571400 | 63688491 | 167 | 5.029 | 2.46e-07 | 0.00476 |
| Caudate nucleus | DVL2 | ENSG00000004975 | 17 | 7118660 | 7172864 | 60 | 5.014 | 2.67e-07 | 0.00515 |
| Caudate nucleus | ELP5 | ENSG00000170291 | 17 | 7119735 | 7173259 | 60 | 5.014 | 2.67e-07 | 0.00515 |
| Caudate nucleus | LRP1 | ENSG00000123384 | 12 | 57487276 | 57617134 | 175 | 4.992 | 2.99e-07 | 0.00578 |
| Caudate nucleus | DNAJC30 | ENSG00000176410 | 7 | 73085299 | 73132783 | 115 | 4.97 | 3.34e-07 | 0.00645 |
| Caudate nucleus | DLG4 | ENSG00000132535 | 17 | 7083209 | 7158021 | 101 | 4.951 | 3.7e-07 | 0.00714 |
| Caudate nucleus | CTD-2545G14.7 | ENSG00000262526 | 17 | 7133746 | 7182954 | 60 | 4.946 | 3.79e-07 | 0.00732 |
| Caudate nucleus | SLC17A4 | ENSG00000146039 | 6 | 25719927 | 25791419 | 244 | 4.938 | 3.95e-07 | 0.00762 |
| Caudate nucleus | PHF23 | ENSG00000040633 | 17 | 7128347 | 7178041 | 59 | 4.929 | 4.14e-07 | 0.00799 |
| Caudate nucleus | GABARAP | ENSG00000170296 | 17 | 7133333 | 7181089 | 57 | 4.91 | 4.56e-07 | 0.00881 |
| Caudate nucleus | ACADVL | ENSG00000072778 | 17 | 7085444 | 7138592 | 81 | 4.9 | 4.8e-07 | 0.00927 |
| Caudate nucleus | CTDNEP1 | ENSG00000175826 | 17 | 7136910 | 7190810 | 71 | 4.892 | 4.99e-07 | 0.00963 |
| Caudate nucleus | SLC2A4 | ENSG00000181856 | 17 | 7149986 | 7201576 | 81 | 4.878 | 5.36e-07 | 0.0104 |
| Caudate nucleus | CLDN7 | ENSG00000181885 | 17 | 7153222 | 7202302 | 82 | 4.876 | 5.41e-07 | 0.0104 |
| Caudate nucleus | RP1-4G17.5 | ENSG00000262302 | 17 | 7140148 | 7200408 | 83 | 4.876 | 5.41e-07 | 0.0105 |
| Caudate nucleus | FNIP2 | ENSG00000052795 | 4 | 159655290 | 159839201 | 245 | 4.854 | 6.05e-07 | 0.0117 |
| Caudate nucleus | CHRNB4 | ENSG00000117971 | 15 | 78906461 | 79055096 | 481 | 4.781 | 8.72e-07 | 0.0168 |
| Caudate nucleus | TEX33 | ENSG00000185264 | 22 | 37377163 | 37438882 | 232 | 4.779 | 8.79e-07 | 0.017 |
| Caudate nucleus | STAT5B | ENSG00000173757 | 17 | 40341186 | 40463725 | 144 | 4.772 | 9.13e-07 | 0.0176 |
| Caudate nucleus | BBX | ENSG00000114439 | 3 | 107206783 | 107540171 | 590 | 4.767 | 9.35e-07 | 0.018 |
| Caudate nucleus | TMEM236 | ENSG00000184040 | 10 | 18006218 | 18099855 | 10 | 4.766 | 9.37e-07 | 0.0181 |
| Caudate nucleus | DUOXA1 | ENSG00000140254 | 15 | 45399569 | 45457136 | 114 | 4.753 | 1e-06 | 0.0194 |
| Caudate nucleus | EZH1 | ENSG00000108799 | 17 | 40842293 | 40932071 | 66 | 4.747 | 1.03e-06 | 0.0199 |
| Caudate nucleus | NTSR2 | ENSG00000169006 | 2 | 11788304 | 11845290 | 211 | 4.743 | 1.06e-06 | 0.0204 |
| Caudate nucleus | MPST | ENSG00000128309 | 22 | 37380676 | 37435863 | 202 | 4.74 | 1.07e-06 | 0.0207 |
| Caudate nucleus | AARSD1 | ENSG00000266967 | 17 | 41092543 | 41151515 | 63 | 4.703 | 1.28e-06 | 0.0247 |
| Caudate nucleus | SLC8B1 | ENSG00000089060 | 12 | 113726564 | 113832298 | 215 | 4.679 | 1.44e-06 | 0.0278 |
| Caudate nucleus | HIST1H2AA | ENSG00000164508 | 6 | 25716291 | 25761790 | 164 | 4.676 | 1.46e-06 | 0.0283 |
| Caudate nucleus | STAB1 | ENSG00000010327 | 3 | 52494354 | 52568511 | 120 | 4.664 | 1.55e-06 | 0.0299 |
| Caudate nucleus | HIST1H3G | ENSG00000256018 | 6 | 26261146 | 26306612 | 133 | 4.654 | 1.63e-06 | 0.0314 |
| Caudate nucleus | RAMP2 | ENSG00000131477 | 17 | 40875465 | 40925059 | 41 | 4.635 | 1.78e-06 | 0.0344 |

| Trait | Gene symbol | Ensembl gene ID | Chromosome | Start (bp) | Stop (bp) | SNPs (n) | Z | P | Bonferroni-adjusted P |
|---|---|---|---|---|---|---|---|---|---|
| Caudate nucleus | RUNX2 | ENSG00000124813 | 6 | 45260894 | 45642086 | 964 | 4.596 | 2.15e-06 | 0.0415 |
| Dentate nucleus | LRRC16A | ENSG00000079691 | 6 | 25244306 | 25630758 | 1409 | 11.13 | 4.23e-29 | 8.17e-25 |
| Dentate nucleus | SLC25A37 | ENSG00000147454 | 8 | 23351318 | 23442976 | 425 | 8.445 | 1.52e-17 | 2.94e-13 |
| Dentate nucleus | SLC39A12 | ENSG00000148482 | 10 | 18205768 | 18342221 | 469 | 8.168 | 1.57e-16 | 3.04e-12 |
| Dentate nucleus | TF | ENSG00000091513 | 3 | 133429800 | 133507850 | 256 | 8.043 | 4.37e-16 | 8.43e-12 |
| Dentate nucleus | TFCP2 | ENSG00000135457 | 12 | 51477446 | 51601926 | 308 | 8.004 | 6.05e-16 | 1.17e-11 |
| Dentate nucleus | HIST1H2AC | ENSG00000180573 | 6 | 26089373 | 26149344 | 159 | 7.911 | 1.28e-15 | 2.46e-11 |
| Dentate nucleus | ABHD12 | ENSG00000100997 | 20 | 25265379 | 25406619 | 341 | 7.832 | 2.41e-15 | 4.65e-11 |
| Dentate nucleus | FNIP2 | ENSG00000052795 | 4 | 159655290 | 159839201 | 245 | 7.754 | 4.44e-15 | 8.57e-11 |
| Dentate nucleus | HIST1H1E | ENSG00000168298 | 6 | 26121559 | 26167343 | 86 | 7.735 | 5.16e-15 | 9.97e-11 |
| Dentate nucleus | GINS1 | ENSG00000101003 | 20 | 25353363 | 25443264 | 216 | 7.715 | 6.05e-15 | 1.17e-10 |
| Dentate nucleus | TMPRSS6 | ENSG00000187045 | 22 | 37451476 | 37540603 | 267 | 7.664 | 9.02e-15 | 1.74e-10 |
| Dentate nucleus | PPID | ENSG00000171497 | 4 | 159620286 | 159679548 | 76 | 7.599 | 1.49e-14 | 2.87e-10 |
| Dentate nucleus | HIST1H1T | ENSG00000187475 | 6 | 26097640 | 26143364 | 127 | 7.596 | 1.52e-14 | 2.94e-10 |
| Dentate nucleus | AC021860.1 | ENSG00000196355 | 4 | 38618029 | 38701430 | 179 | 7.57 | 1.86e-14 | 3.59e-10 |
| Dentate nucleus | SRPRB | ENSG00000144867 | 3 | 133467877 | 133554616 | 218 | 7.497 | 3.27e-14 | 6.31e-10 |
| Dentate nucleus | TST | ENSG00000128311 | 22 | 37396900 | 37450681 | 164 | 7.494 | 3.34e-14 | 6.44e-10 |
| Dentate nucleus | BTN3A2 | ENSG00000186470 | 6 | 26330387 | 26388546 | 250 | 7.485 | 3.57e-14 | 6.89e-10 |
| Dentate nucleus | BCL7B | ENSG00000106635 | 7 | 72940686 | 73007332 | 124 | 7.435 | 5.25e-14 | 1.01e-09 |
| Dentate nucleus | TEX33 | ENSG00000185264 | 22 | 37377163 | 37438882 | 232 | 7.385 | 7.6e-14 | 1.47e-09 |
| Dentate nucleus | HIST1H2BD | ENSG00000158373 | 6 | 26123349 | 26181577 | 98 | 7.366 | 8.77e-14 | 1.69e-09 |
| Dentate nucleus | KCTD17 | ENSG00000100379 | 22 | 37412779 | 37469430 | 102 | 7.342 | 1.06e-13 | 2.04e-09 |
| Dentate nucleus | MPST | ENSG00000128309 | 22 | 37380676 | 37435863 | 202 | 7.296 | 1.49e-13 | 2.87e-09 |
| Dentate nucleus | SLC17A2 | ENSG00000112337 | 6 | 25902982 | 25965946 | 182 | 7.295 | 1.5e-13 | 2.89e-09 |
| Dentate nucleus | HIGD1C | ENSG00000214511 | 12 | 51312705 | 51374289 | 197 | 7.253 | 2.04e-13 | 3.93e-09 |
| Dentate nucleus | PYGB | ENSG00000100994 | 20 | 25193705 | 25288650 | 270 | 7.251 | 2.07e-13 | 4e-09 |
| Dentate nucleus | CDSN | ENSG00000204539 | 6 | 31072867 | 31123223 | 522 | 7.18 | 3.48e-13 | 6.72e-09 |
| Dentate nucleus | RCOR2 | ENSG00000167771 | 11 | 63668693 | 63719316 | 60 | 7.172 | 3.69e-13 | 7.13e-09 |
| Dentate nucleus | SLC39A14 | ENSG00000104635 | 8 | 22189762 | 22301642 | 339 | 7.106 | 5.97e-13 | 1.15e-08 |
| Dentate nucleus | SCGN | ENSG00000079689 | 6 | 25617464 | 25712011 | 282 | 7.102 | 6.14e-13 | 1.19e-08 |
| Dentate nucleus | WDR75 | ENSG00000115368 | 2 | 190271159 | 190350291 | 241 | 7.082 | 7.12e-13 | 1.37e-08 |
| Dentate nucleus | FZD9 | ENSG00000188763 | 7 | 72813109 | 72860450 | 69 | 7.076 | 7.42e-13 | 1.43e-08 |
| Dentate nucleus | PKN1 | ENSG00000123143 | 19 | 14508865 | 14592679 | 201 | 7.072 | 7.61e-13 | 1.47e-08 |
| Dentate nucleus | TTLL5 | ENSG00000119685 | 14 | 76064968 | 76431421 | 613 | 7.071 | 7.71e-13 | 1.49e-08 |
| Dentate nucleus | RNH1 | ENSG00000023191 | 11 | 484512 | 542300 | 271 | 7.046 | 9.18e-13 | 1.77e-08 |
| Dentate nucleus | HIST1H2AA | ENSG00000164508 | 6 | 25716291 | 25761790 | 164 | 7.04 | 9.61e-13 | 1.86e-08 |
| Dentate nucleus | CSRNP2 | ENSG00000110925 | 12 | 51444990 | 51512447 | 166 | 7.032 | 1.02e-12 | 1.96e-08 |
| Dentate nucleus | DDHD1 | ENSG00000100523 | 14 | 53500686 | 53655000 | 259 | 7.018 | 1.12e-12 | 2.17e-08 |
| Dentate nucleus | FP15737 | ENSG00000215298 | 8 | 23395157 | 23442974 | 291 | 7.016 | 1.14e-12 | 2.2e-08 |
| Dentate nucleus | SLC40A1 | ENSG00000138449 | 2 | 190415305 | 190483484 | 125 | 6.969 | 1.59e-12 | 3.08e-08 |
| Dentate nucleus | OSTM1 | ENSG00000081087 | 6 | 108352613 | 108522058 | 394 | 6.917 | 2.31e-12 | 4.46e-08 |
| Dentate nucleus | HIST1H2BA | ENSG00000146047 | 6 | 25692137 | 25737573 | 144 | 6.915 | 2.34e-12 | 4.52e-08 |
| Dentate nucleus | MIEF2 | ENSG00000177427 | 17 | 18128848 | 18179866 | 92 | 6.892 | 2.75e-12 | 5.3e-08 |
| Dentate nucleus | CDV3 | ENSG00000091527 | 3 | 133257574 | 133319105 | 158 | 6.838 | 4.02e-12 | 7.75e-08 |
| Dentate nucleus | LLGL1 | ENSG00000131899 | 17 | 18093901 | 18158189 | 135 | 6.821 | 4.52e-12 | 8.73e-08 |
| Dentate nucleus | NUCKS1 | ENSG00000069275 | 1 | 205671947 | 205754404 | 198 | 6.808 | 4.96e-12 | 9.58e-08 |
| Dentate nucleus | NAA40 | ENSG00000110583 | 11 | 63671431 | 63734800 | 89 | 6.806 | 5e-12 | 9.65e-08 |
| Dentate nucleus | LIN37 | ENSG00000267796 | 19 | 36204262 | 36255420 | 108 | 6.779 | 6.05e-12 | 1.17e-07 |
| Dentate nucleus | AC002398.9 | ENSG00000188223 | 19 | 36201579 | 36255420 | 112 | 6.778 | 6.08e-12 | 1.17e-07 |
| Dentate nucleus | TOPBP1 | ENSG00000163781 | 3 | 133307019 | 133415737 | 271 | 6.775 | 6.24e-12 | 1.2e-07 |
| Dentate nucleus | TBL2 | ENSG00000106638 | 7 | 72973262 | 73028121 | 117 | 6.774 | 6.26e-12 | 1.21e-07 |
| Dentate nucleus | RNF24 | ENSG00000101236 | 20 | 3897956 | 4031229 | 323 | 6.768 | 6.53e-12 | 1.26e-07 |

| Trait | Gene symbol | Ensembl gene ID | Chromosome | Start (bp) | Stop (bp) | SNPs (n) | Z | P | Bonferroni-adjusted P |
|---|---|---|---|---|---|---|---|---|---|
| Dentate nucleus | NEFM | ENSG00000104722 | 8 | 24735525 | 24786607 | 85 | 6.71 | 9.76e-12 | 1.88e-07 |
| Dentate nucleus | C6orf15 | ENSG00000204542 | 6 | 31069000 | 31115336 | 510 | 6.709 | 9.79e-12 | 1.89e-07 |
| Dentate nucleus | ENTPD6 | ENSG00000197586 | 20 | 25141329 | 25217365 | 217 | 6.67 | 1.28e-11 | 2.47e-07 |
| Dentate nucleus | PSORS1C1 | ENSG00000204540 | 6 | 31047527 | 31117869 | 730 | 6.612 | 1.9e-11 | 3.67e-07 |
| Dentate nucleus | MLXIPL | ENSG00000009950 | 7 | 72997524 | 73073873 | 203 | 6.609 | 1.94e-11 | 3.75e-07 |
| Dentate nucleus | GAK | ENSG00000178950 | 4 | 833064 | 961161 | 436 | 6.608 | 1.94e-11 | 3.75e-07 |
| Dentate nucleus | GPX5 | ENSG00000224586 | 6 | 28458702 | 28512729 | 111 | 6.592 | 2.17e-11 | 4.2e-07 |
| Dentate nucleus | NINL | ENSG00000101004 | 20 | 25423341 | 25601153 | 399 | 6.558 | 2.73e-11 | 5.26e-07 |
| Dentate nucleus | BTN2A2 | ENSG00000124508 | 6 | 26348324 | 26405102 | 258 | 6.556 | 2.76e-11 | 5.33e-07 |
| Dentate nucleus | AC051642.1 | ENSG00000268085 | 8 | 23402287 | 23455651 | 319 | 6.505 | 3.88e-11 | 7.49e-07 |
| Dentate nucleus | TMEM236 | ENSG00000184040 | 10 | 18006218 | 18099855 | 10 | 6.476 | 4.69e-11 | 9.06e-07 |
| Dentate nucleus | LRRC56 | ENSG00000161328 | 11 | 502527 | 564916 | 277 | 6.434 | 6.23e-11 | 1.2e-06 |
| Dentate nucleus | WWP2 | ENSG00000198373 | 16 | 69761209 | 69985644 | 496 | 6.433 | 6.25e-11 | 1.21e-06 |
| Dentate nucleus | PTDSS2 | ENSG00000174915 | 11 | 413268 | 501393 | 268 | 6.433 | 6.27e-11 | 1.21e-06 |
| Dentate nucleus | SLC16A6 | ENSG00000108932 | 17 | 66253167 | 66322408 | 153 | 6.399 | 7.8e-11 | 1.51e-06 |
| Dentate nucleus | C2orf76 | ENSG00000186132 | 2 | 120049801 | 120159404 | 351 | 6.389 | 8.34e-11 | 1.61e-06 |
| Dentate nucleus | HLA-E | ENSG00000204592 | 6 | 30422244 | 30471982 | 251 | 6.332 | 1.21e-10 | 2.34e-06 |
| Dentate nucleus | ALKBH5 | ENSG00000091542 | 17 | 18051392 | 18123268 | 131 | 6.318 | 1.33e-10 | 2.56e-06 |
| Dentate nucleus | HIST1H2BI | ENSG00000168242 | 6 | 26238144 | 26283622 | 100 | 6.309 | 1.4e-10 | 2.71e-06 |
| Dentate nucleus | PSENEN | ENSG00000205155 | 19 | 36201015 | 36247911 | 94 | 6.297 | 1.51e-10 | 2.92e-06 |
| Dentate nucleus | GPX6 | ENSG00000198704 | 6 | 28461073 | 28530992 | 149 | 6.294 | 1.55e-10 | 2.99e-06 |
| Dentate nucleus | HIST1H4G | ENSG00000124578 | 6 | 26236886 | 26282259 | 102 | 6.284 | 1.65e-10 | 3.18e-06 |
| Dentate nucleus | HLA-C | ENSG00000204525 | 6 | 31226526 | 31274907 | 1493 | 6.276 | 1.73e-10 | 3.35e-06 |
| Dentate nucleus | HLA-B | ENSG00000234745 | 6 | 31311649 | 31359965 | 1542 | 6.242 | 2.17e-10 | 4.18e-06 |
| Dentate nucleus | HIST1H3F | ENSG00000256316 | 6 | 26240370 | 26285835 | 106 | 6.24 | 2.19e-10 | 4.23e-06 |
| Dentate nucleus | C14orf1 | ENSG00000133935 | 14 | 76106134 | 76162532 | 106 | 6.226 | 2.39e-10 | 4.62e-06 |
| Dentate nucleus | HIST1H2AM | ENSG00000233224 | 6 | 27850477 | 27895963 | 81 | 6.224 | 2.42e-10 | 4.67e-06 |
| Dentate nucleus | IFT43 | ENSG00000119650 | 14 | 76333479 | 76560928 | 462 | 6.185 | 3.1e-10 | 5.98e-06 |
| Dentate nucleus | HIST1H3J | ENSG00000197153 | 6 | 27848093 | 27895884 | 85 | 6.128 | 4.45e-10 | 8.59e-06 |
| Dentate nucleus | ZKSCAN4 | ENSG00000187626 | 6 | 28202401 | 28262011 | 104 | 6.128 | 4.45e-10 | 8.59e-06 |
| Dentate nucleus | HIST1H4J | ENSG00000197238 | 6 | 27756884 | 27802257 | 77 | 6.122 | 4.62e-10 | 8.93e-06 |
| Dentate nucleus | PHRF1 | ENSG00000070047 | 11 | 541486 | 622222 | 312 | 6.12 | 4.67e-10 | 9.01e-06 |
| Dentate nucleus | DBI | ENSG00000155368 | 2 | 120089497 | 120140126 | 163 | 6.116 | 4.8e-10 | 9.28e-06 |
| Dentate nucleus | PTGER1 | ENSG00000160951 | 19 | 14573278 | 14621174 | 187 | 6.11 | 4.97e-10 | 9.59e-06 |
| Dentate nucleus | C4orf45 | ENSG00000164123 | 4 | 159804286 | 159994912 | 466 | 6.11 | 4.97e-10 | 9.6e-06 |
| Dentate nucleus | KLF3 | ENSG00000109787 | 4 | 38630817 | 38712663 | 176 | 6.109 | 5e-10 | 9.65e-06 |
| Dentate nucleus | SLC17A4 | ENSG00000146039 | 6 | 25719927 | 25791419 | 244 | 6.109 | 5e-10 | 9.65e-06 |
| Dentate nucleus | SLC17A1 | ENSG00000124568 | 6 | 25773125 | 25867287 | 232 | 6.109 | 5e-10 | 9.65e-06 |
| Dentate nucleus | SLC17A3 | ENSG00000124564 | 6 | 25823294 | 25917514 | 233 | 6.109 | 5e-10 | 9.65e-06 |
| Dentate nucleus | TRIM38 | ENSG00000112343 | 6 | 25928030 | 25997384 | 160 | 6.109 | 5e-10 | 9.65e-06 |
| Dentate nucleus | HIST1H3A | ENSG00000198366 | 6 | 25985718 | 26031186 | 80 | 6.109 | 5e-10 | 9.65e-06 |
| Dentate nucleus | HIST1H4A | ENSG00000196176 | 6 | 25986907 | 26032278 | 79 | 6.109 | 5e-10 | 9.65e-06 |
| Dentate nucleus | HIST1H1A | ENSG00000124610 | 6 | 26007260 | 26053040 | 93 | 6.109 | 5e-10 | 9.65e-06 |
| Dentate nucleus | HIST1H3C | ENSG00000196532 | 6 | 26010639 | 26056097 | 90 | 6.109 | 5e-10 | 9.65e-06 |
| Dentate nucleus | HIST1H4B | ENSG00000124529 | 6 | 26017124 | 26062480 | 89 | 6.109 | 5e-10 | 9.65e-06 |
| Dentate nucleus | HIST1H3B | ENSG00000124693 | 6 | 26021817 | 26067288 | 86 | 6.109 | 5e-10 | 9.65e-06 |
| Dentate nucleus | HIST1H2AB | ENSG00000137259 | 6 | 26023320 | 26068796 | 82 | 6.109 | 5e-10 | 9.65e-06 |
| Dentate nucleus | HIST1H2BB | ENSG00000196226 | 6 | 26033455 | 26078885 | 94 | 6.109 | 5e-10 | 9.65e-06 |
| Dentate nucleus | HIST1H1C | ENSG00000187837 | 6 | 26045968 | 26091699 | 102 | 6.109 | 5e-10 | 9.65e-06 |
| Dentate nucleus | HFE | ENSG00000010704 | 6 | 26052509 | 26108571 | 135 | 6.109 | 5e-10 | 9.65e-06 |
| Dentate nucleus | HIST1H4C | ENSG00000197061 | 6 | 26069104 | 26114518 | 129 | 6.109 | 5e-10 | 9.65e-06 |
| Dentate nucleus | HIST1H2BC | ENSG00000180596 | 6 | 26105101 | 26159154 | 129 | 6.109 | 5e-10 | 9.65e-06 |

| Trait | Gene symbol | Ensembl gene ID | Chromosome | Start (bp) | Stop (bp) | SNPs (n) | Z | P | Bonferroni-adjusted P |
|---|---|---|---|---|---|---|---|---|---|
| Dentate nucleus | BAZ1B | ENSG00000009954 | 7 | 72844728 | 72971608 | 174 | 6.109 | 5e-10 | 9.65e-06 |
| Dentate nucleus | MARK2 | ENSG00000072518 | 11 | 63571400 | 63688491 | 167 | 6.109 | 5e-10 | 9.65e-06 |
| Dentate nucleus | METTL7A | ENSG00000185432 | 12 | 51282255 | 51336300 | 120 | 6.109 | 5e-10 | 9.65e-06 |
| Dentate nucleus | SLC11A2 | ENSG00000110911 | 12 | 51363184 | 51457349 | 233 | 6.109 | 5e-10 | 9.65e-06 |
| Dentate nucleus | LETMD1 | ENSG00000050426 | 12 | 51406745 | 51464207 | 134 | 6.109 | 5e-10 | 9.65e-06 |
| Dentate nucleus | PANK2 | ENSG00000125779 | 20 | 3834486 | 3917605 | 201 | 6.109 | 5e-10 | 9.65e-06 |
| Dentate nucleus | AL035252.1 | ENSG00000268302 | 20 | 25197122 | 25242370 | 96 | 6.109 | 5e-10 | 9.65e-06 |
| Dentate nucleus | PPP3CC | ENSG00000120910 | 8 | 22263332 | 22408652 | 289 | 6.089 | 5.67e-10 | 1.09e-05 |
| Dentate nucleus | SLC41A1 | ENSG00000133065 | 1 | 205748221 | 205817876 | 178 | 6.082 | 5.92e-10 | 1.14e-05 |
| Dentate nucleus | HIST1H2BO | ENSG00000196331 | 6 | 27826203 | 27871669 | 79 | 6.081 | 5.98e-10 | 1.15e-05 |
| Dentate nucleus | HIST1H4H | ENSG00000158406 | 6 | 26271283 | 26320762 | 164 | 6.056 | 6.97e-10 | 1.35e-05 |
| Dentate nucleus | COX8A | ENSG00000176340 | 11 | 63707079 | 63754015 | 87 | 6.045 | 7.46e-10 | 1.44e-05 |
| Dentate nucleus | RAB6B | ENSG00000154917 | 3 | 133533083 | 133649680 | 298 | 6.041 | 7.66e-10 | 1.48e-05 |
| Dentate nucleus | AP000721.4 | ENSG00000256100 | 11 | 63707092 | 63765818 | 101 | 6.03 | 8.21e-10 | 1.59e-05 |
| Dentate nucleus | ENTPD4 | ENSG00000197217 | 8 | 23233296 | 23350208 | 333 | 6.029 | 8.26e-10 | 1.6e-05 |
| Dentate nucleus | HIST1H3I | ENSG00000182572 | 6 | 27829623 | 27875099 | 86 | 6.025 | 8.44e-10 | 1.63e-05 |
| Dentate nucleus | HIST1H1B | ENSG00000184357 | 6 | 27824570 | 27870359 | 79 | 6.015 | 8.97e-10 | 1.73e-05 |
| Dentate nucleus | NFAT5 | ENSG00000102908 | 16 | 69563997 | 69748569 | 343 | 6.008 | 9.4e-10 | 1.81e-05 |
| Dentate nucleus | FAM182B | ENSG00000175170 | 20 | 25734102 | 25883861 | 233 | 5.998 | 9.99e-10 | 1.93e-05 |
| Dentate nucleus | HIST1H2BL | ENSG00000185130 | 6 | 27765257 | 27810709 | 84 | 5.996 | 1.01e-09 | 1.95e-05 |
| Dentate nucleus | OTUB1 | ENSG00000167770 | 11 | 63718325 | 63779283 | 111 | 5.996 | 1.01e-09 | 1.95e-05 |
| Dentate nucleus | PSORS1C2 | ENSG00000204538 | 6 | 31095313 | 31142127 | 397 | 5.982 | 1.1e-09 | 2.12e-05 |
| Dentate nucleus | INA | ENSG00000148798 | 10 | 105001920 | 105060108 | 95 | 5.973 | 1.16e-09 | 2.24e-05 |
| Dentate nucleus | HIST1H2BN | ENSG00000233822 | 6 | 27771323 | 27833487 | 95 | 5.972 | 1.17e-09 | 2.26e-05 |
| Dentate nucleus | ABT1 | ENSG00000146109 | 6 | 26562180 | 26610278 | 125 | 5.971 | 1.18e-09 | 2.27e-05 |
| Dentate nucleus | HIST1H4L | ENSG00000198558 | 6 | 27830926 | 27876289 | 86 | 5.969 | 1.19e-09 | 2.3e-05 |
| Dentate nucleus | PGBD1 | ENSG00000137338 | 6 | 28214314 | 28280326 | 131 | 5.965 | 1.22e-09 | 2.36e-05 |
| Dentate nucleus | TCF19 | ENSG00000137310 | 6 | 31091319 | 31144936 | 481 | 5.963 | 1.24e-09 | 2.39e-05 |
| Dentate nucleus | HIST1H2BM | ENSG00000196374 | 6 | 27747822 | 27793267 | 70 | 5.958 | 1.28e-09 | 2.46e-05 |
| Dentate nucleus | AGMAT | ENSG00000116771 | 1 | 15888848 | 15946605 | 208 | 5.956 | 1.29e-09 | 2.5e-05 |
| Dentate nucleus | HIST1H2AJ | ENSG00000182611 | 6 | 27772112 | 27817607 | 81 | 5.95 | 1.34e-09 | 2.59e-05 |
| Dentate nucleus | ARSG | ENSG00000141337 | 17 | 66220323 | 66428872 | 589 | 5.939 | 1.43e-09 | 2.76e-05 |
| Dentate nucleus | RAB7L1 | ENSG00000117280 | 1 | 205727114 | 205779588 | 163 | 5.939 | 1.44e-09 | 2.77e-05 |
| Dentate nucleus | DDI2 | ENSG00000197312 | 1 | 15908995 | 16005539 | 273 | 5.935 | 1.47e-09 | 2.84e-05 |
| Dentate nucleus | ASAP1 | ENSG00000153317 | 8 | 131054353 | 131490906 | 1184 | 5.906 | 1.75e-09 | 3.39e-05 |
| Dentate nucleus | CCHCR1 | ENSG00000204536 | 6 | 31100216 | 31161015 | 609 | 5.893 | 1.9e-09 | 3.66e-05 |
| Dentate nucleus | PPP1R21 | ENSG00000162869 | 2 | 48632737 | 48752525 | 297 | 5.891 | 1.92e-09 | 3.7e-05 |
| Dentate nucleus | ANO9 | ENSG00000185101 | 11 | 407933 | 477011 | 173 | 5.878 | 2.07e-09 | 4e-05 |
| Dentate nucleus | HIST1H3H | ENSG00000203813 | 6 | 27742842 | 27788314 | 72 | 5.876 | 2.11e-09 | 4.07e-05 |
| Dentate nucleus | ABLIM1 | ENSG00000099204 | 10 | 116180872 | 116479762 | 787 | 5.869 | 2.2e-09 | 4.24e-05 |
| Dentate nucleus | TIE1 | ENSG00000066056 | 1 | 43731664 | 43798779 | 99 | 5.863 | 2.27e-09 | 4.38e-05 |
| Dentate nucleus | HIST1H3G | ENSG00000256018 | 6 | 26261146 | 26306612 | 133 | 5.858 | 2.35e-09 | 4.53e-05 |
| Dentate nucleus | DVL2 | ENSG00000004975 | 17 | 7118660 | 7172864 | 60 | 5.833 | 2.72e-09 | 5.26e-05 |
| Dentate nucleus | ELP5 | ENSG00000170291 | 17 | 7119735 | 7173259 | 60 | 5.833 | 2.72e-09 | 5.26e-05 |
| Dentate nucleus | DRD4 | ENSG00000069696 | 11 | 602293 | 650706 | 179 | 5.816 | 3.01e-09 | 5.81e-05 |
| Dentate nucleus | IRF7 | ENSG00000185507 | 11 | 602553 | 650999 | 181 | 5.803 | 3.26e-09 | 6.29e-05 |
| Dentate nucleus | TTC9C | ENSG00000162222 | 11 | 62460541 | 62517765 | 82 | 5.801 | 3.3e-09 | 6.38e-05 |
| Dentate nucleus | NKAPL | ENSG00000189134 | 6 | 28192098 | 28238736 | 95 | 5.796 | 3.39e-09 | 6.54e-05 |
| Dentate nucleus | C1orf210 | ENSG00000253313 | 1 | 43737554 | 43786288 | 73 | 5.794 | 3.44e-09 | 6.64e-05 |
| Dentate nucleus | BSCL2 | ENSG00000168000 | 11 | 62447747 | 62512317 | 90 | 5.769 | 3.99e-09 | 7.71e-05 |
| Dentate nucleus | HNRNPUL2 | ENSG00000214753 | 11 | 62470102 | 62529821 | 78 | 5.767 | 4.04e-09 | 7.79e-05 |
| Dentate nucleus | LRRN4CL | ENSG00000177363 | 11 | 62443874 | 62492371 | 66 | 5.767 | 4.04e-09 | 7.8e-05 |

| Trait | Gene symbol | Ensembl gene ID | Chromosome | Start (bp) | Stop (bp) | SNPs (n) | Z | P | Bonferroni-adjusted P |
|---|---|---|---|---|---|---|---|---|---|
| Dentate nucleus | BTN3A1 | ENSG00000026950 | 6 | 26367465 | 26425444 | 261 | 5.763 | 4.13e-09 | 7.98e-05 |
| Dentate nucleus | HNRNPUL2-BSCL2 | ENSG00000234857 | 11 | 62447747 | 62529856 | 113 | 5.752 | 4.42e-09 | 8.53e-05 |
| Dentate nucleus | ZNF322 | ENSG00000181315 | 6 | 26626518 | 26694980 | 147 | 5.74 | 4.72e-09 | 9.12e-05 |
| Dentate nucleus | GNG3 | ENSG00000162188 | 11 | 62440130 | 62486673 | 62 | 5.726 | 5.13e-09 | 9.9e-05 |
| Dentate nucleus | HIST1H4K | ENSG00000197914 | 6 | 27788952 | 27834305 | 64 | 5.724 | 5.2e-09 | 0.0001 |
| Dentate nucleus | HIST1H2AL | ENSG00000198374 | 6 | 27798034 | 27843606 | 65 | 5.722 | 5.26e-09 | 0.000102 |
| Dentate nucleus | PHF23 | ENSG00000040633 | 17 | 7128347 | 7178041 | 59 | 5.718 | 5.38e-09 | 0.000104 |
| Dentate nucleus | MPL | ENSG00000117400 | 1 | 43768478 | 43828443 | 66 | 5.713 | 5.55e-09 | 0.000107 |
| Dentate nucleus | SNAP91 | ENSG00000065609 | 6 | 84252599 | 84454410 | 312 | 5.71 | 5.67e-09 | 0.000109 |
| Dentate nucleus | HIST1H2AH | ENSG00000184825 | 6 | 27079861 | 27125317 | 70 | 5.709 | 5.69e-09 | 0.00011 |
| Dentate nucleus | HIST1H2AK | ENSG00000184348 | 6 | 27795658 | 27841117 | 65 | 5.705 | 5.81e-09 | 0.000112 |

**Supplementary Table 4. Significant genes from MAGMA gene-based analysis of quantitative susceptibility mapping-derived brain iron across six subcortical regions (Bonferroni-corrected P < 2.59 x 10-6).** ***(continued)***

| Trait | Gene symbol | Ensembl gene ID | Chromosome | Start (bp) | Stop (bp) | SNPs (n) | Z | P | Bonferroni-adjusted P |
|---|---|---|---|---|---|---|---|---|---|
| Dentate nucleus | RPEL1 | ENSG00000235376 | 10 | 104970644 | 105017773 | 67 | 5.676 | 6.89e-09 | 0.000133 |
| Dentate nucleus | COL5A2 | ENSG00000204262 | 2 | 189886622 | 190079605 | 456 | 5.676 | 6.9e-09 | 0.000133 |
| Dentate nucleus | CTD-2545G14.7 | ENSG00000262526 | 17 | 7133746 | 7182954 | 60 | 5.669 | 7.18e-09 | 0.000139 |
| Dentate nucleus | RSC1A1 | ENSG00000215695 | 1 | 15951364 | 15998217 | 91 | 5.651 | 7.99e-09 | 0.000154 |
| Dentate nucleus | DLG4 | ENSG00000132535 | 17 | 7083209 | 7158021 | 101 | 5.641 | 8.47e-09 | 0.000164 |
| Dentate nucleus | CDHR5 | ENSG00000099834 | 11 | 606565 | 661078 | 212 | 5.64 | 8.5e-09 | 0.000164 |
| Dentate nucleus | NFU1 | ENSG00000169599 | 2 | 69612882 | 69699760 | 160 | 5.637 | 8.63e-09 | 0.000167 |
| Dentate nucleus | ZSCAN9 | ENSG00000137185 | 6 | 28157664 | 28211260 | 106 | 5.632 | 8.9e-09 | 0.000172 |
| Dentate nucleus | GABARAP | ENSG00000170296 | 17 | 7133333 | 7181089 | 57 | 5.628 | 9.09e-09 | 0.000176 |
| Dentate nucleus | IGFLR1 | ENSG00000126246 | 19 | 36220058 | 36268354 | 124 | 5.627 | 9.15e-09 | 0.000177 |
| Dentate nucleus | CTDNEP1 | ENSG00000175826 | 17 | 7136910 | 7190810 | 71 | 5.611 | 1e-08 | 0.000194 |
| Dentate nucleus | OR2B6 | ENSG00000124657 | 6 | 27890019 | 27935960 | 61 | 5.61 | 1.01e-08 | 0.000195 |
| Dentate nucleus | FAM49B | ENSG00000153310 | 8 | 130841839 | 131064375 | 434 | 5.601 | 1.07e-08 | 0.000206 |
| Dentate nucleus | KMT2B | ENSG00000272333 | 19 | 36173921 | 36239779 | 150 | 5.599 | 1.08e-08 | 0.000209 |
| Dentate nucleus | CDC20 | ENSG00000117399 | 1 | 43789626 | 43838874 | 56 | 5.596 | 1.1e-08 | 0.000212 |
| Dentate nucleus | BTN1A1 | ENSG00000124557 | 6 | 26466449 | 26520650 | 118 | 5.58 | 1.2e-08 | 0.000232 |
| Dentate nucleus | UBXN1 | ENSG00000162191 | 11 | 62433970 | 62481567 | 67 | 5.566 | 1.3e-08 | 0.000252 |
| Dentate nucleus | MOSPD3 | ENSG00000106330 | 7 | 100174725 | 100223007 | 61 | 5.564 | 1.32e-08 | 0.000254 |
| Dentate nucleus | LMF1 | ENSG00000103227 | 16 | 893634 | 1066318 | 956 | 5.561 | 1.34e-08 | 0.000259 |
| Dentate nucleus | PICALM | ENSG00000073921 | 11 | 85658727 | 85815924 | 381 | 5.533 | 1.58e-08 | 0.000305 |
| Dentate nucleus | C19orf55 | ENSG00000167595 | 19 | 36214044 | 36271930 | 143 | 5.52 | 1.69e-08 | 0.000327 |
| Dentate nucleus | HIST1H1D | ENSG00000124575 | 6 | 26224440 | 26270216 | 104 | 5.513 | 1.77e-08 | 0.000342 |
| Dentate nucleus | POLR2G | ENSG00000168002 | 11 | 62494016 | 62544182 | 66 | 5.51 | 1.79e-08 | 0.000347 |
| Dentate nucleus | FLII | ENSG00000177731 | 17 | 18138150 | 18197230 | 107 | 5.499 | 1.91e-08 | 0.000369 |
| Dentate nucleus | OR2B2 | ENSG00000168131 | 6 | 27868963 | 27915174 | 76 | 5.497 | 1.94e-08 | 0.000374 |
| Dentate nucleus | ELOVL1 | ENSG00000066322 | 1 | 43819068 | 43868696 | 66 | 5.494 | 1.97e-08 | 0.00038 |
| Dentate nucleus | RASSF7 | ENSG00000099849 | 11 | 525404 | 574021 | 232 | 5.484 | 2.08e-08 | 0.000401 |
| Dentate nucleus | HRAS | ENSG00000174775 | 11 | 522242 | 572287 | 245 | 5.48 | 2.13e-08 | 0.000411 |
| Dentate nucleus | ACADVL | ENSG00000072778 | 17 | 7085444 | 7138592 | 81 | 5.477 | 2.17e-08 | 0.000418 |
| Dentate nucleus | GFPT1 | ENSG00000198380 | 2 | 69536905 | 69649382 | 197 | 5.47 | 2.25e-08 | 0.000435 |
| Dentate nucleus | DNAJC16 | ENSG00000116138 | 1 | 15818308 | 15928874 | 368 | 5.456 | 2.43e-08 | 0.000469 |
| Dentate nucleus | PCGF6 | ENSG00000156374 | 10 | 105052553 | 105145891 | 135 | 5.445 | 2.59e-08 | 0.000501 |
| Dentate nucleus | ZNF165 | ENSG00000197279 | 6 | 28013753 | 28067341 | 151 | 5.424 | 2.91e-08 | 0.000563 |
| Dentate nucleus | XPO1 | ENSG00000082898 | 2 | 61694984 | 61800761 | 175 | 5.399 | 3.35e-08 | 0.000647 |
| Dentate nucleus | SLC2A4 | ENSG00000181856 | 17 | 7149986 | 7201576 | 81 | 5.394 | 3.45e-08 | 0.000667 |
| Dentate nucleus | CLDN7 | ENSG00000181885 | 17 | 7153222 | 7202302 | 82 | 5.385 | 3.62e-08 | 0.000699 |
| Dentate nucleus | ZSCAN16 | ENSG00000196812 | 6 | 28057338 | 28107860 | 150 | 5.38 | 3.73e-08 | 0.00072 |
| Dentate nucleus | RP1-4G17.5 | ENSG00000262302 | 17 | 7140148 | 7200408 | 83 | 5.375 | 3.84e-08 | 0.000741 |
| Dentate nucleus | C11orf35 | ENSG00000185522 | 11 | 544855 | 595779 | 205 | 5.35 | 4.41e-08 | 0.000851 |
| Dentate nucleus | CLEC18A | ENSG00000157322 | 16 | 69949810 | 70008141 | 142 | 5.346 | 4.5e-08 | 0.000869 |
| Dentate nucleus | C11orf48 | ENSG00000162194 | 11 | 62420287 | 62474727 | 77 | 5.333 | 4.84e-08 | 0.000934 |
| Dentate nucleus | MAS1L | ENSG00000204687 | 6 | 29444474 | 29490738 | 113 | 5.331 | 4.89e-08 | 0.000944 |
| Dentate nucleus | TAF5 | ENSG00000148835 | 10 | 105092724 | 105158822 | 88 | 5.329 | 4.94e-08 | 0.000954 |
| Dentate nucleus | OR2J2 | ENSG00000204700 | 6 | 29106311 | 29152351 | 174 | 5.309 | 5.52e-08 | 0.00107 |
| Dentate nucleus | AD000671.6 | ENSG00000267120 | 19 | 36220153 | 36271333 | 133 | 5.305 | 5.63e-08 | 0.00109 |
| Dentate nucleus | USP34 | ENSG00000115464 | 2 | 61404598 | 61732904 | 607 | 5.302 | 5.73e-08 | 0.00111 |
| Dentate nucleus | U2AF1L4 | ENSG00000161265 | 19 | 36223365 | 36271346 | 125 | 5.287 | 6.23e-08 | 0.0012 |
| Dentate nucleus | AMPD3 | ENSG00000133805 | 11 | 10294860 | 10539126 | 804 | 5.27 | 6.82e-08 | 0.00132 |
| Dentate nucleus | TGFB3 | ENSG00000119699 | 14 | 76414442 | 76484334 | 93 | 5.269 | 6.87e-08 | 0.00133 |

| Trait | Gene symbol | Ensembl gene ID | Chromosome | Start (bp) | Stop (bp) | SNPs (n) | Z | P | Bonferroni-adjusted P |
|---|---|---|---|---|---|---|---|---|---|
| Dentate nucleus | HIST1H2AI | ENSG00000196747 | 6 | 27740899 | 27786429 | 74 | 5.266 | 6.98e-08 | 0.00135 |
| Dentate nucleus | CP | ENSG00000047457 | 3 | 148870197 | 148974842 | 304 | 5.264 | 7.04e-08 | 0.00136 |
| Dentate nucleus | MYOZ2 | ENSG00000172399 | 4 | 120021939 | 120118944 | 301 | 5.264 | 7.05e-08 | 0.00136 |
| Dentate nucleus | TMEM206 | ENSG00000065600 | 1 | 212527273 | 212623243 | 229 | 5.263 | 7.08e-08 | 0.00137 |
| Dentate nucleus | TOP3A | ENSG00000177302 | 17 | 18164742 | 18253321 | 193 | 5.26 | 7.21e-08 | 0.00139 |
| Dentate nucleus | NQO1 | ENSG00000181019 | 16 | 69730899 | 69795854 | 129 | 5.258 | 7.28e-08 | 0.00141 |
| Dentate nucleus | STMN4 | ENSG00000015592 | 8 | 27082840 | 27150937 | 161 | 5.255 | 7.39e-08 | 0.00143 |
| Dentate nucleus | STX6 | ENSG00000135823 | 1 | 180931861 | 181027047 | 200 | 5.25 | 7.59e-08 | 0.00146 |
| Dentate nucleus | HIST1H2BJ | ENSG00000124635 | 6 | 27083676 | 27135541 | 81 | 5.233 | 8.35e-08 | 0.00161 |
| Dentate nucleus | CELA2B | ENSG00000215704 | 1 | 15757404 | 15827895 | 198 | 5.227 | 8.6e-08 | 0.00166 |
| Dentate nucleus | PLEKHM2 | ENSG00000116786 | 1 | 15975827 | 16071264 | 227 | 5.212 | 9.32e-08 | 0.0018 |
| Dentate nucleus | FLVCR2 | ENSG00000119686 | 14 | 76009960 | 76139557 | 283 | 5.197 | 1.01e-07 | 0.00195 |
| Dentate nucleus | STK38L | ENSG00000211455 | 12 | 27361901 | 27488892 | 317 | 5.185 | 1.08e-07 | 0.00208 |
| Dentate nucleus | RAPGEF5 | ENSG00000136237 | 7 | 22147856 | 22431763 | 806 | 5.181 | 1.11e-07 | 0.00214 |
| Dentate nucleus | HIST1H2BK | ENSG00000197903 | 6 | 27096073 | 27149619 | 78 | 5.17 | 1.17e-07 | 0.00226 |
| Dentate nucleus | HIST1H2AG | ENSG00000196787 | 6 | 27065832 | 27113070 | 71 | 5.164 | 1.21e-07 | 0.00234 |
| Dentate nucleus | LTA | ENSG00000226979 | 6 | 31504831 | 31552101 | 198 | 5.162 | 1.22e-07 | 0.00236 |
| Dentate nucleus | ARHGAP33 | ENSG00000004777 | 19 | 36230434 | 36289724 | 142 | 5.162 | 1.22e-07 | 0.00236 |
| Dentate nucleus | ATP6V1G2 | ENSG00000213760 | 6 | 31502239 | 31551204 | 209 | 5.16 | 1.24e-07 | 0.00239 |
| Dentate nucleus | PTK2 | ENSG00000169398 | 8 | 141657999 | 142047315 | 589 | 5.158 | 1.25e-07 | 0.00241 |
| Dentate nucleus | ZKSCAN8 | ENSG00000198315 | 6 | 28074688 | 28137250 | 181 | 5.142 | 1.36e-07 | 0.00262 |
| Dentate nucleus | FOXN2 | ENSG00000170802 | 2 | 48506776 | 48616433 | 240 | 5.141 | 1.37e-07 | 0.00264 |
| Dentate nucleus | AC009041.2 | ENSG00000269047 | 16 | 996252 | 1041663 | 281 | 5.13 | 1.45e-07 | 0.0028 |
| Dentate nucleus | CNNM2 | ENSG00000148842 | 10 | 104643050 | 104859978 | 496 | 5.126 | 1.48e-07 | 0.00286 |
| Dentate nucleus | AAK1 | ENSG00000115977 | 2 | 69678532 | 69936481 | 304 | 5.123 | 1.5e-07 | 0.0029 |
| Dentate nucleus | SCT | ENSG00000070031 | 11 | 616431 | 662143 | 175 | 5.112 | 1.6e-07 | 0.00308 |
| Dentate nucleus | TAF6L | ENSG00000162227 | 11 | 62503775 | 62564814 | 77 | 5.111 | 1.6e-07 | 0.00309 |
| Dentate nucleus | PCOLCE | ENSG00000106333 | 7 | 100164800 | 100215798 | 63 | 5.111 | 1.6e-07 | 0.0031 |
| Dentate nucleus | OR5V1 | ENSG00000243729 | 6 | 29313007 | 29434744 | 411 | 5.108 | 1.63e-07 | 0.00314 |
| Dentate nucleus | AS3MT | ENSG00000214435 | 10 | 104594273 | 104671656 | 213 | 5.106 | 1.64e-07 | 0.00317 |
| Dentate nucleus | AGO2 | ENSG00000123908 | 8 | 141531264 | 141680718 | 376 | 5.101 | 1.69e-07 | 0.00325 |
| Dentate nucleus | LRCH4 | ENSG00000077454 | 7 | 100159855 | 100218776 | 76 | 5.101 | 1.69e-07 | 0.00327 |
| Dentate nucleus | STON1 | ENSG00000243244 | 2 | 48721522 | 48836025 | 363 | 5.07 | 1.99e-07 | 0.00384 |
| Dentate nucleus | DUOX1 | ENSG00000137857 | 15 | 45387131 | 45467774 | 158 | 5.065 | 2.05e-07 | 0.00395 |
| Dentate nucleus | NXNL1 | ENSG00000171773 | 19 | 17556234 | 17606763 | 149 | 5.051 | 2.2e-07 | 0.00424 |
| Dentate nucleus | TFR2 | ENSG00000106327 | 7 | 100208039 | 100275402 | 97 | 5.05 | 2.2e-07 | 0.00426 |
| Dentate nucleus | TRIM31 | ENSG00000204616 | 6 | 30060674 | 30115883 | 372 | 5.044 | 2.28e-07 | 0.0044 |
| Dentate nucleus | HSPB6 | ENSG00000004776 | 19 | 36235469 | 36283980 | 111 | 5.036 | 2.37e-07 | 0.00458 |
| Dentate nucleus | TRIM26 | ENSG00000234127 | 6 | 30142232 | 30216204 | 298 | 5.035 | 2.39e-07 | 0.00462 |
| Dentate nucleus | TLR10 | ENSG00000174123 | 4 | 38763860 | 38819611 | 251 | 5.022 | 2.56e-07 | 0.00493 |
| Dentate nucleus | FLJ27365 | ENSG00000197182 | 22 | 46414749 | 46519808 | 255 | 5.013 | 2.67e-07 | 0.00516 |
| Dentate nucleus | OR12D3 | ENSG00000112462 | 6 | 29331200 | 29378068 | 199 | 5.008 | 2.75e-07 | 0.00532 |
| Dentate nucleus | OR12D2 | ENSG00000168787 | 6 | 29329416 | 29375448 | 202 | 5.006 | 2.77e-07 | 0.00536 |
| Dentate nucleus | DDX39B | ENSG00000198563 | 6 | 31487996 | 31545225 | 269 | 4.999 | 2.88e-07 | 0.00555 |
| Dentate nucleus | TMEM175 | ENSG00000127419 | 4 | 891175 | 962444 | 189 | 4.997 | 2.91e-07 | 0.00561 |
| Dentate nucleus | LST1 | ENSG00000204482 | 6 | 31518901 | 31566686 | 189 | 4.992 | 2.99e-07 | 0.00578 |
| Dentate nucleus | USMG5 | ENSG00000173915 | 10 | 105138798 | 105191223 | 63 | 4.988 | 3.05e-07 | 0.00589 |
| Dentate nucleus | C10orf32-ASMT | ENSG00000270316 | 10 | 104579029 | 104671656 | 245 | 4.986 | 3.08e-07 | 0.00594 |
| Dentate nucleus | AC119673.1 | ENSG00000268313 | 1 | 205647497 | 205694153 | 103 | 4.985 | 3.09e-07 | 0.00597 |
| Dentate nucleus | SMCR8 | ENSG00000176994 | 17 | 18183624 | 18236517 | 123 | 4.982 | 3.14e-07 | 0.00606 |
| Dentate nucleus | ATP6V1G2-DDX39B | ENSG00000254870 | 6 | 31487996 | 31549385 | 278 | 4.981 | 3.16e-07 | 0.00609 |
| Dentate nucleus | MED8 | ENSG00000159479 | 1 | 43839588 | 43890479 | 75 | 4.981 | 3.16e-07 | 0.0061 |

| Trait | Gene symbol | Ensembl gene ID | Chromosome | Start (bp) | Stop (bp) | SNPs (n) | Z | P | Bonferroni-adjusted P |
|---|---|---|---|---|---|---|---|---|---|
| Dentate nucleus | TRIM10 | ENSG00000204613 | 6 | 30109722 | 30163711 | 259 | 4.972 | 3.31e-07 | 0.0064 |
| Dentate nucleus | CAPS | ENSG00000105519 | 19 | 5876718 | 5925888 | 163 | 4.958 | 3.57e-07 | 0.00689 |
| Dentate nucleus | SHMT1 | ENSG00000176974 | 17 | 18221187 | 18301856 | 214 | 4.955 | 3.62e-07 | 0.00699 |
| Dentate nucleus | SOX8 | ENSG00000005513 | 16 | 996808 | 1046979 | 303 | 4.951 | 3.7e-07 | 0.00714 |
| Dentate nucleus | TNF | ENSG00000232810 | 6 | 31508344 | 31556113 | 183 | 4.95 | 3.71e-07 | 0.00716 |
| Dentate nucleus | PDE8A | ENSG00000073417 | 15 | 85488671 | 85692376 | 585 | 4.946 | 3.8e-07 | 0.00733 |
| Dentate nucleus | ZSCAN23 | ENSG00000187987 | 6 | 28389707 | 28446279 | 133 | 4.945 | 3.8e-07 | 0.00734 |
| Dentate nucleus | SZT2 | ENSG00000198198 | 1 | 43820553 | 43928321 | 146 | 4.941 | 3.88e-07 | 0.00749 |
| Dentate nucleus | ZBTB3 | ENSG00000185670 | 11 | 62505791 | 62556660 | 70 | 4.941 | 3.89e-07 | 0.0075 |
| Dentate nucleus | RP11-831H9.11 | ENSG00000255432 | 11 | 62407166 | 62467650 | 73 | 4.929 | 4.13e-07 | 0.00797 |
| Dentate nucleus | CHIA | ENSG00000134216 | 1 | 111798484 | 111873188 | 385 | 4.913 | 4.48e-07 | 0.00865 |
| Dentate nucleus | ACSL1 | ENSG00000151726 | 4 | 185666749 | 185782972 | 420 | 4.913 | 4.49e-07 | 0.00867 |
| Dentate nucleus | VPS37D | ENSG00000176428 | 7 | 73047155 | 73096442 | 133 | 4.907 | 4.63e-07 | 0.00893 |
| Dentate nucleus | HIST1H4I | ENSG00000198339 | 6 | 27072076 | 27118418 | 68 | 4.906 | 4.66e-07 | 0.009 |
| Dentate nucleus | C11orf84 | ENSG00000168005 | 11 | 63545860 | 63605190 | 118 | 4.901 | 4.76e-07 | 0.00919 |
| Dentate nucleus | NENF | ENSG00000117691 | 1 | 212571229 | 212629714 | 155 | 4.889 | 5.06e-07 | 0.00978 |
| Dentate nucleus | TMPRSS12 | ENSG00000186452 | 12 | 51201703 | 51291667 | 192 | 4.875 | 5.45e-07 | 0.0105 |
| Dentate nucleus | MRC1L1 | ENSG00000183748 | 10 | 17816362 | 17963178 | 46 | 4.861 | 5.85e-07 | 0.0113 |
| Dentate nucleus | KIAA1841 | ENSG00000162929 | 2 | 61258006 | 61401960 | 256 | 4.852 | 6.11e-07 | 0.0118 |
| Dentate nucleus | PCDHGC5 | ENSG00000240764 | 5 | 140833808 | 140902546 | 147 | 4.838 | 6.57e-07 | 0.0127 |
| Dentate nucleus | DUOX2 | ENSG00000140279 | 15 | 45374848 | 45441542 | 124 | 4.835 | 6.65e-07 | 0.0128 |
| Dentate nucleus | DDR1 | ENSG00000204580 | 6 | 30809198 | 30877933 | 259 | 4.83 | 6.82e-07 | 0.0132 |
| Dentate nucleus | SLC45A3 | ENSG00000158715 | 1 | 205616979 | 205684587 | 169 | 4.829 | 6.86e-07 | 0.0132 |
| Dentate nucleus | SLC25A34 | ENSG00000162461 | 1 | 16027900 | 16077891 | 131 | 4.816 | 7.34e-07 | 0.0142 |
| Dentate nucleus | CTD-2521M24.10 | ENSG00000269035 | 19 | 17537420 | 17606613 | 197 | 4.815 | 7.36e-07 | 0.0142 |
| Dentate nucleus | PCDHGC4 | ENSG00000242419 | 5 | 140829741 | 140902546 | 155 | 4.812 | 7.48e-07 | 0.0144 |
| Dentate nucleus | VMAC | ENSG00000187650 | 19 | 5869869 | 5920864 | 199 | 4.807 | 7.67e-07 | 0.0148 |
| Dentate nucleus | TIMM23 | ENSG00000138297 | 10 | 51582080 | 51658365 | 20 | 4.8 | 7.94e-07 | 0.0153 |
| Dentate nucleus | AC104532.2 | ENSG00000267314 | 19 | 5869852 | 5924718 | 201 | 4.8 | 7.95e-07 | 0.0154 |
| Dentate nucleus | NFKBIL1 | ENSG00000204498 | 6 | 31479647 | 31536606 | 308 | 4.796 | 8.11e-07 | 0.0157 |
| Dentate nucleus | IER3 | ENSG00000137331 | 6 | 30700976 | 30747331 | 231 | 4.792 | 8.27e-07 | 0.016 |
| Dentate nucleus | NDUFA11 | ENSG00000174886 | 19 | 5881287 | 5939017 | 161 | 4.791 | 8.31e-07 | 0.016 |
| Dentate nucleus | NT5C2 | ENSG00000076685 | 10 | 104835940 | 104988056 | 313 | 4.789 | 8.39e-07 | 0.0162 |
| Dentate nucleus | ARNTL2 | ENSG00000029153 | 12 | 27450787 | 27586241 | 352 | 4.779 | 8.8e-07 | 0.017 |
| Dentate nucleus | PDCD11 | ENSG00000148843 | 10 | 105121405 | 105216049 | 120 | 4.771 | 9.18e-07 | 0.0177 |
| Dentate nucleus | CPED1 | ENSG00000106034 | 7 | 120593731 | 120947498 | 760 | 4.766 | 9.41e-07 | 0.0182 |
| Dentate nucleus | SLC27A1 | ENSG00000130304 | 19 | 17544578 | 17626977 | 238 | 4.762 | 9.59e-07 | 0.0185 |
| Dentate nucleus | PAN3 | ENSG00000152520 | 13 | 28677643 | 28879475 | 306 | 4.744 | 1.05e-06 | 0.0203 |
| Dentate nucleus | C17orf53 | ENSG00000125319 | 17 | 42184274 | 42249844 | 103 | 4.743 | 1.05e-06 | 0.0203 |
| Dentate nucleus | PCDHGC3 | ENSG00000240184 | 5 | 140820580 | 140902542 | 179 | 4.733 | 1.11e-06 | 0.0214 |
| Dentate nucleus | POU5F1 | ENSG00000204531 | 6 | 31122119 | 31183508 | 733 | 4.72 | 1.18e-06 | 0.0227 |
| Dentate nucleus | PGLS | ENSG00000130313 | 19 | 17587438 | 17642097 | 164 | 4.707 | 1.25e-06 | 0.0242 |
| Dentate nucleus | MACROD1 | ENSG00000133315 | 11 | 63756030 | 63968578 | 430 | 4.707 | 1.26e-06 | 0.0243 |
| Dentate nucleus | FAM228A | ENSG00000186453 | 2 | 24362938 | 24433718 | 216 | 4.702 | 1.29e-06 | 0.0249 |
| Dentate nucleus | H3F3B | ENSG00000132475 | 17 | 73762515 | 73816974 | 110 | 4.698 | 1.31e-06 | 0.0253 |
| Dentate nucleus | MYO15A | ENSG00000091536 | 17 | 17977020 | 18093116 | 186 | 4.696 | 1.32e-06 | 0.0256 |
| Dentate nucleus | TMEM82 | ENSG00000162460 | 1 | 16033917 | 16084477 | 131 | 4.685 | 1.4e-06 | 0.027 |
| Dentate nucleus | GFAP | ENSG00000131095 | 17 | 42972376 | 43029305 | 164 | 4.68 | 1.43e-06 | 0.0277 |
| Dentate nucleus | ASGR1 | ENSG00000141505 | 17 | 7066750 | 7117883 | 89 | 4.678 | 1.45e-06 | 0.0279 |
| Dentate nucleus | ADAMTSL4-AS1 | ENSG00000203804 | 1 | 150523480 | 150568969 | 84 | 4.677 | 1.45e-06 | 0.0281 |
| Dentate nucleus | PPP2R5A | ENSG00000066027 | 1 | 212423879 | 212545200 | 288 | 4.677 | 1.46e-06 | 0.0281 |
| Dentate nucleus | CPLX1 | ENSG00000168993 | 4 | 768745 | 854986 | 360 | 4.669 | 1.52e-06 | 0.0293 |

| Trait | Gene symbol | Ensembl gene ID | Chromosome | Start (bp) | Stop (bp) | SNPs (n) | Z | P | Bonferroni-adjusted P |
|---|---|---|---|---|---|---|---|---|---|
| Dentate nucleus | RP11-144F15.1 | ENSG00000257545 | 12 | 106879736 | 107203696 | 629 | 4.666 | 1.54e-06 | 0.0297 |
| Dentate nucleus | RPL31 | ENSG00000071082 | 2 | 101583177 | 101650494 | 154 | 4.652 | 1.64e-06 | 0.0317 |
| Dentate nucleus | MAVS | ENSG00000088888 | 20 | 3792487 | 3859280 | 186 | 4.649 | 1.67e-06 | 0.0322 |
| Dentate nucleus | FAM160A1 | ENSG00000164142 | 4 | 152295368 | 152594784 | 564 | 4.647 | 1.69e-06 | 0.0325 |
| Dentate nucleus | MOG | ENSG00000204655 | 6 | 29589758 | 29650149 | 265 | 4.643 | 1.72e-06 | 0.0331 |
| Dentate nucleus | ZNF160 | ENSG00000170949 | 19 | 53559859 | 53641689 | 369 | 4.642 | 1.72e-06 | 0.0333 |
| Dentate nucleus | AC024592.12 | ENSG00000267740 | 19 | 5856182 | 5938798 | 281 | 4.624 | 1.88e-06 | 0.0364 |
| Dentate nucleus | GABBR1 | ENSG00000204681 | 6 | 29513406 | 29636753 | 422 | 4.622 | 1.9e-06 | 0.0368 |
| Dentate nucleus | PCDHGB7 | ENSG00000254122 | 5 | 140762427 | 140902546 | 297 | 4.617 | 1.95e-06 | 0.0376 |
| Dentate nucleus | CASP9 | ENSG00000132906 | 1 | 15807327 | 15888029 | 234 | 4.615 | 1.97e-06 | 0.038 |
| Dentate nucleus | HDAC5 | ENSG00000108840 | 17 | 42144114 | 42236070 | 137 | 4.611 | 2.01e-06 | 0.0387 |
| Dentate nucleus | DUOXA1 | ENSG00000140254 | 15 | 45399569 | 45457136 | 114 | 4.609 | 2.02e-06 | 0.039 |
| Dentate nucleus | DEFB123 | ENSG00000180424 | 20 | 29993322 | 30048060 | 139 | 4.607 | 2.04e-06 | 0.0395 |
| Dentate nucleus | STON1-GTF2A1L | ENSG00000068781 | 2 | 48722064 | 49013654 | 939 | 4.585 | 2.27e-06 | 0.0437 |
| Dentate nucleus | PCDHGA1 | ENSG00000204956 | 5 | 140675252 | 140902546 | 500 | 4.582 | 2.3e-06 | 0.0444 |
| Dentate nucleus | C16orf95 | ENSG00000260456 | 16 | 87107168 | 87386022 | 1071 | 4.581 | 2.31e-06 | 0.0446 |
| Dentate nucleus | C6orf100 | ENSG00000204709 | 6 | 28876654 | 28922314 | 152 | 4.579 | 2.33e-06 | 0.045 |
| Dentate nucleus | OR14J1 | ENSG00000204695 | 6 | 29239403 | 29285519 | 142 | 4.575 | 2.38e-06 | 0.046 |
| Dentate nucleus | PCDHGA9 | ENSG00000261934 | 5 | 140747520 | 140902546 | 329 | 4.568 | 2.46e-06 | 0.0475 |
| Dentate nucleus | TMUB2 | ENSG00000168591 | 17 | 42229338 | 42279099 | 76 | 4.562 | 2.53e-06 | 0.0489 |
| Globus pallidus | BANK1 | ENSG00000153064 | 4 | 102297443 | 103005969 | 1655 | 9.163 | 2.53e-20 | 4.88e-16 |
| Globus pallidus | SLC39A8 | ENSG00000138821 | 4 | 103162198 | 103387415 | 653 | 8.674 | 2.09e-18 | 4.04e-14 |
| Globus pallidus | COL5A2 | ENSG00000204262 | 2 | 189886622 | 190079605 | 456 | 8.302 | 5.14e-17 | 9.92e-13 |
| Globus pallidus | MRC1L1 | ENSG00000183748 | 10 | 17816362 | 17963178 | 46 | 7.763 | 4.16e-15 | 8.04e-11 |
| Globus pallidus | SLC39A12 | ENSG00000148482 | 10 | 18205768 | 18342221 | 469 | 7.727 | 5.52e-15 | 1.07e-10 |
| Globus pallidus | HIGD1C | ENSG00000214511 | 12 | 51312705 | 51374289 | 197 | 7.65 | 1e-14 | 1.94e-10 |
| Globus pallidus | LETMD1 | ENSG00000050426 | 12 | 51406745 | 51464207 | 134 | 7.64 | 1.08e-14 | 2.09e-10 |
| Globus pallidus | SRPRB | ENSG00000144867 | 3 | 133467877 | 133554616 | 218 | 7.508 | 3e-14 | 5.8e-10 |
| Globus pallidus | HFE | ENSG00000010704 | 6 | 26052509 | 26108571 | 134 | 7.306 | 1.38e-13 | 2.66e-09 |
| Globus pallidus | SLC11A2 | ENSG00000110911 | 12 | 51363184 | 51457349 | 233 | 7.15 | 4.33e-13 | 8.36e-09 |
| Globus pallidus | HIST1H4C | ENSG00000197061 | 6 | 26069104 | 26114518 | 128 | 7.123 | 5.29e-13 | 1.02e-08 |
| Globus pallidus | PSORS1C1 | ENSG00000204540 | 6 | 31047527 | 31117869 | 731 | 6.961 | 1.69e-12 | 3.26e-08 |
| Globus pallidus | HIST1H4A | ENSG00000196176 | 6 | 25986907 | 26032278 | 79 | 6.956 | 1.75e-12 | 3.38e-08 |
| Globus pallidus | HIST1H2BB | ENSG00000196226 | 6 | 26033455 | 26078885 | 94 | 6.944 | 1.9e-12 | 3.67e-08 |
| Globus pallidus | HIST1H3A | ENSG00000198366 | 6 | 25985718 | 26031186 | 80 | 6.929 | 2.12e-12 | 4.09e-08 |
| Globus pallidus | BCL7B | ENSG00000106635 | 7 | 72940686 | 73007332 | 124 | 6.915 | 2.33e-12 | 4.5e-08 |
| Globus pallidus | CSRNP2 | ENSG00000110925 | 12 | 51444990 | 51512447 | 166 | 6.907 | 2.48e-12 | 4.79e-08 |
| Globus pallidus | HIST1H1C | ENSG00000187837 | 6 | 26045968 | 26091699 | 101 | 6.888 | 2.84e-12 | 5.48e-08 |
| Globus pallidus | HIST1H1A | ENSG00000124610 | 6 | 26007260 | 26053040 | 93 | 6.828 | 4.3e-12 | 8.31e-08 |
| Globus pallidus | TFRC | ENSG00000072274 | 3 | 195744054 | 195844060 | 428 | 6.816 | 4.69e-12 | 9.06e-08 |
| Globus pallidus | AP003733.1 | ENSG00000269089 | 11 | 61700453 | 61746755 | 116 | 6.807 | 4.99e-12 | 9.62e-08 |
| Globus pallidus | TMEM236 | ENSG00000184040 | 10 | 18006218 | 18099855 | 10 | 6.798 | 5.32e-12 | 1.03e-07 |
| Globus pallidus | TF | ENSG00000091513 | 3 | 133429800 | 133507850 | 256 | 6.792 | 5.54e-12 | 1.07e-07 |
| Globus pallidus | SCGN | ENSG00000079689 | 6 | 25617464 | 25712011 | 282 | 6.779 | 6.04e-12 | 1.17e-07 |
| Globus pallidus | HIST1H4B | ENSG00000124529 | 6 | 26017124 | 26062480 | 89 | 6.694 | 1.09e-11 | 2.1e-07 |
| Globus pallidus | HIST1H3C | ENSG00000196532 | 6 | 26010639 | 26056097 | 90 | 6.668 | 1.3e-11 | 2.51e-07 |
| Globus pallidus | HIST1H2AB | ENSG00000137259 | 6 | 26023320 | 26068796 | 82 | 6.649 | 1.48e-11 | 2.85e-07 |
| Globus pallidus | HIST1H3B | ENSG00000124693 | 6 | 26021817 | 26067288 | 86 | 6.648 | 1.48e-11 | 2.86e-07 |
| Globus pallidus | CDSN | ENSG00000204539 | 6 | 31072867 | 31123223 | 522 | 6.61 | 1.92e-11 | 3.72e-07 |
| Globus pallidus | HLA-B | ENSG00000234745 | 6 | 31311649 | 31359965 | 1542 | 6.552 | 2.84e-11 | 5.48e-07 |
| Globus pallidus | SLC17A3 | ENSG00000124564 | 6 | 25823294 | 25917514 | 233 | 6.504 | 3.92e-11 | 7.56e-07 |
| Globus pallidus | BAZ1B | ENSG00000009954 | 7 | 72844728 | 72971608 | 174 | 6.479 | 4.61e-11 | 8.89e-07 |

| Trait | Gene symbol | Ensembl gene ID | Chromosome | Start (bp) | Stop (bp) | SNPs (n) | Z | P | Bonferroni-adjusted P |
|---|---|---|---|---|---|---|---|---|---|
| Globus pallidus | C2 | ENSG00000166278 | 6 | 31830562 | 31923449 | 240 | 6.443 | 5.87e-11 | 1.13e-06 |
| Globus pallidus | C6orf15 | ENSG00000204542 | 6 | 31069000 | 31115336 | 511 | 6.44 | 5.95e-11 | 1.15e-06 |
| Globus pallidus | C4A | ENSG00000244731 | 6 | 31914801 | 31980458 | 151 | 6.385 | 8.56e-11 | 1.65e-06 |
| Globus pallidus | TBL2 | ENSG00000106638 | 7 | 72973262 | 73028121 | 117 | 6.36 | 1.01e-10 | 1.95e-06 |
| Globus pallidus | ALKBH5 | ENSG00000091542 | 17 | 18051392 | 18123268 | 131 | 6.337 | 1.17e-10 | 2.26e-06 |
| Globus pallidus | LRRC16A | ENSG00000079691 | 6 | 25244306 | 25630758 | 1409 | 6.323 | 1.28e-10 | 2.47e-06 |
| Globus pallidus | NFKBIL1 | ENSG00000204498 | 6 | 31479647 | 31536606 | 308 | 6.308 | 1.42e-10 | 2.74e-06 |
| Globus pallidus | MCCD1 | ENSG00000204511 | 6 | 31461494 | 31508009 | 380 | 6.301 | 1.48e-10 | 2.86e-06 |
| Globus pallidus | DXO | ENSG00000204348 | 6 | 31927587 | 31975069 | 93 | 6.293 | 1.56e-10 | 3.01e-06 |
| Globus pallidus | LLGL1 | ENSG00000131899 | 17 | 18093901 | 18158189 | 135 | 6.293 | 1.56e-10 | 3.01e-06 |
| Globus pallidus | DDX39B | ENSG00000198563 | 6 | 31487996 | 31545225 | 269 | 6.284 | 1.65e-10 | 3.19e-06 |
| Globus pallidus | MLXIPL | ENSG00000009950 | 7 | 72997524 | 73073873 | 203 | 6.259 | 1.94e-10 | 3.74e-06 |
| Globus pallidus | ATP6V1G2-DDX39B | ENSG00000254870 | 6 | 31487996 | 31549385 | 278 | 6.248 | 2.08e-10 | 4.02e-06 |
| Globus pallidus | ATP6V1G2 | ENSG00000213760 | 6 | 31502239 | 31551204 | 209 | 6.242 | 2.16e-10 | 4.17e-06 |
| Globus pallidus | FZD9 | ENSG00000188763 | 7 | 72813109 | 72860450 | 69 | 6.228 | 2.36e-10 | 4.56e-06 |
| Globus pallidus | TRIM38 | ENSG00000112343 | 6 | 25928030 | 25997384 | 160 | 6.179 | 3.23e-10 | 6.24e-06 |
| Globus pallidus | LTA | ENSG00000226979 | 6 | 31504831 | 31552101 | 198 | 6.161 | 3.62e-10 | 6.98e-06 |
| Globus pallidus | BEST1 | ENSG00000167995 | 11 | 61682293 | 61742987 | 146 | 6.156 | 3.72e-10 | 7.19e-06 |
| Globus pallidus | COL3A1 | ENSG00000168542 | 2 | 189804046 | 189887472 | 170 | 6.149 | 3.89e-10 | 7.51e-06 |
| Globus pallidus | ZBTB12 | ENSG00000204366 | 6 | 31857384 | 31904769 | 116 | 6.144 | 4.03e-10 | 7.77e-06 |
| Globus pallidus | PRRC2A | ENSG00000204469 | 6 | 31553497 | 31615548 | 221 | 6.119 | 4.72e-10 | 9.11e-06 |
| Globus pallidus | WDR75 | ENSG00000115368 | 2 | 190271159 | 190350291 | 241 | 6.109 | 5e-10 | 9.65e-06 |
| Globus pallidus | SLC40A1 | ENSG00000138449 | 2 | 190415305 | 190483484 | 125 | 6.109 | 5e-10 | 9.65e-06 |
| Globus pallidus | PABPC1 | ENSG00000070756 | 8 | 101688044 | 101770037 | 150 | 6.095 | 5.46e-10 | 1.05e-05 |
| Globus pallidus | NELFE | ENSG00000204356 | 6 | 31909864 | 31961887 | 114 | 6.065 | 6.6e-10 | 1.27e-05 |
| Globus pallidus | SLC20A2 | ENSG00000168575 | 8 | 42263993 | 42432069 | 324 | 6.062 | 6.73e-10 | 1.3e-05 |
| Globus pallidus | AL645922.1 | ENSG00000268923 | 6 | 31963945 | 32009881 | 88 | 6.043 | 7.57e-10 | 1.46e-05 |
| Globus pallidus | CFB | ENSG00000244255 | 6 | 31860475 | 31929825 | 169 | 6.037 | 7.86e-10 | 1.52e-05 |
| Globus pallidus | CFB | ENSG00000243649 | 6 | 31860475 | 31929861 | 169 | 6.037 | 7.86e-10 | 1.52e-05 |
| Globus pallidus | TNK2 | ENSG00000061938 | 3 | 195580235 | 195673816 | 298 | 6.017 | 8.89e-10 | 1.72e-05 |
| Globus pallidus | PTPMT1 | ENSG00000110536 | 11 | 47551982 | 47605013 | 73 | 5.995 | 1.02e-09 | 1.96e-05 |
| Globus pallidus | METTL7A | ENSG00000185432 | 12 | 51282255 | 51336300 | 120 | 5.983 | 1.1e-09 | 2.12e-05 |
| Globus pallidus | STK19 | ENSG00000204344 | 6 | 31903868 | 31960598 | 126 | 5.942 | 1.41e-09 | 2.71e-05 |
| Globus pallidus | KBTBD4 | ENSG00000231880 | 11 | 47564277 | 47609823 | 60 | 5.927 | 1.54e-09 | 2.97e-05 |
| Globus pallidus | SLC17A2 | ENSG00000112337 | 6 | 25902982 | 25965946 | 182 | 5.922 | 1.59e-09 | 3.06e-05 |
| Globus pallidus | SLC17A1 | ENSG00000124568 | 6 | 25773125 | 25867287 | 232 | 5.919 | 1.62e-09 | 3.12e-05 |
| Globus pallidus | NDUFS3 | ENSG00000213619 | 11 | 47551888 | 47616114 | 84 | 5.919 | 1.62e-09 | 3.13e-05 |
| Globus pallidus | EHMT2 | ENSG00000204371 | 6 | 31837536 | 31900464 | 171 | 5.902 | 1.8e-09 | 3.47e-05 |
| Globus pallidus | LST1 | ENSG00000204482 | 6 | 31518901 | 31566686 | 189 | 5.877 | 2.09e-09 | 4.04e-05 |
| Globus pallidus | TNF | ENSG00000232810 | 6 | 31508344 | 31556113 | 183 | 5.873 | 2.14e-09 | 4.13e-05 |
| Globus pallidus | CCHCR1 | ENSG00000204536 | 6 | 31100216 | 31161015 | 609 | 5.858 | 2.34e-09 | 4.52e-05 |
| Globus pallidus | FTH1 | ENSG00000167996 | 11 | 61717190 | 61770132 | 169 | 5.84 | 2.61e-09 | 5.04e-05 |
| Globus pallidus | SKIV2L | ENSG00000204351 | 6 | 31891857 | 31947532 | 157 | 5.839 | 2.62e-09 | 5.06e-05 |
| Globus pallidus | ATF6B | ENSG00000213676 | 6 | 32055953 | 32131030 | 141 | 5.821 | 2.92e-09 | 5.63e-05 |
| Globus pallidus | C4B | ENSG00000224389 | 6 | 31947539 | 32013195 | 109 | 5.791 | 3.49e-09 | 6.74e-05 |
| Globus pallidus | FAM180B | ENSG00000196666 | 11 | 47573198 | 47620746 | 45 | 5.766 | 4.05e-09 | 7.83e-05 |
| Globus pallidus | PSORS1C2 | ENSG00000204538 | 6 | 31095313 | 31142127 | 397 | 5.743 | 4.66e-09 | 8.99e-05 |
| Globus pallidus | APOM | ENSG00000204444 | 6 | 31585193 | 31635987 | 157 | 5.743 | 4.66e-09 | 8.99e-05 |
| Globus pallidus | RAB3IL1 | ENSG00000167994 | 11 | 61654773 | 61722741 | 156 | 5.7 | 5.98e-09 | 0.000116 |
| Globus pallidus | BAIAP2L1 | ENSG00000006453 | 7 | 97910963 | 98065380 | 334 | 5.681 | 6.69e-09 | 0.000129 |
| Globus pallidus | PRUNE | ENSG00000143363 | 1 | 150945896 | 151018189 | 159 | 5.675 | 6.94e-09 | 0.000134 |
| Globus pallidus | CSNK2B | ENSG00000204435 | 6 | 31598013 | 31648120 | 143 | 5.665 | 7.35e-09 | 0.000142 |

| Trait | Gene symbol | Ensembl gene ID | Chromosome | Start (bp) | Stop (bp) | SNPs (n) | Z | P | Bonferroni-adjusted P |
|---|---|---|---|---|---|---|---|---|---|
| Globus pallidus | FKBPL | ENSG00000204315 | 6 | 32086484 | 32133068 | 92 | 5.646 | 8.23e-09 | 0.000159 |
| Globus pallidus | TCF19 | ENSG00000137310 | 6 | 31091319 | 31144936 | 481 | 5.608 | 1.02e-08 | 0.000198 |
| Globus pallidus | BAG6 | ENSG00000204463 | 6 | 31596805 | 31655482 | 155 | 5.608 | 1.03e-08 | 0.000198 |
| Globus pallidus | LY6G5B | ENSG00000240053 | 6 | 31602944 | 31651553 | 129 | 5.607 | 1.03e-08 | 0.000199 |
| Globus pallidus | CSNK2B-LY6G5B-1181 | ENSG00000263020 | 6 | 31598879 | 31651323 | 142 | 5.593 | 1.11e-08 | 0.000215 |
| Globus pallidus | LTB | ENSG00000227507 | 6 | 31538302 | 31585299 | 160 | 5.573 | 1.25e-08 | 0.000242 |
| Globus pallidus | HIST1H2BA | ENSG00000146047 | 6 | 25692137 | 25737573 | 144 | 5.536 | 1.55e-08 | 0.000299 |
| Globus pallidus | SMIM19 | ENSG00000176209 | 8 | 42361298 | 42419603 | 127 | 5.519 | 1.7e-08 | 0.000329 |
| Globus pallidus | MIEF2 | ENSG00000177427 | 17 | 18128848 | 18179866 | 92 | 5.511 | 1.79e-08 | 0.000345 |
| Globus pallidus | FAM63A | ENSG00000143409 | 1 | 150959025 | 151015851 | 127 | 5.493 | 1.97e-08 | 0.000381 |
| Globus pallidus | CELF1 | ENSG00000149187 | 11 | 47477496 | 47622121 | 153 | 5.491 | 2e-08 | 0.000386 |
| Globus pallidus | FNBP4 | ENSG00000109920 | 11 | 47728072 | 47823995 | 161 | 5.487 | 2.04e-08 | 0.000394 |
| Globus pallidus | SLC39A13 | ENSG00000165915 | 11 | 47393683 | 47448047 | 115 | 5.487 | 2.05e-08 | 0.000395 |
| Globus pallidus | AIF1 | ENSG00000204472 | 6 | 31547961 | 31594798 | 159 | 5.482 | 2.1e-08 | 0.000406 |
| Globus pallidus | MICB | ENSG00000204516 | 6 | 31427658 | 31488901 | 652 | 5.479 | 2.14e-08 | 0.000414 |
| Globus pallidus | XPO4 | ENSG00000132953 | 13 | 21341469 | 21512187 | 383 | 5.459 | 2.39e-08 | 0.000462 |
| Globus pallidus | MICA | ENSG00000204520 | 6 | 31336356 | 31393092 | 1062 | 5.417 | 3.03e-08 | 0.000584 |
| Globus pallidus | CYP21A2 | ENSG00000231852 | 6 | 31971042 | 32019447 | 117 | 5.409 | 3.17e-08 | 0.000612 |
| Globus pallidus | TFCP2 | ENSG00000135457 | 12 | 51477446 | 51601926 | 308 | 5.398 | 3.38e-08 | 0.000652 |
| Globus pallidus | NCR3 | ENSG00000204475 | 6 | 31546672 | 31595762 | 168 | 5.387 | 3.58e-08 | 0.000691 |
| Globus pallidus | MYBPC3 | ENSG00000134571 | 11 | 47342957 | 47409253 | 149 | 5.387 | 3.58e-08 | 0.000692 |
| Globus pallidus | DBI | ENSG00000155368 | 2 | 120089497 | 120140126 | 163 | 5.383 | 3.66e-08 | 0.000707 |
| Globus pallidus | SPI1 | ENSG00000066336 | 11 | 47366411 | 47435127 | 156 | 5.378 | 3.76e-08 | 0.000725 |
| Globus pallidus | SLC17A4 | ENSG00000146039 | 6 | 25719927 | 25791419 | 244 | 5.368 | 3.99e-08 | 0.00077 |
| Globus pallidus | PPT2 | ENSG00000221988 | 6 | 32086218 | 32144011 | 113 | 5.352 | 4.35e-08 | 0.00084 |
| Globus pallidus | SNX31 | ENSG00000174226 | 8 | 101575116 | 101710643 | 447 | 5.333 | 4.84e-08 | 0.000934 |
| Globus pallidus | HLA-C | ENSG00000204525 | 6 | 31226526 | 31274907 | 1493 | 5.332 | 4.86e-08 | 0.000938 |
| Globus pallidus | EGFL8 | ENSG00000241404 | 6 | 32097360 | 32146058 | 97 | 5.289 | 6.16e-08 | 0.00119 |
| Globus pallidus | NUP160 | ENSG00000030066 | 11 | 47789639 | 47905107 | 188 | 5.263 | 7.09e-08 | 0.00137 |
| Globus pallidus | TNXB | ENSG00000168477 | 6 | 31998931 | 32118111 | 252 | 5.21 | 9.45e-08 | 0.00182 |
| Globus pallidus | MTCH2 | ENSG00000109919 | 11 | 47628867 | 47699175 | 74 | 5.207 | 9.58e-08 | 0.00185 |
| Globus pallidus | HIST1H2AA | ENSG00000164508 | 6 | 25716291 | 25761790 | 164 | 5.2 | 9.99e-08 | 0.00193 |
| Globus pallidus | C16orf95 | ENSG00000260456 | 16 | 87107168 | 87386022 | 1071 | 5.182 | 1.1e-07 | 0.00212 |
| Globus pallidus | DUOX1 | ENSG00000137857 | 15 | 45387131 | 45467774 | 158 | 5.177 | 1.13e-07 | 0.00218 |
| Globus pallidus | AGBL2 | ENSG00000165923 | 11 | 47671143 | 47771941 | 155 | 5.167 | 1.19e-07 | 0.0023 |
| Globus pallidus | C2orf76 | ENSG00000186132 | 2 | 120049801 | 120159404 | 351 | 5.116 | 1.56e-07 | 0.00301 |
| Globus pallidus | POU5F1 | ENSG00000204531 | 6 | 31122119 | 31183508 | 733 | 5.113 | 1.59e-07 | 0.00307 |
| Globus pallidus | KCTD17 | ENSG00000100379 | 22 | 37412779 | 37469430 | 102 | 5.11 | 1.61e-07 | 0.00311 |
| Globus pallidus | KBTBD4 | ENSG00000123444 | 11 | 47583749 | 47635567 | 53 | 5.096 | 1.73e-07 | 0.00334 |
| Globus pallidus | FNIP2 | ENSG00000052795 | 4 | 159655290 | 159839201 | 245 | 5.078 | 1.9e-07 | 0.00367 |
| Globus pallidus | PPT2-EGFL8 | ENSG00000258388 | 6 | 32086622 | 32149755 | 122 | 5.07 | 1.99e-07 | 0.00384 |
| Globus pallidus | BRI3 | ENSG00000164713 | 7 | 97846691 | 97947162 | 240 | 5.067 | 2.02e-07 | 0.0039 |
| Globus pallidus | AP2A2 | ENSG00000183020 | 11 | 889894 | 1022239 | 396 | 5.064 | 2.06e-07 | 0.00397 |
| Globus pallidus | CHID1 | ENSG00000177830 | 11 | 857357 | 950058 | 236 | 5.055 | 2.15e-07 | 0.00415 |
| Globus pallidus | BNIPL | ENSG00000163141 | 1 | 150974046 | 151030076 | 111 | 5.03 | 2.45e-07 | 0.00472 |
| Globus pallidus | DUOX2 | ENSG00000140279 | 15 | 45374848 | 45441542 | 124 | 5.009 | 2.74e-07 | 0.00529 |
| Globus pallidus | VPS37D | ENSG00000176428 | 7 | 73047155 | 73096442 | 133 | 4.989 | 3.03e-07 | 0.00585 |
| Globus pallidus | C1QTNF4 | ENSG00000172247 | 11 | 47601216 | 47651211 | 47 | 4.98 | 3.18e-07 | 0.00614 |
| Globus pallidus | ACSL1 | ENSG00000151726 | 4 | 185666749 | 185782972 | 420 | 4.966 | 3.42e-07 | 0.00661 |
| Globus pallidus | ALPL | ENSG00000162551 | 1 | 21800858 | 21914905 | 469 | 4.962 | 3.49e-07 | 0.00674 |
| Globus pallidus | C1orf56 | ENSG00000143443 | 1 | 150985216 | 151034462 | 97 | 4.95 | 3.72e-07 | 0.00717 |
| Globus pallidus | C6orf47 | ENSG00000204439 | 6 | 31616075 | 31663549 | 102 | 4.912 | 4.5e-07 | 0.00869 |

| Trait | Gene symbol | Ensembl gene ID | Chromosome | Start (bp) | Stop (bp) | SNPs (n) | Z | P | Bonferroni-adjusted P |
|---|---|---|---|---|---|---|---|---|---|
| Globus pallidus | HLA-DRB5 | ENSG00000198502 | 6 | 32475120 | 32533064 | 2421 | 4.889 | 5.07e-07 | 0.00979 |
| Globus pallidus | PRRT1 | ENSG00000204314 | 6 | 32106136 | 32157150 | 103 | 4.83 | 6.82e-07 | 0.0132 |
| Globus pallidus | HSPA1L | ENSG00000204390 | 6 | 31767396 | 31818437 | 128 | 4.829 | 6.85e-07 | 0.0132 |
| Globus pallidus | OR2B6 | ENSG00000124657 | 6 | 27890019 | 27935960 | 61 | 4.829 | 6.85e-07 | 0.0132 |
| Globus pallidus | PPID | ENSG00000171497 | 4 | 159620286 | 159679548 | 76 | 4.781 | 8.73e-07 | 0.0169 |
| Globus pallidus | SLC44A4 | ENSG00000204385 | 6 | 31820969 | 31881823 | 154 | 4.78 | 8.77e-07 | 0.0169 |
| Globus pallidus | MYO15A | ENSG00000091536 | 17 | 17977020 | 18093116 | 186 | 4.777 | 8.88e-07 | 0.0171 |
| Globus pallidus | DDHD1 | ENSG00000100523 | 14 | 53500686 | 53655000 | 259 | 4.776 | 8.95e-07 | 0.0173 |
| Globus pallidus | TECPR1 | ENSG00000205356 | 7 | 97833936 | 97916563 | 192 | 4.767 | 9.37e-07 | 0.0181 |
| Globus pallidus | C6orf48 | ENSG00000204387 | 6 | 31767385 | 31817541 | 124 | 4.758 | 9.8e-07 | 0.0189 |
| Globus pallidus | HSPA1B | ENSG00000204388 | 6 | 31760512 | 31808031 | 115 | 4.735 | 1.1e-06 | 0.0212 |

**Supplementary Table 4. Significant genes from MAGMA gene-based analysis of quantitative susceptibility mapping-derived brain iron across six subcortical regions (Bonferroni-corrected P < 2.59 x 10-6).** ***(continued)***

| Trait | Gene symbol | Ensembl gene ID | Chromosome | Start (bp) | Stop (bp) | SNPs (n) | Z | P | Bonferroni-adjusted P |
|---|---|---|---|---|---|---|---|---|---|
| Globus pallidus | VARS | ENSG00000204394 | 6 | 31735295 | 31798730 | 124 | 4.718 | 1.19e-06 | 0.023 |
| Globus pallidus | DUOXA2 | ENSG00000140274 | 15 | 45371519 | 45420619 | 100 | 4.695 | 1.33e-06 | 0.0258 |
| Globus pallidus | MUC22 | ENSG00000261272 | 6 | 30943251 | 31013179 | 642 | 4.692 | 1.35e-06 | 0.0261 |
| Globus pallidus | IL17D | ENSG00000172458 | 13 | 21241266 | 21307237 | 166 | 4.684 | 1.41e-06 | 0.0272 |
| Globus pallidus | HCG27 | ENSG00000206344 | 6 | 31130537 | 31181745 | 671 | 4.677 | 1.45e-06 | 0.0281 |
| Globus pallidus | DDR1 | ENSG00000204580 | 6 | 30809198 | 30877933 | 259 | 4.672 | 1.49e-06 | 0.0287 |
| Globus pallidus | LSM2 | ENSG00000204392 | 6 | 31755173 | 31809761 | 124 | 4.648 | 1.68e-06 | 0.0324 |
| Globus pallidus | RITA1 | ENSG00000139405 | 12 | 113588331 | 113640173 | 87 | 4.641 | 1.73e-06 | 0.0334 |
| Globus pallidus | RNF5 | ENSG00000204308 | 6 | 32111131 | 32161930 | 103 | 4.641 | 1.73e-06 | 0.0335 |
| Globus pallidus | DUOXA1 | ENSG00000140254 | 15 | 45399569 | 45457136 | 114 | 4.62 | 1.92e-06 | 0.0371 |
| Globus pallidus | GARNL3 | ENSG00000136895 | 9 | 129951544 | 130165939 | 290 | 4.614 | 1.98e-06 | 0.0382 |
| Globus pallidus | RNF24 | ENSG00000101236 | 20 | 3897956 | 4031229 | 323 | 4.609 | 2.02e-06 | 0.039 |
| Globus pallidus | AGPAT1 | ENSG00000204310 | 6 | 32125989 | 32180873 | 136 | 4.608 | 2.03e-06 | 0.0392 |
| Globus pallidus | MAFK | ENSG00000198517 | 7 | 1535350 | 1592679 | 213 | 4.597 | 2.15e-06 | 0.0414 |
| Globus pallidus | GPANK1 | ENSG00000204438 | 6 | 31619006 | 31669060 | 104 | 4.588 | 2.23e-06 | 0.0431 |
| Globus pallidus | DDX54 | ENSG00000123064 | 12 | 113584979 | 113658284 | 117 | 4.588 | 2.23e-06 | 0.0431 |
| Globus pallidus | HLA-DQA1 | ENSG00000196735 | 6 | 32560956 | 32624839 | 3515 | 4.58 | 2.33e-06 | 0.0449 |
| Globus pallidus | HSPA1A | ENSG00000204389 | 6 | 31748291 | 31795723 | 97 | 4.57 | 2.43e-06 | 0.047 |
| Putamen | LRRC16A | ENSG00000079691 | 6 | 25244306 | 25630758 | 1409 | 9.903 | 2.02e-23 | 3.91e-19 |
| Putamen | SLC25A37 | ENSG00000147454 | 8 | 23351318 | 23442976 | 425 | 9.108 | 4.21e-20 | 8.13e-16 |
| Putamen | SLC39A12 | ENSG00000148482 | 10 | 18205768 | 18342221 | 469 | 8.512 | 8.54e-18 | 1.65e-13 |
| Putamen | ABLIM1 | ENSG00000099204 | 10 | 116180872 | 116479762 | 787 | 8.495 | 9.91e-18 | 1.91e-13 |
| Putamen | FP15737 | ENSG00000215298 | 8 | 23395157 | 23442974 | 291 | 8.457 | 1.37e-17 | 2.64e-13 |
| Putamen | TBL2 | ENSG00000106638 | 7 | 72973262 | 73028121 | 117 | 8.099 | 2.78e-16 | 5.36e-12 |
| Putamen | HIST1H2AC | ENSG00000180573 | 6 | 26089373 | 26149344 | 158 | 7.901 | 1.39e-15 | 2.68e-11 |
| Putamen | SLC17A1 | ENSG00000124568 | 6 | 25773125 | 25867287 | 232 | 7.886 | 1.55e-15 | 3e-11 |
| Putamen | KANSL1 | ENSG00000120071 | 17 | 44097282 | 44337733 | 978 | 7.876 | 1.69e-15 | 3.26e-11 |
| Putamen | DHX40 | ENSG00000108406 | 17 | 57607886 | 57695706 | 92 | 7.789 | 3.39e-15 | 6.54e-11 |
| Putamen | VPS37D | ENSG00000176428 | 7 | 73047155 | 73096442 | 133 | 7.715 | 6.05e-15 | 1.17e-10 |
| Putamen | PTRF | ENSG00000177469 | 17 | 40544470 | 40610535 | 170 | 7.694 | 7.11e-15 | 1.37e-10 |
| Putamen | SCGN | ENSG00000079689 | 6 | 25617464 | 25712011 | 282 | 7.692 | 7.25e-15 | 1.4e-10 |
| Putamen | RUNX2 | ENSG00000124813 | 6 | 45260894 | 45642086 | 964 | 7.671 | 8.54e-15 | 1.65e-10 |
| Putamen | SEMA3G | ENSG00000010319 | 3 | 52457069 | 52514101 | 85 | 7.656 | 9.6e-15 | 1.85e-10 |
| Putamen | TFCP2 | ENSG00000135457 | 12 | 51477446 | 51601926 | 308 | 7.643 | 1.06e-14 | 2.05e-10 |
| Putamen | MAPT | ENSG00000186868 | 17 | 43936748 | 44115700 | 910 | 7.618 | 1.29e-14 | 2.48e-10 |
| Putamen | STAT3 | ENSG00000168610 | 17 | 40455342 | 40575586 | 230 | 7.606 | 1.41e-14 | 2.72e-10 |
| Putamen | TST | ENSG00000128311 | 22 | 37396900 | 37450681 | 164 | 7.599 | 1.49e-14 | 2.87e-10 |
| Putamen | LRRC37A | ENSG00000176681 | 17 | 44335099 | 44425160 | 321 | 7.599 | 1.49e-14 | 2.88e-10 |
| Putamen | SRPRB | ENSG00000144867 | 3 | 133467877 | 133554616 | 218 | 7.586 | 1.64e-14 | 3.17e-10 |
| Putamen | NSF | ENSG00000073969 | 17 | 44633035 | 44844830 | 103 | 7.585 | 1.66e-14 | 3.2e-10 |
| Putamen | LRRC37A2 | ENSG00000238083 | 17 | 44553877 | 44643016 | 39 | 7.577 | 1.77e-14 | 3.41e-10 |
| Putamen | TMPRSS6 | ENSG00000187045 | 22 | 37451476 | 37540603 | 267 | 7.543 | 2.3e-14 | 4.45e-10 |
| Putamen | TPCN1 | ENSG00000186815 | 12 | 113623855 | 113746390 | 208 | 7.536 | 2.41e-14 | 4.66e-10 |
| Putamen | MAFK | ENSG00000198517 | 7 | 1535350 | 1592679 | 213 | 7.514 | 2.87e-14 | 5.54e-10 |
| Putamen | HFE | ENSG00000010704 | 6 | 26052509 | 26108571 | 134 | 7.484 | 3.62e-14 | 6.99e-10 |
| Putamen | C2orf76 | ENSG00000186132 | 2 | 120049801 | 120159404 | 351 | 7.48 | 3.71e-14 | 7.17e-10 |
| Putamen | VMP1 | ENSG00000062716 | 17 | 57749553 | 57929616 | 244 | 7.472 | 3.95e-14 | 7.63e-10 |
| Putamen | SLC17A3 | ENSG00000124564 | 6 | 25823294 | 25917514 | 233 | 7.459 | 4.35e-14 | 8.39e-10 |
| Putamen | SLC17A2 | ENSG00000112337 | 6 | 25902982 | 25965946 | 182 | 7.454 | 4.51e-14 | 8.71e-10 |

| Trait | Gene symbol | Ensembl gene ID | Chromosome | Start (bp) | Stop (bp) | SNPs (n) | Z | P | Bonferroni-adjusted P |
|---|---|---|---|---|---|---|---|---|---|
| Putamen | RPS6KB1 | ENSG00000108443 | 17 | 57935447 | 58037925 | 174 | 7.442 | 4.97e-14 | 9.59e-10 |
| Putamen | MLX | ENSG00000108788 | 17 | 40684086 | 40735257 | 95 | 7.438 | 5.1e-14 | 9.84e-10 |
| Putamen | PLEKHM1 | ENSG00000225190 | 17 | 43503266 | 43603115 | 191 | 7.425 | 5.64e-14 | 1.09e-09 |
| Putamen | MRC1L1 | ENSG00000183748 | 10 | 17816362 | 17963178 | 46 | 7.411 | 6.27e-14 | 1.21e-09 |
| Putamen | BAZ1B | ENSG00000009954 | 7 | 72844728 | 72971608 | 174 | 7.405 | 6.56e-14 | 1.27e-09 |
| Putamen | SLC17A4 | ENSG00000146039 | 6 | 25719927 | 25791419 | 244 | 7.399 | 6.88e-14 | 1.33e-09 |
| Putamen | DBI | ENSG00000155368 | 2 | 120089497 | 120140126 | 163 | 7.371 | 8.48e-14 | 1.64e-09 |
| Putamen | TUBG1 | ENSG00000131462 | 17 | 40726694 | 40777252 | 78 | 7.365 | 8.86e-14 | 1.71e-09 |
| Putamen | CRHR1 | ENSG00000120088 | 17 | 43664267 | 43923194 | 1093 | 7.357 | 9.41e-14 | 1.82e-09 |
| Putamen | KCTD17 | ENSG00000100379 | 22 | 37412779 | 37469430 | 102 | 7.316 | 1.27e-13 | 2.46e-09 |
| Putamen | STAB1 | ENSG00000010327 | 3 | 52494354 | 52568511 | 120 | 7.292 | 1.52e-13 | 2.94e-09 |
| Putamen | FAM134C | ENSG00000141699 | 17 | 40721531 | 40797641 | 129 | 7.29 | 1.55e-13 | 2.99e-09 |
| Putamen | CCDC42B | ENSG00000186710 | 12 | 113552663 | 113607081 | 120 | 7.229 | 2.44e-13 | 4.71e-09 |
| Putamen | HIST1H1E | ENSG00000168298 | 6 | 26121559 | 26167343 | 86 | 7.2 | 3e-13 | 5.8e-09 |
| Putamen | FNIP2 | ENSG00000052795 | 4 | 159655290 | 159839201 | 245 | 7.193 | 3.18e-13 | 6.14e-09 |
| Putamen | WNT3 | ENSG00000108379 | 17 | 44829872 | 44945520 | 215 | 7.162 | 3.97e-13 | 7.66e-09 |
| Putamen | RNFT1 | ENSG00000189050 | 17 | 58019601 | 58077122 | 62 | 7.158 | 4.08e-13 | 7.88e-09 |
| Putamen | TNNC1 | ENSG00000114854 | 3 | 52475118 | 52523086 | 78 | 7.144 | 4.53e-13 | 8.75e-09 |
| Putamen | METTL7A | ENSG00000185432 | 12 | 51282255 | 51336300 | 120 | 7.12 | 5.42e-13 | 1.05e-08 |
| Putamen | NISCH | ENSG00000010322 | 3 | 52454134 | 52537087 | 115 | 7.08 | 7.22e-13 | 1.39e-08 |
| Putamen | HIST1H2BC | ENSG00000180596 | 6 | 26105101 | 26159154 | 129 | 7.051 | 8.87e-13 | 1.71e-08 |
| Putamen | PSME3 | ENSG00000131467 | 17 | 40941402 | 41005774 | 33 | 6.999 | 1.29e-12 | 2.49e-08 |
| Putamen | TMEM184A | ENSG00000164855 | 7 | 1571871 | 1635457 | 206 | 6.979 | 1.49e-12 | 2.88e-08 |
| Putamen | HIST1H2BA | ENSG00000146047 | 6 | 25692137 | 25737573 | 144 | 6.978 | 1.5e-12 | 2.89e-08 |
| Putamen | TMEM236 | ENSG00000184040 | 10 | 18006218 | 18099855 | 10 | 6.969 | 1.6e-12 | 3.08e-08 |
| Putamen | HIST1H2BI | ENSG00000168242 | 6 | 26238144 | 26283622 | 100 | 6.934 | 2.05e-12 | 3.95e-08 |
| Putamen | AC051642.1 | ENSG00000268085 | 8 | 23402287 | 23455651 | 319 | 6.934 | 2.05e-12 | 3.95e-08 |
| Putamen | WBSCR22 | ENSG00000071462 | 7 | 73062355 | 73129491 | 180 | 6.929 | 2.11e-12 | 4.08e-08 |
| Putamen | ARL17B | ENSG00000228696 | 17 | 44342150 | 44474130 | 274 | 6.926 | 2.17e-12 | 4.18e-08 |
| Putamen | SPPL2C | ENSG00000185294 | 17 | 43887256 | 43934438 | 268 | 6.92 | 2.25e-12 | 4.35e-08 |
| Putamen | TUBD1 | ENSG00000108423 | 17 | 57926851 | 58005304 | 142 | 6.92 | 2.25e-12 | 4.35e-08 |
| Putamen | ARL17A | ENSG00000185829 | 17 | 44584068 | 44692088 | 32 | 6.917 | 2.31e-12 | 4.47e-08 |
| Putamen | BECN1 | ENSG00000126581 | 17 | 40952152 | 41020367 | 33 | 6.897 | 2.66e-12 | 5.14e-08 |
| Putamen | AOC2 | ENSG00000131480 | 17 | 40961617 | 41012724 | 23 | 6.896 | 2.68e-12 | 5.18e-08 |
| Putamen | HIST1H3F | ENSG00000256316 | 6 | 26240370 | 26285835 | 106 | 6.892 | 2.74e-12 | 5.29e-08 |
| Putamen | HYKK | ENSG00000188266 | 15 | 78764906 | 78839714 | 161 | 6.886 | 2.88e-12 | 5.55e-08 |
| Putamen | PPID | ENSG00000171497 | 4 | 159620286 | 159679548 | 76 | 6.877 | 3.05e-12 | 5.88e-08 |
| Putamen | HIST1H4G | ENSG00000124578 | 6 | 26236886 | 26282259 | 102 | 6.856 | 3.55e-12 | 6.86e-08 |
| Putamen | TF | ENSG00000091513 | 3 | 133429800 | 133507850 | 256 | 6.792 | 5.54e-12 | 1.07e-07 |
| Putamen | TMEM37 | ENSG00000171227 | 2 | 120152477 | 120206096 | 139 | 6.75 | 7.38e-12 | 1.43e-07 |
| Putamen | CHRNA5 | ENSG00000169684 | 15 | 78822862 | 78897611 | 161 | 6.7 | 1.05e-11 | 2.02e-07 |
| Putamen | BAP1 | ENSG00000163930 | 3 | 52425029 | 52479366 | 50 | 6.695 | 1.08e-11 | 2.09e-07 |
| Putamen | PSMA4 | ENSG00000041357 | 15 | 78797747 | 78851604 | 127 | 6.689 | 1.13e-11 | 2.18e-07 |
| Putamen | AC027228.1 | ENSG00000268838 | 15 | 78795023 | 78841288 | 110 | 6.681 | 1.19e-11 | 2.29e-07 |
| Putamen | PHF7 | ENSG00000010318 | 3 | 52409673 | 52467657 | 48 | 6.586 | 2.25e-11 | 4.35e-07 |
| Putamen | HIST1H4H | ENSG00000158406 | 6 | 26271283 | 26320762 | 164 | 6.565 | 2.6e-11 | 5.02e-07 |
| Putamen | HIST1H3G | ENSG00000256018 | 6 | 26261146 | 26306612 | 133 | 6.545 | 2.98e-11 | 5.76e-07 |
| Putamen | AOC3 | ENSG00000131471 | 17 | 40968201 | 41020147 | 24 | 6.543 | 3.01e-11 | 5.81e-07 |
| Putamen | CHRNA3 | ENSG00000080644 | 15 | 78875394 | 78948637 | 155 | 6.509 | 3.79e-11 | 7.31e-07 |
| Putamen | ENTPD4 | ENSG00000197217 | 8 | 23233296 | 23350208 | 333 | 6.488 | 4.34e-11 | 8.39e-07 |
| Putamen | AC021860.1 | ENSG00000196355 | 4 | 38618029 | 38701430 | 179 | 6.471 | 4.85e-11 | 9.37e-07 |
| Putamen | TEX33 | ENSG00000185264 | 22 | 37377163 | 37438882 | 232 | 6.446 | 5.76e-11 | 1.11e-06 |

| Trait | Gene symbol | Ensembl gene ID | Chromosome | Start (bp) | Stop (bp) | SNPs (n) | Z | P | Bonferroni-adjusted P |
|---|---|---|---|---|---|---|---|---|---|
| Putamen | CNTD1 | ENSG00000176563 | 17 | 40915810 | 40973605 | 29 | 6.437 | 6.09e-11 | 1.18e-06 |
| Putamen | MAF | ENSG00000178573 | 16 | 79609740 | 79669611 | 168 | 6.362 | 9.92e-11 | 1.92e-06 |
| Putamen | PLLP | ENSG00000102934 | 16 | 57280004 | 57353599 | 245 | 6.34 | 1.15e-10 | 2.22e-06 |
| Putamen | KLF3 | ENSG00000109787 | 4 | 38630817 | 38712663 | 176 | 6.316 | 1.34e-10 | 2.58e-06 |
| Putamen | DNAJC30 | ENSG00000176410 | 7 | 73085299 | 73132783 | 115 | 6.308 | 1.41e-10 | 2.73e-06 |
| Putamen | SSPN | ENSG00000123096 | 12 | 26239924 | 26462223 | 622 | 6.301 | 1.48e-10 | 2.86e-06 |
| Putamen | MPST | ENSG00000128309 | 22 | 37380676 | 37435863 | 202 | 6.256 | 1.98e-10 | 3.81e-06 |
| Putamen | COA3 | ENSG00000183978 | 17 | 40937165 | 40985722 | 23 | 6.233 | 2.28e-10 | 4.41e-06 |
| Putamen | STMN4 | ENSG00000015592 | 8 | 27082840 | 27150937 | 161 | 6.168 | 3.47e-10 | 6.7e-06 |
| Putamen | CPNE2 | ENSG00000140848 | 16 | 57091449 | 57191878 | 319 | 6.165 | 3.53e-10 | 6.82e-06 |
| Putamen | HCRT | ENSG00000161610 | 17 | 40326078 | 40372470 | 30 | 6.117 | 4.75e-10 | 9.18e-06 |
| Putamen | HIST1H2AA | ENSG00000164508 | 6 | 25716291 | 25761790 | 164 | 6.109 | 5e-10 | 9.65e-06 |
| Putamen | TRIM38 | ENSG00000112343 | 6 | 25928030 | 25997384 | 160 | 6.109 | 5e-10 | 9.65e-06 |
| Putamen | HIST1H3A | ENSG00000198366 | 6 | 25985718 | 26031186 | 80 | 6.109 | 5e-10 | 9.65e-06 |
| Putamen | HIST1H4A | ENSG00000196176 | 6 | 25986907 | 26032278 | 79 | 6.109 | 5e-10 | 9.65e-06 |
| Putamen | HIST1H1A | ENSG00000124610 | 6 | 26007260 | 26053040 | 93 | 6.109 | 5e-10 | 9.65e-06 |
| Putamen | HIST1H3C | ENSG00000196532 | 6 | 26010639 | 26056097 | 90 | 6.109 | 5e-10 | 9.65e-06 |
| Putamen | HIST1H4B | ENSG00000124529 | 6 | 26017124 | 26062480 | 89 | 6.109 | 5e-10 | 9.65e-06 |
| Putamen | HIST1H3B | ENSG00000124693 | 6 | 26021817 | 26067288 | 86 | 6.109 | 5e-10 | 9.65e-06 |
| Putamen | HIST1H2AB | ENSG00000137259 | 6 | 26023320 | 26068796 | 82 | 6.109 | 5e-10 | 9.65e-06 |
| Putamen | HIST1H2BB | ENSG00000196226 | 6 | 26033455 | 26078885 | 94 | 6.109 | 5e-10 | 9.65e-06 |
| Putamen | HIST1H1C | ENSG00000187837 | 6 | 26045968 | 26091699 | 101 | 6.109 | 5e-10 | 9.65e-06 |
| Putamen | HIST1H4C | ENSG00000197061 | 6 | 26069104 | 26114518 | 128 | 6.109 | 5e-10 | 9.65e-06 |
| Putamen | HIST1H1T | ENSG00000187475 | 6 | 26097640 | 26143364 | 127 | 6.109 | 5e-10 | 9.65e-06 |
| Putamen | HIST1H2BD | ENSG00000158373 | 6 | 26123349 | 26181577 | 98 | 6.109 | 5e-10 | 9.65e-06 |
| Putamen | FZD9 | ENSG00000188763 | 7 | 72813109 | 72860450 | 69 | 6.109 | 5e-10 | 9.65e-06 |
| Putamen | BCL7B | ENSG00000106635 | 7 | 72940686 | 73007332 | 124 | 6.109 | 5e-10 | 9.65e-06 |
| Putamen | MLXIPL | ENSG00000009950 | 7 | 72997524 | 73073873 | 203 | 6.109 | 5e-10 | 9.65e-06 |
| Putamen | HIGD1C | ENSG00000214511 | 12 | 51312705 | 51374289 | 197 | 6.109 | 5e-10 | 9.65e-06 |
| Putamen | SLC11A2 | ENSG00000110911 | 12 | 51363184 | 51457349 | 233 | 6.109 | 5e-10 | 9.65e-06 |
| Putamen | LETMD1 | ENSG00000050426 | 12 | 51406745 | 51464207 | 134 | 6.109 | 5e-10 | 9.65e-06 |
| Putamen | CSRNP2 | ENSG00000110925 | 12 | 51444990 | 51512447 | 166 | 6.109 | 5e-10 | 9.65e-06 |
| Putamen | DDX54 | ENSG00000123064 | 12 | 113584979 | 113658284 | 117 | 6.109 | 5e-10 | 9.65e-06 |
| Putamen | RITA1 | ENSG00000139405 | 12 | 113588331 | 113640173 | 87 | 6.109 | 5e-10 | 9.65e-06 |
| Putamen | IQCD | ENSG00000166578 | 12 | 113623246 | 113693899 | 110 | 6.109 | 5e-10 | 9.65e-06 |
| Putamen | ATP6V0A1 | ENSG00000033627 | 17 | 40575862 | 40684629 | 183 | 6.109 | 5e-10 | 9.65e-06 |
| Putamen | NAGLU | ENSG00000108784 | 17 | 40653190 | 40706467 | 84 | 6.109 | 5e-10 | 9.65e-06 |
| Putamen | HSD17B1 | ENSG00000108786 | 17 | 40666232 | 40717231 | 91 | 6.109 | 5e-10 | 9.65e-06 |
| Putamen | COASY | ENSG00000068120 | 17 | 40678485 | 40728295 | 102 | 6.109 | 5e-10 | 9.65e-06 |
| Putamen | PSMC3IP | ENSG00000131470 | 17 | 40714333 | 40764849 | 75 | 6.109 | 5e-10 | 9.65e-06 |
| Putamen | TUBG2 | ENSG00000037042 | 17 | 40776323 | 40829024 | 91 | 6.109 | 5e-10 | 9.65e-06 |
| Putamen | CNTNAP1 | ENSG00000108797 | 17 | 40799631 | 40861832 | 73 | 6.109 | 5e-10 | 9.65e-06 |
| Putamen | PLEKHH3 | ENSG00000068137 | 17 | 40809932 | 40864048 | 54 | 6.109 | 5e-10 | 9.65e-06 |
| Putamen | CCR10 | ENSG00000184451 | 17 | 40820907 | 40870935 | 45 | 6.109 | 5e-10 | 9.65e-06 |
| Putamen | ARHGAP27 | ENSG00000159314 | 17 | 43461275 | 43546787 | 183 | 6.109 | 5e-10 | 9.65e-06 |
| Putamen | STH | ENSG00000256762 | 17 | 44041616 | 44087060 | 218 | 6.109 | 5e-10 | 9.65e-06 |
| Putamen | CLTC | ENSG00000141367 | 17 | 57662219 | 57783671 | 166 | 6.109 | 5e-10 | 9.65e-06 |
| Putamen | PTRH2 | ENSG00000141378 | 17 | 57741997 | 57819987 | 100 | 6.109 | 5e-10 | 9.65e-06 |
| Putamen | RP11-178C3.1 | ENSG00000267318 | 17 | 57983269 | 58060462 | 109 | 6.109 | 5e-10 | 9.65e-06 |
| Putamen | TMPRSS12 | ENSG00000186452 | 12 | 51201703 | 51291667 | 192 | 6.098 | 5.37e-10 | 1.04e-05 |
| Putamen | PTPMT1 | ENSG00000110536 | 11 | 47551982 | 47605013 | 73 | 6.075 | 6.2e-10 | 1.2e-05 |
| Putamen | HLA-B | ENSG00000234745 | 6 | 31311649 | 31359965 | 1543 | 6.073 | 6.27e-10 | 1.21e-05 |

| Trait | Gene symbol | Ensembl gene ID | Chromosome | Start (bp) | Stop (bp) | SNPs (n) | Z | P | Bonferroni-adjusted P |
|---|---|---|---|---|---|---|---|---|---|
| Putamen | SLC8B1 | ENSG00000089060 | 12 | 113726564 | 113832298 | 215 | 6.069 | 6.44e-10 | 1.24e-05 |
| Putamen | WNK4 | ENSG00000126562 | 17 | 40897696 | 40958954 | 35 | 6.055 | 7.01e-10 | 1.35e-05 |
| Putamen | HIST1H1D | ENSG00000124575 | 6 | 26224440 | 26270216 | 104 | 6.017 | 8.91e-10 | 1.72e-05 |
| Putamen | GHDC | ENSG00000167925 | 17 | 40330817 | 40381531 | 35 | 5.985 | 1.08e-09 | 2.09e-05 |
| Putamen | SCTR | ENSG00000080293 | 2 | 120187419 | 120317070 | 358 | 5.982 | 1.1e-09 | 2.12e-05 |
| Putamen | NDUFS3 | ENSG00000213619 | 11 | 47551888 | 47616114 | 84 | 5.981 | 1.11e-09 | 2.14e-05 |
| Putamen | INTS1 | ENSG00000164880 | 7 | 1499913 | 1580489 | 258 | 5.976 | 1.14e-09 | 2.21e-05 |
| Putamen | IREB2 | ENSG00000136381 | 15 | 78694773 | 78803798 | 208 | 5.895 | 1.88e-09 | 3.62e-05 |
| Putamen | RASAL1 | ENSG00000111344 | 12 | 113526624 | 113609044 | 229 | 5.889 | 1.95e-09 | 3.76e-05 |
| Putamen | KBTBD4 | ENSG00000231880 | 11 | 47564277 | 47609823 | 60 | 5.841 | 2.59e-09 | 5.01e-05 |
| Putamen | C4orf45 | ENSG00000164123 | 4 | 159804286 | 159994912 | 466 | 5.82 | 2.94e-09 | 5.68e-05 |
| Putamen | CTSH | ENSG00000103811 | 15 | 79203400 | 79276916 | 218 | 5.817 | 2.99e-09 | 5.78e-05 |
| Putamen | VPS25 | ENSG00000131475 | 17 | 40890454 | 40941617 | 32 | 5.67 | 7.13e-09 | 0.000138 |
| Putamen | DVL2 | ENSG00000004975 | 17 | 7118660 | 7172864 | 60 | 5.657 | 7.69e-09 | 0.000148 |
| Putamen | ELP5 | ENSG00000170291 | 17 | 7119735 | 7173259 | 60 | 5.657 | 7.69e-09 | 0.000148 |
| Putamen | PSORS1C1 | ENSG00000204540 | 6 | 31047527 | 31117869 | 731 | 5.648 | 8.14e-09 | 0.000157 |
| Putamen | MPPED2 | ENSG00000066382 | 11 | 30396040 | 30643419 | 528 | 5.633 | 8.85e-09 | 0.000171 |
| Putamen | PHF23 | ENSG00000040633 | 17 | 7128347 | 7178041 | 59 | 5.632 | 8.91e-09 | 0.000172 |
| Putamen | EZH1 | ENSG00000108799 | 17 | 40842293 | 40932071 | 66 | 5.63 | 9.02e-09 | 0.000174 |
| Putamen | NUP160 | ENSG00000030066 | 11 | 47789639 | 47905107 | 188 | 5.622 | 9.47e-09 | 0.000183 |
| Putamen | FNBP4 | ENSG00000109920 | 11 | 47728072 | 47823995 | 161 | 5.618 | 9.68e-09 | 0.000187 |
| Putamen | MORF4L1 | ENSG00000185787 | 15 | 79067829 | 79200475 | 437 | 5.587 | 1.15e-08 | 0.000223 |
| Putamen | PICALM | ENSG00000073921 | 11 | 85658727 | 85815924 | 381 | 5.586 | 1.16e-08 | 0.000224 |
| Putamen | BTN3A2 | ENSG00000186470 | 6 | 26330387 | 26388546 | 250 | 5.585 | 1.17e-08 | 0.000226 |
| Putamen | CELF1 | ENSG00000149187 | 11 | 47477496 | 47622121 | 153 | 5.569 | 1.28e-08 | 0.000248 |
| Putamen | GABARAP | ENSG00000170296 | 17 | 7133333 | 7181089 | 57 | 5.562 | 1.33e-08 | 0.000257 |
| Putamen | CTD-2545G14.7 | ENSG00000262526 | 17 | 7133746 | 7182954 | 60 | 5.548 | 1.45e-08 | 0.000279 |
| Putamen | RAB6B | ENSG00000154917 | 3 | 133533083 | 133649680 | 298 | 5.541 | 1.5e-08 | 0.00029 |
| Putamen | AGBL2 | ENSG00000165923 | 11 | 47671143 | 47771941 | 155 | 5.54 | 1.51e-08 | 0.000292 |
| Putamen | TMEM206 | ENSG00000065600 | 1 | 212527273 | 212623243 | 229 | 5.53 | 1.6e-08 | 0.000309 |
| Putamen | ALAS1 | ENSG00000023330 | 3 | 52197102 | 52258343 | 77 | 5.529 | 1.61e-08 | 0.000311 |
| Putamen | RAMP2 | ENSG00000131477 | 17 | 40875465 | 40925059 | 41 | 5.518 | 1.71e-08 | 0.000331 |
| Putamen | CTDNEP1 | ENSG00000175826 | 17 | 7136910 | 7190810 | 71 | 5.51 | 1.79e-08 | 0.000346 |
| Putamen | NENF | ENSG00000117691 | 1 | 212571229 | 212629714 | 155 | 5.497 | 1.93e-08 | 0.000373 |
| Putamen | MTCH2 | ENSG00000109919 | 11 | 47628867 | 47699175 | 74 | 5.49 | 2.01e-08 | 0.000388 |
| Putamen | STX1A | ENSG00000106089 | 7 | 73103536 | 73169002 | 129 | 5.468 | 2.27e-08 | 0.000439 |
| Putamen | HLA-C | ENSG00000204525 | 6 | 31226526 | 31274907 | 1493 | 5.466 | 2.3e-08 | 0.000445 |
| Putamen | CDV3 | ENSG00000091527 | 3 | 133257574 | 133319105 | 158 | 5.45 | 2.52e-08 | 0.000486 |
| Putamen | RBBP5 | ENSG00000117222 | 1 | 205045270 | 205126143 | 203 | 5.445 | 2.59e-08 | 0.0005 |
| Putamen | DLG4 | ENSG00000132535 | 17 | 7083209 | 7158021 | 101 | 5.417 | 3.04e-08 | 0.000586 |
| Putamen | MARK2 | ENSG00000072518 | 11 | 63571400 | 63688491 | 167 | 5.416 | 3.05e-08 | 0.000588 |
| Putamen | FAM180B | ENSG00000196666 | 11 | 47573198 | 47620746 | 45 | 5.409 | 3.17e-08 | 0.000612 |
| Putamen | PRUNE | ENSG00000143363 | 1 | 150945896 | 151018189 | 159 | 5.389 | 3.54e-08 | 0.000683 |
| Putamen | ZNF280D | ENSG00000137871 | 15 | 56912379 | 57245769 | 575 | 5.374 | 3.85e-08 | 0.000743 |
| Putamen | SLC2A4 | ENSG00000181856 | 17 | 7149986 | 7201576 | 81 | 5.367 | 4e-08 | 0.000773 |
| Putamen | CLDN7 | ENSG00000181885 | 17 | 7153222 | 7202302 | 82 | 5.363 | 4.09e-08 | 0.00079 |
| Putamen | RP1-4G17.5 | ENSG00000262302 | 17 | 7140148 | 7200408 | 83 | 5.359 | 4.18e-08 | 0.000806 |
| Putamen | CHRNB4 | ENSG00000117971 | 15 | 78906461 | 79055096 | 481 | 5.346 | 4.5e-08 | 0.000868 |
| Putamen | BAIAP2L1 | ENSG00000006453 | 7 | 97910963 | 98065380 | 334 | 5.316 | 5.29e-08 | 0.00102 |
| Putamen | DNAH1 | ENSG00000114841 | 3 | 52315335 | 52444507 | 154 | 5.299 | 5.84e-08 | 0.00113 |
| Putamen | ACADVL | ENSG00000072778 | 17 | 7085444 | 7138592 | 81 | 5.268 | 6.9e-08 | 0.00133 |
| Putamen | TFRC | ENSG00000072274 | 3 | 195744054 | 195844060 | 428 | 5.236 | 8.19e-08 | 0.00158 |

| Trait | Gene symbol | Ensembl gene ID | Chromosome | Start (bp) | Stop (bp) | SNPs (n) | Z | P | Bonferroni-adjusted P |
|---|---|---|---|---|---|---|---|---|---|
| Putamen | TMEM81 | ENSG00000174529 | 1 | 205042258 | 205088645 | 124 | 5.208 | 9.53e-08 | 0.00184 |
| Putamen | CDSN | ENSG00000204539 | 6 | 31072867 | 31123223 | 522 | 5.166 | 1.2e-07 | 0.00231 |
| Putamen | LIMK1 | ENSG00000106683 | 7 | 73462263 | 73546855 | 136 | 5.162 | 1.22e-07 | 0.00236 |
| Putamen | DUOX2 | ENSG00000140279 | 15 | 45374848 | 45441542 | 124 | 5.152 | 1.29e-07 | 0.00249 |
| Putamen | TBC1D4 | ENSG00000136111 | 13 | 75848808 | 76091250 | 563 | 5.118 | 1.54e-07 | 0.00297 |
| Putamen | BHLHE41 | ENSG00000123095 | 12 | 26262959 | 26313060 | 114 | 5.079 | 1.9e-07 | 0.00367 |
| Putamen | POM121 | ENSG00000196313 | 7 | 72314936 | 72431979 | 102 | 5.065 | 2.04e-07 | 0.00394 |
| Putamen | DUOX1 | ENSG00000137857 | 15 | 45387131 | 45467774 | 158 | 5.061 | 2.08e-07 | 0.00402 |
| Putamen | AC005488.1 | ENSG00000267985 | 7 | 72424478 | 72474996 | 13 | 5.046 | 2.26e-07 | 0.00435 |
| Putamen | TRIM74 | ENSG00000155428 | 7 | 72420016 | 72474997 | 15 | 5.046 | 2.26e-07 | 0.00436 |
| Putamen | ATXN7L3 | ENSG00000087152 | 17 | 42259173 | 42312481 | 82 | 5.018 | 2.61e-07 | 0.00503 |
| Putamen | C6orf15 | ENSG00000204542 | 6 | 31069000 | 31115336 | 511 | 4.991 | 3.01e-07 | 0.00581 |
| Putamen | FAM63A | ENSG00000143409 | 1 | 150959025 | 151015851 | 127 | 4.982 | 3.14e-07 | 0.00607 |
| Putamen | POU6F1 | ENSG00000184271 | 12 | 51570719 | 51646477 | 157 | 4.978 | 3.21e-07 | 0.00621 |
| Putamen | MCCD1 | ENSG00000204511 | 6 | 31461494 | 31508009 | 380 | 4.941 | 3.88e-07 | 0.00749 |
| Putamen | ADAMTS7 | ENSG00000136378 | 15 | 79041545 | 79138773 | 388 | 4.935 | 4.01e-07 | 0.00775 |
| Putamen | DUOXA2 | ENSG00000140274 | 15 | 45371519 | 45420619 | 100 | 4.926 | 4.19e-07 | 0.00809 |
| Putamen | G6PC | ENSG00000131482 | 17 | 41017814 | 41075386 | 78 | 4.921 | 4.3e-07 | 0.0083 |
| Putamen | COL5A2 | ENSG00000204262 | 2 | 189886622 | 190079605 | 456 | 4.911 | 4.53e-07 | 0.00875 |
| Putamen | LOXL2 | ENSG00000134013 | 8 | 23144702 | 23317841 | 731 | 4.902 | 4.74e-07 | 0.00915 |
| Putamen | SLC39A13 | ENSG00000165915 | 11 | 47393683 | 47448047 | 115 | 4.896 | 4.9e-07 | 0.00946 |
| Putamen | KBTBD4 | ENSG00000123444 | 11 | 47583749 | 47635567 | 53 | 4.887 | 5.12e-07 | 0.00988 |
| Putamen | TLR9 | ENSG00000173366 | 3 | 52245097 | 52300206 | 62 | 4.878 | 5.37e-07 | 0.0104 |
| Putamen | C17orf53 | ENSG00000125319 | 17 | 42184274 | 42249844 | 103 | 4.821 | 7.15e-07 | 0.0138 |
| Putamen | HSPB9 | ENSG00000197723 | 17 | 40239756 | 40285371 | 76 | 4.818 | 7.26e-07 | 0.014 |
| Putamen | MYBPC3 | ENSG00000134571 | 11 | 47342957 | 47409253 | 149 | 4.803 | 7.81e-07 | 0.0151 |
| Putamen | DHX58 | ENSG00000108771 | 17 | 40243422 | 40299751 | 104 | 4.8 | 7.93e-07 | 0.0153 |
| Putamen | CTD-2132N18.3 | ENSG00000267261 | 17 | 40261692 | 40341934 | 119 | 4.785 | 8.57e-07 | 0.0166 |
| Putamen | BNIPL | ENSG00000163141 | 1 | 150974046 | 151030076 | 111 | 4.759 | 9.72e-07 | 0.0188 |
| Putamen | GPX6 | ENSG00000198704 | 6 | 28461073 | 28530992 | 149 | 4.755 | 9.94e-07 | 0.0192 |
| Putamen | C5orf63 | ENSG00000164241 | 5 | 126368250 | 126444184 | 129 | 4.753 | 1e-06 | 0.0194 |
| Putamen | NPHP3 | ENSG00000113971 | 3 | 132266986 | 132476303 | 333 | 4.752 | 1.01e-06 | 0.0194 |
| Putamen | RAB5C | ENSG00000108774 | 17 | 40266994 | 40342035 | 111 | 4.75 | 1.02e-06 | 0.0196 |
| Putamen | CDKN2A | ENSG00000147889 | 9 | 21957751 | 22030300 | 104 | 4.75 | 1.02e-06 | 0.0197 |
| Putamen | CNTN2 | ENSG00000184144 | 1 | 204977325 | 205057627 | 236 | 4.74 | 1.07e-06 | 0.0206 |
| Putamen | CDKN2B | ENSG00000147883 | 9 | 21992902 | 22044362 | 92 | 4.737 | 1.09e-06 | 0.021 |
| Putamen | GPX5 | ENSG00000224586 | 6 | 28458702 | 28512729 | 111 | 4.729 | 1.13e-06 | 0.0218 |
| Putamen | RPL31 | ENSG00000071082 | 2 | 101583177 | 101650494 | 154 | 4.727 | 1.14e-06 | 0.022 |
| Putamen | HDAC5 | ENSG00000108840 | 17 | 42144114 | 42236070 | 137 | 4.714 | 1.22e-06 | 0.0235 |
| Putamen | TMUB2 | ENSG00000168591 | 17 | 42229338 | 42279099 | 76 | 4.71 | 1.24e-06 | 0.024 |
| Putamen | MICA | ENSG00000204520 | 6 | 31336356 | 31393092 | 1063 | 4.707 | 1.26e-06 | 0.0243 |
| Putamen | HDAC9 | ENSG00000048052 | 7 | 18091572 | 19052039 | 2125 | 4.701 | 1.29e-06 | 0.025 |
| Putamen | ASB16 | ENSG00000161664 | 17 | 42212815 | 42266451 | 78 | 4.685 | 1.4e-06 | 0.027 |
| Putamen | KAT2A | ENSG00000108773 | 17 | 40255126 | 40308376 | 100 | 4.68 | 1.43e-06 | 0.0276 |
| Putamen | SPI1 | ENSG00000066336 | 11 | 47366411 | 47435127 | 156 | 4.674 | 1.47e-06 | 0.0284 |
| Putamen | BTN2A2 | ENSG00000124508 | 6 | 26348324 | 26405102 | 258 | 4.671 | 1.5e-06 | 0.029 |
| Putamen | BRI3 | ENSG00000164713 | 7 | 97846691 | 97947162 | 240 | 4.668 | 1.52e-06 | 0.0294 |
| Putamen | TLR9 | ENSG00000239732 | 3 | 52245096 | 52308183 | 70 | 4.649 | 1.67e-06 | 0.0322 |
| Putamen | C1orf56 | ENSG00000143443 | 1 | 150985216 | 151034462 | 97 | 4.633 | 1.8e-06 | 0.0347 |
| Putamen | C1QTNF4 | ENSG00000172247 | 11 | 47601216 | 47651211 | 47 | 4.63 | 1.83e-06 | 0.0353 |
| Putamen | HIST1H2BH | ENSG00000197459 | 6 | 26216879 | 26262303 | 108 | 4.616 | 1.95e-06 | 0.0377 |
| Putamen | PPM1M | ENSG00000164088 | 3 | 52244841 | 52294613 | 54 | 4.585 | 2.27e-06 | 0.0439 |

| Trait | Gene symbol | Ensembl gene ID | Chromosome | Start (bp) | Stop (bp) | SNPs (n) | Z | P | Bonferroni-adjusted P |
|---|---|---|---|---|---|---|---|---|---|
| Putamen | LLGL1 | ENSG00000131899 | 17 | 18093901 | 18158189 | 135 | 4.577 | 2.36e-06 | 0.0456 |
| Putamen | TWF2 | ENSG00000247596 | 3 | 52252626 | 52308276 | 68 | 4.561 | 2.54e-06 | 0.0491 |
| Red nucleus | LRRC16A | ENSG00000079691 | 6 | 25244306 | 25630758 | 1409 | 9.2 | 1.79e-20 | 3.46e-16 |
| Red nucleus | SLC39A14 | ENSG00000104635 | 8 | 22189762 | 22301642 | 339 | 8.676 | 2.05e-18 | 3.96e-14 |
| Red nucleus | SCGN | ENSG00000079689 | 6 | 25617464 | 25712011 | 282 | 8.621 | 3.3e-18 | 6.38e-14 |
| Red nucleus | TF | ENSG00000091513 | 3 | 133429800 | 133507850 | 256 | 8.129 | 2.17e-16 | 4.19e-12 |
| Red nucleus | COL5A2 | ENSG00000204262 | 2 | 189886622 | 190079605 | 456 | 8.043 | 4.4e-16 | 8.49e-12 |
| Red nucleus | SLC39A12 | ENSG00000148482 | 10 | 18205768 | 18342221 | 469 | 7.832 | 2.4e-15 | 4.64e-11 |
| Red nucleus | PPP3CC | ENSG00000120910 | 8 | 22263332 | 22408652 | 289 | 7.819 | 2.66e-15 | 5.13e-11 |
| Red nucleus | HIST1H3A | ENSG00000198366 | 6 | 25985718 | 26031186 | 80 | 7.795 | 3.22e-15 | 6.22e-11 |
| Red nucleus | SORBS3 | ENSG00000120896 | 8 | 22367499 | 22443301 | 143 | 7.726 | 5.55e-15 | 1.07e-10 |
| Red nucleus | HIST1H1A | ENSG00000124610 | 6 | 26007260 | 26053040 | 93 | 7.712 | 6.22e-15 | 1.2e-10 |
| Red nucleus | SLC17A3 | ENSG00000124564 | 6 | 25823294 | 25917514 | 233 | 7.663 | 9.1e-15 | 1.76e-10 |
| Red nucleus | HIST1H3C | ENSG00000196532 | 6 | 26010639 | 26056097 | 90 | 7.657 | 9.55e-15 | 1.84e-10 |
| Red nucleus | HIST1H2AC | ENSG00000180573 | 6 | 26089373 | 26149344 | 158 | 7.518 | 2.78e-14 | 5.36e-10 |
| Red nucleus | LLGL1 | ENSG00000131899 | 17 | 18093901 | 18158189 | 135 | 7.486 | 3.54e-14 | 6.84e-10 |
| Red nucleus | HFE | ENSG00000010704 | 6 | 26052509 | 26108571 | 134 | 7.462 | 4.25e-14 | 8.21e-10 |
| Red nucleus | RNF24 | ENSG00000101236 | 20 | 3897956 | 4031229 | 323 | 7.456 | 4.47e-14 | 8.64e-10 |
| Red nucleus | HIST1H2AB | ENSG00000137259 | 6 | 26023320 | 26068796 | 82 | 7.445 | 4.85e-14 | 9.36e-10 |
| Red nucleus | ACADVL | ENSG00000072778 | 17 | 7085444 | 7138592 | 81 | 7.442 | 4.95e-14 | 9.56e-10 |
| Red nucleus | KCTD17 | ENSG00000100379 | 22 | 37412779 | 37469430 | 102 | 7.43 | 5.43e-14 | 1.05e-09 |
| Red nucleus | DLG4 | ENSG00000132535 | 17 | 7083209 | 7158021 | 101 | 7.362 | 9.03e-14 | 1.74e-09 |
| Red nucleus | WDR75 | ENSG00000115368 | 2 | 190271159 | 190350291 | 241 | 7.354 | 9.65e-14 | 1.86e-09 |
| Red nucleus | SLC17A2 | ENSG00000112337 | 6 | 25902982 | 25965946 | 182 | 7.348 | 1.01e-13 | 1.94e-09 |
| Red nucleus | ALKBH5 | ENSG00000091542 | 17 | 18051392 | 18123268 | 131 | 7.302 | 1.42e-13 | 2.74e-09 |
| Red nucleus | CSRNP2 | ENSG00000110925 | 12 | 51444990 | 51512447 | 166 | 7.298 | 1.46e-13 | 2.83e-09 |
| Red nucleus | TFCP2 | ENSG00000135457 | 12 | 51477446 | 51601926 | 308 | 7.288 | 1.57e-13 | 3.03e-09 |
| Red nucleus | HIST1H4C | ENSG00000197061 | 6 | 26069104 | 26114518 | 128 | 7.244 | 2.18e-13 | 4.21e-09 |
| Red nucleus | METTL7A | ENSG00000185432 | 12 | 51282255 | 51336300 | 120 | 7.17 | 3.75e-13 | 7.25e-09 |
| Red nucleus | DVL2 | ENSG00000004975 | 17 | 7118660 | 7172864 | 60 | 7.165 | 3.9e-13 | 7.53e-09 |
| Red nucleus | ELP5 | ENSG00000170291 | 17 | 7119735 | 7173259 | 60 | 7.165 | 3.9e-13 | 7.53e-09 |
| Red nucleus | TST | ENSG00000128311 | 22 | 37396900 | 37450681 | 164 | 7.127 | 5.13e-13 | 9.91e-09 |
| Red nucleus | HIST1H2BD | ENSG00000158373 | 6 | 26123349 | 26181577 | 98 | 7.062 | 8.23e-13 | 1.59e-08 |
| Red nucleus | TMPRSS6 | ENSG00000187045 | 22 | 37451476 | 37540603 | 267 | 7.061 | 8.26e-13 | 1.59e-08 |
| Red nucleus | HIST1H1T | ENSG00000187475 | 6 | 26097640 | 26143364 | 127 | 7.058 | 8.45e-13 | 1.63e-08 |
| Red nucleus | PHF23 | ENSG00000040633 | 17 | 7128347 | 7178041 | 59 | 7.021 | 1.1e-12 | 2.12e-08 |
| Red nucleus | HIST1H2BI | ENSG00000168242 | 6 | 26238144 | 26283622 | 100 | 6.987 | 1.4e-12 | 2.71e-08 |
| Red nucleus | NAGLU | ENSG00000108784 | 17 | 40653190 | 40706467 | 84 | 6.902 | 2.57e-12 | 4.96e-08 |
| Red nucleus | GABARAP | ENSG00000170296 | 17 | 7133333 | 7181089 | 57 | 6.868 | 3.26e-12 | 6.29e-08 |
| Red nucleus | HIST1H3F | ENSG00000256316 | 6 | 26240370 | 26285835 | 106 | 6.867 | 3.28e-12 | 6.34e-08 |
| Red nucleus | HIST1H1E | ENSG00000168298 | 6 | 26121559 | 26167343 | 86 | 6.861 | 3.41e-12 | 6.59e-08 |
| Red nucleus | ASGR1 | ENSG00000141505 | 17 | 7066750 | 7117883 | 89 | 6.85 | 3.69e-12 | 7.12e-08 |
| Red nucleus | HIST1H4G | ENSG00000124578 | 6 | 26236886 | 26282259 | 102 | 6.838 | 4.01e-12 | 7.74e-08 |
| Red nucleus | MIEF2 | ENSG00000177427 | 17 | 18128848 | 18179866 | 92 | 6.816 | 4.7e-12 | 9.07e-08 |
| Red nucleus | CTDNEP1 | ENSG00000175826 | 17 | 7136910 | 7190810 | 71 | 6.808 | 4.94e-12 | 9.55e-08 |
| Red nucleus | SLC40A1 | ENSG00000138449 | 2 | 190415305 | 190483484 | 125 | 6.8 | 5.25e-12 | 1.01e-07 |
| Red nucleus | CTD-2545G14.7 | ENSG00000262526 | 17 | 7133746 | 7182954 | 60 | 6.788 | 5.7e-12 | 1.1e-07 |
| Red nucleus | TFRC | ENSG00000072274 | 3 | 195744054 | 195844060 | 428 | 6.756 | 7.11e-12 | 1.37e-07 |
| Red nucleus | DBI | ENSG00000155368 | 2 | 120089497 | 120140126 | 163 | 6.689 | 1.13e-11 | 2.17e-07 |
| Red nucleus | HIST1H2BC | ENSG00000180596 | 6 | 26105101 | 26159154 | 129 | 6.551 | 2.85e-11 | 5.5e-07 |
| Red nucleus | RP1-4G17.5 | ENSG00000262302 | 17 | 7140148 | 7200408 | 83 | 6.53 | 3.28e-11 | 6.33e-07 |
| Red nucleus | SLC2A4 | ENSG00000181856 | 17 | 7149986 | 7201576 | 81 | 6.512 | 3.72e-11 | 7.17e-07 |

| Trait | Gene symbol | Ensembl gene ID | Chromosome | Start (bp) | Stop (bp) | SNPs (n) | Z | P | Bonferroni-adjusted P |
|---|---|---|---|---|---|---|---|---|---|
| Red nucleus | CLDN7 | ENSG00000181885 | 17 | 7153222 | 7202302 | 82 | 6.495 | 4.17e-11 | 8.04e-07 |
| Red nucleus | HIST1H4H | ENSG00000158406 | 6 | 26271283 | 26320762 | 164 | 6.476 | 4.71e-11 | 9.1e-07 |
| Red nucleus | COASY | ENSG00000068120 | 17 | 40678485 | 40728295 | 102 | 6.463 | 5.12e-11 | 9.89e-07 |
| Red nucleus | HSD17B1 | ENSG00000108786 | 17 | 40666232 | 40717231 | 91 | 6.455 | 5.4e-11 | 1.04e-06 |
| Red nucleus | OTX1 | ENSG00000115507 | 2 | 63242192 | 63294971 | 31 | 6.42 | 6.82e-11 | 1.32e-06 |
| Red nucleus | ZC3H12C | ENSG00000149289 | 11 | 109929087 | 110052566 | 326 | 6.405 | 7.52e-11 | 1.45e-06 |
| Red nucleus | HIST1H3G | ENSG00000256018 | 6 | 26261146 | 26306612 | 133 | 6.405 | 7.54e-11 | 1.46e-06 |
| Red nucleus | ATP6V0A1 | ENSG00000033627 | 17 | 40575862 | 40684629 | 183 | 6.322 | 1.29e-10 | 2.49e-06 |
| Red nucleus | SLC25A37 | ENSG00000147454 | 8 | 23351318 | 23442976 | 425 | 6.25 | 2.05e-10 | 3.95e-06 |
| Red nucleus | MLX | ENSG00000108788 | 17 | 40684086 | 40735257 | 95 | 6.189 | 3.03e-10 | 5.85e-06 |
| Red nucleus | TEX33 | ENSG00000185264 | 22 | 37377163 | 37438882 | 232 | 6.152 | 3.83e-10 | 7.4e-06 |

**Supplementary Table 4. Significant genes from MAGMA gene-based analysis of quantitative susceptibility mapping-derived brain iron across six subcortical regions (Bonferroni-corrected P < 2.59 x 10-6).** ***(continued)***

| Trait | Gene symbol | Ensembl gene ID | Chromosome | Start (bp) | Stop (bp) | SNPs (n) | Z | P | Bonferroni-adjusted P |
|---|---|---|---|---|---|---|---|---|---|
| Red nucleus | HIST1H2BA | ENSG00000146047 | 6 | 25692137 | 25737573 | 144 | 6.121 | 4.65e-10 | 8.97e-06 |
| Red nucleus | INA | ENSG00000148798 | 10 | 105001920 | 105060108 | 95 | 6.11 | 4.97e-10 | 9.59e-06 |
| Red nucleus | SRPRB | ENSG00000144867 | 3 | 133467877 | 133554616 | 218 | 6.109 | 5e-10 | 9.65e-06 |
| Red nucleus | SLC17A1 | ENSG00000124568 | 6 | 25773125 | 25867287 | 232 | 6.109 | 5e-10 | 9.65e-06 |
| Red nucleus | TRIM38 | ENSG00000112343 | 6 | 25928030 | 25997384 | 160 | 6.109 | 5e-10 | 9.65e-06 |
| Red nucleus | HIST1H4A | ENSG00000196176 | 6 | 25986907 | 26032278 | 79 | 6.109 | 5e-10 | 9.65e-06 |
| Red nucleus | HIST1H4B | ENSG00000124529 | 6 | 26017124 | 26062480 | 89 | 6.109 | 5e-10 | 9.65e-06 |
| Red nucleus | HIST1H3B | ENSG00000124693 | 6 | 26021817 | 26067288 | 86 | 6.109 | 5e-10 | 9.65e-06 |
| Red nucleus | HIST1H2BB | ENSG00000196226 | 6 | 26033455 | 26078885 | 94 | 6.109 | 5e-10 | 9.65e-06 |
| Red nucleus | HIST1H1C | ENSG00000187837 | 6 | 26045968 | 26091699 | 101 | 6.109 | 5e-10 | 9.65e-06 |
| Red nucleus | BEST1 | ENSG00000167995 | 11 | 61682293 | 61742987 | 146 | 6.109 | 5e-10 | 9.65e-06 |
| Red nucleus | AP003733.1 | ENSG00000269089 | 11 | 61700453 | 61746755 | 116 | 6.109 | 5e-10 | 9.65e-06 |
| Red nucleus | FTH1 | ENSG00000167996 | 11 | 61717190 | 61770132 | 169 | 6.109 | 5e-10 | 9.65e-06 |
| Red nucleus | HIGD1C | ENSG00000214511 | 12 | 51312705 | 51374289 | 197 | 6.109 | 5e-10 | 9.65e-06 |
| Red nucleus | SLC11A2 | ENSG00000110911 | 12 | 51363184 | 51457349 | 233 | 6.109 | 5e-10 | 9.65e-06 |
| Red nucleus | LETMD1 | ENSG00000050426 | 12 | 51406745 | 51464207 | 134 | 6.109 | 5e-10 | 9.65e-06 |
| Red nucleus | C2orf76 | ENSG00000186132 | 2 | 120049801 | 120159404 | 351 | 6.071 | 6.37e-10 | 1.23e-05 |
| Red nucleus | TUBG1 | ENSG00000131462 | 17 | 40726694 | 40777252 | 78 | 6.029 | 8.26e-10 | 1.59e-05 |
| Red nucleus | MPST | ENSG00000128309 | 22 | 37380676 | 37435863 | 202 | 6.019 | 8.76e-10 | 1.69e-05 |
| Red nucleus | PSMC3IP | ENSG00000131470 | 17 | 40714333 | 40764849 | 75 | 5.976 | 1.14e-09 | 2.2e-05 |
| Red nucleus | RAB6B | ENSG00000154917 | 3 | 133533083 | 133649680 | 298 | 5.97 | 1.19e-09 | 2.3e-05 |
| Red nucleus | PTRF | ENSG00000177469 | 17 | 40544470 | 40610535 | 170 | 5.961 | 1.25e-09 | 2.42e-05 |
| Red nucleus | HIST1H1D | ENSG00000124575 | 6 | 26224440 | 26270216 | 104 | 5.892 | 1.91e-09 | 3.68e-05 |
| Red nucleus | FAM134C | ENSG00000141699 | 17 | 40721531 | 40797641 | 129 | 5.871 | 2.17e-09 | 4.19e-05 |
| Red nucleus | TUBG2 | ENSG00000037042 | 17 | 40776323 | 40829024 | 91 | 5.805 | 3.22e-09 | 6.23e-05 |
| Red nucleus | PANK2 | ENSG00000125779 | 20 | 3834486 | 3917605 | 201 | 5.671 | 7.09e-09 | 0.000137 |
| Red nucleus | AC021860.1 | ENSG00000196355 | 4 | 38618029 | 38701430 | 179 | 5.659 | 7.63e-09 | 0.000147 |
| Red nucleus | MYO15A | ENSG00000091536 | 17 | 17977020 | 18093116 | 186 | 5.655 | 7.8e-09 | 0.000151 |
| Red nucleus | RPEL1 | ENSG00000235376 | 10 | 104970644 | 105017773 | 67 | 5.639 | 8.54e-09 | 0.000165 |
| Red nucleus | TNK2 | ENSG00000061938 | 3 | 195580235 | 195673816 | 298 | 5.63 | 9.03e-09 | 0.000174 |
| Red nucleus | EHBP1 | ENSG00000115504 | 2 | 62865986 | 63283622 | 451 | 5.557 | 1.37e-08 | 0.000265 |
| Red nucleus | PCGF6 | ENSG00000156374 | 10 | 105052553 | 105145891 | 135 | 5.542 | 1.49e-08 | 0.000288 |
| Red nucleus | SLC17A4 | ENSG00000146039 | 6 | 25719927 | 25791419 | 244 | 5.46 | 2.39e-08 | 0.000461 |
| Red nucleus | TMPRSS12 | ENSG00000186452 | 12 | 51201703 | 51291667 | 192 | 5.447 | 2.56e-08 | 0.000494 |
| Red nucleus | FNIP2 | ENSG00000052795 | 4 | 159655290 | 159839201 | 245 | 5.447 | 2.56e-08 | 0.000495 |
| Red nucleus | MRC1L1 | ENSG00000183748 | 10 | 17816362 | 17963178 | 46 | 5.428 | 2.85e-08 | 0.000551 |
| Red nucleus | TAF5 | ENSG00000148835 | 10 | 105092724 | 105158822 | 88 | 5.419 | 2.99e-08 | 0.000577 |
| Red nucleus | AKAP10 | ENSG00000108599 | 17 | 19797615 | 19916656 | 182 | 5.381 | 3.71e-08 | 0.000715 |
| Red nucleus | CDV3 | ENSG00000091527 | 3 | 133257574 | 133319105 | 158 | 5.368 | 3.97e-08 | 0.000767 |
| Red nucleus | KLF3 | ENSG00000109787 | 4 | 38630817 | 38712663 | 176 | 5.362 | 4.11e-08 | 0.000793 |
| Red nucleus | IQCD | ENSG00000166578 | 12 | 113623246 | 113693899 | 110 | 5.355 | 4.27e-08 | 0.000825 |
| Red nucleus | TPCN1 | ENSG00000186815 | 12 | 113623855 | 113746390 | 208 | 5.34 | 4.65e-08 | 0.000897 |
| Red nucleus | CNTNAP1 | ENSG00000108797 | 17 | 40799631 | 40861832 | 73 | 5.326 | 5.02e-08 | 0.00097 |
| Red nucleus | DDX54 | ENSG00000123064 | 12 | 113584979 | 113658284 | 117 | 5.319 | 5.23e-08 | 0.00101 |
| Red nucleus | RITA1 | ENSG00000139405 | 12 | 113588331 | 113640173 | 87 | 5.275 | 6.64e-08 | 0.00128 |
| Red nucleus | USMG5 | ENSG00000173915 | 10 | 105138798 | 105191223 | 63 | 5.245 | 7.81e-08 | 0.00151 |
| Red nucleus | HIST1H2AA | ENSG00000164508 | 6 | 25716291 | 25761790 | 164 | 5.181 | 1.11e-07 | 0.00214 |
| Red nucleus | PPID | ENSG00000171497 | 4 | 159620286 | 159679548 | 76 | 5.18 | 1.11e-07 | 0.00214 |
| Red nucleus | COL3A1 | ENSG00000168542 | 2 | 189804046 | 189887472 | 170 | 5.098 | 1.72e-07 | 0.00332 |

| Trait | Gene symbol | Ensembl gene ID | Chromosome | Start (bp) | Stop (bp) | SNPs (n) | Z | P | Bonferroni-adjusted P |
|---|---|---|---|---|---|---|---|---|---|
| Red nucleus | PDCD11 | ENSG00000148843 | 10 | 105121405 | 105216049 | 120 | 5.077 | 1.92e-07 | 0.00371 |
| Red nucleus | ENTPD4 | ENSG00000197217 | 8 | 23233296 | 23350208 | 333 | 5.004 | 2.81e-07 | 0.00543 |
| Red nucleus | TMEM236 | ENSG00000184040 | 10 | 18006218 | 18099855 | 10 | 4.994 | 2.95e-07 | 0.0057 |
| Red nucleus | TMEM17 | ENSG00000186889 | 2 | 62717356 | 62774029 | 193 | 4.983 | 3.13e-07 | 0.00604 |
| Red nucleus | FLII | ENSG00000177731 | 17 | 18138150 | 18197230 | 107 | 4.982 | 3.15e-07 | 0.00608 |
| Red nucleus | FAM185A | ENSG00000222011 | 7 | 102354418 | 102459672 | 276 | 4.979 | 3.2e-07 | 0.00617 |
| Red nucleus | CCDC42B | ENSG00000186710 | 12 | 113552663 | 113607081 | 120 | 4.962 | 3.48e-07 | 0.00672 |
| Red nucleus | STAU1 | ENSG00000124214 | 20 | 47719878 | 47839904 | 255 | 4.956 | 3.59e-07 | 0.00694 |
| Red nucleus | FADS3 | ENSG00000221968 | 11 | 61630991 | 61694523 | 137 | 4.939 | 3.92e-07 | 0.00756 |
| Red nucleus | C4orf45 | ENSG00000164123 | 4 | 159804286 | 159994912 | 466 | 4.925 | 4.21e-07 | 0.00813 |
| Red nucleus | SHMT1 | ENSG00000176974 | 17 | 18221187 | 18301856 | 214 | 4.912 | 4.5e-07 | 0.00868 |
| Red nucleus | SNX31 | ENSG00000174226 | 8 | 101575116 | 101710643 | 447 | 4.89 | 5.03e-07 | 0.00972 |
| Red nucleus | GFAP | ENSG00000131095 | 17 | 42972376 | 43029305 | 164 | 4.886 | 5.16e-07 | 0.00996 |
| Red nucleus | PLEKHH3 | ENSG00000068137 | 17 | 40809932 | 40864048 | 54 | 4.873 | 5.5e-07 | 0.0106 |
| Red nucleus | KIF18B | ENSG00000186185 | 17 | 42992077 | 43060082 | 192 | 4.845 | 6.32e-07 | 0.0122 |
| Red nucleus | ANXA9 | ENSG00000143412 | 1 | 150919493 | 150978110 | 111 | 4.844 | 6.36e-07 | 0.0123 |
| Red nucleus | PRUNE | ENSG00000143363 | 1 | 150945896 | 151018189 | 159 | 4.838 | 6.56e-07 | 0.0127 |
| Red nucleus | CERS2 | ENSG00000143418 | 1 | 150923059 | 150982479 | 115 | 4.821 | 7.14e-07 | 0.0138 |
| Red nucleus | SPECC1 | ENSG00000128487 | 17 | 19877657 | 20232339 | 704 | 4.805 | 7.74e-07 | 0.0149 |
| Red nucleus | PDLIM2 | ENSG00000120913 | 8 | 22400792 | 22465538 | 126 | 4.792 | 8.24e-07 | 0.0159 |
| Red nucleus | TOP3A | ENSG00000177302 | 17 | 18164742 | 18253321 | 193 | 4.75 | 1.02e-06 | 0.0196 |
| Red nucleus | RAB3IL1 | ENSG00000167994 | 11 | 61654773 | 61722741 | 156 | 4.725 | 1.15e-06 | 0.0222 |
| Red nucleus | ABLIM1 | ENSG00000099204 | 10 | 116180872 | 116479762 | 787 | 4.719 | 1.18e-06 | 0.0229 |
| Red nucleus | PABPC1 | ENSG00000070756 | 8 | 101688044 | 101770037 | 150 | 4.718 | 1.19e-06 | 0.023 |
| Red nucleus | FP15737 | ENSG00000215298 | 8 | 23395157 | 23442974 | 291 | 4.696 | 1.33e-06 | 0.0256 |
| Red nucleus | MUC4 | ENSG00000145113 | 3 | 195463636 | 195574148 | 586 | 4.693 | 1.34e-06 | 0.0259 |
| Red nucleus | B4GALT5 | ENSG00000158470 | 20 | 48239482 | 48365415 | 336 | 4.69 | 1.37e-06 | 0.0264 |
| Red nucleus | MAFK | ENSG00000198517 | 7 | 1535350 | 1592679 | 213 | 4.688 | 1.38e-06 | 0.0266 |
| Red nucleus | CNNM2 | ENSG00000148842 | 10 | 104643050 | 104859978 | 496 | 4.661 | 1.57e-06 | 0.0304 |
| Red nucleus | GJA4 | ENSG00000187513 | 1 | 35223599 | 35271348 | 181 | 4.66 | 1.58e-06 | 0.0305 |
| Red nucleus | AS3MT | ENSG00000214435 | 10 | 104594273 | 104671656 | 213 | 4.653 | 1.64e-06 | 0.0316 |
| Red nucleus | EIF5A | ENSG00000132507 | 17 | 7175318 | 7225774 | 113 | 4.62 | 1.92e-06 | 0.0371 |
| Red nucleus | TEX2 | ENSG00000136478 | 17 | 62214587 | 62375661 | 345 | 4.606 | 2.05e-06 | 0.0396 |
| Red nucleus | UGP2 | ENSG00000169764 | 2 | 64033074 | 64128696 | 173 | 4.593 | 2.18e-06 | 0.0421 |
| Red nucleus | BCL7B | ENSG00000106635 | 7 | 72940686 | 73007332 | 124 | 4.587 | 2.24e-06 | 0.0433 |
| Substantia nigra | SLC39A8 | ENSG00000138821 | 4 | 103162198 | 103387415 | 653 | 9.036 | 8.09e-20 | 1.56e-15 |
| Substantia nigra | BANK1 | ENSG00000153064 | 4 | 102297443 | 103005969 | 1655 | 8.431 | 1.72e-17 | 3.32e-13 |
| Substantia nigra | COL5A2 | ENSG00000204262 | 2 | 189886622 | 190079605 | 456 | 8.187 | 1.34e-16 | 2.58e-12 |
| Substantia nigra | TF | ENSG00000091513 | 3 | 133429800 | 133507850 | 256 | 7.944 | 9.75e-16 | 1.88e-11 |
| Substantia nigra | TFRC | ENSG00000072274 | 3 | 195744054 | 195844060 | 428 | 7.664 | 9.01e-15 | 1.74e-10 |
| Substantia nigra | HIST1H1C | ENSG00000187837 | 6 | 26045968 | 26091699 | 101 | 7.581 | 1.71e-14 | 3.3e-10 |
| Substantia nigra | HFE | ENSG00000010704 | 6 | 26052509 | 26108571 | 134 | 7.578 | 1.76e-14 | 3.4e-10 |
| Substantia nigra | HIST1H3B | ENSG00000124693 | 6 | 26021817 | 26067288 | 86 | 7.56 | 2.02e-14 | 3.89e-10 |
| Substantia nigra | HIST1H3A | ENSG00000198366 | 6 | 25985718 | 26031186 | 80 | 7.484 | 3.6e-14 | 6.96e-10 |
| Substantia nigra | HIST1H2AB | ENSG00000137259 | 6 | 26023320 | 26068796 | 82 | 7.461 | 4.31e-14 | 8.32e-10 |
| Substantia nigra | SLC17A3 | ENSG00000124564 | 6 | 25823294 | 25917514 | 233 | 7.382 | 7.78e-14 | 1.5e-09 |
| Substantia nigra | HIST1H4C | ENSG00000197061 | 6 | 26069104 | 26114518 | 128 | 7.37 | 8.51e-14 | 1.64e-09 |
| Substantia nigra | HIST1H3C | ENSG00000196532 | 6 | 26010639 | 26056097 | 90 | 7.229 | 2.44e-13 | 4.71e-09 |
| Substantia nigra | TRIM38 | ENSG00000112343 | 6 | 25928030 | 25997384 | 160 | 7.188 | 3.29e-13 | 6.35e-09 |
| Substantia nigra | HIGD1C | ENSG00000214511 | 12 | 51312705 | 51374289 | 197 | 7.188 | 3.29e-13 | 6.36e-09 |
| Substantia nigra | SLC17A1 | ENSG00000124568 | 6 | 25773125 | 25867287 | 232 | 7.093 | 6.58e-13 | 1.27e-08 |
| Substantia nigra | SCGN | ENSG00000079689 | 6 | 25617464 | 25712011 | 282 | 6.795 | 5.42e-12 | 1.05e-07 |

| Trait | Gene symbol | Ensembl gene ID | Chromosome | Start (bp) | Stop (bp) | SNPs (n) | Z | P | Bonferroni-adjusted P |
|---|---|---|---|---|---|---|---|---|---|
| Substantia nigra | SLC17A2 | ENSG00000112337 | 6 | 25902982 | 25965946 | 182 | 6.769 | 6.47e-12 | 1.25e-07 |
| Substantia nigra | SLC11A2 | ENSG00000110911 | 12 | 51363184 | 51457349 | 233 | 6.714 | 9.5e-12 | 1.83e-07 |
| Substantia nigra | SRPRB | ENSG00000144867 | 3 | 133467877 | 133554616 | 218 | 6.441 | 5.94e-11 | 1.15e-06 |
| Substantia nigra | LRRC16A | ENSG00000079691 | 6 | 25244306 | 25630758 | 1409 | 6.433 | 6.27e-11 | 1.21e-06 |
| Substantia nigra | SLC39A12 | ENSG00000148482 | 10 | 18205768 | 18342221 | 469 | 6.399 | 7.8e-11 | 1.51e-06 |
| Substantia nigra | LETMD1 | ENSG00000050426 | 12 | 51406745 | 51464207 | 134 | 6.174 | 3.32e-10 | 6.41e-06 |
| Substantia nigra | RNF24 | ENSG00000101236 | 20 | 3897956 | 4031229 | 323 | 6.171 | 3.4e-10 | 6.56e-06 |
| Substantia nigra | WDR75 | ENSG00000115368 | 2 | 190271159 | 190350291 | 241 | 6.109 | 5e-10 | 9.65e-06 |
| Substantia nigra | HIST1H4A | ENSG00000196176 | 6 | 25986907 | 26032278 | 79 | 6.109 | 5e-10 | 9.65e-06 |
| Substantia nigra | HIST1H1A | ENSG00000124610 | 6 | 26007260 | 26053040 | 93 | 6.109 | 5e-10 | 9.65e-06 |
| Substantia nigra | HIST1H4B | ENSG00000124529 | 6 | 26017124 | 26062480 | 89 | 6.109 | 5e-10 | 9.65e-06 |
| Substantia nigra | HIST1H2BB | ENSG00000196226 | 6 | 26033455 | 26078885 | 94 | 6.109 | 5e-10 | 9.65e-06 |
| Substantia nigra | CSRNP2 | ENSG00000110925 | 12 | 51444990 | 51512447 | 166 | 5.918 | 1.63e-09 | 3.14e-05 |
| Substantia nigra | HIST1H2BA | ENSG00000146047 | 6 | 25692137 | 25737573 | 144 | 5.868 | 2.2e-09 | 4.25e-05 |
| Substantia nigra | SLC17A4 | ENSG00000146039 | 6 | 25719927 | 25791419 | 244 | 5.564 | 1.32e-08 | 0.000254 |
| Substantia nigra | TFCP2 | ENSG00000135457 | 12 | 51477446 | 51601926 | 308 | 5.366 | 4.02e-08 | 0.000777 |
| Substantia nigra | COL3A1 | ENSG00000168542 | 2 | 189804046 | 189887472 | 170 | 5.277 | 6.57e-08 | 0.00127 |
| Substantia nigra | HIST1H2AA | ENSG00000164508 | 6 | 25716291 | 25761790 | 164 | 5.258 | 7.29e-08 | 0.00141 |
| Substantia nigra | METTL7A | ENSG00000185432 | 12 | 51282255 | 51336300 | 120 | 5.245 | 7.82e-08 | 0.00151 |
| Substantia nigra | KCTD17 | ENSG00000100379 | 22 | 37412779 | 37469430 | 102 | 5.208 | 9.54e-08 | 0.00184 |
| Substantia nigra | AC021860.1 | ENSG00000196355 | 4 | 38618029 | 38701430 | 179 | 5.156 | 1.26e-07 | 0.00244 |
| Substantia nigra | TNK2 | ENSG00000061938 | 3 | 195580235 | 195673816 | 298 | 5.113 | 1.59e-07 | 0.00307 |
| Substantia nigra | PANK2 | ENSG00000125779 | 20 | 3834486 | 3917605 | 201 | 5.11 | 1.61e-07 | 0.00311 |
| Substantia nigra | SNX31 | ENSG00000174226 | 8 | 101575116 | 101710643 | 447 | 5.104 | 1.66e-07 | 0.00321 |
| Substantia nigra | DBI | ENSG00000155368 | 2 | 120089497 | 120140126 | 163 | 5.084 | 1.85e-07 | 0.00357 |
| Substantia nigra | PROX2 | ENSG00000119608 | 14 | 75309736 | 75365537 | 104 | 5.04 | 2.33e-07 | 0.00449 |
| Substantia nigra | DLST | ENSG00000119689 | 14 | 75313594 | 75380448 | 143 | 5.036 | 2.37e-07 | 0.00458 |
| Substantia nigra | KLF3 | ENSG00000109787 | 4 | 38630817 | 38712663 | 176 | 5.028 | 2.48e-07 | 0.0048 |
| Substantia nigra | PABPC1 | ENSG00000070756 | 8 | 101688044 | 101770037 | 150 | 5.026 | 2.5e-07 | 0.00483 |
| Substantia nigra | INA | ENSG00000148798 | 10 | 105001920 | 105060108 | 95 | 4.998 | 2.89e-07 | 0.00559 |
| Substantia nigra | YLPM1 | ENSG00000119596 | 14 | 75195069 | 75332244 | 193 | 4.964 | 3.45e-07 | 0.00666 |
| Substantia nigra | FLVCR2 | ENSG00000119686 | 14 | 76009960 | 76139557 | 282 | 4.955 | 3.61e-07 | 0.00698 |
| Substantia nigra | BCL7B | ENSG00000106635 | 7 | 72940686 | 73007332 | 124 | 4.942 | 3.87e-07 | 0.00747 |
| Substantia nigra | TMEM206 | ENSG00000065600 | 1 | 212527273 | 212623243 | 229 | 4.936 | 3.98e-07 | 0.00768 |
| Substantia nigra | FAM185A | ENSG00000222011 | 7 | 102354418 | 102459672 | 276 | 4.909 | 4.58e-07 | 0.00883 |
| Substantia nigra | RPEL1 | ENSG00000235376 | 10 | 104970644 | 105017773 | 67 | 4.892 | 4.98e-07 | 0.00961 |
| Substantia nigra | USMG5 | ENSG00000173915 | 10 | 105138798 | 105191223 | 63 | 4.875 | 5.43e-07 | 0.0105 |
| Substantia nigra | DUOX1 | ENSG00000137857 | 15 | 45387131 | 45467774 | 158 | 4.852 | 6.12e-07 | 0.0118 |
| Substantia nigra | HIST1H2AC | ENSG00000180573 | 6 | 26089373 | 26149344 | 158 | 4.849 | 6.21e-07 | 0.012 |
| Substantia nigra | LLGL1 | ENSG00000131899 | 17 | 18093901 | 18158189 | 135 | 4.848 | 6.25e-07 | 0.0121 |
| Substantia nigra | POLR2J2 | ENSG00000228049 | 7 | 102267474 | 102347076 | 71 | 4.841 | 6.46e-07 | 0.0125 |
| Substantia nigra | POLR2J2 | ENSG00000267645 | 7 | 102267496 | 102347088 | 71 | 4.841 | 6.46e-07 | 0.0125 |
| Substantia nigra | TIMM23 | ENSG00000138297 | 10 | 51582080 | 51658365 | 20 | 4.811 | 7.5e-07 | 0.0145 |
| Substantia nigra | C2orf76 | ENSG00000186132 | 2 | 120049801 | 120159404 | 351 | 4.791 | 8.31e-07 | 0.016 |
| Substantia nigra | AP003733.1 | ENSG00000269089 | 11 | 61700453 | 61746755 | 116 | 4.783 | 8.63e-07 | 0.0167 |
| Substantia nigra | TBL2 | ENSG00000106638 | 7 | 72973262 | 73028121 | 117 | 4.75 | 1.02e-06 | 0.0196 |
| Substantia nigra | ALKBH5 | ENSG00000091542 | 17 | 18051392 | 18123268 | 131 | 4.738 | 1.08e-06 | 0.0208 |
| Substantia nigra | NENF | ENSG00000117691 | 1 | 212571229 | 212629714 | 155 | 4.734 | 1.1e-06 | 0.0213 |
| Substantia nigra | PCGF6 | ENSG00000156374 | 10 | 105052553 | 105145891 | 135 | 4.731 | 1.12e-06 | 0.0215 |
| Substantia nigra | HLA-C | ENSG00000204525 | 6 | 31226526 | 31274907 | 1493 | 4.704 | 1.28e-06 | 0.0246 |
| Substantia nigra | PDCD11 | ENSG00000148843 | 10 | 105121405 | 105216049 | 120 | 4.697 | 1.32e-06 | 0.0254 |
| Substantia nigra | C4A | ENSG00000244731 | 6 | 31914801 | 31980458 | 151 | 4.696 | 1.33e-06 | 0.0257 |

| Trait | Gene symbol | Ensembl gene ID | Chromosome | Start (bp) | Stop (bp) | SNPs (n) | Z | P | Bonferroni-adjusted P |
|---|---|---|---|---|---|---|---|---|---|
| Substantia nigra | IFI35 | ENSG00000068079 | 17 | 41123742 | 41176473 | 73 | 4.66 | 1.58e-06 | 0.0306 |
| Substantia nigra | TAF5 | ENSG00000148835 | 10 | 105092724 | 105158822 | 88 | 4.659 | 1.59e-06 | 0.0307 |
| Substantia nigra | RPL27 | ENSG00000131469 | 17 | 41115290 | 41164976 | 73 | 4.65 | 1.66e-06 | 0.032 |
| Substantia nigra | LTBP2 | ENSG00000119681 | 14 | 74954873 | 75114306 | 351 | 4.643 | 1.72e-06 | 0.0332 |
| Substantia nigra | TST | ENSG00000128311 | 22 | 37396900 | 37450681 | 164 | 4.639 | 1.75e-06 | 0.0338 |
| Substantia nigra | C2 | ENSG00000166278 | 6 | 31830562 | 31923449 | 240 | 4.639 | 1.75e-06 | 0.0338 |
| Substantia nigra | HIST1H3F | ENSG00000256316 | 6 | 26240370 | 26285835 | 106 | 4.612 | 1.99e-06 | 0.0384 |
| Substantia nigra | PTGES3L | ENSG00000267060 | 17 | 41110105 | 41167545 | 81 | 4.604 | 2.07e-06 | 0.0399 |
| Substantia nigra | NELFE | ENSG00000204356 | 6 | 31909864 | 31961887 | 114 | 4.602 | 2.09e-06 | 0.0403 |
| Substantia nigra | CDSN | ENSG00000204539 | 6 | 31072867 | 31123223 | 522 | 4.601 | 2.1e-06 | 0.0406 |
| Substantia nigra | PTGES3L-AARSD1 | ENSG00000108825 | 17 | 41092543 | 41167545 | 95 | 4.595 | 2.17e-06 | 0.0418 |
| Substantia nigra | AREL1 | ENSG00000119682 | 14 | 75110140 | 75214818 | 112 | 4.564 | 2.5e-06 | 0.0484 |
| Substantia nigra | HIST1H2BI | ENSG00000168242 | 6 | 26238144 | 26283622 | 100 | 4.561 | 2.54e-06 | 0.0491 |

*Abbreviations: Chr = chromosome; MAGMA = Multi-marker Analysis of GenoMic Annotation; NSNPS = number of variants in the gene window; P_bonf = Bonferroni-adjusted P-value; ZSTAT = gene-based Z statistic. Gene windows extended 35 kb upstream and 10 kb downstream. One row per gene per region.*

### Supplementary Table 5. Putative causal genes for quantitative susceptibility mapping-derived brain iron identified by summary data-based Mendelian randomisation using whole blood (eQTLGen) and brain cortex (BrainMeta v2) expression quantitative trait loci.

| Trait | eQTL source | Gene symbol | Probe ID | Probe chromosome | Probe position (bp) | Top SNP | Top SNP chromosome | Top SNP position (bp) | Effect allele | Other allele | Effect allele frequency | Beta (GWAS) | SE (GWAS) | P (GWAS) | Beta (eQTL) | SE (eQTL) | P (eQTL) | Beta (SMR) | SE (SMR) | P (SMR) | P (HEIDI) | HEIDI SNPs (n) | Bonferroni-adjusted P (SMR) | Bonferroni threshold | Unique genes tested (n) | Tests performed (n) | Significant |
|---|---|---|---|---|---|---|---|---|---|---|---|---|---|---|---|---|---|---|---|---|---|---|---|---|---|---|---|
| Caudate nucleus | eQTLGen_Blood | DVL2 | ENSG00000004975 | 17 | 7133262 | rs2017365 | 17 | 7122624 | G | A | 0.3787 | -0.3495 | 0.06755 | 2.5e-07 | 0.1175 | 0.008241 | 3.689e-46 | -2.973 | 0.6113 | 1.154e-06 | 0.8952 | 20 | 0.01806 | 3.194e-06 | 15654 | 344388 | Yes |
| Caudate nucleus | eQTLGen_Blood | CTSH | ENSG00000103811 | 15 | 79227658 | rs34843303 | 15 | 79234470 | C | T | 0.1103 | -0.7619 | 0.1057 | 7.6e-15 | -0.8936 | 0.0117 | 0 | 0.8527 | 0.1188 | 7.13e-13 | 0.7355 | 20 | 1.116e-08 | 3.194e-06 | 15654 | 344388 | Yes |
| Caudate nucleus | eQTLGen_Blood | BCL7B | ENSG00000106635 | 7 | 72961509 | rs7786376 | 7 | 73042614 | G | A | 0.2435 | 0.7082 | 0.07404 | 3e-24 | -0.09835 | 0.009183 | 9.077e-27 | -7.201 | 1.009 | 9.721e-13 | 0.3057 | 20 | 1.522e-08 | 3.194e-06 | 15654 | 344388 | Yes |
| Caudate nucleus | eQTLGen_Blood | MLX | ENSG00000108788 | 17 | 40722171 | rs60929812 | 17 | 40759448 | A | G | 0.2654 | 0.9614 | 0.07268 | 1.9e-40 | 0.1479 | 0.008836 | 6.699e-63 | 6.5 | 0.6263 | 3.112e-25 | 0.01245 | 20 | 4.872e-21 | 3.194e-06 | 15654 | 344388 | Yes |
| Caudate nucleus | eQTLGen_Blood | BST1 | ENSG00000109743 | 4 | 15722254 | rs34559912 | 4 | 15730146 | G | T | 0.4374 | -0.3617 | 0.06609 | 1.2e-07 | 0.4827 | 0.007524 | 0 | -0.7493 | 0.1374 | 4.947e-08 | 0.01559 | 20 | 0.0007744 | 3.194e-06 | 15654 | 344388 | Yes |
| Caudate nucleus | eQTLGen_Blood | BANK1 | ENSG00000153064 | 4 | 102664206 | rs976654 | 4 | 102898570 | C | A | 0.4205 | 0.4105 | 0.06615 | 7.7e-10 | -0.07542 | 0.008036 | 6.292e-21 | -5.443 | 1.052 | 2.26e-07 | 0.04278 | 20 | 0.003537 | 3.194e-06 | 15654 | 344388 | Yes |
| Caudate nucleus | eQTLGen_Blood | IQCD | ENSG00000166578 | 12 | 113646072 | rs729062 | 12 | 113687859 | G | A | 0.08946 | 0.8061 | 0.1295 | 7.2e-11 | 0.1887 | 0.01758 | 7.186e-27 | 4.273 | 0.7934 | 7.232e-08 | 0.03636 | 20 | 0.001132 | 3.194e-06 | 15654 | 344388 | Yes |
| Caudate nucleus | eQTLGen_Blood | ELP5 | ENSG00000170291 | 17 | 7158997 | rs2074219 | 17 | 7156447 | A | G | 0.3648 | -0.3296 | 0.06724 | 2e-06 | -0.2667 | 0.009075 | 8.871e-190 | 1.236 | 0.2557 | 1.333e-06 | 0.644 | 20 | 0.02087 | 3.194e-06 | 15654 | 344388 | Yes |
| Caudate nucleus | eQTLGen_Blood | LINC01089 | ENSG00000212694 | 12 | 122237492 | rs1168664 | 12 | 122234676 | G | A | 0.1899 | -0.3974 | 0.08356 | 1.5e-05 | -0.5996 | 0.0115 | 0 | 0.6627 | 0.1399 | 2.182e-06 | 0.4873 | 20 | 0.03415 | 3.194e-06 | 15654 | 344388 | Yes |
| Caudate nucleus | eQTLGen_Blood | ENSG00000254184 | ENSG00000254184 | 7 | 72161247 | rs66954605 | 7 | 72196999 | A | G | 0.1938 | 0.3996 | 0.0828 | 1.6e-06 | 1.223 | 0.01615 | 0 | 0.3267 | 0.06783 | 1.463e-06 | 0.1828 | 20 | 0.0229 | 3.194e-06 | 15654 | 344388 | Yes |
| Dentate nucleus | eQTLGen_Blood | DVL2 | ENSG00000004975 | 17 | 7133262 | rs2017365 | 17 | 7122624 | G | A | 0.3787 | -0.9025 | 0.1621 | 5.7e-09 | 0.1175 | 0.008241 | 3.689e-46 | -7.678 | 1.48 | 2.138e-07 | 0.7656 | 20 | 0.003347 | 3.194e-06 | 15654 | 344388 | Yes |
| Dentate nucleus | eQTLGen_Blood | RNH1 | ENSG00000023191 | 11 | 500906 | rs12419766 | 11 | 503710 | T | C | 0.09742 | -1.686 | 0.2335 | 1.3e-12 | -0.329 | 0.01306 | 6.502e-140 | 5.125 | 0.7383 | 3.877e-12 | 0.025 | 20 | 6.069e-08 | 3.194e-06 | 15654 | 344388 | Yes |
| Dentate nucleus | eQTLGen_Blood | KIF1B | ENSG00000054523 | 1 | 10356262 | rs6702280 | 1 | 10280774 | A | G | 0.1252 | -1.345 | 0.236 | 4.2e-09 | 0.8977 | 0.01041 | 0 | -1.499 | 0.2634 | 1.28e-08 | 0.02595 | 20 | 0.0002003 | 3.194e-06 | 15654 | 344388 | Yes |
| Dentate nucleus | eQTLGen_Blood | NUCKS1 | ENSG00000069275 | 1 | 205700675 | rs823111 | 1 | 205701870 | T | C | 0.4085 | 0.9393 | 0.1589 | 3.2e-10 | -0.3202 | 0.007894 | 0 | -2.933 | 0.5013 | 4.887e-09 | 0.07164 | 20 | 7.65e-05 | 3.194e-06 | 15654 | 344388 | Yes |
| Dentate nucleus | eQTLGen_Blood | PYGB | ENSG00000100994 | 20 | 25253677 | rs6107017 | 20 | 25232300 | G | A | 0.4573 | 1.145 | 0.1572 | 8.7e-16 | 0.7483 | 0.006756 | 0 | 1.529 | 0.2105 | 3.73e-13 | 0.2807 | 20 | 5.838e-09 | 3.194e-06 | 15654 | 344388 | Yes |
| Dentate nucleus | eQTLGen_Blood | LMF1 | ENSG00000103227 | 16 | 967476 | rs4984720 | 16 | 981861 | G | A | 0.338 | 0.7903 | 0.1671 | 9.2e-07 | 0.2306 | 0.008218 | 2.651e-173 | 3.427 | 0.7348 | 3.116e-06 | 0.02142 | 20 | 0.04878 | 3.194e-06 | 15654 | 344388 | Yes |
| Dentate nucleus | eQTLGen_Blood | MOSPD3 | ENSG00000106330 | 7 | 100211366 | rs7812123 | 7 | 100211556 | A | C | 0.1869 | -1.072 | 0.1975 | 1.5e-08 | -0.3109 | 0.009717 | 1.129e-224 | 3.446 | 0.6441 | 8.768e-08 | 0.5929 | 20 | 0.001372 | 3.194e-06 | 15654 | 344388 | Yes |
| Dentate nucleus | eQTLGen_Blood | BCL7B | ENSG00000106635 | 7 | 72961509 | rs7786376 | 7 | 73042614 | G | A | 0.2435 | 0.942 | 0.1776 | 1.1e-07 | -0.09835 | 0.009183 | 9.077e-27 | -9.578 | 2.015 | 1.995e-06 | 0.09473 | 20 | 0.03123 | 3.194e-06 | 15654 | 344388 | Yes |
| Dentate nucleus | eQTLGen_Blood | EFTUD2 | ENSG00000108883 | 17 | 42952170 | rs1231641 | 17 | 42904720 | A | C | 0.4165 | 0.8569 | 0.1596 | 4.9e-08 | 0.2691 | 0.007963 | 2.019e-250 | 3.184 | 0.6004 | 1.143e-07 | 0.05484 | 20 | 0.001789 | 3.194e-06 | 15654 | 344388 | Yes |
| Dentate nucleus | eQTLGen_Blood | WDR75 | ENSG00000115368 | 2 | 190323225 | rs146991654 | 2 | 190243240 | G | A | 0.03678 | 3.264 | 0.4178 | 2.4e-16 | -0.4894 | 0.02112 | 7.952e-119 | -6.67 | 0.901 | 1.334e-13 | 0.01669 | 20 | 2.088e-09 | 3.194e-06 | 15654 | 344388 | Yes |
| Dentate nucleus | eQTLGen_Blood | RAB29 | ENSG00000117280 | 1 | 205740851 | rs7522056 | 1 | 205735891 | A | G | 0.3251 | -1.043 | 0.166 | 3.8e-11 | -0.3415 | 0.009108 | 1.286e-307 | 3.054 | 0.493 | 5.857e-10 | 0.01106 | 20 | 9.168e-06 | 3.194e-06 | 15654 | 344388 | Yes |
| Dentate nucleus | eQTLGen_Blood | MPL | ENSG00000117400 | 1 | 43810960 | rs11210839 | 1 | 43845658 | G | A | 0.3708 | 0.9035 | 0.1614 | 1.5e-08 | -0.1649 | 0.01234 | 9.788e-41 | -5.48 | 1.061 | 2.429e-07 | 0.2783 | 20 | 0.003802 | 3.194e-06 | 15654 | 344388 | Yes |
| Dentate nucleus | eQTLGen_Blood | TTLL5 | ENSG00000119685 | 14 | 76260694 | rs2059382 | 14 | 76158916 | T | C | 0.3101 | -1.008 | 0.1705 | 2e-10 | -0.312 | 0.008446 | 1.07e-298 | 3.231 | 0.5535 | 5.27e-09 | 0.3193 | 20 | 8.25e-05 | 3.194e-06 | 15654 | 344388 | Yes |
| Dentate nucleus | eQTLGen_Blood | OR2B6 | ENSG00000124657 | 6 | 27925489 | rs34899043 | 6 | 27893166 | C | T | 0.07753 | 3.093 | 0.2785 | 4.7e-31 | -0.1004 | 0.01737 | 7.408e-09 | -30.81 | 6.008 | 2.927e-07 | 0.04942 | 12 | 0.004582 | 3.194e-06 | 15654 | 344388 | Yes |
| Dentate nucleus | eQTLGen_Blood | SLC40A1 | ENSG00000138449 | 2 | 190436894 | rs7586757 | 2 | 190441286 | T | C | 0.3171 | -1.19 | 0.1702 | 3.7e-14 | 0.4568 | 0.008239 | 0 | -2.604 | 0.3755 | 4.056e-12 | 0.3184 | 20 | 6.349e-08 | 3.194e-06 | 15654 | 344388 | Yes |
| Dentate nucleus | eQTLGen_Blood | SUMF1 | ENSG00000144455 | 3 | 4125731 | rs4685744 | 3 | 4403537 | T | C | 0.5129 | -1.048 | 0.1573 | 5.1e-11 | -0.5082 | 0.007415 | 0 | 2.062 | 0.3111 | 3.407e-11 | 0.0237 | 20 | 5.333e-07 | 3.194e-06 | 15654 | 344388 | Yes |
| Dentate nucleus | eQTLGen_Blood | MED8 | ENSG00000159479 | 1 | 43852533 | rs839765 | 1 | 43834998 | G | A | 0.3718 | 0.9016 | 0.1614 | 1.6e-08 | -0.246 | 0.009384 | 1.728e-151 | -3.665 | 0.6707 | 4.628e-08 | 0.01646 | 20 | 0.0007245 | 3.194e-06 | 15654 | 344388 | Yes |
| Dentate nucleus | eQTLGen_Blood | TTC9C | ENSG00000162222 | 11 | 62501653 | rs6675 | 11 | 62505764 | T | C | 0.1938 | -1.073 | 0.1852 | 1.5e-08 | 0.2887 | 0.01057 | 3.28e-164 | -3.717 | 0.6557 | 1.443e-08 | 0.5052 | 20 | 0.0002259 | 3.194e-06 | 15654 | 344388 | Yes |
| Dentate nucleus | eQTLGen_Blood | ELP5 | ENSG00000170291 | 17 | 7158997 | rs2074219 | 17 | 7156447 | A | G | 0.3648 | -0.8744 | 0.1613 | 2.6e-08 | -0.2667 | 0.009075 | 8.871e-190 | 3.279 | 0.6153 | 9.852e-08 | 0.7966 | 20 | 0.001542 | 3.194e-06 | 15654 | 344388 | Yes |
| Dentate nucleus | eQTLGen_Blood | HDAC3 | ENSG00000171720 | 5 | 141008440 | rs1421896 | 5 | 141016288 | T | G | 0.4175 | 0.86 | 0.1619 | 7.1e-09 | -0.1236 | 0.01218 | 3.403e-24 | -6.956 | 1.478 | 2.513e-06 | 0.9639 | 20 | 0.03933 | 3.194e-06 | 15654 | 344388 | Yes |
| Dentate nucleus | eQTLGen_Blood | TMT1A | ENSG00000185432 | 12 | 51321777 | rs12809767 | 12 | 51249207 | C | A | 0.328 | 0.8189 | 0.1641 | 2.5e-07 | -0.2849 | 0.008405 | 7.655e-252 | -2.874 | 0.5821 | 7.898e-07 | 0.3977 | 20 | 0.01236 | 3.194e-06 | 15654 | 344388 | Yes |
| Dentate nucleus | eQTLGen_Blood | BTN3A2 | ENSG00000186470 | 6 | 26371966 | rs9357006 | 6 | 26364628 | C | A | 0.1123 | -1.22 | 0.231 | 5.7e-09 | -1.345 | 0.01019 | 0 | 0.9076 | 0.1719 | 1.301e-07 | 0.07597 | 20 | 0.002037 | 3.194e-06 | 15654 | 344388 | Yes |
| Dentate nucleus | eQTLGen_Blood | ZKSCAN4 | ENSG00000187626 | 6 | 28219706 | rs13200462 | 6 | 28218199 | C | A | 0.1143 | -1.058 | 0.2125 | 5.5e-08 | 0.2895 | 0.01155 | 1.465e-138 | -3.655 | 0.7484 | 1.042e-06 | 0.5172 | 20 | 0.01631 | 3.194e-06 | 15654 | 344388 | Yes |
| Dentate nucleus | eQTLGen_Blood | ZSCAN16 | ENSG00000196812 | 6 | 28095099 | rs9468287 | 6 | 28079741 | A | C | 0.1014 | -1.086 | 0.2178 | 7.2e-08 | 0.2365 | 0.01217 | 4.457e-84 | -4.592 | 0.951 | 1.371e-06 | 0.8284 | 20 | 0.02147 | 3.194e-06 | 15654 | 344388 | Yes |
| Dentate nucleus | eQTLGen_Blood | DDX39B | ENSG00000198563 | 6 | 31504110 | rs3093949 | 6 | 31525184 | T | C | 0.3101 | -0.7963 | 0.1638 | 2.4e-07 | -0.3359 | 0.01685 | 1.89e-88 | 2.371 | 0.5018 | 2.307e-06 | 0.01905 | 20 | 0.03611 | 3.194e-06 | 15654 | 344388 | Yes |
| Dentate nucleus | eQTLGen_Blood | HLA-E | ENSG00000204592 | 6 | 30459613 | rs1265094 | 6 | 31106893 | G | A | 0.5318 | -0.8901 | 0.1586 | 3.5e-10 | 0.1471 | 0.01251 | 6.562e-32 | -6.05 | 1.194 | 4.084e-07 | 0.1007 | 20 | 0.006393 | 3.194e-06 | 15654 | 344388 | Yes |
| Dentate nucleus | eQTLGen_Blood | MARCKSL1P1 | ENSG00000213277 | 10 | 104935581 | rs112699822 | 10 | 104692633 | A | C | 0.08946 | -1.738 | 0.2935 | 3.1e-09 | 0.2257 | 0.01849 | 2.769e-34 | -7.699 | 1.445 | 9.958e-08 | 0.03428 | 20 | 0.001559 | 3.194e-06 | 15654 | 344388 | Yes |
| Dentate nucleus | eQTLGen_Blood | CDC20-DT | ENSG00000234694 | 1 | 43822342 | rs72669002 | 1 | 43821071 | C | T | 0.3708 | 0.9079 | 0.1614 | 1.2e-08 | -0.2844 | 0.02158 | 1.175e-39 | -3.193 | 0.617 | 2.289e-07 | 0.119 | 20 | 0.003583 | 3.194e-06 | 15654 | 344388 | Yes |
| Dentate nucleus | eQTLGen_Blood | LMNTD2-AS1 | ENSG00000254815 | 11 | 558851 | rs35788385 | 11 | 570094 | A | G | 0.1928 | -1.147 | 0.1978 | 2.7e-08 | 0.2478 | 0.02477 | 1.42e-23 | -4.628 | 0.9224 | 5.234e-07 | 0.08156 | 20 | 0.008194 | 3.194e-06 | 15654 | 344388 | Yes |
| Dentate nucleus | eQTLGen_Blood | ENSG00000258985 | ENSG00000258985 | 14 | 53504221 | rs4901343 | 14 | 53565661 | C | T | 0.163 | 1.405 | 0.2132 | 6e-12 | -0.2663 | 0.02841 | 7.063e-21 | -5.276 | 0.9788 | 7.017e-08 | 0.02346 | 20 | 0.001098 | 3.194e-06 | 15654 | 344388 | Yes |
| Putamen | eQTLGen_Blood | DVL2 | ENSG00000004975 | 17 | 7133262 | rs2017365 | 17 | 7122624 | G | A | 0.3787 | -0.6458 | 0.1157 | 4.2e-08 | 0.1175 | 0.008241 | 3.689e-46 | -5.494 | 1.057 | 2.033e-07 | 0.9098 | 20 | 0.003183 | 3.194e-06 | 15654 | 344388 | Yes |
| Putamen | eQTLGen_Blood | STAB1 | ENSG00000010327 | 3 | 52543932 | rs9855470 | 3 | 52493275 | A | G | 0.06064 | -1.643 | 0.2434 | 2.2e-14 | -0.7503 | 0.01785 | 0 | 2.19 | 0.3286 | 2.661e-11 | 0.07125 | 20 | 4.165e-07 | 3.194e-06 | 15654 | 344388 | Yes |
| Putamen | eQTLGen_Blood | FNIP2 | ENSG00000052795 | 4 | 159759745 | rs35747642 | 4 | 159805370 | G | T | 0.3728 | 0.7815 | 0.1165 | 1.7e-11 | -0.2418 | 0.008162 | 7.737e-193 | -3.232 | 0.4941 | 6.076e-11 | 0.01684 | 20 | 9.512e-07 | 3.194e-06 | 15654 | 344388 | Yes |
| Putamen | eQTLGen_Blood | PACC1 | ENSG00000065600 | 1 | 212562758 | rs3767861 | 1 | 212537553 | A | G | 0.3678 | 0.6153 | 0.116 | 2.4e-07 | -0.2709 | 0.008957 | 5.713e-201 | -2.271 | 0.4347 | 1.747e-07 | 0.117 | 20 | 0.002734 | 3.194e-06 | 15654 | 344388 | Yes |

| Trait | eQTL source | Gene symbol | Probe ID | Probe chromosome | Probe position (bp) | Top SNP | Top SNP chromosome | Top SNP position (bp) | Effect allele | Other allele | Effect allele frequency | Beta (GWAS) | SE (GWAS) | P (GWAS) | Beta (eQTL) | SE (eQTL) | P (eQTL) | Beta (SMR) | SE (SMR) | P (SMR) | P (HEIDI) | HEIDI SNPs (n) | Bonferroni-adjusted P (SMR) | Bonferroni threshold | Unique genes tested (n) | Tests performed (n) | Significant |
|---|---|---|---|---|---|---|---|---|---|---|---|---|---|---|---|---|---|---|---|---|---|---|---|---|---|---|---|
| Putamen | eQTLGen_Blood | TRIP6 | ENSG00000087077 | 7 | 100467918 | rs3757868 | 7 | 100482720 | A | G | 0.1829 | -0.7406 | 0.1461 | 2.1e-07 | -0.3709 | 0.01686 | 3.345e-107 | 1.997 | 0.4043 | 7.803e-07 | 0.3658 | 20 | 0.01221 | 3.194e-06 | 15654 | 344388 | Yes |
| Putamen | eQTLGen_Blood | CTSH | ENSG00000103811 | 15 | 79227658 | rs34843303 | 15 | 79234470 | C | T | 0.1103 | -1.781 | 0.1813 | 3.7e-25 | -0.8936 | 0.0117 | 0 | 1.993 | 0.2045 | 1.938e-22 | 0.3914 | 20 | 3.033e-18 | 3.194e-06 | 15654 | 344388 | Yes |
| Putamen | eQTLGen_Blood | BCL7B | ENSG00000106635 | 7 | 72961509 | rs7786376 | 7 | 73042614 | G | A | 0.2435 | 1.556 | 0.1269 | 1.7e-36 | -0.09835 | 0.009183 | 9.077e-27 | -15.82 | 1.961 | 7.164e-16 | 0.1278 | 20 | 1.121e-11 | 3.194e-06 | 15654 | 344388 | Yes |
| Putamen | eQTLGen_Blood | TUBD1 | ENSG00000108423 | 17 | 57953577 | rs112911630 | 17 | 57940172 | A | G | 0.1551 | 1.252 | 0.1536 | 3.3e-17 | -0.1593 | 0.01252 | 4.614e-37 | -7.857 | 1.145 | 6.807e-12 | 0.01698 | 20 | 1.065e-07 | 3.194e-06 | 15654 | 344388 | Yes |
| Putamen | eQTLGen_Blood | MLX | ENSG00000108788 | 17 | 40722171 | rs60929812 | 17 | 40759448 | A | G | 0.2654 | 1.787 | 0.1245 | 1.2e-48 | 0.1479 | 0.008836 | 6.699e-63 | 12.08 | 1.109 | 1.231e-27 | 0.1172 | 20 | 1.927e-23 | 3.194e-06 | 15654 | 344388 | Yes |
| Putamen | eQTLGen_Blood | FNBP4 | ENSG00000109920 | 11 | 47763533 | rs11039337 | 11 | 47707156 | C | T | 0.4304 | -0.6512 | 0.1132 | 1.9e-08 | -0.1343 | 0.007995 | 2.49e-63 | 4.849 | 0.891 | 5.276e-08 | 0.3915 | 20 | 0.0008259 | 3.194e-06 | 15654 | 344388 | Yes |
| Putamen | eQTLGen_Blood | OR2B6 | ENSG00000124657 | 6 | 27925489 | rs34899043 | 6 | 27893166 | C | T | 0.07753 | 1.574 | 0.1992 | 2.8e-16 | -0.1004 | 0.01737 | 7.408e-09 | -15.68 | 3.36 | 3.079e-06 | 0.03581 | 12 | 0.0482 | 3.194e-06 | 15654 | 344388 | Yes |
| Putamen | eQTLGen_Blood | HROB | ENSG00000125319 | 17 | 42229559 | rs730228 | 17 | 42239349 | C | T | 0.3022 | 0.6112 | 0.1248 | 2.9e-06 | -0.1355 | 0.00895 | 9.46e-52 | -4.512 | 0.9686 | 3.187e-06 | 0.0736 | 20 | 0.04989 | 3.194e-06 | 15654 | 344388 | Yes |
| Putamen | eQTLGen_Blood | PSMC3IP | ENSG00000131470 | 17 | 40727091 | rs12601457 | 17 | 40780847 | T | C | 0.2654 | 1.787 | 0.1245 | 9e-49 | 0.08991 | 0.008942 | 8.732e-24 | 19.87 | 2.413 | 1.799e-16 | 0.01948 | 20 | 2.817e-12 | 3.194e-06 | 15654 | 344388 | Yes |
| Putamen | eQTLGen_Blood | RITA1 | ENSG00000139405 | 12 | 113626752 | rs2303616 | 12 | 113729956 | A | G | 0.09344 | 1.799 | 0.2213 | 4e-16 | -0.6277 | 0.0158 | 0 | -2.865 | 0.36 | 1.713e-15 | 0.01079 | 20 | 2.682e-11 | 3.194e-06 | 15654 | 344388 | Yes |
| Putamen | eQTLGen_Blood | TCF12 | ENSG00000140262 | 15 | 57401150 | rs9672676 | 15 | 57496277 | C | T | 0.1938 | -0.7659 | 0.1507 | 1.8e-07 | -0.1685 | 0.01024 | 7.521e-61 | 4.545 | 0.9359 | 1.194e-06 | 0.336 | 20 | 0.01869 | 3.194e-06 | 15654 | 344388 | Yes |
| Putamen | eQTLGen_Blood | USP53 | ENSG00000145390 | 4 | 120175207 | rs7678138 | 4 | 120106766 | A | G | 0.1282 | 0.8137 | 0.1703 | 4.5e-05 | 0.8999 | 0.01079 | 0 | 0.9042 | 0.1895 | 1.833e-06 | 0.02576 | 20 | 0.02869 | 3.194e-06 | 15654 | 344388 | Yes |
| Putamen | eQTLGen_Blood | BRI3 | ENSG00000164713 | 7 | 97909426 | rs6970990 | 7 | 97925382 | G | T | 0.4811 | 0.5456 | 0.1122 | 4.5e-07 | -0.2746 | 0.01162 | 2.205e-123 | -1.987 | 0.4171 | 1.904e-06 | 0.03525 | 20 | 0.0298 | 3.194e-06 | 15654 | 344388 | Yes |
| Putamen | eQTLGen_Blood | IQCD | ENSG00000166578 | 12 | 113646072 | rs729062 | 12 | 113687859 | G | A | 0.08946 | 1.817 | 0.2219 | 2.7e-16 | 0.1887 | 0.01758 | 7.186e-27 | 9.631 | 1.479 | 7.533e-11 | 0.02025 | 20 | 1.179e-06 | 3.194e-06 | 15654 | 344388 | Yes |
| Putamen | eQTLGen_Blood | ELP5 | ENSG00000170291 | 17 | 7158997 | rs2074219 | 17 | 7156447 | A | G | 0.3648 | -0.6359 | 0.1152 | 6.3e-08 | -0.2667 | 0.009075 | 8.871e-190 | 2.385 | 0.4396 | 5.826e-08 | 0.3959 | 20 | 0.000912 | 3.194e-06 | 15654 | 344388 | Yes |
| Putamen | eQTLGen_Blood | LRRC37A4P | ENSG00000214425 | 17 | 43603193 | rs55669501 | 17 | 44202564 | A | G | 0.2406 | 1.439 | 0.1356 | 5.3e-29 | -1.238 | 0.01732 | 0 | -1.162 | 0.1107 | 9.453e-26 | 0.01889 | 20 | 1.48e-21 | 3.194e-06 | 15654 | 344388 | Yes |
| Putamen | eQTLGen_Blood | ENSG00000261575 | ENSG00000261575 | 17 | 44345231 | rs55686102 | 17 | 44202608 | T | C | 0.2406 | 1.438 | 0.1357 | 5.6e-29 | 0.9018 | 0.02488 | 9.653e-288 | 1.595 | 0.1567 | 2.512e-24 | 0.04539 | 20 | 3.933e-20 | 3.194e-06 | 15654 | 344388 | Yes |
| Putamen | eQTLGen_Blood | MAPK8IP1P1 | ENSG00000262500 | 17 | 44321691 | rs55686102 | 17 | 44202608 | T | C | 0.2406 | 1.438 | 0.1357 | 5.6e-29 | 0.3535 | 0.01386 | 1.488e-143 | 4.069 | 0.4156 | 1.227e-22 | 0.1989 | 20 | 1.92e-18 | 3.194e-06 | 15654 | 344388 | Yes |
| Putamen | eQTLGen_Blood | MAPK8IP1P2 | ENSG00000263503 | 17 | 43678970 | rs55669501 | 17 | 44202564 | A | G | 0.2406 | 1.439 | 0.1356 | 5.3e-29 | 1.201 | 0.01779 | 0 | 1.198 | 0.1143 | 1.094e-25 | 0.05678 | 20 | 1.712e-21 | 3.194e-06 | 15654 | 344388 | Yes |
| Putamen | eQTLGen_Blood | DND1P1 | ENSG00000264070 | 17 | 43663766 | rs113991678 | 17 | 43795634 | T | C | 0.2386 | 1.43 | 0.1356 | 7.2e-29 | 1.178 | 0.02033 | 0 | 1.214 | 0.117 | 3.245e-25 | 0.01905 | 20 | 5.08e-21 | 3.194e-06 | 15654 | 344388 | Yes |
| Globus pallidus | eQTLGen_Blood | FNIP2 | ENSG00000052795 | 4 | 159759745 | rs35747642 | 4 | 159805370 | G | T | 0.3728 | 0.8195 | 0.1709 | 9.8e-07 | -0.2418 | 0.008162 | 7.737e-193 | -3.39 | 0.716 | 2.201e-06 | 0.03065 | 20 | 0.03445 | 3.194e-06 | 15654 | 344388 | Yes |
| Globus pallidus | eQTLGen_Blood | KIF1B | ENSG00000054523 | 1 | 10356262 | rs6702280 | 1 | 10280774 | A | G | 0.1252 | -1.247 | 0.2483 | 3.8e-06 | 0.8977 | 0.01041 | 0 | -1.389 | 0.277 | 5.339e-07 | 0.6159 | 20 | 0.008358 | 3.194e-06 | 15654 | 344388 | Yes |
| Globus pallidus | eQTLGen_Blood | CTSH | ENSG00000103811 | 15 | 79227658 | rs34843303 | 15 | 79234470 | C | T | 0.1103 | -1.688 | 0.2663 | 2.7e-11 | -0.8936 | 0.0117 | 0 | 1.889 | 0.299 | 2.698e-10 | 0.871 | 20 | 4.223e-06 | 3.194e-06 | 15654 | 344388 | Yes |
| Globus pallidus | eQTLGen_Blood | BCL7B | ENSG00000106635 | 7 | 72961509 | rs7786376 | 7 | 73042614 | G | A | 0.2435 | 1.087 | 0.1865 | 1.3e-09 | -0.09835 | 0.009183 | 9.077e-27 | -11.05 | 2.158 | 3.043e-07 | 0.7866 | 20 | 0.004764 | 3.194e-06 | 15654 | 344388 | Yes |
| Globus pallidus | eQTLGen_Blood | FNBP4 | ENSG00000109920 | 11 | 47763533 | rs11039337 | 11 | 47707156 | C | T | 0.4304 | -0.8149 | 0.1662 | 6.1e-07 | -0.1343 | 0.007995 | 2.49e-63 | 6.068 | 1.289 | 2.507e-06 | 0.04002 | 20 | 0.03924 | 3.194e-06 | 15654 | 344388 | Yes |
| Globus pallidus | eQTLGen_Blood | ARHGAP22 | ENSG00000128805 | 10 | 49759193 | rs1822861 | 10 | 49834326 | T | G | 0.497 | 0.889 | 0.1656 | 9.7e-09 | 0.3592 | 0.007716 | 0 | 2.475 | 0.4642 | 9.708e-08 | 0.03979 | 20 | 0.00152 | 3.194e-06 | 15654 | 344388 | Yes |
| Globus pallidus | eQTLGen_Blood | XPO4 | ENSG00000132953 | 13 | 21414328 | rs9509423 | 13 | 21467197 | C | T | 0.5169 | 0.892 | 0.1659 | 3.1e-08 | -0.1386 | 0.008335 | 4.128e-62 | -6.435 | 1.258 | 3.106e-07 | 0.1722 | 20 | 0.004862 | 3.194e-06 | 15654 | 344388 | Yes |
| Globus pallidus | eQTLGen_Blood | BANK1 | ENSG00000153064 | 4 | 102664206 | rs976654 | 4 | 102898570 | C | A | 0.4205 | 1.125 | 0.1665 | 7.9e-13 | -0.07542 | 0.008036 | 6.292e-21 | -14.92 | 2.721 | 4.172e-08 | 0.01374 | 20 | 0.0006531 | 3.194e-06 | 15654 | 344388 | Yes |
| Globus pallidus | eQTLGen_Blood | DBI | ENSG00000155368 | 2 | 120127311 | rs12612942 | 2 | 120118237 | G | A | 0.1998 | -1.045 | 0.2021 | 2e-08 | -0.3627 | 0.01438 | 2.275e-140 | 2.88 | 0.5688 | 4.105e-07 | 0.09683 | 20 | 0.006426 | 3.194e-06 | 15654 | 344388 | Yes |
| Globus pallidus | eQTLGen_Blood | CTSS | ENSG00000163131 | 1 | 150720492 | rs41271951 | 1 | 150737220 | G | A | 0.07654 | -1.604 | 0.2948 | 5e-10 | -0.5742 | 0.01594 | 3.911e-284 | 2.794 | 0.5193 | 7.437e-08 | 0.1599 | 20 | 0.001164 | 3.194e-06 | 15654 | 344388 | Yes |
| Globus pallidus | eQTLGen_Blood | SLC20A2 | ENSG00000168575 | 8 | 42335531 | rs2923446 | 8 | 42404177 | G | A | 0.4165 | -0.9507 | 0.1685 | 4.2e-09 | 0.2953 | 0.008622 | 5.45e-257 | -3.22 | 0.5785 | 2.601e-08 | 0.6704 | 20 | 0.0004072 | 3.194e-06 | 15654 | 344388 | Yes |
| Globus pallidus | eQTLGen_Blood | SMIM19 | ENSG00000176209 | 8 | 42402224 | rs3099936 | 8 | 42384331 | T | C | 0.3907 | -0.8977 | 0.1701 | 3e-08 | 0.5677 | 0.008235 | 0 | -1.581 | 0.3005 | 1.423e-07 | 0.8731 | 20 | 0.002227 | 3.194e-06 | 15654 | 344388 | Yes |
| Globus pallidus | eQTLGen_Blood | SND1 | ENSG00000197157 | 7 | 127512447 | rs12706818 | 7 | 127501613 | T | C | 0.2803 | -0.9175 | 0.1814 | 3.3e-07 | -0.1983 | 0.009234 | 2.61e-102 | 4.626 | 0.9395 | 8.48e-07 | 0.2244 | 20 | 0.01327 | 3.194e-06 | 15654 | 344388 | Yes |
| Globus pallidus | eQTLGen_Blood | NELFE | ENSG00000204356 | 6 | 31923375 | rs541862 | 6 | 31916951 | C | T | 0.08847 | 1.395 | 0.2793 | 2.9e-07 | -0.4627 | 0.0298 | 2.292e-54 | -3.016 | 0.6342 | 1.983e-06 | 0.3203 | 20 | 0.03105 | 3.194e-06 | 15654 | 344388 | Yes |
| Globus pallidus | eQTLGen_Blood | MIR570 | ENSG00000207650 | 3 | 195426320 | rs11926771 | 3 | 195656731 | A | C | 0.1879 | -1.194 | 0.2204 | 2.7e-07 | 0.2584 | 0.01405 | 1.539e-75 | -4.621 | 0.8892 | 2.027e-07 | 0.2909 | 20 | 0.003173 | 3.194e-06 | 15654 | 344388 | Yes |
| Globus pallidus | eQTLGen_Blood | ENSG00000225059 | ENSG00000225059 | 6 | 31352008 | rs2516396 | 6 | 31521913 | A | G | 0.2435 | 1.16 | 0.1876 | 1.6e-10 | -0.4186 | 0.02946 | 8.08e-46 | -2.772 | 0.4888 | 1.421e-08 | 0.1612 | 20 | 0.0002224 | 3.194e-06 | 15654 | 344388 | Yes |
| Globus pallidus | eQTLGen_Blood | PLEKHM1 | ENSG00000225190 | 17 | 43540690 | rs55663797 | 17 | 43544379 | A | G | 0.1958 | 1.04 | 0.2147 | 2e-06 | 0.2131 | 0.01102 | 2.924e-83 | 4.88 | 1.039 | 2.624e-06 | 0.03424 | 20 | 0.04107 | 3.194e-06 | 15654 | 344388 | Yes |
| Globus pallidus | eQTLGen_Blood | HCG22 | ENSG00000228789 | 6 | 31024447 | rs1265054 | 6 | 31079643 | C | T | 0.5149 | -0.9592 | 0.1651 | 1.7e-10 | -0.4616 | 0.02 | 7.918e-118 | 2.078 | 0.3688 | 1.76e-08 | 0.02938 | 20 | 0.0002755 | 3.194e-06 | 15654 | 344388 | Yes |
| Red nucleus | eQTLGen_Blood | DVL2 | ENSG00000004975 | 17 | 7133262 | rs2017365 | 17 | 7122624 | G | A | 0.3787 | -1.081 | 0.1521 | 5.1e-14 | 0.1175 | 0.008241 | 3.689e-46 | -9.193 | 1.445 | 2.017e-10 | 0.7531 | 20 | 3.157e-06 | 3.194e-06 | 15654 | 344388 | Yes |
| Red nucleus | eQTLGen_Blood | ACADVL | ENSG00000072778 | 17 | 7124518 | rs507506 | 17 | 7118322 | A | G | 0.4165 | -0.9637 | 0.15 | 1.1e-11 | 0.3217 | 0.007951 | 0 | -2.996 | 0.4721 | 2.207e-10 | 0.07556 | 20 | 3.454e-06 | 3.194e-06 | 15654 | 344388 | Yes |
| Red nucleus | eQTLGen_Blood | AKAP10 | ENSG00000108599 | 17 | 19844635 | rs203477 | 17 | 19831819 | C | A | 0.3946 | 0.8361 | 0.1506 | 1.2e-08 | 0.6212 | 0.007441 | 0 | 1.346 | 0.243 | 3.038e-08 | 0.8521 | 20 | 0.0004756 | 3.194e-06 | 15654 | 344388 | Yes |
| Red nucleus | eQTLGen_Blood | MLX | ENSG00000108788 | 17 | 40722171 | rs60929812 | 17 | 40759448 | A | G | 0.2654 | 1.278 | 0.1636 | 3.3e-16 | 0.1479 | 0.008836 | 6.699e-63 | 8.637 | 1.221 | 1.486e-12 | 0.2792 | 20 | 2.326e-08 | 3.194e-06 | 15654 | 344388 | Yes |
| Red nucleus | eQTLGen_Blood | EFTUD2 | ENSG00000108883 | 17 | 42952170 | rs1231641 | 17 | 42904720 | A | C | 0.4165 | 0.7492 | 0.1497 | 9.7e-07 | 0.2691 | 0.007963 | 2.019e-250 | 2.784 | 0.5625 | 7.459e-07 | 0.1568 | 20 | 0.01168 | 3.194e-06 | 15654 | 344388 | Yes |
| Red nucleus | eQTLGen_Blood | WDR75 | ENSG00000115368 | 2 | 190323225 | rs146991654 | 2 | 190243240 | G | A | 0.03678 | 4.734 | 0.3924 | 2e-36 | -0.4894 | 0.02112 | 7.952e-119 | -9.675 | 0.904 | 9.956e-27 | 0.03382 | 20 | 1.559e-22 | 3.194e-06 | 15654 | 344388 | Yes |
| Red nucleus | eQTLGen_Blood | OTX1 | ENSG00000115507 | 2 | 63281081 | rs76298426 | 2 | 63287328 | C | T | 0.1064 | 1.39 | 0.2137 | 1.1e-11 | 0.5109 | 0.0115 | 0 | 2.72 | 0.4227 | 1.23e-10 | 0.02727 | 20 | 1.925e-06 | 3.194e-06 | 15654 | 344388 | Yes |

| Trait | eQTL source | Gene symbol | Probe ID | Probe chromosome | Probe position (bp) | Top SNP | Top SNP chromosome | Top SNP position (bp) | Effect allele | Other allele | Effect allele frequency | Beta (GWAS) | SE (GWAS) | P (GWAS) | Beta (eQTL) | SE (eQTL) | P (eQTL) | Beta (SMR) | SE (SMR) | P (SMR) | P (HEIDI) | HEIDI SNPs (n) | Bonferroni-adjusted P (SMR) | Bonferroni threshold | Unique genes tested (n) | Tests performed (n) | Significant |
|---|---|---|---|---|---|---|---|---|---|---|---|---|---|---|---|---|---|---|---|---|---|---|---|---|---|---|---|
| Red nucleus | eQTLGen_Blood | TRIM47 | ENSG00000132481 | 17 | 73872449 | rs66496607 | 17 | 73877459 | T | C | 0.1352 | 1.034 | 0.2123 | 3.2e-07 | -0.2701 | 0.01107 | 2.248e-131 | -3.828 | 0.8016 | 1.797e-06 | 0.08218 | 20 | 0.02813 | 3.194e-06 | 15654 | 344388 | Yes |
| Red nucleus | eQTLGen_Blood | RITA1 | ENSG00000139405 | 12 | 113626752 | rs2303616 | 12 | 113729956 | A | G | 0.09344 | 1.54 | 0.2906 | 1.6e-07 | -0.6277 | 0.0158 | 0 | -2.454 | 0.467 | 1.48e-07 | 0.02043 | 20 | 0.002317 | 3.194e-06 | 15654 | 344388 | Yes |
| Red nucleus | eQTLGen_Blood | ACSL1 | ENSG00000151726 | 4 | 185712360 | rs2046814 | 4 | 185704176 | G | T | 0.4215 | -0.9612 | 0.1496 | 1.6e-10 | -0.06123 | 0.008055 | 2.948e-14 | 15.7 | 3.199 | 9.234e-07 | 0.2965 | 20 | 0.01445 | 3.194e-06 | 15654 | 344388 | Yes |
| Red nucleus | eQTLGen_Blood | DBI | ENSG00000155368 | 2 | 120127311 | rs12612942 | 2 | 120118237 | G | A | 0.1998 | -1.049 | 0.1806 | 3.8e-10 | -0.3627 | 0.01438 | 2.275e-140 | 2.893 | 0.5109 | 1.49e-08 | 0.07398 | 20 | 0.0002333 | 3.194e-06 | 15654 | 344388 | Yes |
| Red nucleus | eQTLGen_Blood | IQCD | ENSG00000166578 | 12 | 113646072 | rs729062 | 12 | 113687859 | G | A | 0.08946 | 1.577 | 0.2913 | 7.4e-08 | 0.1887 | 0.01758 | 7.186e-27 | 8.358 | 1.729 | 1.345e-06 | 0.2268 | 20 | 0.02105 | 3.194e-06 | 15654 | 344388 | Yes |
| Red nucleus | eQTLGen_Blood | POLR3D | ENSG00000168495 | 8 | 22106051 | rs4871992 | 8 | 22146385 | A | G | 0.505 | 0.8078 | 0.149 | 3.7e-08 | -0.08518 | 0.007933 | 6.778e-27 | -9.484 | 1.96 | 1.306e-06 | 0.1597 | 20 | 0.02045 | 3.194e-06 | 15654 | 344388 | Yes |
| Red nucleus | eQTLGen_Blood | ELP5 | ENSG00000170291 | 17 | 7158997 | rs2074219 | 17 | 7156447 | A | G | 0.3648 | -0.9623 | 0.1514 | 1.4e-11 | -0.2667 | 0.009075 | 8.871e-190 | 3.609 | 0.5808 | 5.198e-10 | 0.06344 | 20 | 8.136e-06 | 3.194e-06 | 15654 | 344388 | Yes |
| Red nucleus | eQTLGen_Blood | MUC20 | ENSG00000176945 | 3 | 195457873 | rs3103934 | 3 | 195477088 | A | G | 0.1531 | 0.9302 | 0.198 | 3.2e-06 | 0.7071 | 0.01032 | 0 | 1.315 | 0.2807 | 2.774e-06 | 0.148 | 20 | 0.04343 | 3.194e-06 | 15654 | 344388 | Yes |
| Red nucleus | eQTLGen_Blood | TMT1A | ENSG00000185432 | 12 | 51321777 | rs12809767 | 12 | 51249207 | C | A | 0.328 | 0.7311 | 0.154 | 5e-07 | -0.2849 | 0.008405 | 7.655e-252 | -2.566 | 0.5459 | 2.591e-06 | 0.2829 | 20 | 0.04057 | 3.194e-06 | 15654 | 344388 | Yes |
| Red nucleus | eQTLGen_Blood | ENSG00000197459 | ENSG00000197459 | 6 | 26252091 | rs59315883 | 6 | 26302407 | G | A | 0.2127 | 1.163 | 0.1949 | 4.5e-09 | 0.08383 | 0.01019 | 1.913e-16 | 13.87 | 2.872 | 1.362e-06 | 0.2515 | 20 | 0.02132 | 3.194e-06 | 15654 | 344388 | Yes |
| Substantia nigra | eQTLGen_Blood | TRIM38 | ENSG00000112343 | 6 | 25974189 | rs1130000 | 6 | 25985396 | G | A | 0.4374 | 1.27 | 0.1784 | 1.6e-13 | 0.2619 | 0.007985 | 7.483e-236 | 4.849 | 0.697 | 3.473e-12 | 0.01117 | 20 | 5.437e-08 | 3.194e-06 | 15654 | 344388 | Yes |
| Substantia nigra | eQTLGen_Blood | WDR75 | ENSG00000115368 | 2 | 190323225 | rs146991654 | 2 | 190243240 | G | A | 0.03678 | 6.519 | 0.469 | 4e-44 | -0.4894 | 0.02112 | 7.952e-119 | -13.32 | 1.117 | 9.16e-33 | 0.0428 | 20 | 1.434e-28 | 3.194e-06 | 15654 | 344388 | Yes |
| Substantia nigra | eQTLGen_Blood | BLTP1 | ENSG00000138688 | 4 | 123178700 | rs17454584 | 4 | 123353432 | G | A | 0.2396 | -1.2 | 0.2155 | 2.6e-08 | -0.1202 | 0.009273 | 2.073e-38 | 9.983 | 1.952 | 3.146e-07 | 0.617 | 20 | 0.004924 | 3.194e-06 | 15654 | 344388 | Yes |
| Substantia nigra | eQTLGen_Blood | BANK1 | ENSG00000153064 | 4 | 102664206 | rs976654 | 4 | 102898570 | C | A | 0.4205 | 1.012 | 0.178 | 8.3e-09 | -0.07542 | 0.008036 | 6.292e-21 | -13.41 | 2.76 | 1.167e-06 | 0.0292 | 20 | 0.01827 | 3.194e-06 | 15654 | 344388 | Yes |
| Substantia nigra | eQTLGen_Blood | DBI | ENSG00000155368 | 2 | 120127311 | rs12612942 | 2 | 120118237 | G | A | 0.1998 | -1.067 | 0.2158 | 2.4e-07 | -0.3627 | 0.01438 | 2.275e-140 | 2.943 | 0.6064 | 1.217e-06 | 0.2679 | 20 | 0.01905 | 3.194e-06 | 15654 | 344388 | Yes |
| Substantia nigra | eQTLGen_Blood | ENSG00000166667 | ENSG00000166667 | 7 | 101991540 | rs10254446 | 7 | 102378009 | G | A | 0.2028 | -1.04 | 0.2184 | 2.1e-06 | -0.5067 | 0.02136 | 2.264e-124 | 2.053 | 0.4395 | 3.005e-06 | 0.07623 | 20 | 0.04704 | 3.194e-06 | 15654 | 344388 | Yes |
| CommonFactor | eQTLGen_Blood | DVL2 | ENSG00000004975 | 17 | 7133262 | rs2017365 | 17 | 7122624 | G | A | 0.3787 | -0.08585 | 0.01378 | 4.625e-10 | 0.1175 | 0.008241 | 3.689e-46 | -0.7304 | 0.1279 | 1.128e-08 | 0.6688 | 20 | 0.000162 | 3.482e-06 | 14360 | 315920 | Yes |
| CommonFactor | eQTLGen_Blood | STAB1 | ENSG00000010327 | 3 | 52543932 | rs758803 | 3 | 52515533 | T | C | 0.06064 | -0.153 | 0.02914 | 1.512e-07 | -0.7508 | 0.01787 | 0 | 0.2038 | 0.03912 | 1.883e-07 | 0.2017 | 19 | 0.002704 | 3.482e-06 | 14360 | 315920 | Yes |
| CommonFactor | eQTLGen_Blood | UNC13D | ENSG00000092929 | 17 | 73832052 | rs11077798 | 17 | 73953404 | T | C | 0.2376 | 0.08113 | 0.01605 | 4.296e-07 | -0.2221 | 0.009898 | 1.801e-111 | -0.3654 | 0.07408 | 8.151e-07 | 0.02589 | 18 | 0.0117 | 3.482e-06 | 14360 | 315920 | Yes |
| CommonFactor | eQTLGen_Blood | PYGB | ENSG00000100994 | 20 | 25253677 | rs6107017 | 20 | 25232300 | G | A | 0.4573 | 0.07327 | 0.01343 | 4.889e-08 | 0.7483 | 0.006756 | 0 | 0.09791 | 0.01797 | 5.074e-08 | 0.07838 | 20 | 0.0007286 | 3.482e-06 | 14360 | 315920 | Yes |
| CommonFactor | eQTLGen_Blood | CTSH | ENSG00000103811 | 15 | 79227658 | rs12592898 | 15 | 79229199 | A | G | 0.1292 | -0.1285 | 0.02007 | 1.521e-10 | -0.8096 | 0.01101 | 0 | 0.1588 | 0.02489 | 1.781e-10 | 0.3643 | 20 | 2.558e-06 | 3.482e-06 | 14360 | 315920 | Yes |
| CommonFactor | eQTLGen_Blood | TUBD1 | ENSG00000108423 | 17 | 57953577 | rs62081787 | 17 | 57885142 | A | G | 0.1501 | 0.1003 | 0.0185 | 5.898e-08 | -0.1202 | 0.0104 | 6.316e-31 | -0.8341 | 0.1699 | 9.152e-07 | 0.09646 | 20 | 0.01314 | 3.482e-06 | 14360 | 315920 | Yes |
| CommonFactor | eQTLGen_Blood | MLX | ENSG00000108788 | 17 | 40722171 | rs4793090 | 17 | 40686342 | G | A | 0.3231 | 0.1131 | 0.01415 | 1.323e-15 | 0.1323 | 0.008356 | 1.902e-56 | 0.8551 | 0.1198 | 9.698e-13 | 0.1794 | 20 | 1.393e-08 | 3.482e-06 | 14360 | 315920 | Yes |
| CommonFactor | eQTLGen_Blood | ARHGAP22 | ENSG00000128805 | 10 | 49759193 | rs1822861 | 10 | 49834326 | T | G | 0.497 | 0.06757 | 0.01342 | 4.802e-07 | 0.3592 | 0.007716 | 0 | 0.1881 | 0.03759 | 5.59e-07 | 0.01434 | 7 | 0.008027 | 3.482e-06 | 14360 | 315920 | Yes |
| CommonFactor | eQTLGen_Blood | PSMC3IP | ENSG00000131470 | 17 | 40727091 | rs4793090 | 17 | 40686342 | G | A | 0.3231 | 0.1131 | 0.01415 | 1.323e-15 | 0.07409 | 0.008451 | 1.835e-18 | 1.527 | 0.2585 | 3.497e-09 | 0.07189 | 20 | 5.021e-05 | 3.482e-06 | 14360 | 315920 | Yes |
| CommonFactor | eQTLGen_Blood | XPO4 | ENSG00000132953 | 13 | 21414328 | rs9509423 | 13 | 21467197 | C | T | 0.5169 | 0.06778 | 0.01342 | 4.435e-07 | -0.1386 | 0.008335 | 4.128e-62 | -0.489 | 0.1012 | 1.355e-06 | 0.3234 | 20 | 0.01946 | 3.482e-06 | 14360 | 315920 | Yes |
| CommonFactor | eQTLGen_Blood | TCF12 | ENSG00000140262 | 15 | 57401150 | rs2703593 | 15 | 57488767 | C | T | 0.2356 | -0.08962 | 0.01661 | 6.836e-08 | -0.1562 | 0.009516 | 1.601e-60 | 0.5738 | 0.112 | 2.967e-07 | 0.02665 | 20 | 0.004261 | 3.482e-06 | 14360 | 315920 | Yes |
| CommonFactor | eQTLGen_Blood | ACSL1 | ENSG00000151726 | 4 | 185712360 | rs2046814 | 4 | 185704176 | G | T | 0.4215 | -0.09018 | 0.01357 | 2.981e-11 | -0.06123 | 0.008055 | 2.948e-14 | 1.473 | 0.2944 | 5.623e-07 | 0.342 | 20 | 0.008074 | 3.482e-06 | 14360 | 315920 | Yes |
| CommonFactor | eQTLGen_Blood | SCNM1 | ENSG00000163156 | 1 | 151135956 | rs2925740 | 1 | 150969992 | T | C | 0.06561 | -0.1288 | 0.02575 | 5.65e-07 | 0.399 | 0.02942 | 6.709e-42 | -0.3228 | 0.06877 | 2.683e-06 | 0.05156 | 20 | 0.03853 | 3.482e-06 | 14360 | 315920 | Yes |
| CommonFactor | eQTLGen_Blood | UQCC5 | ENSG00000168273 | 3 | 52590641 | rs758800 | 3 | 52529266 | T | C | 0.06064 | -0.1522 | 0.02911 | 1.702e-07 | 0.3034 | 0.02214 | 9.661e-43 | -0.5017 | 0.1027 | 1.031e-06 | 0.2744 | 11 | 0.01481 | 3.482e-06 | 14360 | 315920 | Yes |
| CommonFactor | eQTLGen_Blood | ELP5 | ENSG00000170291 | 17 | 7158997 | rs2074219 | 17 | 7156447 | A | G | 0.3648 | -0.07811 | 0.01372 | 1.252e-08 | -0.2667 | 0.009075 | 8.871e-190 | 0.2929 | 0.05242 | 2.29e-08 | 0.1812 | 20 | 0.0003289 | 3.482e-06 | 14360 | 315920 | Yes |
| CommonFactor | eQTLGen_Blood | ARL17A | ENSG00000185829 | 17 | 44625578 | rs199454 | 17 | 44800110 | G | A | 0.2594 | 0.08741 | 0.01584 | 3.392e-08 | 0.2796 | 0.02057 | 4.335e-42 | 0.3127 | 0.06113 | 3.148e-07 | 0.0277 | 13 | 0.004521 | 3.482e-06 | 14360 | 315920 | Yes |
| CommonFactor | eQTLGen_Blood | DDX39B | ENSG00000198563 | 6 | 31504110 | rs2523500 | 6 | 31518354 | G | A | 0.3608 | 0.07506 | 0.01406 | 9.299e-08 | 0.2298 | 0.01692 | 5.32e-42 | 0.3266 | 0.06572 | 6.713e-07 | 0.2878 | 20 | 0.00964 | 3.482e-06 | 14360 | 315920 | Yes |
| CommonFactor | eQTLGen_Blood | VWA7 | ENSG00000204396 | 6 | 31739237 | rs2523500 | 6 | 31518354 | G | A | 0.3608 | 0.07506 | 0.01406 | 9.299e-08 | 0.1712 | 0.01701 | 8.249e-24 | 0.4385 | 0.09296 | 2.397e-06 | 0.0223 | 20 | 0.03442 | 3.482e-06 | 14360 | 315920 | Yes |
| CommonFactor | eQTLGen_Blood | HLA-E | ENSG00000204592 | 6 | 30459613 | rs1265094 | 6 | 31106893 | G | A | 0.5318 | -0.07037 | 0.01344 | 1.625e-07 | 0.1471 | 0.01251 | 6.562e-32 | -0.4783 | 0.09997 | 1.715e-06 | 0.2598 | 20 | 0.02462 | 3.482e-06 | 14360 | 315920 | Yes |
| CommonFactor | eQTLGen_Blood | KANSL1-AS1 | ENSG00000214401 | 17 | 44272515 | rs199513 | 17 | 44856932 | A | G | 0.2167 | 0.0912 | 0.0164 | 2.677e-08 | 1.235 | 0.01808 | 0 | 0.07384 | 0.01332 | 2.973e-08 | 0.6746 | 20 | 0.000427 | 3.482e-06 | 14360 | 315920 | Yes |
| CommonFactor | eQTLGen_Blood | ENSG00000225059 | ENSG00000225059 | 6 | 31352008 | rs2516396 | 6 | 31521913 | A | G | 0.2435 | 0.0818 | 0.0152 | 7.333e-08 | -0.4186 | 0.02946 | 8.08e-46 | -0.1954 | 0.03882 | 4.811e-07 | 0.2965 | 20 | 0.006908 | 3.482e-06 | 14360 | 315920 | Yes |
| CommonFactor | eQTLGen_Blood | LTA | ENSG00000226979 | 6 | 31540966 | rs2071590 | 6 | 31539768 | A | G | 0.3638 | 0.07443 | 0.01406 | 1.189e-07 | 0.1804 | 0.01255 | 7.891e-47 | 0.4127 | 0.08305 | 6.743e-07 | 0.08248 | 20 | 0.009683 | 3.482e-06 | 14360 | 315920 | Yes |
| CommonFactor | eQTLGen_Blood | HCG22 | ENSG00000228789 | 6 | 31024447 | rs1265054 | 6 | 31079643 | C | T | 0.5149 | -0.08033 | 0.01343 | 2.202e-09 | -0.4616 | 0.02 | 7.918e-118 | 0.174 | 0.03005 | 7.006e-09 | 0.05547 | 20 | 0.0001006 | 3.482e-06 | 14360 | 315920 | Yes |
| CommonFactor | eQTLGen_Blood | PANK2-AS1 | ENSG00000229539 | 20 | 3869200 | rs2300213 | 20 | 3986723 | T | C | 0.2684 | 0.07967 | 0.01562 | 3.405e-07 | 0.3494 | 0.02381 | 8.892e-49 | 0.228 | 0.04733 | 1.457e-06 | 0.01584 | 18 | 0.02092 | 3.482e-06 | 14360 | 315920 | Yes |
| CommonFactor | eQTLGen_Blood | HLA-B | ENSG00000234745 | 6 | 31323307 | rs2523605 | 6 | 31323409 | T | C | 0.169 | 0.07915 | 0.01655 | 1.729e-06 | 0.8445 | 0.02334 | 1.06e-286 | 0.09372 | 0.01977 | 2.121e-06 | 0.05929 | 20 | 0.03046 | 3.482e-06 | 14360 | 315920 | Yes |
| CommonFactor | eQTLGen_Blood | LRRC37A2 | ENSG00000238083 | 17 | 44610946 | rs199514 | 17 | 44856881 | G | A | 0.2187 | 0.0912 | 0.0164 | 2.678e-08 | 0.9179 | 0.01961 | 0 | 0.09936 | 0.01799 | 3.343e-08 | 0.3956 | 15 | 0.00048 | 3.482e-06 | 14360 | 315920 | Yes |
| CommonFactor | eQTLGen_Blood | ENSG00000261575 | ENSG00000261575 | 17 | 44345231 | rs199516 | 17 | 44856485 | C | T | 0.2207 | 0.09062 | 0.0164 | 3.296e-08 | 0.9001 | 0.02558 | 3.563e-271 | 0.1007 | 0.01845 | 4.814e-08 | 0.6102 | 20 | 0.0006913 | 3.482e-06 | 14360 | 315920 | Yes |
| CommonFactor | eQTLGen_Blood | MAPK8IP1P1 | ENSG00000262500 | 17 | 44321691 | rs199514 | 17 | 44856881 | G | A | 0.2187 | 0.0912 | 0.0164 | 2.678e-08 | 0.3483 | 0.01415 | 9.537e-134 | 0.2619 | 0.04827 | 5.81e-08 | 0.233 | 19 | 0.0008344 | 3.482e-06 | 14360 | 315920 | Yes |
| CommonFactor | eQTLGen_Blood | ENSG00000262539 | ENSG00000262539 | 17 | 44337444 | rs199514 | 17 | 44856881 | G | A | 0.2187 | 0.0912 | 0.0164 | 2.678e-08 | 0.5564 | 0.01338 | 0 | 0.1639 | 0.02974 | 3.544e-08 | 0.168 | 19 | 0.0005089 | 3.482e-06 | 14360 | 315920 | Yes |

| Trait | eQTL source | Gene symbol | Probe ID | Probe chromosome | Probe position (bp) | Top SNP | Top SNP chromosome | Top SNP position (bp) | Effect allele | Other allele | Effect allele frequency | Beta (GWAS) | SE (GWAS) | P (GWAS) | Beta (eQTL) | SE (eQTL) | P (eQTL) | Beta (SMR) | SE (SMR) | P (SMR) | P (HEIDI) | HEIDI SNPs (n) | Bonferroni-adjusted P (SMR) | Bonferroni threshold | Unique genes tested (n) | Tests performed (n) | Significant |
|---|---|---|---|---|---|---|---|---|---|---|---|---|---|---|---|---|---|---|---|---|---|---|---|---|---|---|---|
| Caudate nucleus | BrainMeta | AC092957.1 | ENSG00000243620.2 | 3 | 146857958 | rs17567518 | 3 | 147050225 | A | G | 0.2773 | -0.4444 | 0.07639 | 2.7e-10 | 0.6243 | 0.03398 | 2.292e-75 | -0.7118 | 0.1283 | 2.921e-08 | 0.2974 | 20 | 0.0004754 | 3.072e-06 | 16275 | 16284 | Yes |
| Caudate nucleus | BrainMeta | H2BC4 | ENSG00000180596.7 | 6 | 26119628 | rs80215559 | 6 | 25918225 | C | T | 0.04274 | 1.173 | 0.1247 | 2.7e-22 | -0.423 | 0.07171 | 3.664e-09 | -2.772 | 0.5547 | 5.83e-07 | 0.02137 | 18 | 0.009488 | 3.072e-06 | 16275 | 16284 | Yes |
| Caudate nucleus | BrainMeta | TYW1B | ENSG00000277149.5 | 7 | 72161254 | rs59973580 | 7 | 72236982 | A | G | 0.1938 | 0.3924 | 0.08275 | 2.3e-06 | 1.277 | 0.03461 | 3.073e-298 | 0.3072 | 0.06531 | 2.567e-06 | 0.6971 | 20 | 0.04177 | 3.072e-06 | 16275 | 16284 | Yes |
| Caudate nucleus | BrainMeta | STMN4 | ENSG00000015592.16 | 8 | 27104388 | rs17366947 | 8 | 27103024 | G | A | 0.3767 | 0.416 | 0.06749 | 7.7e-11 | -0.9061 | 0.02981 | 6.056e-203 | -0.459 | 0.076 | 1.539e-09 | 0.8347 | 20 | 2.505e-05 | 3.072e-06 | 16275 | 16284 | Yes |
| Caudate nucleus | BrainMeta | ACO1 | ENSG00000122729.19 | 9 | 32419684 | rs10970987 | 9 | 32457189 | C | T | 0.06859 | -0.8461 | 0.1209 | 5.9e-13 | 1.268 | 0.05872 | 2.225e-103 | -0.6674 | 0.1003 | 2.791e-11 | 0.1093 | 20 | 4.543e-07 | 3.072e-06 | 16275 | 16284 | Yes |
| Caudate nucleus | BrainMeta | SLC39A12-AS1 | ENSG00000226083.5 | 10 | 18295103 | rs11012783 | 10 | 18425418 | G | A | 0.4205 | 1.425 | 0.06783 | 1.6e-100 | 0.5626 | 0.03112 | 4.606e-73 | 2.534 | 0.1849 | 9.416e-43 | 0.3785 | 20 | 1.532e-38 | 3.072e-06 | 16275 | 16284 | Yes |
| Caudate nucleus | BrainMeta | CACNB2 | ENSG00000165995.22 | 10 | 18630920 | rs12357580 | 10 | 18450062 | T | C | 0.2823 | 1.282 | 0.07509 | 6.2e-68 | 0.222 | 0.03311 | 1.99e-11 | 5.772 | 0.9247 | 4.316e-10 | 0.04248 | 20 | 7.024e-06 | 3.072e-06 | 16275 | 16284 | Yes |
| Caudate nucleus | BrainMeta | VPS11 | ENSG00000160695.15 | 11 | 118945606 | rs592190 | 11 | 118955314 | A | G | 0.4712 | 0.3464 | 0.06654 | 1.4e-06 | 0.7859 | 0.02944 | 5.147e-157 | 0.4408 | 0.08626 | 3.228e-07 | 0.1544 | 20 | 0.005253 | 3.072e-06 | 16275 | 16284 | Yes |
| Caudate nucleus | BrainMeta | RITA1 | ENSG00000139405.16 | 12 | 113626752 | rs7135901 | 12 | 113705962 | G | T | 0.09443 | 0.7884 | 0.129 | 1.5e-10 | -1.135 | 0.06081 | 8.456e-78 | -0.6943 | 0.1195 | 6.273e-09 | 0.04885 | 20 | 0.0001021 | 3.072e-06 | 16275 | 16284 | Yes |
| Caudate nucleus | BrainMeta | HYKK | ENSG00000188266.14 | 15 | 78814810 | rs1504546 | 15 | 78824235 | T | C | 0.3708 | -0.4081 | 0.06617 | 1.4e-10 | -0.5726 | 0.02906 | 2.067e-86 | 0.7127 | 0.1211 | 3.967e-09 | 0.1113 | 20 | 6.457e-05 | 3.072e-06 | 16275 | 16284 | Yes |
| Caudate nucleus | BrainMeta | CHRNA5 | ENSG00000169684.13 | 15 | 78872736 | rs8053 | 15 | 78841220 | T | C | 0.3718 | -0.4046 | 0.06602 | 1.4e-10 | 1.06 | 0.02545 | 0 | -0.3819 | 0.06298 | 1.333e-09 | 0.05568 | 20 | 2.169e-05 | 3.072e-06 | 16275 | 16284 | Yes |
| Caudate nucleus | BrainMeta | CHRNA3 | ENSG00000080644.16 | 15 | 78899516 | rs12903129 | 15 | 78819606 | T | C | 0.3698 | -0.4146 | 0.06623 | 7e-11 | 0.5156 | 0.05376 | 8.713e-22 | -0.804 | 0.1534 | 1.588e-07 | 0.04213 | 20 | 0.002584 | 3.072e-06 | 16275 | 16284 | Yes |
| Caudate nucleus | BrainMeta | CTSH | ENSG00000103811.18 | 15 | 79227658 | rs2289702 | 15 | 79237293 | T | C | 0.1054 | -0.8119 | 0.1077 | 6.7e-16 | -0.7446 | 0.04959 | 5.941e-51 | 1.09 | 0.1619 | 1.636e-11 | 0.6241 | 20 | 2.663e-07 | 3.072e-06 | 16275 | 16284 | Yes |
| Dentate nucleus | BrainMeta | TMEM125 | ENSG00000179178.11 | 1 | 43737664 | rs7549876 | 1 | 43760236 | T | G | 0.3658 | 0.9854 | 0.1665 | 3.7e-10 | -0.3087 | 0.03108 | 3.031e-23 | -3.192 | 0.6277 | 3.674e-07 | 0.2538 | 20 | 0.00598 | 3.072e-06 | 16276 | 16285 | Yes |
| Dentate nucleus | BrainMeta | TIE1 | ENSG00000066056.14 | 1 | 43777716 | rs7549876 | 1 | 43760236 | T | G | 0.3658 | 0.9854 | 0.1665 | 3.7e-10 | -0.5411 | 0.03396 | 3.643e-57 | -1.821 | 0.3281 | 2.867e-08 | 0.4925 | 20 | 0.0004667 | 3.072e-06 | 16276 | 16285 | Yes |
| Dentate nucleus | BrainMeta | MED8 | ENSG00000159479.17 | 1 | 43852534 | rs11172 | 1 | 43850473 | G | A | 0.3738 | 0.8951 | 0.1614 | 2.6e-08 | 0.3835 | 0.02994 | 1.404e-37 | 2.334 | 0.4585 | 3.581e-07 | 0.1478 | 20 | 0.005829 | 3.072e-06 | 16276 | 16285 | Yes |
| Dentate nucleus | BrainMeta | STX6 | ENSG00000135823.14 | 1 | 180967052 | rs11586493 | 1 | 180961245 | G | A | 0.4314 | 0.8813 | 0.1595 | 6.6e-08 | 0.6704 | 0.03326 | 2.528e-90 | 1.315 | 0.2467 | 9.849e-08 | 0.2454 | 20 | 0.001603 | 3.072e-06 | 16276 | 16285 | Yes |
| Dentate nucleus | BrainMeta | RAB29 | ENSG00000117280.13 | 1 | 205740862 | rs823118 | 1 | 205723572 | C | T | 0.4652 | -1.067 | 0.158 | 5.9e-13 | -0.53 | 0.03361 | 5.178e-56 | 2.014 | 0.3243 | 5.345e-10 | 0.3983 | 20 | 8.7e-06 | 3.072e-06 | 16276 | 16285 | Yes |
| Dentate nucleus | BrainMeta | WDR75 | ENSG00000115368.10 | 2 | 190323225 | rs142714202 | 2 | 190360073 | G | A | 0.03479 | 3.357 | 0.4279 | 1.2e-16 | -0.9401 | 0.08842 | 2.115e-26 | -3.571 | 0.5657 | 2.743e-10 | 0.07664 | 20 | 4.465e-06 | 3.072e-06 | 16276 | 16285 | Yes |
| Dentate nucleus | BrainMeta | SLC40A1 | ENSG00000138449.11 | 2 | 190436900 | rs35623329 | 2 | 190443445 | C | T | 0.2893 | -1.061 | 0.1808 | 1e-09 | 0.6622 | 0.03671 | 9.788e-73 | -1.602 | 0.2871 | 2.382e-08 | 0.03846 | 20 | 0.0003877 | 3.072e-06 | 16276 | 16285 | Yes |
| Dentate nucleus | BrainMeta | SUMF1 | ENSG00000144455.14 | 3 | 4125728 | rs6762654 | 3 | 4405167 | T | C | 0.4105 | 0.8746 | 0.159 | 9.9e-08 | 0.4932 | 0.03521 | 1.396e-44 | 1.773 | 0.3463 | 3.033e-07 | 0.02037 | 20 | 0.004937 | 3.072e-06 | 16276 | 16285 | Yes |
| Dentate nucleus | BrainMeta | AC023483.1 | ENSG00000286579.1 | 3 | 4332008 | rs11915920 | 3 | 4410534 | T | C | 0.506 | -1.071 | 0.1572 | 1.7e-11 | 0.3462 | 0.0285 | 5.784e-34 | -3.094 | 0.5206 | 2.801e-09 | 0.01514 | 20 | 4.559e-05 | 3.072e-06 | 16276 | 16285 | Yes |
| Dentate nucleus | BrainMeta | KIAA1109 | ENSG00000138688.16 | 4 | 123178700 | rs56313700 | 4 | 123287573 | A | C | 0.2396 | -1.187 | 0.1919 | 1.4e-09 | -0.6283 | 0.03719 | 4.853e-64 | 1.889 | 0.3252 | 6.334e-09 | 0.02737 | 20 | 0.0001031 | 3.072e-06 | 16276 | 16285 | Yes |
| Dentate nucleus | BrainMeta | H2BC4 | ENSG00000180596.7 | 6 | 26119628 | rs80215559 | 6 | 25918225 | C | T | 0.04274 | 6.174 | 0.299 | 6.8e-104 | -0.423 | 0.07171 | 3.664e-09 | -14.6 | 2.573 | 1.413e-08 | 0.1117 | 18 | 0.0002299 | 3.072e-06 | 16276 | 16285 | Yes |
| Dentate nucleus | BrainMeta | BTN3A2 | ENSG00000186470.14 | 6 | 26371968 | rs71557332 | 6 | 26356853 | T | C | 0.1123 | -1.205 | 0.2309 | 8.8e-09 | -1.311 | 0.07194 | 3.694e-74 | 0.9196 | 0.1833 | 5.222e-07 | 0.01202 | 20 | 0.008499 | 3.072e-06 | 16276 | 16285 | Yes |
| Dentate nucleus | BrainMeta | HLA-H | ENSG00000206341.7 | 6 | 29856894 | rs34302671 | 6 | 28454795 | G | A | 0.09742 | 2.268 | 0.2566 | 1.9e-20 | -3.25 | 0.1252 | 1.182e-148 | -0.6976 | 0.0834 | 6.032e-17 | 0.02555 | 20 | 9.818e-13 | 3.072e-06 | 16276 | 16285 | Yes |
| Dentate nucleus | BrainMeta | RUNX2 | ENSG00000124813.23 | 6 | 45463990 | rs2790100 | 6 | 45430336 | A | C | 0.334 | -1.012 | 0.167 | 1.3e-10 | -0.378 | 0.0314 | 2.24e-33 | 2.678 | 0.4946 | 6.121e-08 | 0.1214 | 20 | 0.0009963 | 3.072e-06 | 16276 | 16285 | Yes |
| Dentate nucleus | BrainMeta | SNAP91 | ENSG00000065609.14 | 6 | 84341004 | rs2224195 | 6 | 84307726 | C | T | 0.4582 | -0.8125 | 0.1584 | 2.7e-07 | -0.4207 | 0.03401 | 3.841e-35 | 1.931 | 0.4075 | 2.146e-06 | 0.1919 | 20 | 0.03493 | 3.072e-06 | 16276 | 16285 | Yes |
| Dentate nucleus | BrainMeta | OSTM1 | ENSG00000081087.15 | 6 | 108424836 | rs6940713 | 6 | 108392809 | G | A | 0.2843 | 1.303 | 0.1743 | 7.3e-14 | 0.5757 | 0.03422 | 1.724e-63 | 2.263 | 0.3313 | 8.514e-12 | 0.2643 | 20 | 1.386e-07 | 3.072e-06 | 16276 | 16285 | Yes |
| Dentate nucleus | BrainMeta | SLC25A37 | ENSG00000147454.14 | 8 | 23409722 | rs4872141 | 8 | 23384559 | A | G | 0.4523 | -1.181 | 0.1593 | 2.2e-14 | -0.3216 | 0.02763 | 2.657e-31 | 3.674 | 0.5875 | 4.036e-10 | 0.5764 | 20 | 6.568e-06 | 3.072e-06 | 16276 | 16285 | Yes |
| Dentate nucleus | BrainMeta | ADAM7 | ENSG00000069206.15 | 8 | 24341463 | rs1457266 | 8 | 24769852 | A | G | 0.3529 | -0.974 | 0.1696 | 9.1e-10 | 0.4055 | 0.04975 | 3.574e-16 | -2.402 | 0.5116 | 2.666e-06 | 0.336 | 20 | 0.04339 | 3.072e-06 | 16276 | 16285 | Yes |
| Dentate nucleus | BrainMeta | STMN4 | ENSG00000015592.16 | 8 | 27104388 | rs17366947 | 8 | 27103024 | G | A | 0.3767 | 0.8653 | 0.1617 | 3.3e-08 | -0.9061 | 0.02981 | 6.056e-203 | -0.955 | 0.1812 | 1.364e-07 | 0.4187 | 20 | 0.00222 | 3.072e-06 | 16276 | 16285 | Yes |
| Dentate nucleus | BrainMeta | ASAP1 | ENSG00000153317.15 | 8 | 131260135 | rs62524625 | 8 | 131084728 | T | C | 0.3101 | -0.9582 | 0.1745 | 2.8e-09 | 0.4166 | 0.03814 | 8.937e-28 | -2.3 | 0.4687 | 9.245e-07 | 0.02493 | 20 | 0.01505 | 3.072e-06 | 16276 | 16285 | Yes |
| Dentate nucleus | BrainMeta | INA | ENSG00000148798.11 | 10 | 105043499 | rs4293062 | 10 | 105052624 | A | G | 0.4841 | -0.994 | 0.1583 | 7.2e-10 | 0.6211 | 0.02935 | 2.286e-99 | -1.6 | 0.2659 | 1.767e-09 | 0.02079 | 20 | 2.876e-05 | 3.072e-06 | 16276 | 16285 | Yes |
| Dentate nucleus | BrainMeta | HRAS | ENSG00000174775.17 | 11 | 534764 | rs35333170 | 11 | 526478 | G | A | 0.0994 | -1.733 | 0.2341 | 3.2e-13 | -0.7617 | 0.04546 | 5.187e-63 | 2.275 | 0.336 | 1.272e-11 | 0.1571 | 20 | 2.07e-07 | 3.072e-06 | 16276 | 16285 | Yes |
| Dentate nucleus | BrainMeta | CD82 | ENSG00000085117.12 | 11 | 44613942 | rs2303865 | 11 | 44636833 | A | G | 0.1322 | -1.401 | 0.2332 | 4.9e-10 | 0.8945 | 0.04141 | 1.798e-103 | -1.567 | 0.2706 | 7.064e-09 | 0.05007 | 13 | 0.000115 | 3.072e-06 | 16276 | 16285 | Yes |
| Dentate nucleus | BrainMeta | TTC9C | ENSG00000162222.14 | 11 | 62501653 | rs10897290 | 11 | 62451681 | T | C | 0.2097 | -1.077 | 0.1824 | 5.2e-09 | 0.4218 | 0.03606 | 1.326e-31 | -2.553 | 0.4844 | 1.368e-07 | 0.4127 | 20 | 0.002227 | 3.072e-06 | 16276 | 16285 | Yes |
| Dentate nucleus | BrainMeta | DDHD1 | ENSG00000100523.16 | 14 | 53561752 | rs8015438 | 14 | 53602160 | G | T | 0.1511 | 1.519 | 0.2202 | 6.8e-13 | -0.6512 | 0.04054 | 4.71e-58 | -2.333 | 0.368 | 2.299e-10 | 0.3472 | 20 | 3.742e-06 | 3.072e-06 | 16276 | 16285 | Yes |
| Dentate nucleus | BrainMeta | TTLL5 | ENSG00000119685.20 | 14 | 76260695 | rs11623813 | 14 | 76138194 | C | T | 0.33 | -0.9747 | 0.1666 | 3.4e-10 | -0.5639 | 0.03262 | 6.155e-67 | 1.729 | 0.312 | 3.016e-08 | 0.3737 | 20 | 0.0004909 | 3.072e-06 | 16276 | 16285 | Yes |
| Dentate nucleus | BrainMeta | OTUD7A | ENSG00000169918.10 | 15 | 31965296 | rs4779931 | 15 | 32156572 | G | A | 0.4771 | 1.008 | 0.158 | 3.1e-11 | 0.6405 | 0.02997 | 2.321e-101 | 1.574 | 0.2575 | 9.903e-10 | 0.1162 | 20 | 1.612e-05 | 3.072e-06 | 16276 | 16285 | Yes |
| Dentate nucleus | BrainMeta | COX6B1 | ENSG00000126267.11 | 19 | 36144454 | rs61218414 | 19 | 36171716 | T | C | 0.5 | -0.9133 | 0.1576 | 4.5e-09 | 0.6705 | 0.02962 | 1.93e-113 | -1.362 | 0.2426 | 1.986e-08 | 0.02817 | 20 | 0.0003233 | 3.072e-06 | 16276 | 16285 | Yes |
| Dentate nucleus | BrainMeta | ZBTB32 | ENSG00000011590.14 | 19 | 36201700 | rs107068 | 19 | 36204690 | G | A | 0.4791 | 1.108 | 0.1579 | 6.4e-13 | 0.5656 | 0.0397 | 4.694e-46 | 1.958 | 0.3112 | 3.122e-10 | 0.5725 | 20 | 5.081e-06 | 3.072e-06 | 16276 | 16285 | Yes |
| Dentate nucleus | BrainMeta | KMT2B | ENSG00000272333.7 | 19 | 36219327 | rs231591 | 19 | 36224705 | G | A | 0.4811 | 1.098 | 0.1579 | 1.1e-12 | 0.2102 | 0.02967 | 1.39e-12 | 5.224 | 1.053 | 6.937e-07 | 0.8153 | 20 | 0.01129 | 3.072e-06 | 16276 | 16285 | Yes |
| Dentate nucleus | BrainMeta | ZNF160 | ENSG00000170949.17 | 19 | 53588274 | rs12985137 | 19 | 53617818 | C | T | 0.4205 | 0.8359 | 0.1585 | 3.6e-08 | -0.5373 | 0.02701 | 4.791e-88 | -1.556 | 0.3053 | 3.456e-07 | 0.2946 | 20 | 0.005624 | 3.072e-06 | 16276 | 16285 | Yes |
| Dentate nucleus | BrainMeta | ZNF347 | ENSG00000197937.13 | 19 | 53644826 | rs12985137 | 19 | 53617818 | C | T | 0.4205 | 0.8359 | 0.1585 | 3.6e-08 | -0.5056 | 0.03015 | 4.14e-63 | -1.654 | 0.3287 | 4.906e-07 | 0.2358 | 20 | 0.007985 | 3.072e-06 | 16276 | 16285 | Yes |

| Trait | eQTL source | Gene symbol | Probe ID | Probe chromosome | Probe position (bp) | Top SNP | Top SNP chromosome | Top SNP position (bp) | Effect allele | Other allele | Effect allele frequency | Beta (GWAS) | SE (GWAS) | P (GWAS) | Beta (eQTL) | SE (eQTL) | P (eQTL) | Beta (SMR) | SE (SMR) | P (SMR) | P (HEIDI) | HEIDI SNPs (n) | Bonferroni-adjusted P (SMR) | Bonferroni threshold | Unique genes tested (n) | Tests performed (n) | Significant |
|---|---|---|---|---|---|---|---|---|---|---|---|---|---|---|---|---|---|---|---|---|---|---|---|---|---|---|---|
| Dentate nucleus | BrainMeta | GINS1 | ENSG00000101003.10 | 20 | 25410786 | rs6083809 | 20 | 25329309 | A | G | 0.4374 | 1.059 | 0.1586 | 5.2e-14 | 0.348 | 0.03156 | 2.842e-28 | 3.042 | 0.5328 | 1.134e-08 | 0.1556 | 20 | 0.0001845 | 3.072e-06 | 16276 | 16285 | Yes |
| Dentate nucleus | BrainMeta | AL450124.1 | ENSG00000204556.4 | 20 | 26000394 | rs6050936 | 20 | 25834073 | A | G | 0.3201 | 0.9436 | 0.1712 | 7.6e-10 | -0.5441 | 0.04858 | 4.058e-29 | -1.734 | 0.3507 | 7.603e-07 | 0.02327 | 20 | 0.01237 | 3.072e-06 | 16276 | 16285 | Yes |
| Putamen | BrainMeta | LINC02278 | ENSG00000251635.2 | 4 | 38569634 | rs4833079 | 4 | 38654681 | C | T | 0.3559 | 0.863 | 0.1142 | 9.4e-14 | -0.3647 | 0.03503 | 2.217e-25 | -2.366 | 0.3868 | 9.514e-10 | 0.08591 | 20 | 1.548e-05 | 3.072e-06 | 16275 | 16284 | Yes |
| Putamen | BrainMeta | H2BC4 | ENSG00000180596.7 | 6 | 26119628 | rs80215559 | 6 | 25918225 | C | T | 0.04274 | 3.605 | 0.2139 | 5.4e-68 | -0.423 | 0.07171 | 3.664e-09 | -8.521 | 1.531 | 2.583e-08 | 0.02427 | 18 | 0.0004205 | 3.072e-06 | 16275 | 16284 | Yes |
| Putamen | BrainMeta | HLA-H | ENSG00000206341.7 | 6 | 29856894 | rs34302671 | 6 | 28454795 | G | A | 0.09742 | 1.254 | 0.1836 | 2.6e-12 | -3.25 | 0.1252 | 1.182e-148 | -0.3858 | 0.0584 | 3.971e-11 | 0.2136 | 20 | 6.462e-07 | 3.072e-06 | 16275 | 16284 | Yes |
| Putamen | BrainMeta | RUNX2 | ENSG00000124813.23 | 6 | 45463990 | rs2790100 | 6 | 45430336 | A | C | 0.334 | -0.9276 | 0.1194 | 1.3e-17 | -0.378 | 0.0314 | 2.24e-33 | 2.454 | 0.376 | 6.755e-11 | 0.0818 | 20 | 1.099e-06 | 3.072e-06 | 16275 | 16284 | Yes |
| Putamen | BrainMeta | AC004130.2 | ENSG00000271133.5 | 7 | 20369646 | rs3807933 | 7 | 20383123 | T | C | 0.05865 | -1.474 | 0.2347 | 1.2e-10 | -1.157 | 0.05537 | 6.926e-97 | 1.275 | 0.2119 | 1.779e-09 | 0.3545 | 20 | 2.896e-05 | 3.072e-06 | 16275 | 16284 | Yes |
| Putamen | BrainMeta | SLC25A37 | ENSG00000147454.14 | 8 | 23409722 | rs4872141 | 8 | 23384559 | A | G | 0.4523 | -0.8636 | 0.114 | 1.5e-15 | -0.3216 | 0.02763 | 2.657e-31 | 2.686 | 0.423 | 2.16e-10 | 0.1078 | 20 | 3.516e-06 | 3.072e-06 | 16275 | 16284 | Yes |
| Putamen | BrainMeta | STMN4 | ENSG00000015592.16 | 8 | 27104388 | rs17366947 | 8 | 27103024 | G | A | 0.3767 | 0.8356 | 0.1157 | 1.5e-12 | -0.9061 | 0.02981 | 6.056e-203 | -0.9221 | 0.1312 | 2.104e-12 | 0.2198 | 20 | 3.424e-08 | 3.072e-06 | 16275 | 16284 | Yes |
| Putamen | BrainMeta | AC022509.5 | ENSG00000286752.1 | 12 | 26306428 | rs7136036 | 12 | 26327295 | A | G | 0.4453 | -0.5521 | 0.1124 | 2.1e-06 | -0.7211 | 0.0424 | 7.098e-65 | 0.7657 | 0.1622 | 2.346e-06 | 0.02571 | 20 | 0.03817 | 3.072e-06 | 16275 | 16284 | Yes |
| Putamen | BrainMeta | RITA1 | ENSG00000139405.16 | 12 | 113626752 | rs7135901 | 12 | 113705962 | G | T | 0.09443 | 1.798 | 0.2211 | 5.4e-16 | -1.135 | 0.06081 | 8.456e-78 | -1.584 | 0.2124 | 8.869e-14 | 0.5532 | 20 | 1.443e-09 | 3.072e-06 | 16275 | 16284 | Yes |
| Putamen | BrainMeta | CHRNA5 | ENSG00000169684.13 | 15 | 78872736 | rs8053 | 15 | 78841220 | T | C | 0.3718 | -0.7393 | 0.1132 | 2.2e-11 | 1.06 | 0.02545 | 0 | -0.6978 | 0.1082 | 1.105e-10 | 0.01309 | 20 | 1.798e-06 | 3.072e-06 | 16275 | 16284 | Yes |
| Putamen | BrainMeta | CTSH | ENSG00000103811.18 | 15 | 79227658 | rs2289702 | 15 | 79237293 | T | C | 0.1054 | -1.843 | 0.1847 | 2.4e-25 | -0.7446 | 0.04959 | 5.941e-51 | 2.476 | 0.2979 | 9.558e-17 | 0.4714 | 20 | 1.556e-12 | 3.072e-06 | 16275 | 16284 | Yes |
| Putamen | BrainMeta | MAF | ENSG00000178573.7 | 16 | 79627187 | rs250167 | 16 | 79596365 | C | T | 0.3569 | 1.199 | 0.1155 | 1.6e-26 | 0.1711 | 0.0302 | 1.464e-08 | 7.007 | 1.409 | 6.578e-07 | 0.1052 | 19 | 0.01071 | 3.072e-06 | 16275 | 16284 | Yes |
| Putamen | BrainMeta | TUBB8P7 | ENSG00000261812.6 | 16 | 90161162 | rs138169571 | 16 | 90162340 | T | C | 0.07952 | 1.038 | 0.2092 | 2.8e-07 | 1.048 | 0.06126 | 1.168e-65 | 0.9902 | 0.2078 | 1.878e-06 | 0.2407 | 20 | 0.03056 | 3.072e-06 | 16275 | 16284 | Yes |
| Putamen | BrainMeta | SLC2A4 | ENSG00000181856.15 | 17 | 7188306 | rs5418 | 17 | 7185092 | G | A | 0.4314 | -0.5897 | 0.114 | 5e-07 | -0.4373 | 0.02756 | 1.095e-56 | 1.348 | 0.2742 | 8.815e-07 | 0.2821 | 20 | 0.01435 | 3.072e-06 | 16275 | 16284 | Yes |
| Putamen | BrainMeta | PLEKHM1 | ENSG00000225190.11 | 17 | 43540690 | rs55746869 | 17 | 43551083 | T | C | 0.1958 | 1.469 | 0.1461 | 3.8e-26 | 0.3923 | 0.05379 | 3.014e-13 | 3.745 | 0.6343 | 3.555e-09 | 0.01227 | 12 | 5.785e-05 | 3.072e-06 | 16275 | 16284 | Yes |
| Putamen | BrainMeta | LRRC37A4P | ENSG00000214425.8 | 17 | 43606505 | rs17577975 | 17 | 44210442 | T | C | 0.2396 | 1.447 | 0.1359 | 3.5e-29 | -1.301 | 0.03932 | 5.428e-240 | -1.112 | 0.1097 | 3.779e-24 | | | 6.151e-20 | 3.072e-06 | 16275 | 16284 | Yes |
| Putamen | BrainMeta | AC091132.3 | ENSG00000266918.1 | 17 | 43610074 | rs17689918 | 17 | 43910088 | A | G | 0.2396 | 1.425 | 0.1357 | 9.6e-29 | 1.055 | 0.06109 | 7.244e-67 | 1.35 | 0.1505 | 2.85e-19 | | | 4.638e-15 | 3.072e-06 | 16275 | 16284 | Yes |
| Putamen | BrainMeta | LINC02210 | ENSG00000204650.14 | 17 | 43715108 | rs17660337 | 17 | 44169605 | C | T | 0.2406 | 1.434 | 0.1356 | 6.9e-29 | 1.155 | 0.04562 | 1.667e-141 | 1.241 | 0.1272 | 1.666e-22 | | | 2.712e-18 | 3.072e-06 | 16275 | 16284 | Yes |
| Putamen | BrainMeta | AC005829.2 | ENSG00000262539.1 | 17 | 44337444 | rs75257416 | 17 | 44349079 | A | C | 0.2008 | 1.504 | 0.1509 | 1.5e-26 | 1.43 | 0.05978 | 2.202e-126 | 1.052 | 0.1143 | 3.595e-20 | 0.0248 | 6 | 5.851e-16 | 3.072e-06 | 16275 | 16284 | Yes |
| Globus pallidus | BrainMeta | DBI | ENSG00000155368.16 | 2 | 120127316 | rs956309 | 2 | 120128482 | T | C | 0.1998 | -1.052 | 0.2021 | 1.9e-08 | 0.4826 | 0.0382 | 1.372e-36 | -2.18 | 0.4529 | 1.488e-06 | 0.01131 | 20 | 0.02422 | 3.072e-06 | 16276 | 16285 | Yes |
| Globus pallidus | BrainMeta | WDR75 | ENSG00000115368.10 | 2 | 190323225 | rs142714202 | 2 | 190360073 | G | A | 0.03479 | 6.603 | 0.4498 | 4.6e-52 | -0.9401 | 0.08842 | 2.115e-26 | -7.024 | 0.8157 | 7.251e-18 | 0.02453 | 20 | 1.18e-13 | 3.072e-06 | 16276 | 16285 | Yes |
| Globus pallidus | BrainMeta | AC092957.1 | ENSG00000243620.2 | 3 | 146857958 | rs17567518 | 3 | 147050225 | A | G | 0.2773 | -1.023 | 0.1922 | 4.5e-09 | 0.6243 | 0.03398 | 2.292e-75 | -1.639 | 0.3205 | 3.157e-07 | 0.2812 | 20 | 0.005138 | 3.072e-06 | 16276 | 16285 | Yes |
| Globus pallidus | BrainMeta | SMBD1P | ENSG00000283426.1 | 3 | 195433502 | rs6804822 | 3 | 195709662 | A | G | 0.2455 | -1.214 | 0.2008 | 9.5e-09 | 0.7998 | 0.03679 | 8.516e-105 | -1.518 | 0.2605 | 5.671e-09 | 0.1862 | 20 | 9.23e-05 | 3.072e-06 | 16276 | 16285 | Yes |
| Globus pallidus | BrainMeta | MUC20 | ENSG00000176945.17 | 3 | 195457874 | rs6804822 | 3 | 195709662 | A | G | 0.2455 | -1.214 | 0.2008 | 9.5e-09 | 0.859 | 0.03186 | 4.53e-160 | -1.414 | 0.2396 | 3.619e-09 | 0.09513 | 20 | 5.89e-05 | 3.072e-06 | 16276 | 16285 | Yes |
| Globus pallidus | BrainMeta | AC024937.2 | ENSG00000231464.1 | 3 | 195724356 | rs7614767 | 3 | 195753451 | G | A | 0.3419 | 1.208 | 0.1759 | 2.1e-12 | -0.4971 | 0.05976 | 8.88e-17 | -2.43 | 0.4588 | 1.188e-07 | 0.1877 | 20 | 0.001933 | 3.072e-06 | 16276 | 16285 | Yes |
| Globus pallidus | BrainMeta | LINC02278 | ENSG00000251635.2 | 4 | 38569634 | rs4833079 | 4 | 38654681 | C | T | 0.3559 | 0.9343 | 0.1674 | 2.1e-07 | -0.3647 | 0.03503 | 2.217e-25 | -2.562 | 0.5208 | 8.726e-07 | 0.2055 | 20 | 0.0142 | 3.072e-06 | 16276 | 16285 | Yes |
| Globus pallidus | BrainMeta | H2BC4 | ENSG00000180596.7 | 6 | 26119628 | rs80215559 | 6 | 25918225 | C | T | 0.04274 | 2.814 | 0.314 | 6.4e-20 | -0.423 | 0.07171 | 3.664e-09 | -6.652 | 1.35 | 8.335e-07 | 0.02349 | 18 | 0.01357 | 3.072e-06 | 16276 | 16285 | Yes |
| Globus pallidus | BrainMeta | HCG22 | ENSG00000228789.8 | 6 | 31024447 | rs2517534 | 6 | 31017334 | A | G | 0.4264 | 1.031 | 0.1667 | 1.2e-11 | -0.7991 | 0.03955 | 8.905e-91 | -1.29 | 0.2182 | 3.372e-09 | 0.03793 | 20 | 5.488e-05 | 3.072e-06 | 16276 | 16285 | Yes |
| Globus pallidus | BrainMeta | MICB-DT | ENSG00000286940.1 | 6 | 31455231 | rs2534676 | 6 | 31464029 | C | T | 0.4155 | 0.9716 | 0.1696 | 2.8e-10 | 0.327 | 0.03682 | 6.556e-19 | 2.971 | 0.617 | 1.476e-06 | 0.03908 | 20 | 0.02402 | 3.072e-06 | 16276 | 16285 | Yes |
| Globus pallidus | BrainMeta | SND1 | ENSG00000197157.11 | 7 | 127512454 | rs9649027 | 7 | 127354818 | C | T | 0.2803 | -0.9331 | 0.1816 | 1.9e-07 | -0.4781 | 0.03949 | 9.702e-34 | 1.952 | 0.4126 | 2.238e-06 | 0.6954 | 20 | 0.03642 | 3.072e-06 | 16276 | 16285 | Yes |
| Globus pallidus | BrainMeta | SLC20A2 | ENSG00000168575.10 | 8 | 42335531 | rs2923405 | 8 | 42448126 | T | G | 0.4205 | -0.9281 | 0.1684 | 8.2e-09 | 0.3258 | 0.02529 | 5.588e-38 | -2.849 | 0.5622 | 4.041e-07 | 0.7163 | 20 | 0.006578 | 3.072e-06 | 16276 | 16285 | Yes |
| Globus pallidus | BrainMeta | SMIM19 | ENSG00000176209.12 | 8 | 42403318 | rs2974355 | 8 | 42414117 | C | T | 0.3877 | -0.8965 | 0.1699 | 2.9e-08 | 0.9442 | 0.03062 | 8.959e-209 | -0.9495 | 0.1826 | 1.982e-07 | 0.597 | 20 | 0.003225 | 3.072e-06 | 16276 | 16285 | Yes |
| Globus pallidus | BrainMeta | SNX31 | ENSG00000174226.9 | 8 | 101630380 | rs1693567 | 8 | 101677386 | C | T | 0.4672 | -1.892 | 0.1664 | 1.4e-33 | 0.7291 | 0.02919 | 1.17e-137 | -2.595 | 0.2508 | 4.351e-25 | 0.8826 | 20 | 7.082e-21 | 3.072e-06 | 16276 | 16285 | Yes |
| Globus pallidus | BrainMeta | PABPC1 | ENSG00000070756.17 | 8 | 101716540 | rs1786345 | 8 | 101674751 | C | A | 0.4414 | -1.824 | 0.1676 | 3.4e-31 | 0.3413 | 0.02869 | 1.243e-32 | -5.346 | 0.6656 | 9.588e-16 | 0.4348 | 20 | 1.561e-11 | 3.072e-06 | 16276 | 16285 | Yes |
| Globus pallidus | BrainMeta | CTSH | ENSG00000103811.18 | 15 | 79227658 | rs2289702 | 15 | 79237293 | T | C | 0.1054 | -1.703 | 0.2714 | 7e-11 | -0.7446 | 0.04959 | 5.941e-51 | 2.287 | 0.3951 | 7.065e-09 | 0.6061 | 20 | 0.000115 | 3.072e-06 | 16276 | 16285 | Yes |
| Globus pallidus | BrainMeta | TUBB8P7 | ENSG00000261812.6 | 16 | 90161162 | rs138169571 | 16 | 90162340 | T | C | 0.07952 | 1.493 | 0.3072 | 2.1e-07 | 1.048 | 0.06126 | 1.168e-65 | 1.424 | 0.3046 | 2.954e-06 | 0.3319 | 20 | 0.04808 | 3.072e-06 | 16276 | 16285 | Yes |
| Red nucleus | BrainMeta | WDR75 | ENSG00000115368.10 | 2 | 190323225 | rs142714202 | 2 | 190360073 | G | A | 0.03479 | 4.696 | 0.4019 | 4.6e-34 | -0.9401 | 0.08842 | 2.115e-26 | -4.995 | 0.6352 | 3.732e-15 | 0.2095 | 20 | 6.074e-11 | 3.072e-06 | 16275 | 16284 | Yes |
| Red nucleus | BrainMeta | SMBD1P | ENSG00000283426.1 | 3 | 195433502 | rs6804822 | 3 | 195709662 | A | G | 0.2455 | -0.9555 | 0.1796 | 3.7e-07 | 0.7998 | 0.03679 | 8.516e-105 | -1.195 | 0.2311 | 2.361e-07 | 0.02333 | 20 | 0.003843 | 3.072e-06 | 16275 | 16284 | Yes |
| Red nucleus | BrainMeta | MUC20 | ENSG00000176945.17 | 3 | 195457874 | rs6804822 | 3 | 195709662 | A | G | 0.2455 | -0.9555 | 0.1796 | 3.7e-07 | 0.859 | 0.03186 | 4.53e-160 | -1.112 | 0.2131 | 1.787e-07 | 0.03453 | 20 | 0.002908 | 3.072e-06 | 16275 | 16284 | Yes |
| Red nucleus | BrainMeta | LINC02278 | ENSG00000251635.2 | 4 | 38569634 | rs4833079 | 4 | 38654681 | C | T | 0.3559 | 1.075 | 0.1498 | 6.7e-12 | -0.3647 | 0.03503 | 2.217e-25 | -2.946 | 0.4987 | 3.473e-09 | 0.04501 | 20 | 5.652e-05 | 3.072e-06 | 16275 | 16284 | Yes |
| Red nucleus | BrainMeta | PIWIL2 | ENSG00000197181.12 | 8 | 22173955 | rs4556088 | 8 | 22198981 | A | G | 0.4304 | -0.8311 | 0.1482 | 7.8e-09 | 0.6101 | 0.0271 | 2.908e-112 | -1.362 | 0.2503 | 5.235e-08 | 0.1289 | 20 | 0.000852 | 3.072e-06 | 16275 | 16284 | Yes |
| Red nucleus | BrainMeta | SLC25A37 | ENSG00000147454.14 | 8 | 23409722 | rs4872141 | 8 | 23384559 | A | G | 0.4523 | -1.016 | 0.1495 | 1.9e-12 | -0.3216 | 0.02763 | 2.657e-31 | 3.16 | 0.5384 | 4.38e-09 | 0.254 | 20 | 7.129e-05 | 3.072e-06 | 16275 | 16284 | Yes |
| Red nucleus | BrainMeta | SNX31 | ENSG00000174226.9 | 8 | 101630380 | rs1693567 | 8 | 101677386 | C | T | 0.4672 | -1.391 | 0.1487 | 2.6e-24 | 0.7291 | 0.02919 | 1.17e-137 | -1.907 | 0.2177 | 1.951e-18 | 0.5393 | 20 | 3.176e-14 | 3.072e-06 | 16275 | 16284 | Yes |

| Trait | eQTL source | Gene symbol | Probe ID | Probe chromosome | Probe position (bp) | Top SNP | Top SNP chromosome | Top SNP position (bp) | Effect allele | Other allele | Effect allele frequency | Beta (GWAS) | SE (GWAS) | P (GWAS) | Beta (eQTL) | SE (eQTL) | P (eQTL) | Beta (SMR) | SE (SMR) | P (SMR) | P (HEIDI) | HEIDI SNPs (n) | Bonferroni-adjusted P (SMR) | Bonferroni threshold | Unique genes tested (n) | Tests performed (n) | Significant |
|---|---|---|---|---|---|---|---|---|---|---|---|---|---|---|---|---|---|---|---|---|---|---|---|---|---|---|---|
| Red nucleus | BrainMeta | PABPC1 | ENSG00000070756.17 | 8 | 101716540 | rs1786345 | 8 | 101674751 | C | A | 0.4414 | -1.322 | 0.1497 | 4.6e-22 | 0.3413 | 0.02869 | 1.243e-32 | -3.874 | 0.5462 | 1.318e-12 | 0.1629 | 20 | 2.144e-08 | 3.072e-06 | 16275 | 16284 | Yes |
| Red nucleus | BrainMeta | SPAAR | ENSG00000235387.5 | 9 | 35923318 | rs10972618 | 9 | 35908313 | C | A | 0.3211 | 0.8386 | 0.1601 | 1.7e-07 | -0.3291 | 0.03127 | 6.508e-26 | -2.548 | 0.5432 | 2.733e-06 | 0.1431 | 20 | 0.04448 | 3.072e-06 | 16275 | 16284 | Yes |
| Red nucleus | BrainMeta | LINC01505 | ENSG00000234323.8 | 9 | 109129374 | rs145825805 | 9 | 109099744 | A | G | 0.0835 | 2.232 | 0.2885 | 4.1e-14 | 0.4886 | 0.06421 | 2.772e-14 | 4.568 | 0.842 | 5.804e-08 | 0.5946 | 15 | 0.0009446 | 3.072e-06 | 16275 | 16284 | Yes |
| Red nucleus | BrainMeta | SLC39A12 | ENSG00000148482.12 | 10 | 18286494 | rs690759 | 10 | 18243703 | A | G | 0.3708 | -1.841 | 0.1539 | 1e-34 | -0.3751 | 0.03032 | 3.75e-35 | 4.909 | 0.5707 | 7.885e-18 | 0.0218 | 20 | 1.283e-13 | 3.072e-06 | 16275 | 16284 | Yes |
| Red nucleus | BrainMeta | INA | ENSG00000148798.11 | 10 | 105043499 | rs4293062 | 10 | 105052624 | A | G | 0.4841 | -0.9314 | 0.1487 | 1.1e-10 | 0.6211 | 0.02935 | 2.286e-99 | -1.5 | 0.2496 | 1.886e-09 | 0.0575 | 20 | 3.069e-05 | 3.072e-06 | 16275 | 16284 | Yes |
| Red nucleus | BrainMeta | RITA1 | ENSG00000139405.16 | 12 | 113626752 | rs7135901 | 12 | 113705962 | G | T | 0.09443 | 1.578 | 0.2902 | 7.1e-08 | -1.135 | 0.06081 | 8.456e-78 | -1.39 | 0.2662 | 1.786e-07 | 0.07805 | 20 | 0.002907 | 3.072e-06 | 16275 | 16284 | Yes |
| Red nucleus | BrainMeta | ACADVL | ENSG00000072778.20 | 17 | 7124514 | rs507506 | 17 | 7118322 | A | G | 0.4165 | -0.9637 | 0.15 | 1.1e-11 | 0.4706 | 0.02971 | 1.696e-56 | -2.048 | 0.3439 | 2.611e-09 | 0.06057 | 20 | 4.249e-05 | 3.072e-06 | 16275 | 16284 | Yes |
| Red nucleus | BrainMeta | SLC2A4 | ENSG00000181856.15 | 17 | 7188306 | rs5418 | 17 | 7185092 | G | A | 0.4314 | -0.839 | 0.1498 | 6.1e-09 | -0.4373 | 0.02756 | 1.095e-56 | 1.918 | 0.3633 | 1.289e-07 | 0.0118 | 20 | 0.002098 | 3.072e-06 | 16275 | 16284 | Yes |
| Substantia nigra | BrainMeta | WDR75 | ENSG00000115368.10 | 2 | 190323225 | rs142714202 | 2 | 190360073 | G | A | 0.03479 | 6.482 | 0.4803 | 1.4e-41 | -0.9401 | 0.08842 | 2.115e-26 | -6.895 | 0.8256 | 6.759e-17 | 0.2002 | 20 | 1.1e-12 | 3.072e-06 | 16275 | 16284 | Yes |
| Substantia nigra | BrainMeta | SMBD1P | ENSG00000283426.1 | 3 | 195433502 | rs6804822 | 3 | 195709662 | A | G | 0.2455 | -1.085 | 0.2145 | 7.8e-07 | 0.7998 | 0.03679 | 8.516e-105 | -1.356 | 0.2754 | 8.469e-07 | 0.07811 | 20 | 0.01378 | 3.072e-06 | 16275 | 16284 | Yes |
| Substantia nigra | BrainMeta | MUC20 | ENSG00000176945.17 | 3 | 195457874 | rs6804822 | 3 | 195709662 | A | G | 0.2455 | -1.085 | 0.2145 | 7.8e-07 | 0.859 | 0.03186 | 4.53e-160 | -1.263 | 0.2541 | 6.729e-07 | 0.05291 | 20 | 0.01095 | 3.072e-06 | 16275 | 16284 | Yes |
| Substantia nigra | BrainMeta | AC024937.2 | ENSG00000231464.1 | 3 | 195724356 | rs7614767 | 3 | 195753451 | G | A | 0.3419 | 1.164 | 0.188 | 8.1e-10 | -0.4971 | 0.05976 | 8.88e-17 | -2.342 | 0.4714 | 6.738e-07 | 0.1114 | 20 | 0.01097 | 3.072e-06 | 16275 | 16284 | Yes |
| Substantia nigra | BrainMeta | LINC02278 | ENSG00000251635.2 | 4 | 38569634 | rs4833079 | 4 | 38654681 | C | T | 0.3559 | 1.148 | 0.179 | 9.2e-10 | -0.3647 | 0.03503 | 2.217e-25 | -3.147 | 0.5764 | 4.775e-08 | 0.03663 | 20 | 0.0007771 | 3.072e-06 | 16275 | 16284 | Yes |
| Substantia nigra | BrainMeta | KIAA1109 | ENSG00000138688.16 | 4 | 123178700 | rs56313700 | 4 | 123287573 | A | C | 0.2396 | -1.174 | 0.2154 | 5.1e-08 | -0.6283 | 0.03719 | 4.853e-64 | 1.868 | 0.3602 | 2.143e-07 | 0.07602 | 20 | 0.003488 | 3.072e-06 | 16275 | 16284 | Yes |
| Substantia nigra | BrainMeta | H2BC4 | ENSG00000180596.7 | 6 | 26119628 | rs80215559 | 6 | 25918225 | C | T | 0.04274 | 3.064 | 0.3354 | 2.7e-21 | -0.423 | 0.07171 | 3.664e-09 | -7.244 | 1.462 | 7.218e-07 | 0.01229 | 18 | 0.01175 | 3.072e-06 | 16275 | 16284 | Yes |
| Substantia nigra | BrainMeta | SNX31 | ENSG00000174226.9 | 8 | 101630380 | rs1693567 | 8 | 101677386 | C | T | 0.4672 | -1.742 | 0.1777 | 9.5e-24 | 0.7291 | 0.02919 | 1.17e-137 | -2.389 | 0.2618 | 7.032e-20 | 0.5191 | 20 | 1.144e-15 | 3.072e-06 | 16275 | 16284 | Yes |
| Substantia nigra | BrainMeta | PABPC1 | ENSG00000070756.17 | 8 | 101716540 | rs1786345 | 8 | 101674751 | C | A | 0.4414 | -1.708 | 0.1788 | 1.2e-22 | 0.3413 | 0.02869 | 1.243e-32 | -5.005 | 0.672 | 9.492e-14 | 0.2284 | 20 | 1.545e-09 | 3.072e-06 | 16275 | 16284 | Yes |
| Substantia nigra | BrainMeta | LINC01505 | ENSG00000234323.8 | 9 | 109129374 | rs145825805 | 9 | 109099744 | A | G | 0.0835 | 3.239 | 0.3447 | 4.7e-21 | 0.4886 | 0.06421 | 2.772e-14 | 6.629 | 1.121 | 3.356e-09 | 0.5129 | 15 | 5.462e-05 | 3.072e-06 | 16275 | 16284 | Yes |
| Substantia nigra | BrainMeta | AREL1 | ENSG00000119682.18 | 14 | 75149980 | rs17782683 | 14 | 75167706 | A | G | 0.4682 | -0.9529 | 0.1769 | 2.4e-07 | 0.6743 | 0.02855 | 2.357e-123 | -1.413 | 0.269 | 1.498e-07 | 0.01446 | 20 | 0.002439 | 3.072e-06 | 16275 | 16284 | Yes |
| CommonFactor | BrainMeta | AC092957.1 | ENSG00000243620.2 | 3 | 146857958 | rs2654817 | 3 | 147066186 | A | G | 0.2783 | -0.07774 | 0.01556 | 5.879e-07 | 0.6182 | 0.03412 | 2.23e-73 | -0.1258 | 0.02612 | 1.468e-06 | 0.3841 | 20 | 0.01955 | 3.754e-06 | 13318 | 13327 | Yes |
| CommonFactor | BrainMeta | MUC4 | ENSG00000145113.22 | 3 | 195506240 | rs7614767 | 3 | 195753451 | G | A | 0.3419 | 0.08502 | 0.01429 | 2.709e-09 | -0.468 | 0.04931 | 2.279e-21 | -0.1817 | 0.03604 | 4.648e-07 | 0.01722 | 20 | 0.006191 | 3.754e-06 | 13318 | 13327 | Yes |
| CommonFactor | BrainMeta | AC024937.2 | ENSG00000231464.1 | 3 | 195724356 | rs7614767 | 3 | 195753451 | G | A | 0.3419 | 0.08502 | 0.01429 | 2.709e-09 | -0.4971 | 0.05976 | 8.88e-17 | -0.171 | 0.03535 | 1.308e-06 | 0.04699 | 20 | 0.01741 | 3.754e-06 | 13318 | 13327 | Yes |
| CommonFactor | BrainMeta | LINC02278 | ENSG00000251635.2 | 4 | 38569634 | rs6845639 | 4 | 38716721 | G | A | 0.3509 | 0.0947 | 0.01353 | 2.611e-12 | -0.2819 | 0.03552 | 2.088e-15 | -0.336 | 0.06402 | 1.534e-07 | 0.4206 | 20 | 0.002043 | 3.754e-06 | 13318 | 13327 | Yes |
| CommonFactor | BrainMeta | H3C3 | ENSG00000287080.2 | 6 | 26045854 | rs807212 | 6 | 26065621 | A | G | 0.327 | -0.1472 | 0.01474 | 1.721e-23 | -0.3297 | 0.0449 | 2.09e-13 | 0.4464 | 0.07546 | 3.295e-09 | 0.05287 | 20 | 4.389e-05 | 3.754e-06 | 13318 | 13327 | Yes |
| CommonFactor | BrainMeta | HCG22 | ENSG00000228789.8 | 6 | 31024447 | rs2517534 | 6 | 31017334 | A | G | 0.4264 | 0.08552 | 0.01354 | 2.716e-10 | -0.7991 | 0.03955 | 8.905e-91 | -0.107 | 0.01776 | 1.673e-09 | 0.01025 | 20 | 2.228e-05 | 3.754e-06 | 13318 | 13327 | Yes |
| CommonFactor | BrainMeta | TCF19 | ENSG00000137310.12 | 6 | 31130630 | rs1265081 | 6 | 31111675 | C | A | 0.4712 | 0.07038 | 0.01343 | 1.615e-07 | -0.4271 | 0.03688 | 5.217e-31 | -0.1648 | 0.03453 | 1.814e-06 | 0.1948 | 20 | 0.02415 | 3.754e-06 | 13318 | 13327 | Yes |
| CommonFactor | BrainMeta | RUNX2 | ENSG00000124813.23 | 6 | 45463990 | rs6458443 | 6 | 45408440 | T | G | 0.335 | -0.09977 | 0.0141 | 1.461e-12 | -0.3673 | 0.03136 | 1.099e-31 | 0.2717 | 0.04484 | 1.379e-09 | 0.1954 | 5 | 1.837e-05 | 3.754e-06 | 13318 | 13327 | Yes |
| CommonFactor | BrainMeta | SLC25A37 | ENSG00000147454.14 | 8 | 23409722 | rs4872141 | 8 | 23384559 | A | G | 0.4523 | -0.1018 | 0.0135 | 4.625e-14 | -0.3216 | 0.02763 | 2.657e-31 | 0.3167 | 0.05003 | 2.465e-10 | 0.3374 | 20 | 3.283e-06 | 3.754e-06 | 13318 | 13327 | Yes |
| CommonFactor | BrainMeta | SNX31 | ENSG00000174226.9 | 8 | 101630380 | rs1693567 | 8 | 101677386 | C | T | 0.4672 | -0.1283 | 0.01346 | 1.603e-21 | 0.7291 | 0.02919 | 1.17e-137 | -0.1759 | 0.01976 | 5.478e-19 | 0.3456 | 18 | 7.295e-15 | 3.754e-06 | 13318 | 13327 | Yes |
| CommonFactor | BrainMeta | PABPC1 | ENSG00000070756.17 | 8 | 101716540 | rs1786345 | 8 | 101674751 | C | A | 0.4414 | -0.1246 | 0.01353 | 3.285e-20 | 0.3413 | 0.02869 | 1.243e-32 | -0.365 | 0.05012 | 3.287e-13 | 0.073 | 14 | 4.377e-09 | 3.754e-06 | 13318 | 13327 | Yes |
| CommonFactor | BrainMeta | INA | ENSG00000148798.11 | 10 | 105043499 | rs4293062 | 10 | 105052624 | A | G | 0.4841 | -0.07225 | 0.01352 | 9.031e-08 | 0.6211 | 0.02935 | 2.286e-99 | -0.1163 | 0.02245 | 2.19e-07 | 0.1728 | 17 | 0.002917 | 3.754e-06 | 13318 | 13327 | Yes |
| CommonFactor | BrainMeta | DDHD1 | ENSG00000100523.16 | 14 | 53561752 | rs8015438 | 14 | 53602160 | G | T | 0.1511 | 0.09912 | 0.01868 | 1.127e-07 | -0.6512 | 0.04054 | 4.71e-58 | -0.1522 | 0.03022 | 4.72e-07 | 0.1708 | 20 | 0.006286 | 3.754e-06 | 13318 | 13327 | Yes |
| CommonFactor | BrainMeta | TTLL5 | ENSG00000119685.20 | 14 | 76260695 | rs11623813 | 14 | 76138194 | C | T | 0.33 | -0.0704 | 0.01422 | 7.392e-07 | -0.5639 | 0.03262 | 6.155e-67 | 0.1248 | 0.02623 | 1.942e-06 | 0.1305 | 20 | 0.02586 | 3.754e-06 | 13318 | 13327 | Yes |
| CommonFactor | BrainMeta | SORD2P | ENSG00000259479.6 | 15 | 45147418 | rs199138 | 15 | 45387550 | G | A | 0.9344 | 0.161 | 0.02533 | 2.068e-10 | -1.217 | 0.0775 | 1.343e-55 | -0.1323 | 0.02245 | 3.813e-09 | 0.05537 | 20 | 5.079e-05 | 3.754e-06 | 13318 | 13327 | Yes |
| CommonFactor | BrainMeta | SORD | ENSG00000140263.15 | 15 | 45342364 | rs2554451 | 15 | 45388382 | T | C | 0.9344 | 0.1623 | 0.0254 | 1.648e-10 | 0.9433 | 0.08299 | 6.134e-30 | 0.1721 | 0.03089 | 2.535e-08 | 0.05505 | 20 | 0.0003376 | 3.754e-06 | 13318 | 13327 | Yes |
| CommonFactor | BrainMeta | CTSH | ENSG00000103811.18 | 15 | 79227658 | rs12592898 | 15 | 79229199 | A | G | 0.1292 | -0.1285 | 0.02007 | 1.521e-10 | -0.5877 | 0.0459 | 1.53e-37 | 0.2187 | 0.03818 | 1.022e-08 | 0.2478 | 12 | 0.0001361 | 3.754e-06 | 13318 | 13327 | Yes |
| CommonFactor | BrainMeta | SLC2A4 | ENSG00000181856.15 | 17 | 7188306 | rs5418 | 17 | 7185092 | G | A | 0.4314 | -0.06768 | 0.01355 | 5.861e-07 | -0.4373 | 0.02756 | 1.095e-56 | 0.1548 | 0.03248 | 1.888e-06 | 0.03965 | 20 | 0.02514 | 3.754e-06 | 13318 | 13327 | Yes |
| CommonFactor | BrainMeta | LLGL1 | ENSG00000131899.11 | 17 | 18138569 | rs3862147 | 17 | 18133713 | G | T | 0.2445 | 0.1047 | 0.01577 | 3.188e-11 | -0.3285 | 0.03478 | 3.54e-21 | -0.3187 | 0.05868 | 5.612e-08 | 0.1268 | 8 | 0.0007473 | 3.754e-06 | 13318 | 13327 | Yes |
| CommonFactor | BrainMeta | RETREG3 | ENSG00000141699.11 | 17 | 40747086 | rs1024091 | 17 | 40771994 | T | C | 0.5239 | 0.08686 | 0.01353 | 1.358e-10 | -0.2043 | 0.02632 | 8.519e-15 | -0.4252 | 0.08597 | 7.547e-07 | 0.04867 | 12 | 0.01005 | 3.754e-06 | 13318 | 13327 | Yes |
| CommonFactor | BrainMeta | RN7SL739P | ENSG00000265821.1 | 17 | 43603966 | rs199516 | 17 | 44856485 | C | T | 0.2207 | 0.09062 | 0.0164 | 3.296e-08 | -0.5433 | 0.06125 | 7.232e-19 | -0.1668 | 0.03556 | 2.735e-06 | 0.134 | 11 | 0.03642 | 3.754e-06 | 13318 | 13327 | Yes |
| CommonFactor | BrainMeta | CRHR1 | ENSG00000120088.15 | 17 | 43887420 | rs199516 | 17 | 44856485 | C | T | 0.2207 | 0.09062 | 0.0164 | 3.296e-08 | 0.5535 | 0.05021 | 2.966e-28 | 0.1637 | 0.03315 | 7.843e-07 | 0.01104 | 20 | 0.01045 | 3.754e-06 | 13318 | 13327 | Yes |
| CommonFactor | BrainMeta | MAPT-IT1 | ENSG00000279685.2 | 17 | 43974656 | rs242561 | 17 | 44026548 | T | C | 0.2396 | 0.1035 | 0.01617 | 1.517e-10 | 0.3862 | 0.05374 | 6.661e-13 | 0.2681 | 0.05609 | 1.745e-06 | 0.05665 | 20 | 0.02324 | 3.754e-06 | 13318 | 13327 | Yes |
| CommonFactor | BrainMeta | FBF1 | ENSG00000188878.19 | 17 | 73921438 | rs11077798 | 17 | 73953404 | T | C | 0.2376 | 0.08113 | 0.01605 | 4.296e-07 | 0.5374 | 0.03515 | 9.415e-53 | 0.151 | 0.03145 | 1.589e-06 | 0.3721 | 16 | 0.02116 | 3.754e-06 | 13318 | 13327 | Yes |

*Abbreviations: eQTL = expression quantitative trait locus; HEIDI = heterogeneity in dependent instruments; SE = standard error; SMR = summary data-based Mendelian randomisation; SNP = single-nucleotide polymorphism. One row per gene per eQTL source, so a gene identified in both whole blood and brain cortex contributes two rows. Genes were retained where P_HEIDI > 0.01 with at least five variants in the HEIDI test.*

**Supplementary Table 6. Genetic pleiotropy between quantitative susceptibility mapping-derived brain iron and Alzheimer's disease across six subcortical regions, estimated in two independent Alzheimer's disease genome-wide association datasets.**

| QSM Brain Region | AD dataset | APOE region | PM11 | PAR | PAR (%) | P (GPA) |
|---|---|---|---|---|---|---|
| Caudate nucleus | Jansen 2019 | Excluded | 0.009057 | 0.1294 | 12.94% | <1E-300 |
| Dentate nucleus | Jansen 2019 | Excluded | 0.01353 | 0.1714 | 17.14% | <1E-300 |
| Globus pallidus | Jansen 2019 | Excluded | 0.01546 | 0.2654 | 26.54% | <1E-300 |
| Putamen | Jansen 2019 | Excluded | 0.008562 | 0.1789 | 17.89% | <1E-300 |
| Red nucleus | Jansen 2019 | Excluded | 0.01196 | 0.1624 | 16.24% | <1E-300 |
| Substantia nigra | Jansen 2019 | Excluded | 0.01597 | 0.1927 | 19.27% | <1E-300 |
| Caudate nucleus | Jansen 2019 | Included | 0.004451 | 0.0756 | 7.56% | <1E-300 |
| Dentate nucleus | Jansen 2019 | Included | 0.006746 | 0.0972 | 9.72% | <1E-300 |
| Globus pallidus | Jansen 2019 | Included | 0.009138 | 0.1784 | 17.84% | <1E-300 |
| Putamen | Jansen 2019 | Included | 0.004806 | 0.1312 | 13.12% | <1E-300 |
| Red nucleus | Jansen 2019 | Included | 0.005774 | 0.0908 | 9.08% | <1E-300 |
| Substantia nigra | Jansen 2019 | Included | 0.008162 | 0.1087 | 10.87% | <1E-300 |
| Caudate nucleus | Kunkle 2019 | Excluded | 0.02188 | 0.2280 | 22.80% | <1E-300 |
| Dentate nucleus | Kunkle 2019 | Excluded | 0.01853 | 0.1628 | 16.28% | <1E-300 |
| Globus pallidus | Kunkle 2019 | Excluded | 0.02218 | 0.2613 | 26.13% | <1E-300 |
| Putamen | Kunkle 2019 | Excluded | 0.0134 | 0.1686 | 16.86% | <1E-300 |
| Red nucleus | Kunkle 2019 | Excluded | 0.01298 | 0.1156 | 11.56% | 1.315e-184 |
| Substantia nigra | Kunkle 2019 | Excluded | 0.02048 | 0.1736 | 17.36% | <1E-300 |
| Caudate nucleus | Kunkle 2019 | Included | 0.01289 | 0.1802 | 18.02% | <1E-300 |
| Dentate nucleus | Kunkle 2019 | Included | 0.01016 | 0.1167 | 11.67% | <1E-300 |
| Globus pallidus | Kunkle 2019 | Included | 0.01346 | 0.2138 | 21.38% | <1E-300 |
| Putamen | Kunkle 2019 | Included | 0.00829 | 0.1588 | 15.88% | <1E-300 |
| Red nucleus | Kunkle 2019 | Included | 0.007045 | 0.0849 | 8.49% | <1E-300 |
| Substantia nigra | Kunkle 2019 | Included | 0.01164 | 0.1262 | 12.62% | <1E-300 |

*Abbreviations: GPA = genetic analysis incorporating pleiotropy and annotation; PAR = pleiotropic association ratio; PM11 = estimated proportion of variants associated with both traits; QSM = quantitative susceptibility mapping. PAR = PM11 / (PM10 + PM01 + PM11). The APOE region was defined as chr19:44.9-45.9 Mb (GRCh37).*

### Supplementary Table 7. Pleiotropic association ratios and genetic correlations between regional brain iron and Alzheimer's disease, with and without the APOE region.

| Brain region | AD dataset | APOE region | Method | rg | SE | P | 95% CI | Significant at P < 0.05 |
|---|---|---|---|---|---|---|---|---|
| Caudate nucleus | Jansen 2019 | Excluded | HDL | 0.1511 | 0.0534 | 0.00467 | (0.046, 0.256) | Yes |
| Dentate nucleus | Jansen 2019 | Excluded | HDL | 0.0248 | 0.0491 | 0.613 | (-0.071, 0.121) | No |
| Globus pallidus | Jansen 2019 | Excluded | HDL | 0.0893 | 0.054 | 0.0981 | (-0.017, 0.195) | No |
| Putamen | Jansen 2019 | Excluded | HDL | 0.1111 | 0.0534 | 0.0374 | (0.006, 0.216) | Yes |
| Red nucleus | Jansen 2019 | Excluded | HDL | -0.0491 | 0.0532 | 0.356 | (-0.153, 0.055) | No |
| Substantia nigra | Jansen 2019 | Excluded | HDL | 0.0274 | 0.0565 | 0.627 | (-0.083, 0.138) | No |
| Caudate nucleus | Jansen 2019 | Excluded | LDSC | 0.1613 | 0.0533 | 0.0025 | (0.057, 0.266) | Yes |
| Dentate nucleus | Jansen 2019 | Excluded | LDSC | 0.0037 | 0.0499 | 0.941 | (-0.094, 0.102) | No |
| Globus pallidus | Jansen 2019 | Excluded | LDSC | 0.0862 | 0.0573 | 0.133 | (-0.026, 0.199) | No |
| Putamen | Jansen 2019 | Excluded | LDSC | 0.0765 | 0.0556 | 0.169 | (-0.032, 0.185) | No |
| Red nucleus | Jansen 2019 | Excluded | LDSC | -0.0239 | 0.0581 | 0.68 | (-0.138, 0.090) | No |
| Substantia nigra | Jansen 2019 | Excluded | LDSC | 0.0116 | 0.0561 | 0.836 | (-0.098, 0.122) | No |
| Caudate nucleus | Jansen 2019 | Included | HDL | 0.1588 | 0.0558 | 0.00444 | (0.049, 0.268) | Yes |
| Dentate nucleus | Jansen 2019 | Included | HDL | 0.0231 | 0.0523 | 0.658 | (-0.079, 0.126) | No |
| Globus pallidus | Jansen 2019 | Included | HDL | 0.0929 | 0.0556 | 0.0944 | (-0.016, 0.202) | No |
| Putamen | Jansen 2019 | Included | HDL | 0.1163 | 0.0547 | 0.0335 | (0.009, 0.224) | Yes |
| Red nucleus | Jansen 2019 | Included | HDL | -0.051 | 0.0572 | 0.373 | (-0.163, 0.061) | No |
| Substantia nigra | Jansen 2019 | Included | HDL | 0.0278 | 0.0587 | 0.636 | (-0.087, 0.143) | No |
| Caudate nucleus | Jansen 2019 | Included | LDSC | 0.2129 | 0.0874 | 0.0149 | (0.042, 0.384) | Yes |
| Dentate nucleus | Jansen 2019 | Included | LDSC | 0.0059 | 0.0609 | 0.923 | (-0.113, 0.125) | No |
| Globus pallidus | Jansen 2019 | Included | LDSC | 0.1083 | 0.0739 | 0.143 | (-0.037, 0.253) | No |
| Putamen | Jansen 2019 | Included | LDSC | 0.092 | 0.0696 | 0.186 | (-0.044, 0.228) | No |
| Red nucleus | Jansen 2019 | Included | LDSC | -0.0206 | 0.0711 | 0.772 | (-0.160, 0.119) | No |
| Substantia nigra | Jansen 2019 | Included | LDSC | 0.0141 | 0.0703 | 0.841 | (-0.124, 0.152) | No |
| Caudate nucleus | Kunkle 2019 | Excluded | HDL | -0.0176 | 0.0339 | 0.603 | (-0.084, 0.049) | No |
| Dentate nucleus | Kunkle 2019 | Excluded | HDL | 0.05 | 0.0446 | 0.262 | (-0.037, 0.137) | No |
| Globus pallidus | Kunkle 2019 | Excluded | HDL | 0.0262 | 0.0306 | 0.392 | (-0.034, 0.086) | No |
| Putamen | Kunkle 2019 | Excluded | HDL | 0.0404 | 0.0445 | 0.364 | (-0.047, 0.128) | No |
| Red nucleus | Kunkle 2019 | Excluded | HDL | -0.0453 | 0.0314 | 0.149 | (-0.107, 0.016) | No |
| Substantia nigra | Kunkle 2019 | Excluded | HDL | -0.0189 | 0.0331 | 0.568 | (-0.084, 0.046) | No |
| Caudate nucleus | Kunkle 2019 | Excluded | LDSC | 0.0193 | 0.0631 | 0.76 | (-0.104, 0.143) | No |
| Dentate nucleus | Kunkle 2019 | Excluded | LDSC | 0.0844 | 0.054 | 0.118 | (-0.021, 0.190) | No |
| Globus pallidus | Kunkle 2019 | Excluded | LDSC | 0.0494 | 0.0635 | 0.437 | (-0.075, 0.174) | No |
| Putamen | Kunkle 2019 | Excluded | LDSC | 0.0765 | 0.0556 | 0.169 | (-0.032, 0.185) | No |
| Red nucleus | Kunkle 2019 | Excluded | LDSC | 0.0041 | 0.0571 | 0.943 | (-0.108, 0.116) | No |
| Substantia nigra | Kunkle 2019 | Excluded | LDSC | 0.0385 | 0.0636 | 0.545 | (-0.086, 0.163) | No |
| Caudate nucleus | Kunkle 2019 | Included | HDL | -0.0124 | 0.0445 | 0.781 | (-0.100, 0.075) | No |
| Dentate nucleus | Kunkle 2019 | Included | HDL | 0.0479 | 0.0526 | 0.362 | (-0.055, 0.151) | No |
| Globus pallidus | Kunkle 2019 | Included | HDL | -0.0079 | 0.0548 | 0.885 | (-0.115, 0.100) | No |
| Putamen | Kunkle 2019 | Included | HDL | 0.0456 | 0.0516 | 0.377 | (-0.056, 0.147) | No |
| Red nucleus | Kunkle 2019 | Included | HDL | -0.0576 | 0.0449 | 0.199 | (-0.146, 0.030) | No |
| Substantia nigra | Kunkle 2019 | Included | HDL | -0.026 | 0.0536 | 0.627 | (-0.131, 0.079) | No |
| Caudate nucleus | Kunkle 2019 | Included | LDSC | 0.0362 | 0.0748 | 0.629 | (-0.110, 0.183) | No |
| Dentate nucleus | Kunkle 2019 | Included | LDSC | 0.1039 | 0.0672 | 0.122 | (-0.028, 0.236) | No |
| Globus pallidus | Kunkle 2019 | Included | LDSC | 0.0553 | 0.0794 | 0.486 | (-0.100, 0.211) | No |
| Putamen | Kunkle 2019 | Included | LDSC | 0.0836 | 0.0725 | 0.249 | (-0.058, 0.226) | No |
| Red nucleus | Kunkle 2019 | Included | LDSC | 0.0146 | 0.0681 | 0.83 | (-0.119, 0.148) | No |
| Substantia nigra | Kunkle 2019 | Included | LDSC | 0.0474 | 0.0753 | 0.529 | (-0.100, 0.195) | No |

*Abbreviations: CI = confidence interval; HDL = high-definition likelihood; LDSC = linkage disequilibrium score regression; rg = genetic correlation; SE = standard error. The APOE region was defined as chr19:44.9-45.9 Mb (GRCh37). Significance is reported at P < 0.05, uncorrected.*

**Supplementary Table 8. Genome-wide significant loci from the multivariate genome-wide association analysis of the shared Alzheimer's disease-basal ganglia iron latent factor (r2 < 0.1, P < 5 x 10-8).**

| No | GenomicLocus | Unique variant ID | rsID | Chromosome | pos | p | nIndSigSNPs | IndSigSNPs |
|---|---|---|---|---|---|---|---|---|
| 1 | 1 | 1:161155392:A:G | rs4575098 | 1 | 161155392 | 1.61e-10 | 2 | rs4575098;rs4379692 |
| 2 | 2 | 1:207786828:A:G | rs2093760 | 1 | 207786828 | 7.179e-18 | 4 | rs2093760;rs11576522;rs11118328;rs6690215 |
| 3 | 3 | 2:127891427:A:C | rs4663105 | 2 | 127891427 | 8.337e-44 | 12 | rs4663105;rs744373;rs1060743;rs10194375;rs72838215; rs6431219;rs6743470;rs4663099 ... |
| 4 | 4 | 3:195836575:C:T | rs62282694 | 3 | 195836575 | 1.028e-08 | 1 | rs62282694 |
| 5 | 5 | 4:11026028:A:G | rs6448453 | 4 | 11026028 | 6.91e-09 | 1 | rs6448453 |
| 6 | 6 | 4:102681041:A:G | rs201081507 | 4 | 102681041 | 3.672e-12 | 1 | rs201081507 |
| 7 | 7 | 4:103128298:A:G | rs151431 | 4 | 103128298 | 6.766e-14 | 2 | rs151431;rs2033900 |
| 8 | 7 | 4:103288255:A:G | rs35139692 | 4 | 103288255 | 2.749e-08 | 1 | rs35139692 |
| 9 | 8 | 6:22306698:A:C | rs6910948 | 6 | 22306698 | 1.052e-08 | 1 | rs6910948 |
| 10 | 9 | 6:47432637:C:T | rs9381563 | 6 | 47432637 | 1.631e-10 | 1 | rs9381563 |
| 11 | 10 | 7:99774327:A:G | rs12705074 | 7 | 99774327 | 3.852e-09 | 3 | rs12705074;rs858502;rs13246354 |
| 12 | 10 | 7:99971834:A:G | rs1859788 | 7 | 99971834 | 1.337e-15 | 5 | rs1859788;rs2734897;rs866500;rs858502;rs35305377 |
| 13 | 11 | 7:143108158:C:T | rs7810606 | 7 | 143108158 | 4.051e-10 | 3 | rs7810606;rs35251323;rs56402156 |
| 14 | 12 | 8:27208126:A:G | rs6987305 | 8 | 27208126 | 1.598e-11 | 2 | rs6987305;rs2741342 |
| 15 | 12 | 8:27322974:A:G | rs34181358 | 8 | 27322974 | 1.162e-08 | 2 | rs34181358;rs2741342 |
| 16 | 12 | 8:27466315:C:T | rs1532278 | 8 | 27466315 | 1.79e-18 | 2 | rs1532278;rs9331908 |
| 17 | 13 | 10:11717397:C:T | rs11257238 | 10 | 11717397 | 2.008e-08 | 1 | rs11257238 |
| 18 | 14 | 11:59958380:A:C | rs2081545 | 11 | 59958380 | 6.001e-16 | 6 | rs2081545;rs11824734;rs11559565;rs554311;rs580064; rs1786140 |
| 19 | 15 | 11:85776544:A:G | rs867611 | 11 | 85776544 | 2.905e-22 | 5 | rs867611;rs7938634;rs11234556;rs527162;rs12808312 |
| 20 | 16 | 14:92938855:A:G | rs12590654 | 14 | 92938855 | 1.841e-11 | 3 | rs12590654;rs2896209;rs36026988 |
| 21 | 17 | 15:59022615:C:T | rs442495 | 15 | 59022615 | 1.133e-08 | 1 | rs442495 |
| 22 | 18 | 15:63569902:C:T | rs117618017 | 15 | 63569902 | 9.251e-09 | 1 | rs117618017 |
| 23 | 19 | 17:5138980:A:G | rs113260531 | 17 | 5138980 | 1.754e-10 | 1 | rs113260531 |
| 24 | 20 | 19:1039444:C:T | rs3795065 | 19 | 1039444 | 1.487e-09 | 3 | rs3795065;rs3752231;rs12151021 |
| 25 | 21 | 19:45071070:G:T | rs11083742 | 19 | 45071070 | 3.245e-14 | 3 | rs11083742;rs846881;rs73046410 |
| 26 | 22 | 19:51727962:A:C | rs3865444 | 19 | 51727962 | 3.643e-09 | 1 | rs3865444 |
| 27 | 23 | 20:54982351:G:T | rs6014720 | 20 | 54982351 | 3.964e-09 | 1 | rs6014720 |

*Abbreviations: Chr = chromosome; IndSig = independent significant; SNP = single-nucleotide polymorphism; uniqID = unique variant identifier. Variants showing significant Q heterogeneity, and their partners in high linkage disequilibrium, were excluded. Positions are given on GRCh37. Long variant and gene lists are abbreviated in this document; complete values are given in the accompanying spreadsheet workbook.*

### Supplementary Table 9. Significant genes from MAGMA gene-based analysis of the shared Alzheimer's disease-basal ganglia iron latent factor genome-wide association analysis (Bonferroni-corrected P < 2.62 x 10-6).

| Trait | Ensembl gene ID | Gene symbol | Chromosome | Start (bp) | Stop (bp) | SNPs (n) | Model parameters | n | Z | P | Bonferroni-adjusted P | Bonferroni threshold | Genes tested (n) | Significant |
|---|---|---|---|---|---|---|---|---|---|---|---|---|---|---|
| Shared liability factor | ENSG00000110077 | MS4A6A | 11 | 59929081 | 59987139 | 68 | 3 | 1252282 | 7.478 | 3.786e-14 | 7.224e-10 | 2.62e-06 | 19081 | Yes |
| Shared liability factor | ENSG00000136717 | BIN1 | 2 | 127795603 | 127899931 | 276 | 21 | 1252282 | 7.354 | 9.597e-14 | 1.831e-09 | 2.62e-06 | 19081 | Yes |
| Shared liability factor | ENSG00000110079 | MS4A4A | 11 | 60013014 | 60086445 | 122 | 12 | 1252282 | 7.28 | 1.67e-13 | 3.186e-09 | 2.62e-06 | 19081 | Yes |
| Shared liability factor | ENSG00000214787 | MS4A4E | 11 | 59958726 | 60045561 | 142 | 8 | 1252282 | 7.168 | 3.798e-13 | 7.246e-09 | 2.62e-06 | 19081 | Yes |
| Shared liability factor | ENSG00000140090 | SLC24A4 | 14 | 92753925 | 92972596 | 479 | 46 | 1252282 | 6.54 | 3.069e-11 | 5.855e-07 | 2.62e-06 | 19081 | Yes |
| Shared liability factor | ENSG00000149534 | MS4A2 | 11 | 59820734 | 59873444 | 96 | 7 | 1252282 | 6.485 | 4.442e-11 | 8.475e-07 | 2.62e-06 | 19081 | Yes |
| Shared liability factor | ENSG00000146904 | EPHA1 | 7 | 143077382 | 143140985 | 83 | 15 | 1252282 | 6.327 | 1.251e-10 | 2.387e-06 | 2.62e-06 | 19081 | Yes |
| Shared liability factor | ENSG00000213420 | GPC2 | 7 | 99757229 | 99809995 | 61 | 5 | 1252282 | 6.269 | 1.816e-10 | 3.464e-06 | 2.62e-06 | 19081 | Yes |
| Shared liability factor | ENSG00000073921 | PICALM | 11 | 85658727 | 85815924 | 272 | 12 | 1252282 | 6.263 | 1.89e-10 | 3.606e-06 | 2.62e-06 | 19081 | Yes |
| Shared liability factor | ENSG00000066923 | STAG3 | 7 | 99740186 | 99829111 | 114 | 7 | 1252282 | 6.127 | 4.47e-10 | 8.528e-06 | 2.62e-06 | 19081 | Yes |
| Shared liability factor | ENSG00000197093 | GAL3ST4 | 7 | 99746867 | 99801373 | 56 | 5 | 1252282 | 6.123 | 4.585e-10 | 8.748e-06 | 2.62e-06 | 19081 | Yes |
| Shared liability factor | ENSG00000213413 | PVRIG | 7 | 99780864 | 99829113 | 76 | 4 | 1252282 | 6.075 | 6.183e-10 | 1.18e-05 | 2.62e-06 | 19081 | Yes |
| Shared liability factor | ENSG00000204296 | C6orf10 | 6 | 32246303 | 32374684 | 720 | 19 | 1252282 | 6.068 | 6.484e-10 | 1.237e-05 | 2.62e-06 | 19081 | Yes |
| Shared liability factor | ENSG00000105383 | CD33 | 19 | 51693320 | 51757115 | 51 | 9 | 1252282 | 6.003 | 9.689e-10 | 1.849e-05 | 2.62e-06 | 19081 | Yes |
| Shared liability factor | ENSG00000160844 | GATS | 7 | 99788283 | 99904855 | 100 | 7 | 1252282 | 5.963 | 1.241e-09 | 2.367e-05 | 2.62e-06 | 19081 | Yes |
| Shared liability factor | ENSG00000203710 | CR1 | 1 | 207634492 | 207823992 | 198 | 18 | 1252282 | 5.941 | 1.42e-09 | 2.71e-05 | 2.62e-06 | 19081 | Yes |
| Shared liability factor | ENSG00000146826 | C7orf43 | 7 | 99742043 | 99791338 | 52 | 6 | 1252282 | 5.918 | 1.632e-09 | 3.114e-05 | 2.62e-06 | 19081 | Yes |
| Shared liability factor | ENSG00000204287 | HLA-DRA | 6 | 32372619 | 32422823 | 427 | 17 | 1252282 | 5.752 | 4.423e-09 | 8.439e-05 | 2.62e-06 | 19081 | Yes |
| Shared liability factor | ENSG00000120885 | CLU | 8 | 27444434 | 27507548 | 115 | 12 | 1252282 | 5.729 | 5.037e-09 | 9.612e-05 | 2.62e-06 | 19081 | Yes |
| Shared liability factor | ENSG00000158864 | NDUFS2 | 1 | 161131894 | 161194185 | 49 | 9 | 1252282 | 5.573 | 1.249e-08 | 0.0002383 | 2.62e-06 | 19081 | Yes |
| Shared liability factor | ENSG00000166926 | MS4A6E | 11 | 60067304 | 60174069 | 233 | 11 | 1252282 | 5.558 | 1.361e-08 | 0.0002597 | 2.62e-06 | 19081 | Yes |
| Shared liability factor | ENSG00000185899 | TAS2R60 | 7 | 143105546 | 143151502 | 58 | 9 | 1252282 | 5.528 | 1.616e-08 | 0.0003083 | 2.62e-06 | 19081 | Yes |
| Shared liability factor | ENSG00000204290 | BTNL2 | 6 | 32351740 | 32409905 | 439 | 17 | 1252282 | 5.422 | 2.946e-08 | 0.0005621 | 2.62e-06 | 19081 | Yes |
| Shared liability factor | ENSG00000158859 | ADAMTS4 | 1 | 161144098 | 161203846 | 56 | 10 | 1252282 | 5.411 | 3.139e-08 | 0.000599 | 2.62e-06 | 19081 | Yes |
| Shared liability factor | ENSG00000149516 | MS4A3 | 11 | 59789060 | 59848601 | 100 | 12 | 1252282 | 5.401 | 3.307e-08 | 0.000631 | 2.62e-06 | 19081 | Yes |
| Shared liability factor | ENSG00000158869 | FCER1G | 1 | 161150024 | 161200489 | 50 | 9 | 1252282 | 5.382 | 3.69e-08 | 0.0007042 | 2.62e-06 | 19081 | Yes |
| Shared liability factor | ENSG00000158850 | B4GALT3 | 1 | 161131100 | 161182287 | 27 | 5 | 1252282 | 5.375 | 3.826e-08 | 0.0007301 | 2.62e-06 | 19081 | Yes |
| Shared liability factor | ENSG00000268387 | AL590714.1 | 1 | 161155160 | 161202296 | 53 | 9 | 1252282 | 5.299 | 5.817e-08 | 0.00111 | 2.62e-06 | 19081 | Yes |
| Shared liability factor | ENSG00000064687 | ABCA7 | 19 | 1005102 | 1075571 | 191 | 27 | 1252282 | 5.269 | 6.877e-08 | 0.001312 | 2.62e-06 | 19081 | Yes |
| Shared liability factor | ENSG00000180448 | HMHA1 | 19 | 1030922 | 1096627 | 160 | 27 | 1252282 | 5.206 | 9.669e-08 | 0.001845 | 2.62e-06 | 19081 | Yes |
| Shared liability factor | ENSG00000182087 | TMEM259 | 19 | 999647 | 1056117 | 190 | 26 | 1252282 | 5.17 | 1.172e-07 | 0.002236 | 2.62e-06 | 19081 | Yes |
| Shared liability factor | ENSG00000064666 | CNN2 | 19 | 991298 | 1049068 | 217 | 28 | 1252282 | 5.046 | 2.25e-07 | 0.004293 | 2.62e-06 | 19081 | Yes |
| Shared liability factor | ENSG00000197721 | CR1L | 1 | 207783458 | 207921761 | 266 | 12 | 1252282 | 5.029 | 2.462e-07 | 0.004698 | 2.62e-06 | 19081 | Yes |
| Shared liability factor | ENSG00000160862 | AZGP1 | 7 | 99554343 | 99608780 | 57 | 6 | 1252282 | 5.028 | 2.475e-07 | 0.004723 | 2.62e-06 | 19081 | Yes |
| Shared liability factor | ENSG00000143224 | PPOX | 1 | 161101200 | 161157803 | 27 | 2 | 1252282 | 5.025 | 2.519e-07 | 0.004806 | 2.62e-06 | 19081 | Yes |
| Shared liability factor | ENSG00000264813 | ACE | 17 | 61527184 | 61609209 | 103 | 10 | 1252282 | 4.971 | 3.326e-07 | 0.006346 | 2.62e-06 | 19081 | Yes |
| Shared liability factor | ENSG00000159640 | ACE | 17 | 61519422 | 61609205 | 105 | 10 | 1252282 | 4.967 | 3.398e-07 | 0.006484 | 2.62e-06 | 19081 | Yes |
| Shared liability factor | ENSG00000151006 | PRSS53 | 16 | 31084746 | 31135949 | 50 | 4 | 1252282 | 4.905 | 4.66e-07 | 0.008892 | 2.62e-06 | 19081 | Yes |
| Shared liability factor | ENSG00000103507 | BCKDK | 16 | 31082428 | 31134110 | 53 | 5 | 1252282 | 4.9 | 4.796e-07 | 0.009151 | 2.62e-06 | 19081 | Yes |
| Shared liability factor | ENSG00000106261 | ZKSCAN1 | 7 | 99578204 | 99649312 | 74 | 9 | 1252282 | 4.899 | 4.829e-07 | 0.009214 | 2.62e-06 | 19081 | Yes |
| Shared liability factor | ENSG00000128923 | FAM63B | 15 | 59028391 | 59164099 | 142 | 10 | 1252282 | 4.871 | 5.543e-07 | 0.01058 | 2.62e-06 | 19081 | Yes |
| Shared liability factor | ENSG00000255439 | RP11-196G11.1 | 16 | 31084760 | 31141277 | 55 | 5 | 1252282 | 4.834 | 6.704e-07 | 0.01279 | 2.62e-06 | 19081 | Yes |
| Shared liability factor | ENSG00000167397 | VKORC1 | 16 | 31092163 | 31142301 | 52 | 5 | 1252282 | 4.807 | 7.674e-07 | 0.01464 | 2.62e-06 | 19081 | Yes |
| Shared liability factor | ENSG00000143258 | USP21 | 1 | 161094240 | 161145513 | 26 | 2 | 1252282 | 4.776 | 8.943e-07 | 0.01706 | 2.62e-06 | 19081 | Yes |
| Shared liability factor | ENSG00000052344 | PRSS8 | 16 | 31132756 | 31182083 | 34 | 6 | 1252282 | 4.772 | 9.128e-07 | 0.01742 | 2.62e-06 | 19081 | Yes |
| Shared liability factor | ENSG00000103510 | KAT8 | 16 | 31092075 | 31152714 | 57 | 5 | 1252282 | 4.752 | 1.009e-06 | 0.01925 | 2.62e-06 | 19081 | Yes |
| Shared liability factor | ENSG00000198087 | CD2AP | 6 | 47410525 | 47604999 | 267 | 8 | 1252282 | 4.739 | 1.073e-06 | 0.02047 | 2.62e-06 | 19081 | Yes |
| Shared liability factor | ENSG00000143222 | UFC1 | 1 | 161087566 | 161138646 | 23 | 2 | 1252282 | 4.733 | 1.108e-06 | 0.02115 | 2.62e-06 | 19081 | Yes |
| Shared liability factor | ENSG00000158882 | TOMM40L | 1 | 161160793 | 161210408 | 62 | 10 | 1252282 | 4.73 | 1.122e-06 | 0.02141 | 2.62e-06 | 19081 | Yes |

| Trait | Ensembl gene ID | Gene symbol | Chromosome | Start (bp) | Stop (bp) | SNPs (n) | Model parameters | n | Z | P | Bonferroni-adjusted P | Bonferroni threshold | Genes tested (n) | Significant |
|---|---|---|---|---|---|---|---|---|---|---|---|---|---|---|
| Shared liability factor | ENSG00000268927 | FLJ00418 | 16 | 70660570 | 70709739 | 86 | 5 | 1252282 | 4.693 | 1.344e-06 | 0.02564 | 2.62e-06 | 19081 | Yes |
| Shared liability factor | ENSG00000180787 | ZFP3 | 17 | 4946543 | 5009669 | 125 | 12 | 1252282 | 4.681 | 1.429e-06 | 0.02727 | 2.62e-06 | 19081 | Yes |
| Shared liability factor | ENSG00000159840 | ZYX | 7 | 143043173 | 143098204 | 62 | 15 | 1252282 | 4.676 | 1.46e-06 | 0.02785 | 2.62e-06 | 19081 | Yes |
| Shared liability factor | ENSG00000158796 | DEDD | 1 | 161080764 | 161137478 | 27 | 2 | 1252282 | 4.668 | 1.518e-06 | 0.02897 | 2.62e-06 | 19081 | Yes |
| Shared liability factor | ENSG00000143256 | PFDN2 | 1 | 161060346 | 161122901 | 37 | 2 | 1252282 | 4.659 | 1.59e-06 | 0.03034 | 2.62e-06 | 19081 | Yes |
| Shared liability factor | ENSG00000161929 | SCIMP | 17 | 5102256 | 5173155 | 97 | 10 | 1252282 | 4.565 | 2.503e-06 | 0.04775 | 2.62e-06 | 19081 | Yes |
| Shared liability factor | ENSG00000168918 | INPP5D | 2 | 233889677 | 234126549 | 365 | 24 | 1252282 | 4.556 | 2.604e-06 | 0.04968 | 2.62e-06 | 19081 | Yes |

*Abbreviations: Chr = chromosome; MAGMA = Multi-marker Analysis of GenoMic Annotation; NPARAM = number of model parameters; NSNPS = number of variants in the gene window; P_bonf = Bonferroni-adjusted P-value; ZSTAT = gene-based Z statistic.*

### Supplementary Table 10. Putative causal genes for the shared Alzheimer's disease-basal ganglia iron latent factor identified by summary data-based Mendelian randomisation, after Bonferroni correction and HEIDI filtering.

| Trait | eQTL source | Gene symbol | Probe ID | Probe chromosome | Probe position (bp) | Top SNP | Top SNP chromosome | Top SNP position (bp) | Effect allele | Other allele | Effect allele frequency | Beta (GWAS) | SE (GWAS) | P (GWAS) | Beta (eQTL) | SE (eQTL) | P (eQTL) | Beta (SMR) | SE (SMR) | P (SMR) | P (HEIDI) | HEIDI SNPs (n) | Bonferroni-adjusted P (SMR) | Bonferroni threshold | Unique genes tested (n) | Tests performed (n) | Significant |
|---|---|---|---|---|---|---|---|---|---|---|---|---|---|---|---|---|---|---|---|---|---|---|---|---|---|---|---|
| Shared liability factor | eQTLGen_Blood | RIN3 | ENSG00000100599 | 14 | 93067728 | rs17783630 | 14 | 92955385 | A | C | 0.4592 | -0.006342 | 0.001197 | 1.185e-07 | 0.1856 | 0.007918 | 1.785e-121 | -0.03417 | 0.006615 | 2.396e-07 | 0.01534 | 9 | 0.003378 | 3.547e-06 | 14095 | 310090 | Yes |
| Shared liability factor | eQTLGen_Blood | KAT8 | ENSG00000103510 | 16 | 31134894 | rs1549299 | 16 | 31154146 | G | A | 0.3022 | -0.006786 | 0.001286 | 1.323e-07 | 0.2396 | 0.009451 | 9.407e-142 | -0.02833 | 0.005485 | 2.405e-07 | 0.1317 | 20 | 0.00339 | 3.547e-06 | 14095 | 310090 | Yes |
| Shared liability factor | eQTLGen_Blood | SLC24A4 | ENSG00000140090 | 14 | 92875760 | rs17783630 | 14 | 92955385 | A | C | 0.4592 | -0.006342 | 0.001197 | 1.185e-07 | 0.4672 | 0.007543 | 0 | -0.01357 | 0.002572 | 1.316e-07 | 0.02037 | 9 | 0.001855 | 3.547e-06 | 14095 | 310090 | Yes |
| Shared liability factor | eQTLGen_Blood | EPHA1 | ENSG00000146904 | 7 | 143096683 | rs12703526 | 7 | 143107588 | G | T | 0.5219 | -0.007409 | 0.001202 | 7.163e-10 | 0.2004 | 0.007923 | 3.377e-141 | -0.03696 | 0.006174 | 2.133e-09 | 0.7275 | 9 | 3.006e-05 | 3.547e-06 | 14095 | 310090 | Yes |
| Shared liability factor | eQTLGen_Blood | ZNF232 | ENSG00000167840 | 17 | 5017623 | rs934631 | 17 | 5075838 | C | A | 0.1163 | 0.008974 | 0.001867 | 1.534e-06 | -0.2498 | 0.01233 | 3.538e-91 | -0.03593 | 0.007683 | 2.914e-06 | 0.2933 | 20 | 0.04107 | 3.547e-06 | 14095 | 310090 | Yes |
| Shared liability factor | eQTLGen_Blood | CTSW | ENSG00000172543 | 11 | 65649246 | rs658938 | 11 | 65651830 | A | G | 0.1889 | -0.006946 | 0.00149 | 3.135e-06 | -0.9867 | 0.008568 | 0 | 0.00704 | 0.001511 | 3.194e-06 | 0.7696 | 20 | 0.04502 | 3.547e-06 | 14095 | 310090 | Yes |
| Shared liability factor | eQTLGen_Blood | PRSS36 | ENSG00000178226 | 16 | 31155830 | rs1549299 | 16 | 31154146 | G | A | 0.3022 | -0.006786 | 0.001286 | 1.323e-07 | 0.1307 | 0.00944 | 1.349e-43 | -0.05192 | 0.01053 | 8.228e-07 | 0.02456 | 20 | 0.0116 | 3.547e-06 | 14095 | 310090 | Yes |
| Shared liability factor | eQTLGen_Blood | SPACDR | ENSG00000185955 | 7 | 100058066 | rs34919929 | 7 | 100012334 | G | A | 0.3052 | -0.009675 | 0.001285 | 5.102e-14 | -0.08424 | 0.008703 | 3.703e-22 | 0.1148 | 0.01933 | 2.8e-09 | 0.01806 | 20 | 3.947e-05 | 3.547e-06 | 14095 | 310090 | Yes |
| Shared liability factor | eQTLGen_Blood | ZNF232-AS1 | ENSG00000234327 | 17 | 5016531 | rs934631 | 17 | 5075838 | C | A | 0.1163 | 0.008974 | 0.001867 | 1.534e-06 | -0.8651 | 0.01538 | 0 | -0.01037 | 0.002166 | 1.674e-06 | 0.0252 | 20 | 0.02359 | 3.547e-06 | 14095 | 310090 | Yes |
| Shared liability factor | eQTLGen_Blood | CR1-AS1 | ENSG00000236911 | 1 | 207752585 | rs2274566 | 1 | 207753345 | C | T | 0.4026 | 0.006105 | 0.001196 | 3.34e-07 | -0.3381 | 0.02024 | 1.189e-62 | -0.01806 | 0.0037 | 1.058e-06 | 0.06169 | 13 | 0.01491 | 3.547e-06 | 14095 | 310090 | Yes |
| Shared liability factor | eQTLGen_Blood | SIGLEC22P | ENSG00000268849 | 19 | 51714840 | rs12609179 | 19 | 51722582 | G | A | 0.4483 | 0.006 | 0.001205 | 6.439e-07 | -0.2633 | 0.019 | 1.098e-43 | -0.02278 | 0.004864 | 2.804e-06 | 0.4109 | 7 | 0.03953 | 3.547e-06 | 14095 | 310090 | Yes |
| Shared liability factor | BrainMeta | CR1 | ENSG00000203710.12 | 1 | 207742301 | rs679515 | 1 | 207750568 | T | C | 0.172 | 0.01359 | 0.001578 | 7.219e-18 | 0.8347 | 0.03594 | 2.48e-119 | 0.01628 | 0.002017 | 6.788e-16 | 0.04336 | 20 | 8.829e-12 | 3.844e-06 | 13006 | 13014 | Yes |
| Shared liability factor | BrainMeta | TSBP1-AS1 | ENSG00000225914.3 | 6 | 32298978 | rs1967688 | 6 | 32340068 | T | C | 0.4791 | -0.006092 | 0.001209 | 4.724e-07 | 0.5822 | 0.04428 | 1.706e-39 | -0.01046 | 0.002224 | 2.552e-06 | 0.0728 | 20 | 0.0332 | 3.844e-06 | 13006 | 13014 | Yes |
| Shared liability factor | BrainMeta | HLA-DRB6 | ENSG00000229391.7 | 6 | 32524144 | rs9271515 | 6 | 32589645 | A | G | 0.3837 | 0.006848 | 0.001233 | 2.761e-08 | -0.887 | 0.0399 | 1.769e-109 | -0.00772 | 0.001432 | 7.04e-08 | 0.01568 | 20 | 0.0009156 | 3.844e-06 | 13006 | 13014 | Yes |
| Shared liability factor | BrainMeta | HLA-DRB1 | ENSG00000196126.11 | 6 | 32552086 | rs9270971 | 6 | 32573598 | T | C | 0.2674 | 0.007201 | 0.00132 | 4.897e-08 | 0.912 | 0.04064 | 1.557e-111 | 0.007896 | 0.00149 | 1.154e-07 | 0.04825 | 20 | 0.001501 | 3.844e-06 | 13006 | 13014 | Yes |
| Shared liability factor | BrainMeta | AL355353.1 | ENSG00000270761.1 | 6 | 47445144 | rs9381563 | 6 | 47432637 | C | T | 0.3221 | 0.008004 | 0.001252 | 1.631e-10 | -0.2552 | 0.03689 | 4.519e-12 | -0.03136 | 0.006678 | 2.658e-06 | 0.1141 | 8 | 0.03457 | 3.844e-06 | 13006 | 13014 | Yes |
| Shared liability factor | BrainMeta | CD2AP | ENSG00000198087.7 | 6 | 47520262 | rs9381563 | 6 | 47432637 | C | T | 0.3221 | 0.008004 | 0.001252 | 1.631e-10 | 0.2665 | 0.03415 | 5.964e-15 | 0.03003 | 0.006072 | 7.598e-07 | 0.06662 | 19 | 0.009882 | 3.844e-06 | 13006 | 13014 | Yes |
| Shared liability factor | BrainMeta | AP4M1 | ENSG00000221838.10 | 7 | 99703570 | rs858505 | 7 | 99819577 | G | A | 0.2674 | -0.00834 | 0.001338 | 4.554e-10 | 0.2928 | 0.03608 | 4.822e-16 | -0.02848 | 0.005761 | 7.664e-07 | 0.1443 | 20 | 0.009968 | 3.844e-06 | 13006 | 13014 | Yes |
| Shared liability factor | BrainMeta | PRSS36 | ENSG00000178226.11 | 16 | 31155826 | rs1549299 | 16 | 31154146 | G | A | 0.3022 | -0.006786 | 0.001286 | 1.323e-07 | 0.6842 | 0.03474 | 2.325e-86 | -0.009919 | 0.001946 | 3.469e-07 | 0.01457 | 20 | 0.004512 | 3.844e-06 | 13006 | 13014 | Yes |
| Shared liability factor | BrainMeta | ACE | ENSG00000159640.17 | 17 | 61565082 | rs4292 | 17 | 61554341 | C | T | 0.3678 | -0.006082 | 0.001236 | 8.673e-07 | 0.5639 | 0.03182 | 2.704e-70 | -0.01079 | 0.002275 | 2.133e-06 | 0.05793 | 20 | 0.02774 | 3.844e-06 | 13006 | 13014 | Yes |

*Abbreviations: eQTL = expression quantitative trait locus; HEIDI = heterogeneity in dependent instruments; SE = standard error; SMR = summary data-based Mendelian randomisation. One row per gene per eQTL source.*

**Supplementary Table 11. Competitive gene-set analysis of the shared Alzheimer's disease-basal ganglia iron latent factor genome-wide association analysis. (A) The 50 most significant gene sets. (B) All gene sets relating to ferroptosis, oxidative stress-induced cell death, lipid peroxidation, glutathione metabolism or iron handling.**

| Panel | Rank | Gene set | Genes (n) | Beta | Standardised beta | SE | P |
|---|---|---|---|---|---|---|---|
| A (50 most significant) | 1 | WP_COMPLEMENT_AND_COAGULATION_CASCADES | 53 | 0.7115 | 0.03745 | 0.1269 | 1.053e-08 |
| A (50 most significant) | 2 | GOBP_IMMUNE_COMPLEX_CLEARANCE | 5 | 1.87 | 0.03026 | 0.3367 | 1.436e-08 |
| A (50 most significant) | 3 | GOCC_NEUROFIBRILLARY_TANGLE | 5 | 1.51 | 0.02445 | 0.2786 | 3.013e-08 |
| A (50 most significant) | 4 | GOBP_NEGATIVE_REGULATION_OF_AMYLOID_PRECURSOR_PROTEIN_CATABOLIC_PROCESS | 18 | 1.14 | 0.03499 | 0.2123 | 4.033e-08 |
| A (50 most significant) | 5 | GOBP_NEGATIVE_REGULATION_OF_METALLOENDOPEPTIDASE_ACTIVITY | 4 | 2.363 | 0.03421 | 0.4704 | 2.57e-07 |
| A (50 most significant) | 6 | GOBP_MICROGLIAL_CELL_PROLIFERATION | 8 | 1.25 | 0.02559 | 0.2672 | 1.46e-06 |
| A (50 most significant) | 7 | GOBP_MACROPHAGE_ACTIVATION_INVOLVED_IN_IMMUNE_RESPONSE | 19 | 0.8921 | 0.02814 | 0.1968 | 2.936e-06 |
| A (50 most significant) | 8 | GOBP_NEGATIVE_REGULATION_OF_METALLOPEPTIDASE_ACTIVITY | 6 | 1.865 | 0.03306 | 0.4134 | 3.264e-06 |
| A (50 most significant) | 9 | GOBP_MACROPHAGE_PROLIFERATION | 11 | 1.069 | 0.02566 | 0.2412 | 4.676e-06 |
| A (50 most significant) | 10 | GOBP_REGULATION_OF_ASPARTIC_TYPE_PEPTIDASE_ACTIVITY | 12 | 1.34 | 0.03359 | 0.3027 | 4.857e-06 |
| A (50 most significant) | 11 | GOBP_GLIAL_CELL_PROLIFERATION | 49 | 0.547 | 0.02769 | 0.1246 | 5.661e-06 |
| A (50 most significant) | 12 | GOMF_COMPLEMENT_BINDING | 25 | 0.9068 | 0.0328 | 0.2098 | 7.764e-06 |
| A (50 most significant) | 13 | GOBP_TRANSDIFFERENTIATION | 7 | 1.22 | 0.02337 | 0.2878 | 1.129e-05 |
| A (50 most significant) | 14 | GOBP_POSITIVE_REGULATION_OF_COMPLEMENT_ACTIVATION | 5 | 1.582 | 0.0256 | 0.3795 | 1.541e-05 |
| A (50 most significant) | 15 | GOBP_ANTIGEN_RECEPTOR_MEDIATED_SIGNALING_PATHWAY | 168 | 0.2685 | 0.02508 | 0.06585 | 2.295e-05 |
| A (50 most significant) | 16 | GOMF_OPSONIN_BINDING | 20 | 0.8406 | 0.0272 | 0.207 | 2.468e-05 |
| A (50 most significant) | 17 | GOMF_GTPASE_BINDING | 287 | 0.2036 | 0.02479 | 0.05202 | 4.548e-05 |
| A (50 most significant) | 18 | GOBP_BEHAVIORAL_RESPONSE_TO_ETHANOL | 9 | 0.9488 | 0.0206 | 0.2433 | 4.825e-05 |
| A (50 most significant) | 19 | GOCC_EXTRINSIC_COMPONENT_OF_PRESYNAPTIC_MEMBRANE | 5 | 1.587 | 0.02568 | 0.4093 | 5.339e-05 |
| A (50 most significant) | 20 | GOCC_TRANSPORT_VESICLE | 395 | 0.1783 | 0.02539 | 0.04637 | 6.047e-05 |
| A (50 most significant) | 21 | GOBP_COMPLEMENT_ACTIVATION_CLASSICAL_PATHWAY | 30 | 0.6277 | 0.02487 | 0.1665 | 8.214e-05 |
| A (50 most significant) | 22 | GOBP_POSITIVE_REGULATION_OF_PROTEIN_CONTAINING_COMPLEX_ASSEMBLY | 186 | 0.2427 | 0.02385 | 0.06487 | 9.168e-05 |
| A (50 most significant) | 23 | GOBP_ACTIVATION_OF_IMMUNE_RESPONSE | 430 | 0.1616 | 0.02398 | 0.04329 | 9.495e-05 |
| A (50 most significant) | 24 | GOBP_REGULATION_OF_ASPARTIC_TYPE_ENDOPEPTIDASE_ACTIVITY_INVOLVED_IN_AMYLOID_PREC ... | 10 | 1.244 | 0.02846 | 0.3333 | 9.578e-05 |
| A (50 most significant) | 25 | GOBP_OLFACTORY_NERVE_DEVELOPMENT | 6 | 1.368 | 0.02425 | 0.3668 | 9.632e-05 |
| A (50 most significant) | 26 | GOMF_LOW_DENSITY_LIPOPROTEIN_PARTICLE_RECEPTOR_BINDING | 23 | 0.6596 | 0.02289 | 0.1782 | 0.0001079 |
| A (50 most significant) | 27 | CHEN_METABOLIC_SYNDROM_NETWORK | 1162 | 0.1007 | 0.02407 | 0.02733 | 0.0001158 |
| A (50 most significant) | 28 | GOBP_PROTEIN_LOCALIZATION_TO_CILIUM | 67 | 0.3909 | 0.02312 | 0.1075 | 0.0001391 |
| A (50 most significant) | 29 | GOBP_REGULATION_OF_COMPLEMENT_ACTIVATION | 18 | 0.7744 | 0.02378 | 0.2153 | 0.0001612 |
| A (50 most significant) | 30 | GOBP_IMMUNE_RESPONSE_REGULATING_SIGNALING_PATHWAY | 392 | 0.1573 | 0.02231 | 0.04375 | 0.0001629 |
| A (50 most significant) | 31 | REACTOME_RAS_SIGNALING_DOWNSTREAM_OF_NF1_LOSS_OF_FUNCTION_VARIANTS | 7 | 1.236 | 0.02367 | 0.3469 | 0.0001844 |
| A (50 most significant) | 32 | GOBP_HUMORAL_IMMUNE_RESPONSE_MEDIATED_BY_CIRCULATING_IMMUNOGLOBULIN | 41 | 0.5008 | 0.02319 | 0.1411 | 0.0001938 |
| A (50 most significant) | 33 | GOBP_REGULATION_OF_LYSOSOME_ORGANIZATION | 6 | 1.647 | 0.02919 | 0.4643 | 0.0001959 |
| A (50 most significant) | 34 | GOBP_REGULATION_OF_METALLOENDOPEPTIDASE_ACTIVITY | 7 | 1.159 | 0.0222 | 0.3279 | 0.0002036 |
| A (50 most significant) | 35 | BIOCARTA_ION_PATHWAY | 5 | 1.701 | 0.02753 | 0.4815 | 0.0002062 |
| A (50 most significant) | 36 | GOCC_ENDOSOME | 976 | 0.1035 | 0.02281 | 0.0294 | 0.0002154 |
| A (50 most significant) | 37 | GOMF_CLATHRIN_HEAVY_CHAIN_BINDING | 10 | 1.143 | 0.02617 | 0.3257 | 0.0002248 |
| A (50 most significant) | 38 | FRASOR_RESPONSE_TO_ESTRADIOL_UP | 35 | 0.5605 | 0.02398 | 0.1598 | 0.0002272 |

| Panel | Rank | Gene set | Genes (n) | Beta | Standardised beta | SE | P |
|---|---|---|---|---|---|---|---|
| A (50 most significant) | 39 | GOBP_COMPLEMENT_ACTIVATION | 50 | 0.4728 | 0.02417 | 0.1358 | 0.0002508 |
| A (50 most significant) | 40 | GOBP_POSITIVE_REGULATION_OF_IMMUNE_SYSTEM_PROCESS | 925 | 0.1045 | 0.02245 | 0.03026 | 0.0002767 |
| A (50 most significant) | 41 | GOBP_B_CELL_RECEPTOR_SIGNALING_PATHWAY | 57 | 0.3972 | 0.02168 | 0.1161 | 0.000313 |
| A (50 most significant) | 42 | GOBP_POSITIVE_REGULATION_OF_CELL_DEVELOPMENT | 403 | 0.1566 | 0.02251 | 0.04581 | 0.0003162 |
| A (50 most significant) | 43 | GOMF_PROTEIN_CONTAINING_COMPLEX_BINDING | 1224 | 0.08831 | 0.02164 | 0.02591 | 0.0003272 |
| A (50 most significant) | 44 | GOBP_ANTIGEN_PROCESSING_AND_PRESENTATION_OF_PEPTIDE_ANTIGEN | 68 | 0.4223 | 0.02517 | 0.1239 | 0.0003287 |
| A (50 most significant) | 45 | GOCC_ENDOSOME_MEMBRANE | 507 | 0.1372 | 0.02206 | 0.04028 | 0.0003318 |
| A (50 most significant) | 46 | GOBP_NEGATIVE_REGULATION_OF_DOPAMINE_SECRETION | 5 | 1.51 | 0.02444 | 0.449 | 0.0003871 |
| A (50 most significant) | 47 | GOMF_INTEGRIN_BINDING | 146 | 0.2364 | 0.0206 | 0.07065 | 0.0004109 |
| A (50 most significant) | 48 | WP_B_CELL_RECEPTOR_SIGNALING_PATHWAY | 92 | 0.3018 | 0.0209 | 0.09073 | 0.0004419 |
| A (50 most significant) | 49 | GOBP_MICROGLIAL_CELL_ACTIVATION_INVOLVED_IN_IMMUNE_RESPONSE | 5 | 1.041 | 0.01685 | 0.3145 | 0.0004676 |
| A (50 most significant) | 50 | GOBP_SYNAPTIC_VESICLE_BUDDING | 9 | 0.9937 | 0.02158 | 0.3014 | 0.0004897 |
| B (iron and oxidative stress) | 437 | GOBP_IRON_ION_TRANSMEMBRANE_TRANSPORT | 16 | 0.5219 | 0.01511 | 0.231 | 0.01194 |
| B (iron and oxidative stress) | 915 | KEGG_GLUTATHIONE_METABOLISM | 47 | 0.2631 | 0.01304 | 0.1442 | 0.03399 |
| B (iron and oxidative stress) | 1295 | GOBP_REGULATION_OF_IRON_ION_TRANSPORT | 5 | 0.4501 | 0.007286 | 0.2768 | 0.05199 |
| B (iron and oxidative stress) | 1338 | GOMF_GLUTATHIONE_PEROXIDASE_ACTIVITY | 20 | 0.3488 | 0.01129 | 0.217 | 0.05399 |
| B (iron and oxidative stress) | 1462 | GOBP_IRON_ION_TRANSPORT | 50 | 0.2021 | 0.01033 | 0.1298 | 0.05969 |
| B (iron and oxidative stress) | 1793 | GOBP_IRON_ION_IMPORT_ACROSS_PLASMA_MEMBRANE | 4 | 0.5262 | 0.007618 | 0.3677 | 0.07624 |
| B (iron and oxidative stress) | 1821 | REACTOME_GLUTATHIONE_CONJUGATION | 35 | 0.2154 | 0.009216 | 0.1518 | 0.07797 |
| B (iron and oxidative stress) | 1831 | GOBP_GLUTATHIONE_TRANSMEMBRANE_TRANSPORT | 7 | 0.495 | 0.00948 | 0.3497 | 0.07848 |
| B (iron and oxidative stress) | 2698 | GOBP_POSITIVE_REGULATION_OF_OXIDATIVE_STRESS_INDUCED_CELL_DEATH | 17 | 0.242 | 0.007221 | 0.2099 | 0.1245 |
| B (iron and oxidative stress) | 2938 | GOMF_IRON_ION_TRANSMEMBRANE_TRANSPORTER_ACTIVITY | 8 | 0.4334 | 0.008873 | 0.3973 | 0.1377 |
| B (iron and oxidative stress) | 2964 | GOBP_RESPONSE_TO_IRON_ION | 29 | 0.1796 | 0.006997 | 0.1657 | 0.1392 |
| B (iron and oxidative stress) | 3143 | GOBP_REGULATION_OF_OXIDATIVE_STRESS_INDUCED_CELL_DEATH | 64 | 0.124 | 0.007167 | 0.1185 | 0.1477 |
| B (iron and oxidative stress) | 3231 | GOMF_GLUTATHIONE_TRANSFERASE_ACTIVITY | 26 | 0.1998 | 0.007371 | 0.1943 | 0.1519 |
| B (iron and oxidative stress) | 3495 | GOBP_POSITIVE_REGULATION_OF_OXIDATIVE_STRESS_INDUCED_NEURON_DEATH | 7 | 0.3252 | 0.006227 | 0.3349 | 0.1658 |
| B (iron and oxidative stress) | 3699 | GOBP_GLUTATHIONE_TRANSPORT | 9 | 0.2736 | 0.00594 | 0.2953 | 0.1772 |
| B (iron and oxidative stress) | 4158 | REACTOME_GLUTATHIONE_SYNTHESIS_AND_RECYCLING | 12 | 0.1929 | 0.004836 | 0.2303 | 0.2011 |
| B (iron and oxidative stress) | 4205 | WP_GLUTATHIONE_METABOLISM | 18 | 0.1814 | 0.005569 | 0.2184 | 0.2031 |
| B (iron and oxidative stress) | 4292 | GOBP_MULTICELLULAR_ORGANISMAL_LEVEL_IRON_ION_HOMEOSTASIS | 9 | 0.2821 | 0.006125 | 0.3464 | 0.2077 |
| B (iron and oxidative stress) | 4305 | GOMF_GLUTATHIONE_DISULFIDE_OXIDOREDUCTASE_ACTIVITY | 5 | 0.3567 | 0.005774 | 0.439 | 0.2082 |
| B (iron and oxidative stress) | 5774 | GOBP_INTRACELLULAR_IRON_ION_HOMEOSTASIS | 56 | 0.06813 | 0.003685 | 0.1224 | 0.2888 |
| B (iron and oxidative stress) | 6051 | GOMF_FERRIC_IRON_BINDING | 9 | 0.1701 | 0.003693 | 0.3336 | 0.3051 |
| B (iron and oxidative stress) | 6719 | GOMF_GLUTATHIONE_TRANSMEMBRANE_TRANSPORTER_ACTIVITY | 5 | 0.1623 | 0.002628 | 0.4039 | 0.3439 |
| B (iron and oxidative stress) | 6981 | GOBP_IRON_ION_HOMEOSTASIS | 78 | 0.03556 | 0.002269 | 0.09835 | 0.3588 |
| B (iron and oxidative stress) | 7082 | GOBP_NEGATIVE_REGULATION_OF_OXIDATIVE_STRESS_INDUCED_CELL_DEATH | 44 | 0.05245 | 0.002516 | 0.1506 | 0.3638 |
| B (iron and oxidative stress) | 8723 | GOBP_GLUTATHIONE_CATABOLIC_PROCESS | 8 | 0.03047 | 0.0006237 | 0.2979 | 0.4593 |
| B (iron and oxidative stress) | 8970 | WP_FERROPTOSIS | 58 | 0.007214 | 0.0003972 | 0.1118 | 0.4743 |
| B (iron and oxidative stress) | 9145 | GOBP_NEGATIVE_REGULATION_OF_OXIDATIVE_STRESS_INDUCED_NEURON_DEATH | 20 | 0.008342 | 0.0002699 | 0.2242 | 0.4852 |
| B (iron and oxidative stress) | 9663 | GOMF_IRON_ION_BINDING | 140 | -0.003472 | -0.0002963 | 0.08052 | 0.5172 |
| B (iron and oxidative stress) | 9856 | GOBP_POSITIVE_REGULATION_OF_OXIDATIVE_STRESS_INDUCED_INTRINSIC_APOPTOTIC_SIGNALI ... | 6 | -0.02434 | -0.0004316 | 0.3519 | 0.5276 |
| B (iron and oxidative stress) | 10352 | GOMF_FERROUS_IRON_TRANSMEMBRANE_TRANSPORTER_ACTIVITY | 5 | -0.07895 | -0.001278 | 0.5482 | 0.5573 |

| Panel | Rank | Gene set | Genes (n) | Beta | Standardised beta | SE | P |
|---|---|---|---|---|---|---|---|
| B (iron and oxidative stress) | 10498 | GOBP_GLUTATHIONE_METABOLIC_PROCESS | 50 | -0.02286 | -0.001169 | 0.1364 | 0.5666 |
| B (iron and oxidative stress) | 10601 | GOBP_CELLULAR_RESPONSE_TO_IRON_ION | 8 | -0.05056 | -0.001035 | 0.2752 | 0.5729 |
| B (iron and oxidative stress) | 10636 | GOMF_FERROUS_IRON_BINDING | 24 | -0.03535 | -0.001253 | 0.1866 | 0.5751 |
| B (iron and oxidative stress) | 11876 | GOBP_SEQUESTERING_OF_IRON_ION | 5 | -0.1832 | -0.002965 | 0.4693 | 0.6518 |
| B (iron and oxidative stress) | 12249 | GOMF_GLUTATHIONE_BINDING | 10 | -0.157 | -0.003595 | 0.346 | 0.675 |
| B (iron and oxidative stress) | 12360 | WP_GAMMAGLUTAMYL_CYCLE_FOR_THE_BIOSYNTHESIS_AND_DEGRADATION_OF_GLUTATHIONE_INCLU ... | 6 | -0.1676 | -0.002972 | 0.3565 | 0.6809 |
| B (iron and oxidative stress) | 13387 | GOMF_GLUTATHIONE_HYDROLASE_ACTIVITY | 6 | -0.276 | -0.004894 | 0.4169 | 0.746 |
| B (iron and oxidative stress) | 15118 | GOMF_ABC_TYPE_GLUTATHIONE_S_CONJUGATE_TRANSPORTER_ACTIVITY | 8 | -0.3287 | -0.00673 | 0.305 | 0.8594 |
| B (iron and oxidative stress) | 15210 | REACTOME_OXIDATIVE_STRESS_INDUCED_SENESCENCE | 89 | -0.1096 | -0.007471 | 0.09941 | 0.865 |
| B (iron and oxidative stress) | 15639 | GOBP_REGULATION_OF_OXIDATIVE_STRESS_INDUCED_INTRINSIC_APOPTOTIC_SIGNALING_PATHWA ... | 26 | -0.2399 | -0.008849 | 0.189 | 0.8978 |
| B (iron and oxidative stress) | 16727 | GOBP_NEGATIVE_REGULATION_OF_OXIDATIVE_STRESS_INDUCED_NEURON_INTRINSIC_APOPTOTIC_ ... | 4 | -0.7964 | -0.01153 | 0.4045 | 0.9755 |
| B (iron and oxidative stress) | 16887 | GOBP_NEGATIVE_REGULATION_OF_OXIDATIVE_STRESS_INDUCED_INTRINSIC_APOPTOTIC_SIGNALI ... | 17 | -0.5933 | -0.0177 | 0.262 | 0.9882 |

*Abbreviations: BETA_STD = standardised effect size; MAGMA = Multi-marker Analysis of GenoMic Annotation; NGENES = number of genes in the set; RANK = rank by competitive gene-set P-value. Competitive gene-set analysis was run across 17 009 MSigDB gene sets; the Bonferroni threshold was P < 2.94 x 10-6. Panel A lists the 50 highest-ranked gene sets; panel B lists all 42 gene sets relating to ferroptosis, oxidative stress-induced cell death, lipid peroxidation, glutathione metabolism or iron handling. Long variant and gene lists are abbreviated in this document; complete values are given in the accompanying spreadsheet workbook.*

### Supplementary Table 12. Pathway and biological process enrichment analysis of genes underlying the shared Alzheimer's disease-basal ganglia iron latent factor genome-wide association analysis.

| Term ID | Source | Term | Genes (n) | Gene coverage (%) | Log■■(P) | Log■■(q) | Contributing genes |
|---|---|---|---|---|---|---|---|
| GO:0031341 | GO Biological Processes | regulation of cell killing | 7 | 11.29 | -8.482 | -4.146 | AZGP1;CLU;CR1;CR1L;HLA-DRA;HLA-DRB1;INPP5D |
| GO:0002703 | GO Biological Processes | regulation of leukocyte mediated immunity | 8 | 12.9 | -7.165 | -3.129 | AZGP1;CR1;CR1L;FCER1G;HLA-DRA;HLA-DRB1;INPP5D; SCIMP |
| GO:0002697 | GO Biological Processes | regulation of immune effector process | 9 | 14.52 | -6.815 | -3.013 | AZGP1;CLU;CR1;CR1L;FCER1G;HLA-DRA;HLA-DRB1; INPP5D;SCIMP |
| GO:0002706 | GO Biological Processes | regulation of lymphocyte mediated immunity | 7 | 11.29 | -6.685 | -3.013 | AZGP1;CR1;CR1L;FCER1G;HLA-DRA;HLA-DRB1;INPP5D |
| GO:0002683 | GO Biological Processes | negative regulation of immune system process | 10 | 16.13 | -6.507 | -3.013 | CD33;CLU;CR1;CR1L;FCER1G;HLA-DRB1;INPP5D;BTNL2; PVRIG;RIN3 |
| GO:0002768 | GO Biological Processes | immune response-regulating cell surface receptor signaling pathway | 8 | 12.9 | -6.506 | -3.013 | CD33;CR1;FCER1G;HLA-DRB1;INPP5D;CD2AP;BTNL2; SCIMP |
| GO:0048002 | GO Biological Processes | antigen processing and presentation of peptide antigen | 5 | 8.06 | -6.505 | -3.013 | AZGP1;ACE;FCER1G;HLA-DRA;HLA-DRB1 |
| hsa05310 | KEGG Pathway | Asthma | 4 | 6.45 | -6.258 | -2.824 | MS4A2;FCER1G;HLA-DRA;HLA-DRB1 |
| GO:1903659 | GO Biological Processes | regulation of complement-dependent cytotoxicity | 3 | 4.84 | -6.165 | -2.782 | CLU;CR1;CR1L |
| GO:0034248 | GO Biological Processes | regulation of amide metabolic process | 5 | 8.06 | -6.025 | -2.689 | BIN1;CLU;PICALM;BCKDK;ABCA7 |
| GO:0002429 | GO Biological Processes | immune response-activating cell surface receptor signaling pathway | 7 | 11.29 | -5.955 | -2.659 | CR1;FCER1G;HLA-DRB1;INPP5D;CD2AP;BTNL2;SCIMP |
| GO:0050778 | GO Biological Processes | positive regulation of immune response | 10 | 16.13 | -5.821 | -2.564 | AZGP1;CLU;CR1;FCER1G;HLA-DRA;HLA-DRB1;INPP5D; CD2AP;BTNL2;SCIMP |
| GO:0002764 | GO Biological Processes | immune response-regulating signaling pathway | 8 | 12.9 | -5.735 | -2.539 | CD33;CR1;FCER1G;HLA-DRB1;INPP5D;CD2AP;BTNL2; SCIMP |
| GO:0002253 | GO Biological Processes | activation of immune response | 8 | 12.9 | -5.72 | -2.539 | CLU;CR1;FCER1G;HLA-DRB1;INPP5D;CD2AP;BTNL2; SCIMP |
| hsa04640 | KEGG Pathway | Hematopoietic cell lineage | 5 | 8.06 | -5.7 | -2.539 | CD33;CR1;CR1L;HLA-DRA;HLA-DRB1 |
| GO:0019882 | GO Biological Processes | antigen processing and presentation | 5 | 8.06 | -5.595 | -2.463 | AZGP1;ACE;FCER1G;HLA-DRA;HLA-DRB1 |
| GO:0045916 | GO Biological Processes | negative regulation of complement activation | 3 | 4.84 | -5.531 | -2.425 | CLU;CR1;CR1L |
| GO:1902003 | GO Biological Processes | regulation of amyloid-beta formation | 4 | 6.45 | -5.429 | -2.347 | BIN1;CLU;PICALM;ABCA7 |
| GO:0030100 | GO Biological Processes | regulation of endocytosis | 7 | 11.29 | -5.389 | -2.331 | BIN1;CLU;CNN2;FCER1G;PICALM;ABCA7;RIN3 |
| GO:0002822 | GO Biological Processes | regulation of adaptive immune response based on somatic recombination of immune ... | 6 | 9.68 | -5.332 | -2.297 | AZGP1;CR1;CR1L;FCER1G;HLA-DRA;HLA-DRB1 |
| GO:0002819 | GO Biological Processes | regulation of adaptive immune response | 6 | 9.68 | -5.194 | -2.199 | AZGP1;CR1;CR1L;FCER1G;HLA-DRA;HLA-DRB1 |
| GO:0002921 | GO Biological Processes | negative regulation of humoral immune response | 3 | 4.84 | -5.183 | -2.199 | CLU;CR1;CR1L |
| GO:1902991 | GO Biological Processes | regulation of amyloid precursor protein catabolic process | 4 | 6.45 | -5.173 | -2.199 | BIN1;CLU;PICALM;ABCA7 |
| GO:0002757 | GO Biological Processes | immune response-activating signaling pathway | 7 | 11.29 | -5.151 | -2.194 | CR1;FCER1G;HLA-DRB1;INPP5D;CD2AP;BTNL2;SCIMP |
| GO:0002483 | GO Biological Processes | antigen processing and presentation of endogenous peptide antigen | 3 | 4.84 | -4.85 | -1.911 | AZGP1;HLA-DRA;HLA-DRB1 |
| GO:0045806 | GO Biological Processes | negative regulation of endocytosis | 4 | 6.45 | -4.804 | -1.887 | CNN2;PICALM;ABCA7;RIN3 |
| GO:0045591 | GO Biological Processes | positive regulation of regulatory T cell differentiation | 3 | 4.84 | -4.793 | -1.887 | CR1;HLA-DRA;HLA-DRB1 |
| GO:0002705 | GO Biological Processes | positive regulation of leukocyte mediated immunity | 5 | 8.06 | -4.77 | -1.881 | AZGP1;FCER1G;HLA-DRA;HLA-DRB1;SCIMP |
| GO:0002460 | GO Biological Processes | adaptive immune response based on somatic recombination of immune receptors buil ... | 6 | 9.68 | -4.748 | -1.874 | CLU;CR1;CR1L;FCER1G;HLA-DRB1;INPP5D |
| GO:0002699 | GO Biological Processes | positive regulation of immune effector process | 6 | 9.68 | -4.703 | -1.843 | AZGP1;CR1;FCER1G;HLA-DRA;HLA-DRB1;SCIMP |
| GO:0030449 | GO Biological Processes | regulation of complement activation | 3 | 4.84 | -4.685 | -1.84 | CLU;CR1;CR1L |
| hsa05140 | KEGG Pathway | Leishmaniasis | 4 | 6.45 | -4.668 | -1.837 | CR1;CR1L;HLA-DRA;HLA-DRB1 |
| GO:0097242 | GO Biological Processes | amyloid-beta clearance | 3 | 4.84 | -4.635 | -1.816 | CLU;PICALM;ABCA7 |
| GO:0002443 | GO Biological Processes | leukocyte mediated immunity | 6 | 9.68 | -4.513 | -1.708 | CLU;CR1;CR1L;ACE;FCER1G;INPP5D |
| GO:0019883 | GO Biological Processes | antigen processing and presentation of endogenous antigen | 3 | 4.84 | -4.494 | -1.701 | AZGP1;HLA-DRA;HLA-DRB1 |
| GO:0019886 | GO Biological Processes | antigen processing and presentation of exogenous peptide antigen via MHC class I ... | 3 | 4.84 | -4.45 | -1.67 | FCER1G;HLA-DRA;HLA-DRB1 |
| hsa05152 | KEGG Pathway | Tuberculosis | 5 | 8.06 | -4.435 | -1.667 | CR1;CR1L;FCER1G;HLA-DRA;HLA-DRB1 |
| GO:1902430 | GO Biological Processes | negative regulation of amyloid-beta formation | 3 | 4.84 | -4.408 | -1.651 | BIN1;CLU;ABCA7 |
| GO:0002495 | GO Biological Processes | antigen processing and presentation of peptide antigen via MHC class II | 3 | 4.84 | -4.368 | -1.622 | FCER1G;HLA-DRA;HLA-DRB1 |

| Term ID | Source | Term | Genes (n) | Gene coverage (%) | Log■■(P) | Log■■(q) | Contributing genes |
|---|---|---|---|---|---|---|---|
| GO:0002504 | GO Biological Processes | antigen processing and presentation of peptide or polysaccharide antigen via MHC ... | 3 | 4.84 | -4.328 | -1.604 | FCER1G;HLA-DRA;HLA-DRB1 |
| GO:1902992 | GO Biological Processes | negative regulation of amyloid precursor protein catabolic process | 3 | 4.84 | -4.328 | -1.604 | BIN1;CLU;ABCA7 |
| GO:0031342 | GO Biological Processes | negative regulation of cell killing | 3 | 4.84 | -4.253 | -1.555 | CLU;CR1;INPP5D |
| GO:0002474 | GO Biological Processes | antigen processing and presentation of peptide antigen via MHC class I | 3 | 4.84 | -4.253 | -1.555 | AZGP1;ACE;FCER1G |
| GO:0001910 | GO Biological Processes | regulation of leukocyte mediated cytotoxicity | 4 | 6.45 | -4.249 | -1.555 | AZGP1;HLA-DRA;HLA-DRB1;INPP5D |
| GO:0001916 | GO Biological Processes | positive regulation of T cell mediated cytotoxicity | 3 | 4.84 | -4.182 | -1.498 | AZGP1;HLA-DRA;HLA-DRB1 |
| GO:0034249 | GO Biological Processes | negative regulation of amide metabolic process | 3 | 4.84 | -4.147 | -1.473 | BIN1;CLU;ABCA7 |
| GO:2000008 | GO Biological Processes | regulation of protein localization to cell surface | 3 | 4.84 | -4.019 | -1.355 | FCER1G;PICALM;ABCA7 |
| GO:0050852 | GO Biological Processes | T cell receptor signaling pathway | 4 | 6.45 | -3.986 | -1.33 | HLA-DRB1;INPP5D;CD2AP;BTNL2 |
| GO:0050777 | GO Biological Processes | negative regulation of immune response | 5 | 8.06 | -3.973 | -1.326 | CLU;CR1;CR1L;HLA-DRB1;INPP5D |
| GO:0002478 | GO Biological Processes | antigen processing and presentation of exogenous peptide antigen | 3 | 4.84 | -3.96 | -1.322 | FCER1G;HLA-DRA;HLA-DRB1 |

*Abbreviations: GO = Gene Ontology. Terms were retained where P < 0.01, gene count was at least three and the enrichment factor exceeded 1.5, with significance assessed after Benjamini-Hochberg correction. Enrichment was tested in Metascape against all genes in the genome as background. Long variant and gene lists are abbreviated in this document; complete values are given in the accompanying spreadsheet workbook.*

### Supplementary Table 13. Inverse-variance weighted Mendelian randomisation estimates for the bidirectional association between brain iron and Alzheimer's disease (main analysis, Jansen 2019).

| Direction | Brain region | Instruments (n) | IVW beta (95% CI) | SE | p-value | FDR q | MRlap β (95% CI) | MRlap SE | MRlap p | Δβ (correction) | Mean F | Minimum F |
|---|---|---|---|---|---|---|---|---|---|---|---|---|
| Forward | Caudate nucleus | 93 | -0.0007 (-0.0016, 0.0002) | 0.0005 | 0.148 | 0.295 | -0.0068 (-0.0166, 0.0029) | 0.0050 | 0.170 | -0.0062 | 77.2 | 24.9 |
| Forward | Dentate nucleus | 152 | -0.0003 (-0.0006, 0.0000) | 0.0002 | 0.079 | 0.220 | -0.0060 (-0.0146, 0.0027) | 0.0044 | 0.176 | -0.0057 | 60.8 | 25.3 |
| Forward | Globus pallidus | 86 | -0.0001 (-0.0005, 0.0003) | 0.0002 | 0.651 | 0.760 | -0.0018 (-0.0135, 0.0098) | 0.0060 | 0.758 | -0.0017 | 68.4 | 23.1 |
| Forward | Putamen | 122 | 0.0003 (-0.0001, 0.0008) | 0.0002 | 0.174 | 0.325 | 0.0051 (-0.0058, 0.0161) | 0.0056 | 0.359 | 0.0048 | 65.1 | 23.8 |
| Forward | Red nucleus | 87 | -0.0007 (-0.0011, -0.0002) * | 0.0002 | 0.005 | 0.093 | -0.0141 (-0.0252, -0.0031) | 0.0056 | 0.012 | -0.0135 | 55.3 | 27.3 |
| Forward | Substantia nigra | 49 | -0.0004 (-0.0010, 0.0001) | 0.0003 | 0.100 | 0.254 | -0.0110 (-0.0261, 0.0042) | 0.0077 | 0.156 | -0.0105 | 53.6 | 27.5 |
| Forward | Basal ganglia iron factor | 117 | 0.0046 (-0.0022, 0.0114) | 0.0035 | 0.188 | 0.329 | 0.0072 (-0.0088, 0.0232) | 0.0082 | 0.376 | 0.0026 | 65.1 | 29.9 |
| Reverse | Caudate nucleus | 33 | 0.8686 (-0.5195, 2.2567) | 0.7082 | 0.220 | 0.362 | 0.1208 (-0.1552, 0.3968) | 0.1408 | 0.391 | -0.7478 | 48.8 | 30.3 |
| Reverse | Dentate nucleus | 33 | 1.5620 (-1.7654, 4.8894) | 1.6977 | 0.358 | 0.556 | 0.0925 (-0.1849, 0.3698) | 0.1415 | 0.513 | -1.4695 | 48.8 | 30.3 |
| Reverse | Globus pallidus | 33 | 4.5759 (1.0808, 8.0711) * | 1.7832 | 0.010 | 0.093 | 0.2527 (-0.0507, 0.5560) | 0.1548 | 0.103 | -4.3233 | 48.8 | 30.3 |
| Reverse | Putamen | 33 | 3.0503 (0.6703, 5.4304) * | 1.2143 | 0.012 | 0.093 | 0.2512 (-0.0732, 0.5755) | 0.1655 | 0.129 | -2.7992 | 48.8 | 30.3 |
| Reverse | Red nucleus | 33 | 0.4562 (-2.6674, 3.5798) | 1.5937 | 0.775 | 0.834 | 0.0182 (-0.1933, 0.2297) | 0.1079 | 0.866 | -0.4380 | 48.8 | 30.3 |
| Reverse | Substantia nigra | 33 | 2.8358 (-0.8963, 6.5679) | 1.9041 | 0.136 | 0.295 | 0.1399 (-0.0691, 0.3488) | 0.1066 | 0.190 | -2.6959 | 48.8 | 30.3 |
| Reverse | Basal ganglia iron factor | 32 | 0.2127 (0.0443, 0.3810) * | 0.0859 | 0.013 | 0.093 | 0.1728 (-0.0441, 0.3897) | 0.1107 | 0.118 | -0.0398 | 49.4 | 30.3 |

*Abbreviations: CI = confidence interval; FDR = false discovery rate; IV = instrumental variable; IVW = inverse-variance weighted; MRlap = correction for sample overlap between exposure and outcome datasets; SE = standard error. Beta is the log-odds of Alzheimer's disease per standard deviation of brain iron in the forward direction, and the standard deviation of brain iron per log-odds of Alzheimer's disease in the reverse direction. * P < 0.05 (nominal); dagger, FDR q < 0.05 (Benjamini-Hochberg). The APOE region was excluded throughout.*

### Supplementary Table 14. Sensitivity analysis: inverse-variance weighted Mendelian randomisation estimates using the Kunkle 2019 dataset (no UK Biobank sample overlap; MRlap correction not required).

| Direction | Brain region | Instruments (n) | IVW beta (95% CI) | SE | p-value | FDR q | Mean F | Minimum F | Consistent with Jansen? | Note |
|---|---|---|---|---|---|---|---|---|---|---|
| ■ FORWARD: Brain Iron → AD | | | | | | | | | | |
| Forward | Caudate nucleus | 84 | -0.0005 (-0.0061, 0.0052) | 0.0029 | 0.870 | 0.870 | 73.5 | 24.9 | No | |
| Forward | Dentate nucleus | 145 | 0.0008 (-0.0011, 0.0027) | 0.0010 | 0.423 | 0.601 | 61.5 | 25.3 | No | |
| Forward | Globus pallidus | 80 | 0.0007 (-0.0017, 0.0032) | 0.0013 | 0.567 | 0.690 | 65.3 | 26.6 | No | |
| Forward | Putamen | 116 | -0.0012 (-0.0042, 0.0018) | 0.0015 | 0.429 | 0.601 | 64.9 | 23.8 | No | |
| Forward | Red nucleus | 85 | -0.0029 (-0.0057, -0.0001) * | 0.0014 | 0.046 | 0.142 | 55.6 | 27.3 | Yes | Replicates Jansen result |
| Forward | Substantia nigra | 45 | -0.0011 (-0.0043, 0.0022) | 0.0017 | 0.514 | 0.654 | 53.6 | 27.5 | No | |
| Forward | Basal ganglia iron factor | 112 | 0.0042 (-0.0394, 0.0479) | 0.0223 | 0.849 | 0.870 | 64.4 | 29.9 | No | |
| ■ REVERSE: AD → Brain Iron | | | | | | | | | | |
| Reverse | Caudate nucleus | 17 | 0.3459 (0.0396, 0.6521) * | 0.1563 | 0.027 | 0.107 | 48.1 | 29.8 | No | Novel signal |
| Reverse | Dentate nucleus | 17 | 0.8357 (0.1019, 1.5695) * | 0.3744 | 0.026 | 0.107 | 48.1 | 29.8 | No | Novel signal |
| Reverse | Globus pallidus | 17 | 0.5729 (-0.1981, 1.3439) | 0.3934 | 0.145 | 0.295 | 48.1 | 29.8 | Yes | |
| Reverse | Putamen | 17 | 0.5767 (0.0516, 1.1019) * | 0.2679 | 0.031 | 0.110 | 48.1 | 29.8 | Yes | Replicates Jansen result |
| Reverse | Red nucleus | 17 | 0.1109 (-0.5780, 0.7999) | 0.3515 | 0.752 | 0.834 | 48.1 | 29.8 | No | |
| Reverse | Substantia nigra | 17 | 0.3108 (-0.5125, 1.1340) | 0.4200 | 0.459 | 0.612 | 48.1 | 29.8 | No | |
| Reverse | Basal ganglia iron factor | 17 | 0.0442 (0.0074, 0.0809) * | 0.0187 | 0.018 | 0.103 | 48.1 | 29.8 | Yes | Replicates Jansen result |

*Abbreviations: CI = confidence interval; FDR = false discovery rate; IV = instrumental variable; IVW = inverse-variance weighted; SE = standard error. The Kunkle 2019 dataset contains no UK Biobank participants, so MRlap correction was not applied. * P < 0.05 (nominal); dagger, FDR q < 0.05.*

### Supplementary Table 15. Sensitivity analysis: all Mendelian randomisation estimators for brain iron to Alzheimer's disease (forward direction, Jansen 2019).

| Brain region | Method | Instruments (n) | β | SE | 95% CI Lower | 95% CI Upper | p-value | Egger Intercept | Egger Intercept p | Pleiotropy? |
|---|---|---|---|---|---|---|---|---|---|---|
| Caudate nucleus | | | | | | | | | | |
| Caudate nucleus | IVW | 93 | -0.00068 | 0.00047 | -0.00159 | 0.00024 | 0.148 | — | — | N/A |
| Caudate nucleus | MR-Egger | 93 | 0.00032 | 0.00090 | -0.00144 | 0.00208 | 0.721 | -0.00086 | 0.290 | |
| Caudate nucleus | Weighted Median | 93 | -0.00082 | 0.00083 | -0.00244 | 0.00081 | 0.324 | — | — | N/A |
| Caudate nucleus | Simple Mode | 93 | -0.00118 | 0.00144 | -0.00401 | 0.00165 | 0.415 | — | — | N/A |
| Caudate nucleus | Weighted Mode | 93 | -0.00118 | 0.00085 | -0.00285 | 0.00049 | 0.167 | — | — | N/A |
| Dentate nucleus | | | | | | | | | | |
| Dentate nucleus | IVW | 152 | -0.00029 | 0.00016 | -0.00061 | 0.00003 | 0.079 | — | — | N/A |
| Dentate nucleus | MR-Egger | 152 | -0.00050 | 0.00043 | -0.00134 | 0.00034 | 0.242 | 0.00033 | 0.665 | |
| Dentate nucleus | Weighted Median | 152 | -0.00018 | 0.00026 | -0.00068 | 0.00032 | 0.487 | — | — | N/A |
| Dentate nucleus | Simple Mode | 152 | -0.00018 | 0.00058 | -0.00133 | 0.00097 | 0.760 | — | — | N/A |
| Dentate nucleus | Weighted Mode | 152 | -0.00018 | 0.00042 | -0.00101 | 0.00065 | 0.671 | — | — | N/A |
| Globus pallidus | | | | | | | | | | |
| Globus pallidus | IVW | 86 | -0.00009 | 0.00020 | -0.00049 | 0.00030 | 0.651 | — | — | N/A |
| Globus pallidus | MR-Egger | 86 | 0.00021 | 0.00040 | -0.00057 | 0.00099 | 0.595 | -0.00062 | 0.493 | |
| Globus pallidus | Weighted Median | 86 | -0.00012 | 0.00031 | -0.00073 | 0.00049 | 0.694 | — | — | N/A |
| Globus pallidus | Simple Mode | 86 | -0.00092 | 0.00064 | -0.00217 | 0.00033 | 0.148 | — | — | N/A |
| Globus pallidus | Weighted Mode | 86 | 0.00024 | 0.00038 | -0.00050 | 0.00098 | 0.519 | — | — | N/A |
| Putamen | | | | | | | | | | |
| Putamen | IVW | 122 | 0.00033 | 0.00024 | -0.00015 | 0.00081 | 0.174 | — | — | N/A |
| Putamen | MR-Egger | 122 | -0.00055 | 0.00059 | -0.00171 | 0.00061 | 0.352 | 0.00114 | 0.268 | |
| Putamen | Weighted Median | 122 | -0.00023 | 0.00041 | -0.00104 | 0.00057 | 0.575 | — | — | N/A |
| Putamen | Simple Mode | 122 | -0.00062 | 0.00077 | -0.00213 | 0.00089 | 0.422 | — | — | N/A |
| Putamen | Weighted Mode | 122 | -0.00046 | 0.00052 | -0.00148 | 0.00056 | 0.376 | — | — | N/A |
| Red nucleus | | | | | | | | | | |
| Red nucleus | IVW | 87 | -0.00066 | 0.00024 | -0.00113 | -0.00020 | 0.005* | — | — | N/A |
| Red nucleus | MR-Egger | 87 | -0.00158 | 0.00063 | -0.00281 | -0.00036 | 0.013* | 0.00138 | 0.152 | |
| Red nucleus | Weighted Median | 87 | -0.00093 | 0.00036 | -0.00164 | -0.00022 | 0.011* | — | — | N/A |
| Red nucleus | Simple Mode | 87 | -0.00122 | 0.00080 | -0.00278 | 0.00035 | 0.129 | — | — | N/A |
| Red nucleus | Weighted Mode | 87 | -0.00122 | 0.00066 | -0.00250 | 0.00007 | 0.064 | — | — | N/A |
| Substantia nigra | | | | | | | | | | |
| Substantia nigra | IVW | 49 | -0.00044 | 0.00027 | -0.00096 | 0.00008 | 0.100 | — | — | N/A |
| Substantia nigra | MR-Egger | 49 | 0.00069 | 0.00055 | -0.00038 | 0.00177 | 0.212 | -0.00212 | 0.025 | Possible |
| Substantia nigra | Weighted Median | 49 | -0.00074 | 0.00041 | -0.00155 | 0.00008 | 0.076 | — | — | N/A |
| Substantia nigra | Simple Mode | 49 | -0.00080 | 0.00077 | -0.00230 | 0.00071 | 0.300 | — | — | N/A |
| Substantia nigra | Weighted Mode | 49 | 0.00000 | 0.00060 | -0.00117 | 0.00118 | 0.994 | — | — | N/A |
| Basal ganglia iron factor | | | | | | | | | | |
| Basal ganglia iron factor | IVW | 117 | 0.00460 | 0.00349 | -0.00225 | 0.01144 | 0.188 | — | — | N/A |
| Basal ganglia iron factor | MR-Egger | 117 | -0.00287 | 0.00801 | -0.01857 | 0.01283 | 0.721 | 0.00069 | 0.490 | |
| Basal ganglia iron factor | Weighted Median | 117 | -0.00352 | 0.00574 | -0.01476 | 0.00772 | 0.540 | — | — | N/A |
| Basal ganglia iron factor | Simple Mode | 117 | -0.00297 | 0.01183 | -0.02617 | 0.02022 | 0.802 | — | — | N/A |
| Basal ganglia iron factor | Weighted Mode | 117 | -0.00646 | 0.00823 | -0.02259 | 0.00968 | 0.433 | — | — | N/A |

*Abbreviations: CI = confidence interval; IV = instrumental variable; IVW = inverse-variance weighted; MR = Mendelian randomisation; SE = standard error. IVW was the primary estimator. The MR-Egger intercept tests for directional pleiotropy. * P < 0.05.*

### Supplementary Table 16. Sensitivity analysis: all Mendelian randomisation estimators for Alzheimer's disease to brain iron (reverse direction, Jansen 2019).

| Brain region | Method | Instruments (n) | β | SE | 95% CI Lower | 95% CI Upper | p-value | Egger Intercept | Egger Intercept p | Pleiotropy? |
|---|---|---|---|---|---|---|---|---|---|---|
| Caudate nucleus | | | | | | | | | | |
| Caudate nucleus | IVW | 33 | 0.86859 | 0.70822 | -0.51952 | 2.25670 | 0.220 | — | — | N/A |
| Caudate nucleus | MR-Egger | 33 | 5.69178 | 2.10732 | 1.56144 | 9.82212 | 0.011* | -0.10028 | 0.101 | |
| Caudate nucleus | Weighted Median | 33 | -0.43252 | 1.11849 | -2.62475 | 1.75971 | 0.699 | — | — | N/A |
| Caudate nucleus | Simple Mode | 33 | -1.53217 | 2.43619 | -6.30711 | 3.24277 | 0.529 | — | — | N/A |
| Caudate nucleus | Weighted Mode | 33 | -0.81961 | 1.67999 | -4.11238 | 2.47316 | 0.626 | — | — | N/A |
| Dentate nucleus | | | | | | | | | | |
| Dentate nucleus | IVW | 33 | 1.56198 | 1.69767 | -1.76545 | 4.88941 | 0.358 | — | — | N/A |
| Dentate nucleus | MR-Egger | 33 | 7.56958 | 5.05238 | -2.33308 | 17.47224 | 0.144 | -0.12493 | 0.411 | |
| Dentate nucleus | Weighted Median | 33 | 4.12052 | 2.58738 | -0.95075 | 9.19178 | 0.111 | — | — | N/A |
| Dentate nucleus | Simple Mode | 33 | 2.35945 | 5.27268 | -7.97500 | 12.69391 | 0.655 | — | — | N/A |
| Dentate nucleus | Weighted Mode | 33 | 5.09803 | 3.47725 | -1.71739 | 11.91344 | 0.143 | — | — | N/A |
| Globus pallidus | | | | | | | | | | |
| Globus pallidus | IVW | 33 | 4.57594 | 1.78324 | 1.08080 | 8.07109 | 0.010* | — | — | N/A |
| Globus pallidus | MR-Egger | 33 | 5.54040 | 5.30647 | -4.86029 | 15.94109 | 0.305 | -0.02005 | 0.908 | |
| Globus pallidus | Weighted Median | 33 | 5.25470 | 2.99421 | -0.61396 | 11.12336 | 0.079 | — | — | N/A |
| Globus pallidus | Simple Mode | 33 | 7.25212 | 7.43571 | -7.32187 | 21.82610 | 0.329 | — | — | N/A |
| Globus pallidus | Weighted Mode | 33 | 7.92869 | 6.24810 | -4.31758 | 20.17497 | 0.204 | — | — | N/A |
| Putamen | | | | | | | | | | |
| Putamen | IVW | 33 | 3.05034 | 1.21430 | 0.67032 | 5.43037 | 0.012* | — | — | N/A |
| Putamen | MR-Egger | 33 | 5.36200 | 3.61351 | -1.72048 | 12.44449 | 0.148 | -0.04806 | 0.702 | |
| Putamen | Weighted Median | 33 | 1.63527 | 1.98976 | -2.26466 | 5.53520 | 0.411 | — | — | N/A |
| Putamen | Simple Mode | 33 | 0.30772 | 3.82522 | -7.18972 | 7.80516 | 0.936 | — | — | N/A |
| Putamen | Weighted Mode | 33 | 0.86594 | 2.62150 | -4.27220 | 6.00408 | 0.741 | — | — | N/A |
| Red nucleus | | | | | | | | | | |
| Red nucleus | IVW | 33 | 0.45618 | 1.59366 | -2.66740 | 3.57976 | 0.775 | — | — | N/A |
| Red nucleus | MR-Egger | 33 | 3.90255 | 4.74240 | -5.39256 | 13.19765 | 0.417 | -0.07166 | 0.508 | |
| Red nucleus | Weighted Median | 33 | -0.05084 | 2.49080 | -4.93281 | 4.83114 | 0.984 | — | — | N/A |
| Red nucleus | Simple Mode | 33 | -4.68720 | 5.05122 | -14.58759 | 5.21319 | 0.353 | — | — | N/A |
| Red nucleus | Weighted Mode | 33 | -3.80550 | 4.18612 | -12.01029 | 4.39930 | 0.363 | — | — | N/A |
| Substantia nigra | | | | | | | | | | |
| Substantia nigra | IVW | 33 | 2.83581 | 1.90413 | -0.89629 | 6.56791 | 0.136 | — | — | N/A |
| Substantia nigra | MR-Egger | 33 | 4.67436 | 5.66630 | -6.43159 | 15.78031 | 0.416 | -0.03823 | 0.764 | |
| Substantia nigra | Weighted Median | 33 | 2.92435 | 2.92284 | -2.80441 | 8.65311 | 0.317 | — | — | N/A |
| Substantia nigra | Simple Mode | 33 | 2.52017 | 5.54425 | -8.34656 | 13.38690 | 0.649 | — | — | N/A |
| Substantia nigra | Weighted Mode | 33 | 3.40108 | 5.17961 | -6.75095 | 13.55312 | 0.511 | — | — | N/A |
| Basal ganglia iron factor | | | | | | | | | | |
| Basal ganglia iron factor | IVW | 32 | 0.21265 | 0.08592 | 0.04425 | 0.38105 | 0.013* | — | — | N/A |
| Basal ganglia iron factor | MR-Egger | 32 | 0.48211 | 0.26390 | -0.03515 | 0.99936 | 0.078 | -0.00553 | 0.522 | |
| Basal ganglia iron factor | Weighted Median | 32 | 0.15816 | 0.13580 | -0.10802 | 0.42433 | 0.244 | — | — | N/A |
| Basal ganglia iron factor | Simple Mode | 32 | 0.14796 | 0.25668 | -0.35512 | 0.65105 | 0.564 | — | — | N/A |
| Basal ganglia iron factor | Weighted Mode | 32 | 0.13585 | 0.17517 | -0.20748 | 0.47918 | 0.438 | — | — | N/A |
| * p <0.05. Coloured rows by method. | | | | | | | | | | |

*Abbreviations: CI = confidence interval; IV = instrumental variable; IVW = inverse-variance weighted; MR = Mendelian randomisation; SE = standard error. Format as for Supplementary Table 15, for the reverse direction. * P < 0.05.*

**Supplementary Table 17. Instrument strength and heterogeneity for the bidirectional Mendelian randomisation analysis of brain iron and Alzheimer's disease.**

| Direction | Exposure | Outcome | Instruments (n) | Mean F | Minimum F | Instrument $R^2$ (%) | Cochran's Q | Q degrees of freedom | Q P | $I^2$ (%) |
|---|---|---|---|---|---|---|---|---|---|---|
| Forward | Caudate nucleus | Jansen 2019 | 93 | 77.17 | 24.93 | 20.39 | 138.7 | 92 | 0.001209 | 33.65 |
| Reverse | Jansen 2019 | Caudate nucleus | 33 | 48.78 | 30.29 | 0.3535 | 70.09 | 32 | 0.0001153 | 54.34 |
| Forward | Caudate nucleus | Kunkle 2019 | 84 | 73.55 | 24.93 | 17.55 | 103.1 | 83 | 0.06664 | 19.51 |
| Reverse | Kunkle 2019 | Caudate nucleus | 17 | 48.13 | 29.82 | 1.279 | 40.85 | 16 | 0.0005853 | 60.83 |
| Forward | DentateNucleus | Jansen 2019 | 152 | 60.81 | 25.29 | 26.31 | 234.4 | 151 | 1.588e-05 | 35.59 |
| Reverse | Jansen 2019 | DentateNucleus | 33 | 48.78 | 30.29 | 0.3535 | 72.84 | 32 | 5.067e-05 | 56.07 |
| Forward | DentateNucleus | Kunkle 2019 | 145 | 61.54 | 25.29 | 25.4 | 207.9 | 144 | 0.0003947 | 30.73 |
| Reverse | Kunkle 2019 | DentateNucleus | 17 | 48.13 | 29.82 | 1.279 | 25.64 | 16 | 0.05937 | 37.59 |
| Forward | GlobusPallidus | Jansen 2019 | 86 | 68.4 | 23.09 | 16.74 | 138.8 | 85 | 0.0002059 | 38.78 |
| Reverse | Jansen 2019 | GlobusPallidus | 33 | 48.78 | 30.29 | 0.3535 | 84.47 | 32 | 1.283e-06 | 62.12 |
| Forward | GlobusPallidus | Kunkle 2019 | 80 | 65.31 | 26.65 | 14.87 | 148.4 | 79 | 3.79e-06 | 46.77 |
| Reverse | Kunkle 2019 | GlobusPallidus | 17 | 48.13 | 29.82 | 1.279 | 75.79 | 16 | 9.462e-10 | 78.89 |
| Forward | Putamen | Jansen 2019 | 122 | 65.1 | 23.84 | 22.62 | 264.4 | 121 | 1.068e-12 | 54.23 |
| Reverse | Jansen 2019 | Putamen | 33 | 48.78 | 30.29 | 0.3535 | 96.27 | 32 | 2.358e-08 | 66.76 |
| Forward | Putamen | Kunkle 2019 | 116 | 64.88 | 23.84 | 21.44 | 227.6 | 115 | 2.048e-09 | 49.47 |
| Reverse | Kunkle 2019 | Putamen | 17 | 48.13 | 29.82 | 1.279 | 62.54 | 16 | 1.945e-07 | 74.42 |
| Forward | RedNucleus | Jansen 2019 | 87 | 55.3 | 27.31 | 13.71 | 104.6 | 86 | 0.08427 | 17.78 |
| Reverse | Jansen 2019 | RedNucleus | 33 | 48.78 | 30.29 | 0.3535 | 41.68 | 32 | 0.1174 | 23.23 |
| Forward | RedNucleus | Kunkle 2019 | 85 | 55.57 | 27.31 | 13.47 | 111.3 | 84 | 0.02463 | 24.56 |
| Reverse | Kunkle 2019 | RedNucleus | 17 | 48.13 | 29.82 | 1.279 | 21.07 | 16 | 0.176 | 24.05 |
| Forward | SubstantiaNigra | Jansen 2019 | 49 | 53.6 | 27.54 | 7.484 | 54.24 | 48 | 0.2487 | 11.5 |
| Reverse | Jansen 2019 | SubstantiaNigra | 33 | 48.78 | 30.29 | 0.3535 | 40.18 | 32 | 0.152 | 20.35 |
| Forward | SubstantiaNigra | Kunkle 2019 | 45 | 53.65 | 27.54 | 6.879 | 49.63 | 44 | 0.259 | 11.34 |
| Reverse | Kunkle 2019 | SubstantiaNigra | 17 | 48.13 | 29.82 | 1.279 | 29.33 | 16 | 0.02182 | 45.44 |
| Forward | Basal ganglia iron factor | Jansen 2019 | 117 | 65.13 | 29.87 | 21.69 | 258.9 | 116 | 6.469e-13 | 55.19 |
| Reverse | Jansen 2019 | Basal ganglia iron factor | 32 | 49.35 | 30.29 | 0.3468 | 84.45 | 31 | 7.639e-07 | 63.29 |
| Forward | Basal ganglia iron factor | Kunkle 2019 | 112 | 64.42 | 29.87 | 20.54 | 215.5 | 111 | 1.078e-08 | 48.5 |
| Reverse | Kunkle 2019 | Basal ganglia iron factor | 17 | 48.13 | 29.82 | 1.279 | 67.62 | 16 | 2.595e-08 | 76.34 |

*Abbreviations: I2 = proportion of between-instrument variance attributable to heterogeneity; IV = instrumental variable; Mean F = mean F statistic across instruments; Q = Cochran's Q. Instruments with F below 10 were treated as weak and excluded.*
